# The long-term impact of vaginal surgical mesh devices in UK primary care: a cohort study in the CPRD

**DOI:** 10.1101/2021.07.27.21261171

**Authors:** Emily McFadden, Sarah Lay-Flurrie, Constantinos Koshiaris, Georgia C Richards, Carl Heneghan

## Abstract

**Objective:** Stress urinary incontinence (SUI) and pelvic organ prolapse (POP) are treated with surgical mesh devices; evidence of their long-term complications is lacking. To examine long-term complications in women with SUI and/or POP, with and without surgical mesh implants.

**Design:** Longitudinal open cohort study from April 01 2006 to November 30 2018

**Setting:** The Clinical Practice Research Datalink (CPRD) Gold database, linked to Hospital Episodes Statistics (HES) inpatient data, Office for National Statistics mortality data, and Index of Multiple Deprivation socioeconomic status data.

**Participants:** Women aged ≥18 years with a diagnostic SUI/POP code.

**Exposure:** Mesh surgery coded in HES or CPRD data, compared to no mesh surgery.

**Main Outcomes measures:** Rates of diagnoses of depression, anxiety or self-harm (composite measure) and sexual dysfunction, using Cox proportional hazards regression, and rates of prescriptions for antibiotics and opioids, using negative binomial regression.

**Results:** There were 220,544 women eligible for inclusion; 74% (n=162,687) had SUI, 37% (n=82,123) had POP and 11% (n=24,266) had both. Women undergoing mesh surgery for SUI or POP had higher rates of antibiotic use (SUI: IRR 1.15 (95% CI 1.13 to 1.18; p<0.001); POP: IRR 1.09 (95% CI 1.04 to 1.14; p<0.001)). Women with no previous history of the outcome, who underwent mesh surgery for SUI or POP, had higher rates of depression, anxiety, or self-harm (SUI: HR 2.43 (95% CI 2.19 to 2.70; p<0.001; POP: HR 1.47 (95% CI 1.19 to 1.81; p<0.001)), sexual dysfunction (SUI: HR 1.88 (1.50 to 2.36; p<0.001); POP: HR 1.64 (95% CI 1.02 to 2.63; p=0.04)) and opioid use (SUI: IRR 1.40 (95% CI 1.26 to 1.56, p<0.001); POP: IRR 1.23 (95% CI 1.01 to 1.49; p=0.04)). Women with a history of depression, anxiety and self-harm had lower rates of these outcomes with SUI or POP mesh surgery (SUI: HR 0.70 (95% CI 0.67 to 0.73; p<0.001), POP: HR 0.72 (95% CI 0.65 to 0.79; p<0.001). Women with a history of opioid use who had POP mesh surgery had lower rates of prescriptions (IRR 0.91 95% CI (0.86 to 0.96); p=0.001).

**Conclusions:** Mesh surgery was associated with poor mental and sexual health outcomes, alongside increased opioid and antibiotic use, in women with no history of these outcomes and improved mental health, and lower opioid use, in women with a previous history of these outcomes. Careful consideration of the benefits and risk of mesh surgery for women with SUI or POP on an individual basis is required.

**What is known on this topic:** There are well publicised concerns about high complication rates and harms associated with surgical mesh, a treatment that has been used in urogynaecological procedures to treat stress urinary incontinence (SUI) and pelvic organ prolapse (POP) for the past 20 years. Yet, there is a consistent lack of long-term evidence over the use of surgical mesh beyond surgical readmission, with NICE, Cochrane, and the FDA all reporting a lack of long-term outcome data. This study reports long-term complications in women with stress urinary incontinence (SUI) and/or pelvic organ prolapse (POP), with and without surgical mesh implants.

**What this study adds:** In this cohort of 162,687 women with SUI and 82,123 women with POP, rates of depression, anxiety, self-harm, sexual dysfunction and opioid prescriptions were higher in women with mesh-surgery who had no previous history of these outcomes. Lower rates of depression, anxiety and self-harm were seen in women with a history of these outcomes and SUI or POP mesh surgery. Opioid prescribing was lower in women with a previous history of prescriptions and POP mesh surgery. Careful consideration of the benefits and risk of mesh surgery for women with SUI or POP on an individual basis is required.

## Introduction

Surgical mesh has been used in urogynaecological procedures to treat stress urinary incontinence (SUI) and pelvic organ prolapse (POP) for the past 20 years. There have been concerns, however, about high rates of complications and harms. Complications may include pain, infection, depression, anxiety, sexual dysfunction, mesh erosion and further surgery[1–5]. More than 100,000 women are suing manufacturers globally due to the accumulating reports of harms attributed to mesh[6–8].

Pressures from patient advocacy groups alongside media exposure of harms and of failings in market approvals of mesh devices led the UK government to suspend transvaginal mesh devices in July 2018[9]. However, recommendations differ between national bodies, and evidence is still lacking on the long-term health outcomes for patients who receive surgical mesh devices[10].

Previous research in the UK has used Hospital Episodes Statistics (HES) data to describe complications (mainly rates of reoperation) related to mesh surgery in the hospital setting[5,11]. There is a lack of evidence on long-term outcomes arising from mesh devices beyond readmission[1]. Therefore this study will use the Clinical Practice Research Datalink (CPRD) linked to HES data to examine long-term patient outcomes affecting comorbidity and quality of life, including depression, anxiety and self-harm, and sexual dysfunction, and numbers of prescriptions for antibiotics and opioid pain relief, in patients with SUI and/or POP, both with and without surgical mesh implants.

## Methods

### Study design

We conducted an open cohort study of women aged 18 years and over, registered at practices contributing to the CPRD Gold database from April 01 2006. Women were eligible for inclusion if they had a diagnostic code for SUI or POP recorded in their primary care electronic medical record and they met the following criteria: they were registered at an “up-to-standard” practice, their record was deemed “acceptable” for research based on CPRD defined quality indicators[12], and they were eligible for data linkage. Women entered the study on the latest of the study start date (April 01 2006) or the date they met all eligibility criteria. End of follow-up was the earliest date of death, date of deregistration with the practice or the last date of available primary care and linked data (November 30 2018). CPRD data were linked to HES inpatient data, Office for National Statistics (ONS) mortality data and Index of Multiple Deprivation (IMD) socioeconomic status data.

### Exposures, outcomes, and covariates

The primary exposure was a record of mesh surgery in HES inpatient or CPRD data, compared to those without mesh surgery codes (includes non-mesh surgery). Women who start out in the unexposed (no mesh surgery group) can later enter the exposed group if they have mesh insertion surgery during follow-up; their contribution to the unexposed group was censored the day before the date of insertion. For the exposed group: women who had surgery before study entry were considered exposed from the point of entry; women who had surgery during follow-up entered the study, in the exposed group, at the date of insertion; where relevant, follow-up was censored at the date of mesh removal. Both groups were considered at risk of the outcome from the point of study entry. Codes for mesh surgery for SUI (tension free vaginal tape and transobturator tapes) were introduced in April 2006[1]. Sensitivity analyses excluded women with mesh surgery prior to study entry.

Outcomes considered were new episodes of depression, anxiety or self-harm (composite measure), new episodes of sexual dysfunction, and numbers of prescriptions for antibiotics and opioids. These were identified from coded diagnoses and/or prescriptions in the primary care record (eTable 1 in supplement).

Covariates, measured at study entry, included in the analysis were age, body mass index (BMI), ethnicity, deprivation (quintile of IMD) and general practice region.

### Statistical analysis

All analyses were conducted using Stata, version 14 (StataCorp). Data for women with SUI and POP were analysed separately. Baseline summary statistics (means and standard deviations or numbers and proportions) were calculated for the covariates and summarised by diagnosis and surgery status.

The crude rate of each outcome per 100-person years of follow-up was calculated in each calendar year and presented graphically.

Cox proportional hazards models were used to examine the relationship between mesh surgery and time to new diagnoses of depression, anxiety or self-harm, adjusting for covariates and history of the outcome prior to study entry. Equivalent analyses were carried out for sexual dysfunction. Negative binomial regression modelling was used to examine associations between rates of antibiotic prescriptions and mesh-surgery exposure, adjusting for covariates and previous use. Equivalent analyses were carried out for opioid prescriptions. In subgroup analyses, interaction terms between exposure and previous history of each outcome were fitted, and when interactions were significant (p≤0.05), models were fitted stratifying according to the previous history of each outcome instead of adjusting for this variable.

Post hoc analyses estimated numbers needed to treat (NNT) for outcomes significantly associated with mesh surgery from event rates in each group at 5 and 10 years.[13,14] Prescriptions were analysed using a time to first prescription model.[15]

### Missing data

For diagnoses and prescriptions, an absence of relevant codes was assumed to reflect the absence of disease and treatment, respectively. Data on age and practice region were complete. A missing indicator was included in models where ethnic group data was missing. Multiple imputation was used for missing BMI data: 40 imputed datasets were created by log-transforming BMI and, including all covariates, exposures and outcomes in the imputation model. Final model estimates were derived using Rubin’s rules[16].

### Ethical approval

The protocol for this research was approved by the Independent Scientific Advisory Committee (ISAC) of the Medicines and Healthcare Products Regulatory Agency (protocol number 19_167), and the approved protocol was made available to the journal and reviewers during peer review. Ethical approval for observational research using the CPRD with approval from ISAC has been granted by a National Research Ethics Service Committee (Trent Multi Research Ethics Committee, REC reference number 05/MRE04/87).

## Results

In total, 220,544 women, from 400 practices, were included in the cohort: 74% (n=162,687) with SUI and 37% (n=82,123) with POP, of which 11% (n=24,266) had codes for both SUI and POP (eFigure 1). For SUI there were 657,169 person-years of eligible follow-up and for POP there were 391,481 person-years of follow-up. The median length of follow-up was 2.7 years (IQR: 0.6-6.7 years) for SUI and 4.1 years (IQR: 4.2-7.9) for POP, with a maximum of 12.7 years follow-up in both groups.

Table 1 shows baseline characteristics for included participants by the diagnosis of SUI and POP. More women had mesh surgery after study entry for SUI than for POP (6475 [3.9%] vs 2108 [2.5%). Women with SUI were slightly younger than those with POP (58.9 vs 64.5 years). Other characteristics were similar. Table 2 shows the characteristics of women who had received a mesh device by diagnosis. BMI, and deprivation were similar in those with and without a mesh device. Age was similar in women with POP with and without a mesh device, but women with mesh surgery for SUI were slightly younger. Proportions receiving surgery were slightly higher in white women and slightly lower in all other ethnic groups. Mesh operations were largely carried out after April 01 2006 for SUI, as we would expect with the introduction of specific codes for tension free vaginal tape and transobturator tapes) in April 2006[1]. Operations for POP occurred prior to 2000 through to the end of follow-up.

**Table 1:**
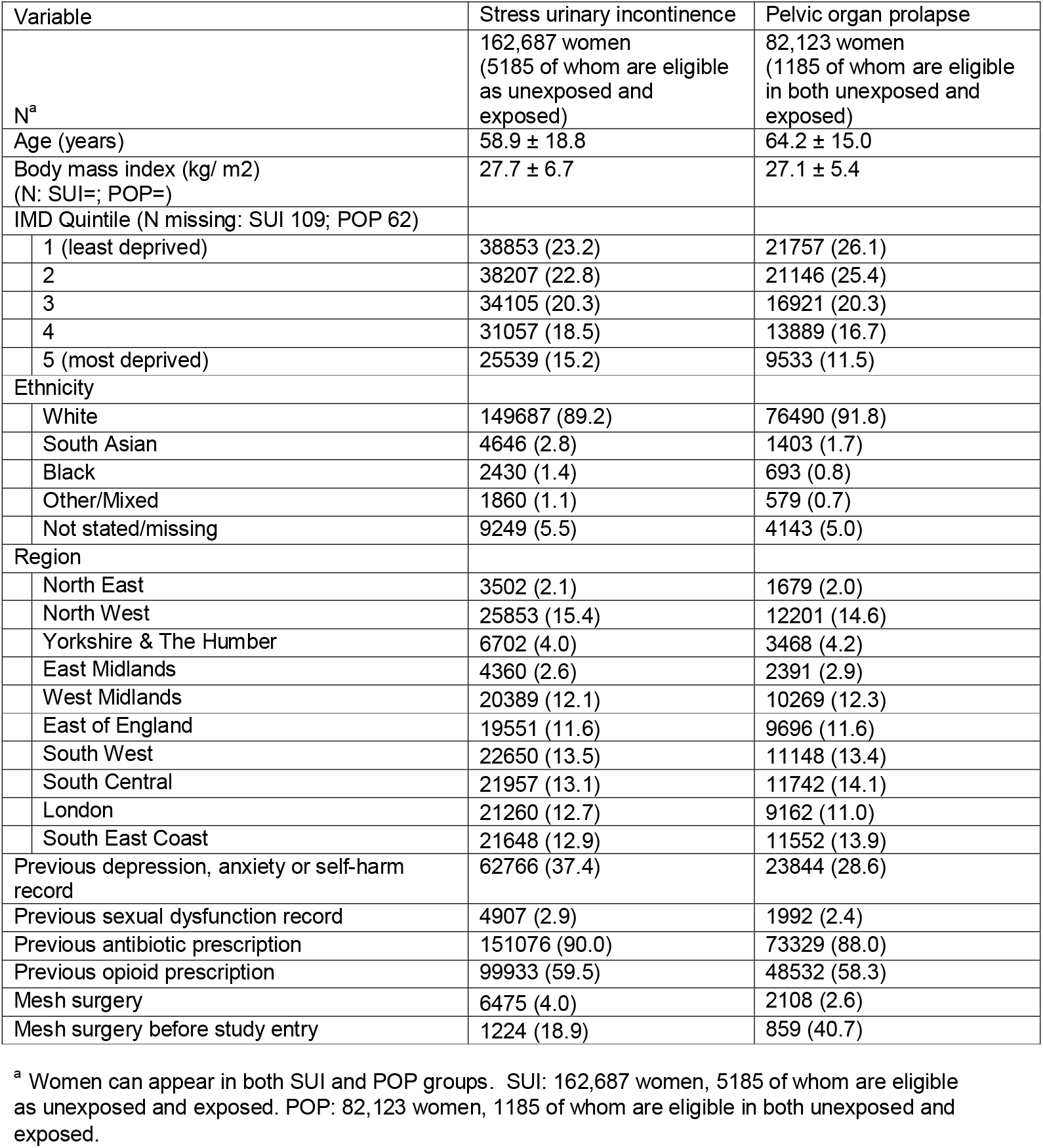
Baseline characteristics of 162,687 women with stress urinary incontinence and 82,123 women with pelvic organ prolapse.

| Variable | Stress urinary incontinence | Pelvic organ prolapse |
| --- | --- | --- |
| N<sup>a</sup> | 162,687 women<br>(5185 of whom are eligible<br>as unexposed and<br>exposed) | 82,123 women<br>(1185 of whom are eligible<br>in both unexposed and<br>exposed) |
| Age (years) | 58.9 ± 18.8 | 64.2 ± 15.0 |
| Body mass index (kg/ m<sup>2</sup>)<br>(N: SUI=; POP=) | 27.7 ± 6.7 | 27.1 ± 5.4 |
| IMD Quintile (N missing: SUI 109; POP 62) |  |  |
| 1 (least deprived) | 38853 (23.2) | 21757 (26.1) |
| 2 | 38207 (22.8) | 21146 (25.4) |
| 3 | 34105 (20.3) | 16921 (20.3) |
| 4 | 31057 (18.5) | 13889 (16.7) |
| 5 (most deprived) | 25539 (15.2) | 9533 (11.5) |
| Ethnicity |  |  |
| White | 149687 (89.2) | 76490 (91.8) |
| South Asian | 4646 (2.8) | 1403 (1.7) |
| Black | 2430 (1.4) | 693 (0.8) |
| Other/Mixed | 1860 (1.1) | 579 (0.7) |
| Not stated/missing | 9249 (5.5) | 4143 (5.0) |
| Region |  |  |
| North East | 3502 (2.1) | 1679 (2.0) |
| North West | 25853 (15.4) | 12201 (14.6) |
| Yorkshire & The Humber | 6702 (4.0) | 3468 (4.2) |
| East Midlands | 4360 (2.6) | 2391 (2.9) |
| West Midlands | 20389 (12.1) | 10269 (12.3) |
| East of England | 19551 (11.6) | 9696 (11.6) |
| South West | 22650 (13.5) | 11148 (13.4) |
| South Central | 21957 (13.1) | 11742 (14.1) |
| London | 21260 (12.7) | 9162 (11.0) |
| South East Coast | 21648 (12.9) | 11552 (13.9) |
| Previous depression, anxiety or self-harm<br>record | 62766 (37.4) | 23844 (28.6) |
| Previous sexual dysfunction record | 4907 (2.9) | 1992 (2.4) |
| Previous antibiotic prescription | 151076 (90.0) | 73329 (88.0) |
| Previous opioid prescription | 99933 (59.5) | 48532 (58.3) |
| Mesh surgery | 6475 (4.0) | 2108 (2.6) |
| Mesh surgery before study entry | 1224 (18.9) | 859 (40.7) |

<sup>a</sup> Women can appear in both SUI and POP groups. SUI: 162,687 women, 5185 of whom are eligible as unexposed and exposed. POP: 82,123 women, 1185 of whom are eligible in both unexposed and exposed.

**Table 2:** Baseline characteristics of 162,687 women with stress urinary incontinence and 82,123 women with pelvic organ prolapse by mesh surgery status.

| Data are mean ± SD or % (N) | | Stress urinary incontinence | | Pelvic organ prolapse | |
| --- | --- | --- | --- | --- | --- |
| Variable |  | No mesh surgery | mesh surgery | No mesh surgery | mesh surgery |
| N<sup>a</sup> |  | 161397 | 6475 | 81200 | 2108 |
| Age (years) | | 59.1 ± 19.0 | 54.4 ± 12.5 | 64.2 ± 15.0 | 63.9 ± 11.8 |
| Body mass index (kg/ m2)<br>(N: SUI=; POP=) | | 27.7 ± 6.7 | 28.4 ± 6.0 | 27.1 ± 5.4 | 26.9 ± 4.7 |
| IMD Quintile (N missing: SUI 109; POP 62) |  |  |  |  |  |
| 1 (least deprived) |  | 37368 (23.2) | 1485 (22.9) | 21216 (26.1) | 541 (25.7) |
| 2 |  | 36664 (22.7) | 1543 (23.8) | 20596 (25.4) | 550 (26.1) |
| 3 |  | 32737 (20.3) | 1368 (21.1) | 16496 (20.3) | 425 (20.2) |
| 4 |  | 29822 (18.5) | 1235 (19.1) | 13530 (16.7) | 359 (17.0) |
| 5 (most deprived) |  | 24699 (15.3) | 840 (13.0) | 9301 (11.5) | 232 (11.0) |
| Ethnicity |  |  |  |  |  |
| White |  | 143502 (88.9) | 6185 (95.5) | 74452 (91.7) | 2038 (96.7) |
| South Asian |  | 4558 (2.8) | 88 (1.4) | 1384 (1.7) | 19 (0.9) |
| Black |  | 2393 (1.5) | 37 (0.6) | 677 (0.8) | 16 (0.8) |
| Other/Mixed |  | 1810 (1.1) | 50 (0.8) | 570 (0.7) | 9 (0.4) |
| Not stated |  | 9134 (5.7) | 115 (1.8) | 4117 (5.1) | 26 (1.2) |
| Region |  |  |  |  |  |
| North East |  | 3391 (2.1) | 111 (1.7) | 1656 (2.0) | 23 (1.1) |
| North West |  | 24965 (15.5) | 888 (13.7) | 11914 (14.7) | 287 (13.6) |
| Yorkshire & The Humber |  | 6496 (4.0) | 206 (3.2) | 3404 (4.2) | 64 (3.0) |
| East Midlands |  | 4230 (2.6) | 130 (2.0) | 2329 (2.9) | 62 (2.9) |
| West Midlands |  | 19687 (12.2) | 702 (10.8) | 10034 (12.4) | 235 (11.1) |
| East of England |  | 18859 (11.7) | 692 (10.7) | 9403 (11.6) | 293 (13.9) |
| South West |  | 21783 (13.5) | 867 (13.4) | 10758 (13.2) | 390 (18.5) |
| South Central |  | 20919 (13.0) | 1038 (16.0) | 11473 (14.1) | 269 (12.8) |
| London |  | 20564 (12.7) | 696 (10.7) | 8976 (11.1) | 186 (8.8) |
| South East Coast |  | 20503 (12.7) | 1145 (17.7) | 11253 (13.9) | 299 (14.2) |
| Previous depression, anxiety or self-harm record |  | 58728 (36.4) | 4038 (62.4) | 22985 (28.3) | 859 (40.7) |
| Previous sexual dysfunction record |  | 4499 (2.8) | 408 (6.3) | 1916 (2.4) | 76 (3.6) |
| Previous antibiotic prescription |  | 145114 (89.9) | 5962 (92.1) | 71436 (88.0) | 1893 (89.8) |
| Previous opioid prescription |  | 95935 (59.4) | 3998 (61.7) | 47175 (58.1) | 1357 (64.4) |
| Mesh surgery before entry |  |  | 1224 (18.9) |  | 859 (40.7) |
| Surgical speciality |  |  |  |  |  |
| Gynaecology |  |  | 5119 (79.1) |  | 1814 (86.1) |
| Obstetric |  |  | 637 (9.8) |  | 214 (10.2) |
| Urology |  |  | 704 (10.9) |  | 10 (0.5) |
| Other |  |  | 15 (0.2) |  | 70 (3.3) |
| Year of mesh surgery |  |  |  |  |  |
| Before 1/4/2006<sup>b</sup> |  |  | 151 (2.3) |  | 638 (32.0) |
| 1/4/2006-2009 |  |  | 2786 (43.0) |  | 507 (25.5) |
| 2010-2014 |  |  | 2986 (46.1) |  | 688 (34.5) |
| $\geq 2015$ | | | 552 (8.5) | | 159 (8.0) |

<sup>a</sup> Women can appear in both surgery groups: women were in the unexposed group up until the date of mesh surgery, at which point they joined the exposed group and were followed until the end-of-follow up or date of mesh removal. Women who had mesh surgery before study entry were considered exposed from the point of study entry. SUI: 162,687 women, 5185 of whom are eligible as unexposed and exposed. POP: 82,123 women, 1185 of whom are eligible in both unexposed and exposed.
<sup>b</sup> April 01 2006 is the study start date as specific codes for SUI mesh only introduced on this date

### Stress urinary incontinence

Unadjusted rates of depression, anxiety, and self-harm were higher in women who had mesh surgery (Figure 1A; Table 3: unadjusted HR 1.60, 95% CI 1.54 to 1.66; p<0.001). After adjustment the association was reversed (adjusted HR 0.78, 95% CI 0.78 to 0.81; p<0.001). Subgroup analyses showed that in women with no previous history of depression, anxiety or self-harm, the risk of an episode was greater with mesh surgery (adjusted HR 2.43 (2.19 to 2.70; p<0.001). In women who had a previous history, the risk of further episodes of depression, anxiety or self-harm was lower in women who had mesh surgery, even after adjustment for covariates (adjusted HR 0.70, 95% CI 0.67 to 0.73; p<0.001).

**Figure 1:**
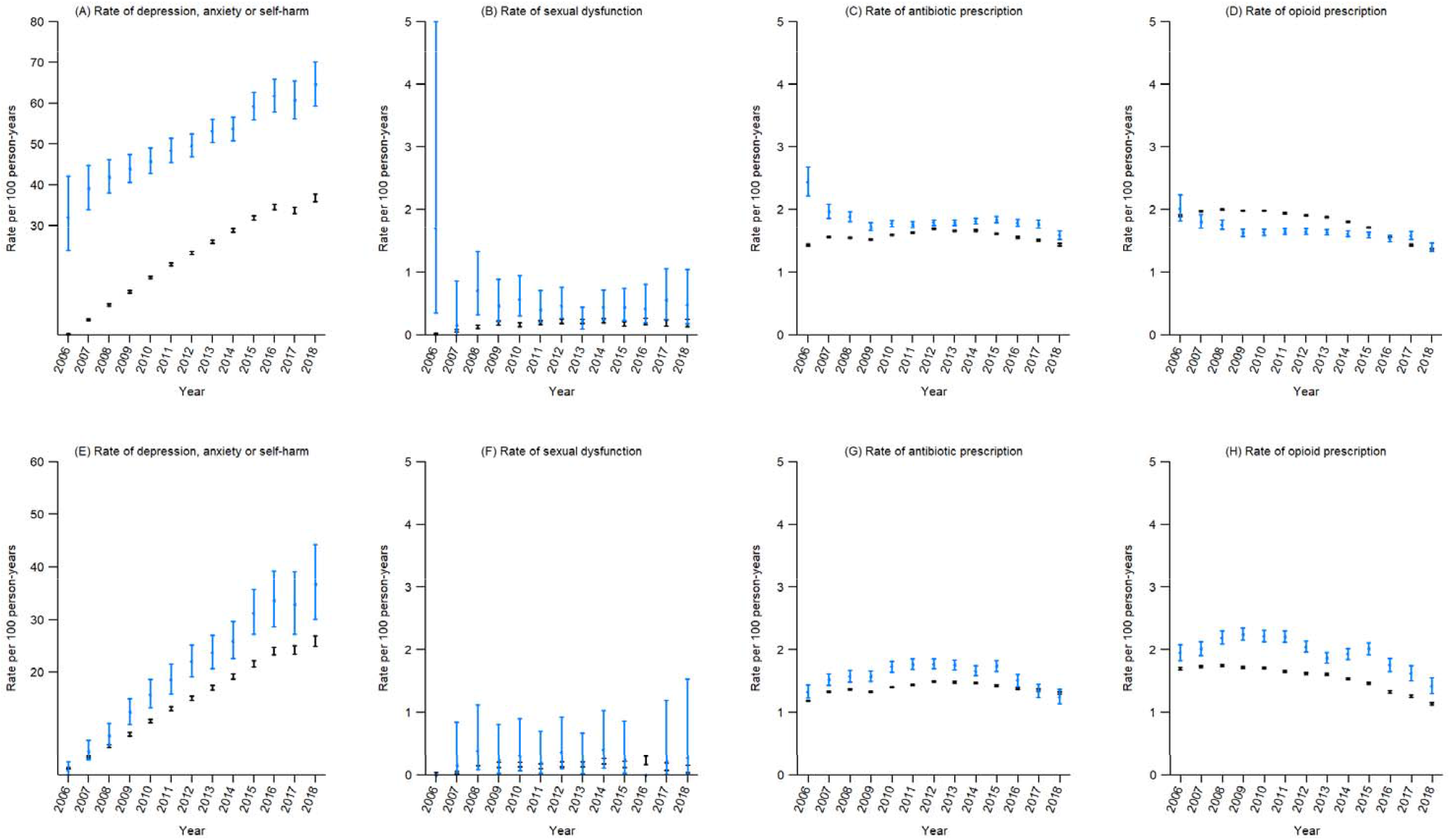
Annual rates and 95% confidence intervals of each outcome in 162,687 women with stress urinary incontinence (A to D) and 82,123 women with pelvic organ prolapse (E to H)

**Table 3:** Unadjusted and adjusted^a^ hazard ratios/incidence rate ratios for outcomes, comparing mesh surgery with no surgery, in 162,578 women with stress urinary incontinence and 82,061 women with pelvic organ prolapse.

| Ratios are for mesh surgery, reference group of no surgery | Depression anxiety and self-harm |  |  |  | Sexual dysfunction |  |  |  | Antibiotic prescriptions |  |  |  | Opioid prescriptions |  |  |  |
| --- | --- | --- | --- | --- | --- | --- | --- | --- | --- | --- | --- | --- | --- | --- | --- | --- |
|  | Unadjusted |  | Adjusted <sup>a</sup> |  | Unadjusted |  | Adjusted <sup>a</sup> |  | Unadjusted |  | Adjusted <sup>a</sup> |  | Unadjusted |  | Adjusted <sup>a</sup> |  |
|  | HR | p-value | HR | p-value | HR | p-value | HR | p-value | IRR | p-value | IRR | p-value | IRR | p-value | IRR | p-value |
|  | (95% CI) |  | (95% CI) |  | (95% CI) |  | (95% CI) |  | (95% CI) |  | (95% CI) |  | (95% CI) |  | (95% CI) |  |
| **Stress urinary incontinence** |  |  |  |  |  |  |  |  |  |  |  |  |  |  |  |  |
| All women | 1.60<br>(1.54,1.66) | <0.001 | 0.78<br>(0.75,0.81) | <0.001 | 1.88<br>(1.55,2.29) | <0.001 | 1.55<br>(1.27,1.90) | <0.001 | 1.15<br>(1.12,1.18) | <0.001 | 1.15<br>(1.13,1.18) | <0.001 | 1.20<br>(1.17,1.23) | <0.001 | 0.93<br>(0.90,0.96) | <0.001 |
| No previous history of the outcome | 2.18<br>(1.97,2.42) | <0.001 | 2.43<br>(2.19,2.70) | <0.001 | 1.93<br>(1.55,2.41) | <0.001 | 1.88<br>(1.50,2.36) | <0.001 | 1.37<br>(1.21,1.55) | <0.001 | 1.47<br>(1.30,1.66) | <0.001 | 1.29<br>(1.16,1.44) | <0.001 | 1.40<br>(1.26,1.56) | <0.001 |
| Previous history of the outcome | 0.70<br>(0.67,0.73) | <0.001 | 0.70<br>(0.67,0.73) | <0.001 | 0.74<br>(0.48,1.15) | 0.19 | 0.86<br>(0.55,1.35) | 0.52 | 1.14<br>(1.11,1.17) | <0.001 | 1.17<br>(1.14,1.20) | <0.001 | 1.15<br>(1.12,1.18) | <0.001 | 1.02<br>(0.99,1.05) | 0.25 |
| **Pelvic organ prolapse** |  |  |  |  |  |  |  |  |  |  |  |  |  |  |  |  |
| All women | 1.28<br>(1.17,1.39) | <0.001 | 0.77<br>(0.70,0.84) | <0.001 | 1.33<br>(0.85,2.07) | 0.22 | 1.19<br>(0.76,1.87) | 0.45 | 1.13<br>(1.08,1.19) | <0.001 | 1.09<br>(1.04,1.14) | <0.001 | 1.03<br>(0.98,1.08) | 0.29 | 0.79<br>(0.74,0.84) | <0.001 |
| No previous history of the outcome | 1.39<br>(1.12,1.71) | 0.002 | 1.47<br>(1.19,1.81) | <0.001 | 1.52<br>(0.95,2.44) | 0.08 | 1.64<br>(1.02,2.63) | 0.04 |  |  |  |  | 1.27<br>(1.05,1.55) | 0.01 | 1.23<br>(1.01,1.49) | 0.04 |
| Previous history of the outcome | 0.70<br>(0.64,0.77) | <0.001 | 0.72<br>(0.65,0.79) | <0.001 | 0.34<br>(0.08,1.36) | 0.13 | 0.32<br>(0.08,1.31) | 0.11 |  |  |  |  | 1.01<br>(0.96,1.05) | 0.77 | 0.91<br>(0.86,0.96) | 0.001 |

<sup>a</sup> Rows for all women were adjusted for age (5 year groups), body mass index (<18.5, 18.5-24.9, 25-29.9, ≥30 kg/m<sup>2</sup>), deprivation quintiles, ethnicity ("white", "Asian", "black", "other"/"mixed", missing), Strategic Health Authority region, history of the outcome. Rows stratified by previous history of the outcome are only presented for models where the interaction between exposure and previous history of the outcome were significant (p≤0.05). These rows were adjusted for all the above variables except for previous history of the outcome.

Rates of sexual dysfunction were higher in women with SUI who had mesh surgery compared with those who had not (Figure 1B; Table 3: unadjusted HR 1.88, 95% CI 1.55 to 2.29; p<0.001; adjusted HR 1.55, 95% CI 1.27 to 1.90; p<0.001). Subgroup analyses showed an increased risk of sexual dysfunction in women who had mesh surgery in women who had a previous history of sexual dysfunction, adjusted HR 1.88 (95% CI 1.50 to 2.36; p<0.001) and no significant association in those with no previous history.

Rates of antibiotic prescriptions were higher in women who had mesh surgery (Figure 1C; Table 3: unadjusted IRR 1.15, 95% CI 1.12 to 1.18; p<0.001; adjusted IRR 1.15, 95% CI 1.13 to 1.18; p<0.001). There was an increased rate of antibiotic prescriptions in women who had mesh surgery, in both women with and without a previous history of antibiotic prescriptions; the rate appeared higher in women with no previous history (Table 3).

There was no clear trend over time in the rate of opioid prescriptions in women who did and did not have mesh surgery (Figure 1D). In multivariate analyses, an increased rate of opioid prescription was seen in women who had mesh surgery, but this association was reversed after adjustment (Table 3). Subgroup analyses showed that in women with no previous history of an opioid prescription there was an increased rate of a new prescription in those who had mesh surgery, adjusted IRR 1.40 (95% CI 1.26 to 1.56; p<0.001), which was not seen in those with prior prescription of opioids, adjusted IRR 1.02 (95% CI 0.99 to 1.05; p=0.25).

### Pelvic organ prolapse

Unadjusted rates of depression, anxiety, and self-harm were higher in women who had mesh surgery (Figure 1E; Table 3: unadjusted HR 1.28, 95% CI 1.17 to 1.39; p<0.01). After adjustment, there was a lower risk of depression, anxiety and self-harm in those with mesh surgery (adjusted HR 0.77, 95% CI 0.70 to 0.84; p<0.001). Subgroup analyses showed that women without a previous history of depression, anxiety or self-harm had a greater risk of these outcomes after mesh-surgery, adjusted HR 1.47 (95% CI 1.19 to 1.81; p<0.001), and women with a history of these outcomes had a lower risk after mesh surgery (adjusted HR 0.72 (95% CI 0.65 to 0.79; p<0.001).

No clear association between rates of sexual dysfunction and mesh surgery were found (Figure 1F; Table 3); event numbers are small, and confidence intervals are wide. Subgroup analyses suggest that women with no previous history of sexual dysfunction had a greater risk of these outcomes after mesh-surgery, adjusted HR 1.64 (95% CI 1.02 to 2.64; p=0.04). In women with a previous history of these outcomes, the risk of a further episode was not associated with mesh surgery.

Rates of antibiotic prescriptions were higher in women who had mesh surgery (Figure 1G, Table 3: unadjusted IRR 1.13, 95% CI 1.08 to 1.19; p<0.001; adjusted IRR 1.09, 95% CI 1.04 to 1.14; p<0.001). Interaction terms were not significant, so subgroup analyses are not presented.

Unadjusted rates of opioids were higher in women with mesh surgery (Figure 1H; Table3). After adjustment, rates of opioid prescriptions were reduced in women who had mesh surgery (IRR 0.79, 95% CI 0.74 to 0.84; p<0.001). Subgroup analyses showed that in women with no previous history of an opioid prescription, there was an increased rate of a new opioid prescription, adjusted IRR 1.23 (95% CI 1.01 to 1.49; p=0.04), but in those with a previous prescription, rates of further prescriptions were lower, adjusted IRR 0.91 (95% CI 0.86 to 0.96; p=0.001).

Table 4 presents NNT and NNH for each outcome at 5 and 10 years; positive numbers should be interpreted as NNT, and negative numbers should be interpreted as NNH. For example, at 5 years, overall, for every 10.6 (10.6 to 10.7) women with no previous history of depression, anxiety and self-harm, who undergo SUI mesh surgery, there is one additional episode of depression, anxiety and self-harm. Whereas in women with a previous history of depression, anxiety and self-harm, at 5 years, for every 18 women (95% CI 17.6 to 18.3) who undergo SUI mesh surgery, there is one fewer episode of depression, anxiety and self-harm. There was no significant interaction between a previous history of antibiotic or opioid prescriptions and mesh exposure, thus these analyses and NNH are presented for all women.

**Table 4:** Numbers needed to treat/harm^a^ and estimated 95% confidence at 5 and 10 years follow-up, for each outcome, for 162,578 women with stress urinary incontinence and 82,061 women with pelvic organ prolapse, and stratified by a previous history of the outcome^b^.

|  | Depression anxiety and self-harm |  | Sexual dysfunction |  | Antibiotic prescriptions |  | Opioid prescriptions |  |
| --- | --- | --- | --- | --- | --- | --- | --- | --- |
|  | NNT 5 years | NNT 10 years | NNT 5 years | NNT 10 years | NNT 5 years | NNT 10 years | NNT 5 years | NNT 10 years |
| Stress urinary incontinence |  |  |  |  |  |  |  |  |
| All women | 23.1<br>(22.8,23.3) | 22.9<br>(22.7,23.1) | -206.9<br>(-271.4,-142.4) | -143.1<br>(-186.8,-99.4) | -21.3<br>(-21.4,-21.1) | -42.9<br>(-43.3,-42.6) | -276.0<br>(-376.9,-175.1) | -292.4<br>(-399.1,-185.6) |
| No previous history of the outcome | -10.6<br>(-10.7,-10.6) | -8.2<br>(-8.3,-8.2) | -159.3<br>(-199.8,-118.8) | -110.2<br>(-137.9,-82.5) |  |  |  |  |
| Previous history of the outcome | 18.0<br>(17.6,18.3) | 19.2<br>(18.8,19.6) | <sup>c</sup> | <sup>c</sup> |  |  |  |  |
| Pelvic organ prolapse |  |  |  |  |  |  |  |  |
| All women | 28.9<br>(28.4,29.3) | 27.1<br>(26.7,27.5) | -726.1<br>(-901.2,-550.9) | -544.2<br>(-674.0,-414.4) | -22.4<br>(-22.9,-22.0) | -41.3<br>(-42.0,-40.5) | -42.8<br>(-49.0,-36.6) | -44.6<br>(-51.0,-38.2) |
| No previous history of the outcome | -45.1<br>(-46.2,-43.9) | -32.6<br>(-33.5,-31.8) | -271.5<br>(-328.3,-214.8) | -205.7<br>(-248.4,-163.0) |  |  |  |  |
| Previous history of the outcome | 15.7<br>(15.4,16.0) | 15.7<br>(15.4,16.1) | <sup>c</sup> | <sup>c</sup> |  |  |  |  |

<sup>a</sup> Positive numbers should be interpreted as the number needed to treat <sup>d</sup> for example, at 5 years, for every 18.0 (95% CI 17.6 to 18.3) women with a previous history of depression, anxiety and self-harm who undergo SUI mesh surgery, there is 1 fewer diagnosis of depression, anxiety and self-harm. Negative numbers should be interpreted as a number needed to harm: for example, at 5 years, for every 10.6 women (95% CI 10.6 to 10.7) with no previous history of depression, anxiety and self-harm who undergo SUI mesh surgery, there is 1 extra episode of these outcomes.
<sup>b</sup> estimated from a Cox proportional hazards model, so time to first diagnosis of depression, anxiety or self-harm, or time to first prescription of antibiotics, adjusted for age (5 year groups), body mass index (<18.5, 18.5-24.9, 25-29.9, ≥30 kg/m<sup>2</sup>), deprivation quintiles, ethnicity ("white", "Asian", "black", "other"/"mixed", missing), Strategic Health Authority region, history of the outcome. Rows stratified by previous history of the outcome are only presented for models where the interaction between exposure and previous history of the outcome were significant (p≤0.05). These rows were adjusted for all the above variables except for previous history of the outcome. Hazard ratios and 95% confidence intervals for depression, anxiety and self-harm and sexual dysfunction are shown in Table 3, and in eTable2 for antibiotic and opioid prescriptions; NNT are only calculated for significant HR.
<sup>c</sup> due to very small numbers in women aged ≥80 years with mesh surgery (<5 women), confidence intervals could not be properly estimated

Sensitivity analyses excluding women who underwent mesh surgery prior to study entry gave similar results (eTable 3).

## Discussion

In this large study of electronic health records, rates of antibiotic prescriptions were higher in women undergoing mesh surgery for SUI or POP. Rates of depression, anxiety, self-harm, sexual dysfunction and opioid prescriptions were higher in women with no previous history of these outcomes who had mesh surgery for SUI or POP, with NNH at 5 years ranging from 10.6 to 271.5. A lower rate of depression, anxiety and self-harm was seen in women with a history of these outcomes who had mesh surgery for SUI or POP, with NNT at 5 years ranging from 15.7 to 19.2. Lower rates of opioid prescriptions in were seen in women with a previous history of prescriptions who had mesh surgery for POP. Our results suggest careful consideration of the benefits and risk of mesh surgery for women with SUI or POP on an individual basis is required.

### Strength and limitations

This is a large-scale analysis of high-quality data, representative of the UK population[21], however we were only able to study women in English practices as linkage is required to obtain data on our primary exposure, mesh surgery. Previous studies[22] have shown that age distributions of patients with and without linkage are similar and, since everyone living in England is entitled to access national health services, we believe selection bias was minimal.

Mesh surgery status should be well recorded in HES data, but there may be some misclassification of exposure status if patients have mesh insertion surgery prior to registering with the CPRD practice contributing data. Given it is relatively major surgery, it may be coded retrospectively. We also used codes for repair/removal of mesh devices to assess exposure status. It is plausible that different mesh devices may have different complication rates however information on the specific type of mesh device is not available. There is often a lag between allocating a specific code to a procedure and the procedure coming into use. Codes for mesh surgery for SUI (tension free vaginal tape and transobturator tapes) were introduced in April 2006[1]. Prior to this date more general codes were used, and we therefore may have missed some women who had surgery and misclassified them as unexposed to mesh surgery.

A major limitation of this study is that those who choose to undergo mesh surgery may differ from those who do not, including by disease severity. The prevalence of a previous history of depression, anxiety and self-harm and sexual dysfunction was greater in those who had mesh surgery. Our multivariate subgroup analyses were considered hypothesis generating, with the aim to identify factors that may increase the risk of patient outcomes. Our descriptive comparisons between patients who did and did not have mesh devices fitted may be particularly vulnerable to confounding by indication, but we still feel that this comparison is worth including to give some indication of rates of each outcome in a group of patients who may have been eligible for a mesh device. Sensitivity analyses including women with mesh surgery at or after study entry showed similar results.

Depression, anxiety and self-harm, and sexual dysfunction may have been under ascertained, as symptom data is known to not be well recorded: some patients with these conditions will not be diagnosed and some will not have their diagnosis recorded[23]. We used published strategies and code lists that included both diagnostic and prescription codes to try minimise this[24]. Given the media exposure about mesh surgery, sexual dysfunction recording may also be subject to reporting bias.

### Comparison with the existing literature

In 2017, NICE draft guidance[17] recommended the repair of POP using transvaginal mesh should not be done except in the context of research. And in July 2018, Baroness Cumberlege, chair of the Independent Medicines and Medical Devices Safety Review (IMMDS) [1], called for a halt in the use of mesh for SUI until conditions affecting training, registration, and licensing were met. Based on updated NICE guidance[18], in April 2019, the mesh ban was lifted, with changes. NICE recommended that mesh could be used once certain conditions were met. Although it is still subject to a period of “high vigilance restriction.”

One of the main problems was the lack of evidence informing long term complications in NICE’s evidence review. For SUI, NICE found 141 studies and reported 109 RCTs on clinical effectiveness and short- and medium-term surgery complications[18]. There were 259 instances of very low-quality evidence across the statements and only one high-quality evidence statement in one RCT. Overall, interpretation of the results was limited by the quality of the data, and the trials were too short to inform long term complications and were not able to detect serious adverse events or postoperative complications. For POP, NICE reported 46 studies of short-term complications following surgery, 24 for mid-term complications, and 17 on long-term complications[18]. Only one instance of high-quality evidence was reported. For POP with SUI, four articles reported data from three trials. Because of the lack of evidence, NICE recommended that it is essential to explain this gap in longer-term outcomes to women prior to treatment.

There has been a consistent lack of long-term evidence over the use of surgical mesh. NICE, Cochrane, and the FDA have reported a lack of long-term outcome data. In the Cochrane review on surgery for women with POP with or without SUI, 19 RCTs reported that ‘adverse events were infrequently reported in all studies’[19]. A further Cochrane review of mid-urethral sling operations for SUI in women including 81 studies, reported the occurrence of problems with sexual intercourse involving pain was low, in contrast to our findings. The review, however, reports only ten trials assessed this outcome with follow-up that spanned six to 24 months[20]. To date there has been a lack of data in primary care, and long-term follow up and hence the need for this study.

### Implications for clinicians and policymakers

Our results suggest careful consideration of the benefits and risk of mesh surgery for women with SUI or POP on an individual basis is required. The IMMDS review, published in 2020[1], reported ‘the system does not know the true long-term complication rate for pelvic mesh procedures’ and that it ‘is impossible to know how many women would have chosen a different form of treatment – a different care pathway – if only they had been given the information they needed to make a fully-informed choice.’ We consider the primary care data presented here essential to informed decision making.

Recommendation 7 in the IMMDS review[1] sets out the need for a central patient-identifiable database. The implantation of devices at the time of the operation should then ‘be linked to specifically created registers to research and audit the outcomes both in terms of the device safety and patient reported outcomes measures.’ We consider the linking of the data to primary care essential and that the data analysis and dissemination be done in real time to facilitate timely decision making.

## Conclusions

In a large cohort of women in primary care, rates of antibiotic prescriptions were increased in those who underwent mesh surgery for SUI and POP; rates of depression, anxiety, and self-harm, sexual dysfunction and opioids prescriptions were increased in those who had no history of these outcomes and underwent SUI or POP mesh surgery; rates of depression, anxiety and self-harm were lower in those with a previous history who underwent SUI or POP mesh surgery; rates of opioid use were lower in women with a previous history who underwent POP mesh surgery. Careful consideration of the benefits and risk of mesh surgery for women with SUI or POP on an individual basis is required, and the linking of registers and auditing of clinical outcomes for mesh is essential to inform decision making.

## Supporting information

eFigure 1, eTable 2, eTable 3

eTable 1

## Data Availability

Data sharing: To guarantee the confidentiality of personal and health information only the authors have had access to the data during the study in accordance with the relevant licence agreements. CPRD linked data were provided under a licence that does not permit sharing.

## Footnotes

## Acknowledgements

CH conceived the study. EM, GR and CH designed the study. EM and SLF carried out the acquisition of data, data management and statistical analysis, with advice from CK. SLF and EM had full access to all the data in the study and takes responsibility for the integrity of the data and the accuracy of the data analysis. EM and CH wrote the first draft of the manuscript. All authors were involved in the drafting and commenting on further revisions of the manuscript. All authors read and approved the final manuscript.

## Competing interests

All authors have completed the ICMJE uniform disclosure form at www.icmje.org/coi_disclosure.pdf. EM has no conflicts of interest. SLF was supported by the NIHR Biomedical Research Centre (Oxford) and NIHR Applied Research Collaboration Oxford and Thames Valley during the conduct of the study, but outside the submitted work. CK is currently supported by a Wellcome Trust/Royal Society Sir Henry Dale Fellowship, but outside the submitted work. GCR was financially supported by the National Institute of Health Research (NIHR) School for Primary Care Research (SPCR), the Naji Foundation, and the Rotary Foundation to study for a Doctor of Philosophy, but no longer has any conflicts of interest. CH holds grant funding from the NIHR School of Primary Care Research Evidence Synthesis Working Group [Project 390], the NIHR BRC Oxford and the World Health Organization for a series of Living rapid review on the modes of transmission of SARs-CoV-2. CH has received financial remuneration from an asbestos case and given legal advice on mesh and hormone pregnancy tests cases. He has received expenses and fees for his media work including occasional payments from BBC Radio 4 Inside Health and The Spectator. He receives expenses for teaching EBM and is also paid for his GP work in NHS out of hours (contract Oxford Health NHS Foundation Trust). He has also received income from the publication of a series of toolkit books and for appraising treatment recommendations in non NHS settings. He is Director of CEBM and an NIHR Senior Investigator and an advisor to Collateral Global.

## Funding

This work was funded by the NIHR School for Primary Care Research [project no. 438, round FR17].

## Transparency

The lead author (EM) affirms that the manuscript is an honest, accurate, and transparent account of the study being reported; that no important aspects of the study have been omitted; and that any discrepancies from the study as planned (and, if relevant, registered) have been explained.

## Patient or user group involvement statement

Patient representatives from patient advocacy group, Sling the Mesh, were involved (via conference call) with the protocol, research question, objectives, and outcome measures of the study to ensure they were patient-specific, appropriate and relevant. This study uses routinely collected data, so no recruitment was required.

## Data sharing

To guarantee the confidentiality of personal and health information only the authors have had access to the data during the study in accordance with the relevant licence agreements. CPRD linked data were provided under a licence that does not permit sharing.

## Supplementary material

eFigure 1: Flow diagram of patients for inclusion in this study

eTable 1: Code lists for outcome and exposure variables.

eTable 2: Unadjusted and adjusted^a^ hazard ratios for antibiotic prescriptions and opioid prescriptions, comparing mesh surgery with no surgery, in 162,578 women with stress urinary incontinence and 82,061 women with pelvic organ prolapse

eTable 3: Unadjusted and adjusted^a^ hazard ratios/incidence rate ratios for outcomes, comparing mesh surgery with no surgery, in 161,356 women with stress urinary incontinence and 81,203 women with pelvic organ prolapse, including only women with surgery at/after study entry.

## Notes

### Author Declarations

The protocol for this research was approved by the Independent Scientific Advisory Committee (ISAC) of the Medicines and Healthcare Products Regulatory Agency (protocol number 19_167), and the approved protocol is available on request. Ethical approval for observational research using the CPRD with approval from ISAC has been granted by a National Research Ethics Service Committee (Trent Multi Research Ethics Committee, REC reference number 05/MRE04/87).

### Summary of Updates

What this study adds - In this cohort of 162,687 women with SUI and 82,123 women with POP, rates of depression, anxiety, self-harm, sexual dysfunction and opioid prescriptions were higher in women with mesh-surgery who had no previous history of these outcomes. Lower rates of depression, anxiety and self-harm were seen in women with a history of these outcomes and SUI or POP mesh surgery. Opioid prescribing was lower in women with a previous history of prescriptions and POP mesh surgery. Careful consideration of the benefits and risk of mesh surgery for women with SUI or POP on an individual basis is required.

