## Supplementary material for "The long-term impact of vaginal surgical mesh devices in UK primary care: a cohort study in the CPRD": eFigure 1, eTable 2, eTable 3

eFigure 1: Flow diagram of patients for inclusion in this study

eTable 1: Code lists for outcome and exposure variables. **Please refer to separate document**

eTable 2: Unadjusted and adjusted^a^ hazard ratios for antibiotic prescriptions and opioid prescriptions, comparing mesh surgery with no surgery, in 167,210 women with stress urinary incontinence and 82,123 women with pelvic organ prolapse

eTable 3: Unadjusted and adjusteda hazard ratios/incidence rate ratios for outcomes, comparing mesh surgery with no surgery, in 161,356 women with stress urinary incontinence and 81,203 women with pelvic organ prolapse, including only women with surgery at/after study entry.

eFigure 1: Flow diagram of patients for inclusion in this study

1.	Herrett E, Gallagher AM, Bhaskaran K, et al. Data Resource Profile: Clinical Practice Research Datalink (CPRD). *Int J Epidemiol*. June 2015:dyv098-. doi:10.1093/ije/dyv098

284,462 women with acceptable data1, codes for SUI and/or POP, and record linkage in CPRD Set 17

63,853 patients with no eligible follow-up data

220,609 women with codes for SUI and/or POP (24,266 women have both)

82,123 women with POP codes and eligible follow-up data

162,752 women with SUI codes

65 women with removal of mesh before study entry

162,687 women with SUI codes and eligible follow up data

eTable1: Code lists for outcome and exposure variables. **Please refer to separate document**

eTable2: Unadjusted and adjusted^a^ hazard ratios for antibiotic prescriptions and opioid prescriptions, comparing mesh surgery with no surgery, in 162,578 women with stress urinary incontinence and 82,061 women with pelvic organ prolapse

| Ratios are for mesh surgery, reference group of no surgery | **Antibiotic prescriptions** | | | | **Opioid prescriptions** | | | |
| --- | --- | --- | --- | --- | --- | --- | --- | --- |
|  | Unadjusted | | Adjusted ^a^ | | Unadjusted | | Adjusted ^a^ | |
|  | HR | p-value | HR | p-value | HR | p-value | HR | p-value |
|  | (95% CI) |  | (95% CI) |  | (95% CI) |  | (95% CI) |  |
| **Stress urinary incontinence** |  |  |  |  |  |  |  |  |
| All women | 1.26 ^b^  (1.22,1.31) | <0.001 | 1.33 (1.28,1.38) | <0.001 | 1.12 (1.08,1.18) | <0.001 | 1.15 (1.10,1.20) | <0.001 |
| No previous history of the outcome | 1.37 (1.22,1.53) | <0.001 | 1.45 (1.30,1.63) | <0.001 | 1.14 (1.05,1.23) | <0.01 | 1.17 (1.08,1.27) | <0.001 |
| Previous history of the outcome | 1.25 (1.20,1.30) | <0.001 | 1.31 (1.26,1.37) | <0.001 | 1.06 (1.00,1.12) | 0.04 | 1.13 (1.07,1.20) | <0.001 |
| **Pelvic organ prolapse** |  |  |  |  |  |  |  |  |
| All women | 1.21 (1.15,1.29) | <0.001 | 1.22 (1.15,1.29) | <0.001 | 1.20 (1.12,1.29) | <0.001 | 1.13 (1.06,1.22) | <0.001 |
| No previous history of the outcome | 1.10 (0.93,1.30) | 0.29 | 1.12 (0.94,1.33) | 0.19 | 1.12 (0.98,1.29) | 0.10 | 1.10 (0.96,1.26) | 0.17 |
| Previous history of the outcome | 1.24 (1.17,1.32) | <0.001 | 1.23 (1.16,1.31) | <0.001 | 1.12 (1.03,1.21) | <0.01 | 1.15 (1.06,1.25) | <0.001 |

a Rows for all women were adjusted for age (5 year groups), body mass index (<18.5, 18.5-24.9, 25-29.9, ≥30 kg/m2), deprivation quintiles, ethnicity (“white”, “Asian”, “black”, “other”/”mixed”, missing), Strategic Health Authority region, history of the outcome. Rows stratified by previous history of the outcome are only presented for models where the interaction between exposure and previous history of the outcome were significant (p≤0.05). These rows were adjusted for all the above variables except for previous history of the outcome.

^b^ HR shows that women with stress urinary incontinence, who had mesh surgery, were at 1.26 times (95% CI 1.22 to 1.31) the risk of a first antibiotic prescription compared with those who did not have mesh surgery, this association was statistically significant, p<0.001.

**eTable 3: Unadjusted and adjusted**^a^ **hazard ratios/incidence rate ratios for outcomes, comparing mesh surgery with no surgery,** **in 161,356 women with stress urinary incontinence and 81,203 women with pelvic organ prolapse, including only women with surgery at/after study entry.**

| Ratios are for mesh surgery, reference group of no surgery | **Depression anxiety and self-harm** | | | | **Sexual dysfunction** | | | | **Antibiotic prescriptions** | | | | **Opioid prescriptions** | | | |
| --- | --- | --- | --- | --- | --- | --- | --- | --- | --- | --- | --- | --- | --- | --- | --- | --- |
|  | Unadjusted | | Adjusted ^a^ | | Unadjusted | | Adjusted ^a^ | | Unadjusted | Adjusted ^a^ | | | Unadjusted | | Adjusted ^a^ | |
|  | HR | p-value | HR | p-value | HR | p-value | HR | p-value | IRR | p-value | IRR | p-value | IRR | p-value | IRR | p-value |
|  | (95% CI) |  | (95% CI) |  | (95% CI) |  | (95% CI) |  | (95% CI) |  | (95% CI) |  | (95% CI) |  | (95% CI) |  |
| **Stress urinary incontinence** |  |  |  |  |  |  |  |  |  |  |  |  |  |  |  |  |
| All women | 1.54 (1.48,1.61) | <0.001 | 0.72 (0.69,0.75) | <0.001 | 1.85 (1.49,2.28) | <0.001 | 1.49 (1.20,1.85) | <0.001 | 1.15 (1.12,1.18) | <0.001 | 1.14 (1.11,1.17) | <0.001 | 1.20 (1.17,1.24) | <0.001 | 0.91 (0.88,0.94) | <0.001 |
| No previous history of the outcome | 2.18 (1.95,2.44) | <0.001 | 2.43 (2.17,2.73) | <0.001 | 1.97 (1.56,2.49) | <0.001 | 1.90 (1.50,2.40) | <0.001 | 1.45 (1.24,1.70) | <0.001 | 1.56 (1.34,1.83) | <0.001 | 1.26 (1.11,1.42) | <0.001 | 1.42 (1.25,1.60) | <0.001 |
| Previous history of the outcome | 0.65 (0.62,0.68) | <0.001 | 0.65 (0.62,0.68) | <0.001 | 0.59 (0.36,0.99) | 0.04 | 0.69 (0.41,1.15) | 0.15 | 1.13 (1.10,1.16) | <0.001 | 1.15 (1.12,1.18) | <0.001 | 1.15 (1.12,1.18) | <0.001 | 1.01 (0.97,1.04) | 0.68 |
| **Pelvic organ prolapse** |  |  |  |  |  |  |  |  |  |  |  |  |  |  |  |  |
| All women | 1.60 (1.45,1.77) | <0.001 | 0.70 (0.64,0.78) | <0.001 | 1.91 (1.19,3.06) | 0.007 | 1.49 (0.93,2.39) | 0.10 | 1.10 (1.04,1.17) | <0.001 | 1.04 (0.99,1.10) | 0.14 | 0.99 (0.94,1.04) | 0.78 | 0.72 (0.67,0.77) | <0.001 |
| No previous history of the outcome | 2.21 (1.74,2.82) | <0.001 | 2.21 (1.74,2.82) | <0.001 | 2.20 (1.33,3.62) | 0.002 | 2.23 (1.35,3.68) | 0.002 |  |  |  |  | 1.18 (0.91,1.53) | 0.21 | 1.21 (0.94,1.57) | 0.15 |
| Previous history of the outcome | 0.62 (0.56,0.69) | <0.001 | 0.63 (0.57,0.70) | <0.001 | 0.40 (0.10,1.61) | 0.20 | 0.38 (0.09,1.55) | 0.18 |  |  |  |  | 0.99 (0.94,1.03) | 0.60 | 0.87 (0.81,0.92) | <0.001 |

^a^ Rows for all women were adjusted for age (5 year groups), body mass index (<18.5, 18.5-24.9, 25-29.9, ≥30 kg/m2), deprivation quintiles, ethnicity (“white”, “Asian”, “black”, “other”/”mixed”, missing), Strategic Health Authority region, history of the outcome. Rows stratified by previous history of the outcome are only presented for models where the interaction between exposure and previous history of the outcome were significant (p≤0.05). These rows were adjusted for all the above variables except for previous history of the outcome.
