## Supplementary material for "The long-term impact of vaginal surgical mesh devices in UK primary care: a cohort study in the CPRD": eTable 1

eTable 1: Codes lists for Outcomes and exposures

Codes are those used within the CPRD dataset^1^

1.	Herrett E, Gallagher AM, Bhaskaran K, et al. Data Resource Profile: Clinical Practice Research Datalink (CPRD). *Int J Epidemiol*. June 2015:dyv098-. doi:10.1093/ije/dyv098

Depression, anxiety and self-harm medical codes

| Medical code | Read code | Read term |
| --- | --- | --- |
| depression | |  |
| 324 | E2B..00 | Depressive disorder NEC |
| 543 | Eu32z11 | [X]Depression NOS |
| 595 | E112.14 | Endogenous depression |
| 655 | E200300 | Anxiety with depression |
| 1055 | E135.00 | Agitated depression |
| 1131 | E204.00 | Neurotic depression reactive type |
| 1531 | Eu31.11 | [X]Manic-depressive illness |
| 1533 | E290.00 | Brief depressive reaction |
| 1908 | 2257 | O/E - depressed |
| 1996 | 1B17.00 | Depressed |
| 2639 | E204.11 | Postnatal depression |
| 2716 | 1465 | H/O: depression |
| 2923 | 62T1.00 | Puerperal depression |
| 2970 | Eu32z00 | [X]Depressive episode, unspecified |
| 2972 | E2B0.00 | Postviral depression |
| 3291 | Eu32z12 | [X]Depressive disorder NOS |
| 3292 | Eu33.00 | [X]Recurrent depressive disorder |
| 4323 | E2B1.00 | Chronic depression |
| 4639 | Eu32.00 | [X]Depressive episode |
| 4824 | 1B17.11 | C/O - feeling depressed |
| 4979 | Eu53012 | [X]Postpartum depression NOS |
| 5879 | E112.11 | Agitated depression |
| 5987 | Eu32z14 | [X] Reactive depression NOS |
| 6482 | E113700 | Recurrent depression |
| 6546 | E112.12 | Endogenous depression first episode |
| 6710 | Eu31.12 | [X]Manic-depressive psychosis |
| 6854 | Eu32y00 | [X]Other depressive episodes |
| 6932 | E113.11 | Endogenous depression - recurrent |
| 6950 | E112.13 | Endogenous depression first episode |
| 7011 | E112z00 | Single major depressive episode NOS |
| 7604 | Eu32.13 | [X]Single episode of reactive depression |
| 7737 | Eu34113 | [X]Neurotic depression |
| 7749 | Eu41211 | [X]Mild anxiety depression |
| 7953 | Eu34100 | [X]Dysthymia |
| 8478 | E130.00 | Reactive depressive psychosis |
| 8584 | Eu34111 | [X]Depressive neurosis |
| 8826 | Eu33.15 | [X]SAD - Seasonal affective disorder |
| 8851 | Eu33.11 | [X]Recurrent episodes of depressive reaction |
| 8902 | Eu33.13 | [X]Recurrent episodes of reactive depression |
| 9055 | Eu32.11 | [X]Single episode of depressive reaction |
| 9183 | E11z200 | Masked depression |
| 9211 | Eu32100 | [X]Moderate depressive episode |
| 9667 | Eu32200 | [X]Severe depressive episode without psychotic symptoms |
| 9796 | 1B1U.00 | Symptoms of depression |
| 10015 | 1BT..00 | Depressed mood |
| 10290 | Eu34112 | [X]Depressive personality disorder |
| 10438 | 1B1U.11 | Depressive symptoms |
| 10455 | E211200 | Depressive personality disorder |
| 10610 | E112.00 | Single major depressive episode |
| 10667 | Eu32400 | [X]Mild depression |
| 10720 | Eu32y11 | [X]Atypical depression |
| 10825 | E118.00 | Seasonal affective disorder |
| 11055 | Eu25100 | [X]Schizoaffective disorder, depressive type |
| 11252 | Eu33212 | [X]Major depression, recurrent without psychotic symptoms |
| 11329 | Eu33211 | [X]Endogenous depression without psychotic symptoms |
| 11717 | Eu32000 | [X]Mild depressive episode |
| 11913 | Eu41200 | [X]Mixed anxiety and depressive disorder |
| 12099 | Eu32300 | [X]Severe depressive episode with psychotic symptoms |
| 12122 | 9H91.00 | Depression medication review |
| 12399 | 9H90.00 | Depression annual review |
| 13307 | Eu53011 | [X]Postnatal depression NOS |
| 14709 | E113200 | Recurrent major depressive episodes, moderate |
| 15099 | E113.00 | Recurrent major depressive episode |
| 15155 | E112200 | Single major depressive episode, moderate |
| 15219 | E112300 | Single major depressive episode, severe, without psychosis |
| 15220 | Eu34114 | [X]Persistant anxiety depression |
| 16506 | E112100 | Single major depressive episode, mild |
| 16632 | E291.00 | Prolonged depressive reaction |
| 16861 | Eu33315 | [X]Recurrent severe episodes of psychotic depression |
| 18510 | Eu32.12 | [X]Single episode of psychogenic depression |
| 19054 | Eu3y111 | [X]Recurrent brief depressive episodes |
| 19439 | 212S.00 | Depression resolved |
| 19696 | Eu33.12 | [X]Recurrent episodes of psychogenic depression |
| 20785 | Eu20400 | [X]Post-schizophrenic depression |
| 21887 | E002100 | Senile dementia with depression |
| 22116 | Eu33400 | [X]Recurrent depressive disorder, currently in remission |
| 22806 | Eu32212 | [X]Single episode major depression w'out psychotic symptoms |
| 23731 | Eu33311 | [X]Endogenous depression with psychotic symptoms |
| 24112 | Eu32313 | [X]Single episode of psychotic depression |
| 24117 | Eu32311 | [X]Single episode of major depression and psychotic symptoms |
| 24171 | E113400 | Recurrent major depressive episodes, severe, with psychosis |
| 25563 | E113z00 | Recurrent major depressive episode NOS |
| 25697 | E113300 | Recurrent major depressive episodes, severe, no psychosis |
| 27491 | E11y200 | Atypical depressive disorder |
| 27677 | E001300 | Presenile dementia with depression |
| 28248 | Eu32z13 | [X]Prolonged single episode of reactive depression |
| 28756 | Eu33.14 | [X]Seasonal depressive disorder |
| 28863 | Eu32314 | [X]Single episode of reactive depressive psychosis |
| 28970 | 9hC0.00 | Excepted from depression quality indicators: Patient unsuita |
| 29342 | E113100 | Recurrent major depressive episodes, mild |
| 29520 | Eu33100 | [X]Recurrent depressive disorder, current episode moderate |
| 29784 | Eu33000 | [X]Recurrent depressive disorder, current episode mild |
| 30405 | 9H92.00 | Depression interim review |
| 30483 | 8CAa.00 | Patient given advice about management of depression |
| 31757 | Eu33314 | [X]Recurr severe episodes/psychogenic depressive psychosis |
| 32159 | E112400 | Single major depressive episode, severe, with psychosis |
| 32841 | 8HHq.00 | Referral for guided self-help for depression |
| 32845 | Eu92000 | [X]Depressive conduct disorder |
| 32941 | Eu33313 | [X]Recurr severe episodes/major depression+psychotic symptom |
| 33469 | Eu33200 | [X]Recurr depress disorder cur epi severe without psyc sympt |
| 34390 | E112000 | Single major depressive episode, unspecified |
| 35274 | Eu25111 | [X]Schizoaffective psychosis, depressive type |
| 35671 | E113000 | Recurrent major depressive episodes, unspecified |
| 36246 | E290z00 | Brief depressive reaction NOS |
| 36616 | Eu33z11 | [X]Monopolar depression NOS |
| 37764 | Eu33316 | [X]Recurrent severe episodes/reactive depressive psychosis |
| 41022 | Eu25112 | [X]Schizophreniform psychosis, depressive type |
| 41989 | Eu32211 | [X]Single episode agitated depressn w'out psychotic symptoms |
| 42931 | 9HA0.00 | On depression register |
| 43239 | 9hC1.00 | Excepted from depression quality indicators: Informed dissen |
| 43292 | E004300 | Arteriosclerotic dementia with depression |
| 43324 | E112500 | Single major depressive episode, partial or unspec remission |
| 44300 | Eu33z00 | [X]Recurrent depressive disorder, unspecified |
| 44936 | 9HA1.00 | Removed from depression register |
| 47731 | Eu33y00 | [X]Other recurrent depressive disorders |
| 52678 | Eu32312 | [X]Single episode of psychogenic depressive psychosis |
| 55384 | E113600 | Recurrent major depressive episodes, in full remission |
| 56273 | E113500 | Recurrent major depressive episodes,partial/unspec remission |
| 56609 | Eu32y12 | [X]Single episode of masked depression NOS |
| 57409 | E112600 | Single major depressive episode, in full remission |
| 59386 | Eu32213 | [X]Single episode vital depression w'out psychotic symptoms |
| 66153 | Eu31.13 | [X]Manic-depressive reaction |
| 73991 | Eu33214 | [X]Vital depression, recurrent without psychotic symptoms |
| 74050 | 3004A |  |
| 74079 | 3004CA |  |
| 74155 | 3004ER |  |
| 74293 | 3000E |  |
| 74296 | 2962R |  |
| 74352 | 3004AM |  |
| 74470 | 2962A |  |
| 74624 | 3004 |  |
| 74775 | 2962EN |  |
| 75087 | 2962B |  |
| 75612 | 3004C |  |
| 76768 | 2960AD |  |
| 77005 | 3091N |  |
| 77229 | 2962AF |  |
| 78918 | 3004AB |  |
| 79163 | 7902DC |  |
| 79246 | 3004B |  |
| 79553 | 3091PL |  |
| 80858 | 3004E |  |
| 83126 | 3091PN |  |
| 83254 | 3004PP |  |
| 84210 | 3091PF |  |
| 84554 | 3004M |  |
| 85574 | 2960AC |  |
| 98252 | Eu32600 | [X]Major depression, moderately severe |
| 98346 | Eu32500 | [X]Major depression, mild |
| 98414 | Eu32700 | [X]Major depression, severe without psychotic symptoms |
| 98417 | Eu32800 | [X]Major depression, severe with psychotic symptoms |
| 103677 | Eu32B00 | [X]Antenatal depression |
| **anxiety** |  |  |
| Medical code | Read code | Read term |
| 191 | E278100 | Tension headache |
| 276 | E28..00 | Acute reaction to stress |
| 636 | E200.00 | Anxiety states |
| 655 | E200300 | Anxiety with depression |
| 966 | E207.00 | Hypochondriasis |
| 1510 | E202B00 | Cancer phobia |
| 1723 | E202800 | Claustrophobia |
| 1758 | E200400 | Chronic anxiety |
| 1907 | E202.00 | Phobic disorders |
| 2030 | E203100 | Obsessional neurosis |
| 2076 | E21..00 | Personality disorders |
| 2188 | E201.00 | Hysteria |
| 2300 | E202000 | Phobia unspecified |
| 2366 | E202C00 | Dental phobia |
| 2571 | Eu40000 | [X]Agoraphobia |
| 2775 | E290000 | Grief reaction |
| 2826 | E29..00 | Adjustment reaction |
| 2871 | E264200 | Cyclical vomiting - psychogenic |
| 3076 | E202100 | Agoraphobia with panic attacks |
| 3208 | E203.00 | Obsessive-compulsive disorders |
| 3361 | E205.00 | Neurasthenia - nervous debility |
| 3438 | E201100 | Hysterical blindness |
| 3685 | E20y100 | Writer's cramp neurosis |
| 3869 | E264400 | Psychogenic dyspepsia |
| 4069 | E200100 | Panic disorder |
| 4167 | E202A00 | Fear of flying |
| 4171 | Eu43100 | [X]Post - traumatic stress disorder |
| 4199 | E26..00 | Physiological malfunction arising from mental factors |
| 4269 | E201700 | Hysterical amnesia |
| 4534 | E200z00 | Anxiety state NOS |
| 4634 | E200500 | Recurrent anxiety |
| 4659 | E200200 | Generalised anxiety disorder |
| 4775 | E201800 | Hysterical fugue |
| 4963 | E264000 | Psychogenic aerophagy |
| 5067 | E26z.00 | Psychosomatic disorder NOS |
| 5249 | E20..00 | Neurotic disorders |
| 5304 | Eu42.00 | [X]Obsessive - compulsive disorder |
| 5305 | E206.00 | Depersonalisation syndrome |
| 5385 | Eu41.00 | [X]Other anxiety disorders |
| 5652 | E210.00 | Paranoid personality disorder |
| 5678 | E203000 | Compulsive neurosis |
| 6071 | E202E00 | Fear of pregnancy |
| 6075 | E293000 | Adjustment reaction with aggression |
| 6221 | E292000 | Separation anxiety disorder |
| 6939 | E200000 | Anxiety state unspecified |
| 7537 | Eu45200 | [X]Hypochondriacal disorder |
| 7716 | E29y300 | Elective mutism due to an adjustment reaction |
| 8205 | Eu41000 | [X]Panic disorder [episodic paroxysmal anxiety] |
| 9125 | 8G94.00 | Anxiety management training |
| 9265 | Eu46100 | [X]Depersonalization - derealization syndrome |
| 9386 | Eu40.00 | [X]Phobic anxiety disorders |
| 9686 | E2...00 | Neurotic, personality and other nonpsychotic disorders |
| 9785 | Eu40200 | [X]Specific (isolated) phobias |
| 10001 | E26y000 | Bruxism (teeth grinding) |
| 10344 | Eu41100 | [X]Generalized anxiety disorder |
| 10390 | E202D00 | Fear of death |
| 10455 | E211200 | Depressive personality disorder |
| 11098 | Eu43.00 | [X]Reaction to severe stress, and adjustment disorders |
| 11336 | Eu43200 | [X]Adjustment disorders |
| 11602 | Eu40100 | [X]Social phobias |
| 11607 | Eu43000 | [X]Acute stress reaction |
| 11913 | Eu41200 | [X]Mixed anxiety and depressive disorder |
| 11940 | E280.00 | Acute panic state due to acute stress reaction |
| 12228 | E211100 | Hypomanic personality disorder |
| 12453 | Eu45500 | [X]Globus pharyngeus |
| 12508 | Eu40300 | [X]Needle phobia |
| 12707 | E211300 | Cyclothymic personality disorder |
| 12838 | E202200 | Agoraphobia without mention of panic attacks |
| 14729 | E202z00 | Phobic disorder NOS |
| 14780 | E20z.00 | Neurotic disorder NOS |
| 14979 | E211.00 | Affective personality disorder |
| 15034 | E262z00 | Psychogenic cardiovascular symptom NOS |
| 15035 | E260z00 | Psychogenic musculoskeletal symptoms NOS |
| 15224 | E263z00 | Psychogenic skin symptoms NOS |
| 15284 | E262200 | Neurocirculatory asthenia |
| 15292 | E262000 | Cardiac neurosis |
| 15321 | E20y000 | Somatization disorder |
| 15371 | E264300 | Psychogenic diarrhoea |
| 15431 | E201600 | Other conversion disorder |
| 15483 | E261100 | Psychogenic cough |
| 15551 | E282.00 | Acute stupor state due to acute stress reaction |
| 15566 | E203z00 | Obsessive-compulsive disorder NOS |
| 15665 | E292z00 | Adjustment reaction with disturbance of other emotion NOS |
| 15939 | E264500 | Psychogenic constipation |
| 15959 | E263000 | Psychogenic pruritus |
| 16178 | E211000 | Unspecified affective personality disorder |
| 16199 | E202300 | Social phobia, fear of eating in public |
| 16415 | E293.00 | Adjustment reaction with predominant disturbance of conduct |
| 16484 | E201500 | Hysterical seizures |
| 16561 | Eu46000 | [X]Neurasthenia |
| 18049 | Eu45.00 | [X]Somatoform disorders |
| 18399 | Eu42200 | [X]Mixed obsessional thoughts and acts |
| 18603 | E202500 | Social phobia, fear of public washing |
| 19921 | E29y500 | Other adjustment reaction with withdrawal |
| 20053 | E261300 | Psychogenic hyperventilation |
| 20109 | E265100 | Psychogenic vaginismus |
| 20245 | E283000 | Acute situational disturbance |
| 20634 | Eu42000 | [X]Predominantly obsessional thoughts or ruminations |
| 21753 | Eu43y00 | [X]Other reactions to severe stress |
| 22019 | Eu42100 | [X]Predominantly compulsive acts [obsessional rituals] |
| 22136 | Eu44.00 | [X]Dissociative [conversion] disorders |
| 22721 | Eu42z00 | [X]Obsessive-compulsive disorder, unspecified |
| 23327 | E292100 | Adolescent emancipation disorder |
| 23354 | E201z00 | Hysteria NOS |
| 23413 | E261400 | Psychogenic yawning |
| 23462 | E29yz00 | Other adjustment reactions NOS |
| 23490 | E201A00 | Dissociative reaction unspecified |
| 23598 | E201300 | Hysterical tremor |
| 23808 | Eu4..00 | [X]Neurotic, stress - related and somoform disorders |
| 23838 | Eu41z00 | [X]Anxiety disorder, unspecified |
| 23869 | E284.00 | Stress reaction causing mixed disturbance of emotion/conduct |
| 24066 | Eu41y00 | [X]Other specified anxiety disorders |
| 24212 | E292.00 | Adjustment reaction, predominant disturbance other emotions |
| 24439 | Eu45000 | [X]Somatization disorder |
| 24525 | E201C00 | Phantom pregnancy |
| 24847 | E283100 | Acute posttrauma stress state |
| 26138 | E28z.00 | Acute stress reaction NOS |
| 27390 | E29y400 | Adjustment reaction due to hospitalisation |
| 27633 | Eu44600 | [X]Dissociative anaesthesia and sensory loss |
| 27685 | Eu40y00 | [X]Other phobic anxiety disorders |
| 28090 | Eu46.00 | [X]Other neurotic disorders |
| 28106 | E202600 | Acrophobia |
| 28938 | E202700 | Animal phobia |
| 29322 | E201B00 | Compensation neurosis |
| 29448 | E262.00 | Psychogenic cardiovascular symptoms |
| 29461 | E263.00 | Psychogenic skin symptoms |
| 29707 | E283z00 | Other acute stress reaction NOS |
| 30179 | Eu45400 | [X]Persistent somatoform pain disorder |
| 30961 | E262300 | Psychogenic cardiovascular disorder |
| 31422 | E264.00 | Psychogenic gastrointestinal tract symptoms |
| 31515 | Eu43z00 | [X]Reaction to severe stress, unspecified |
| 31672 | E202900 | Fear of crowds |
| 31957 | E202400 | Social phobia, fear of public speaking |
| 32034 | E261000 | Psychogenic air hunger |
| 32387 | E29y100 | Other post-traumatic stress disorder |
| 34064 | Eu40z00 | [X]Phobic anxiety disorder, unspecified |
| 34664 | E261z00 | Psychogenic respiratory symptom NOS |
| 34696 | E201400 | Hysterical paralysis |
| 34978 | Eu44400 | [X]Dissociative motor disorders |
| 35632 | E29y200 | Adjustment reaction with physical symptoms |
| 35914 | E293100 | Adjustment reaction with antisocial behaviour |
| 37669 | E29z.00 | Adjustment reaction NOS |
| 38134 | E261.00 | Psychogenic respiratory symptoms |
| 38640 | E283.00 | Other acute stress reactions |
| 38809 | Eu42y00 | [X]Other obsessive-compulsive disorders |
| 39518 | E20y200 | Other occupational neurosis |
| 39747 | Eu44300 | [X]Trance and possession disorders |
| 39826 | Eu44100 | [X]Dissociative fugue |
| 40311 | E278.00 | Psychalgia |
| 40994 | Eu44000 | [X]Dissociative amnesia |
| 41038 | Eu45300 | [X]Somatoform autonomic dysfunction |
| 41455 | E29y.00 | Other adjustment reactions |
| 41572 | E201000 | Hysteria unspecified |
| 41615 | E261500 | Psychogenic aphonia |
| 42000 | E20y.00 | Other neurotic disorders |
| 42737 | E281.00 | Acute fugue state due to acute stress reaction |
| 43050 | E20yz00 | Other neurotic disorder NOS |
| 43302 | E201900 | Multiple personality |
| 44212 | E260.00 | Psychogenic musculoskeletal symptoms |
| 44321 | Eu41300 | [X]Other mixed anxiety disorders |
| 44331 | Eu46y00 | [X]Other specified neurotic disorders |
| 44547 | E265200 | Psychogenic dysmenorrhea |
| 44739 | E201200 | Hysterical deafness |
| 45205 | E278200 | Psychogenic backache |
| 45603 | E294.00 | Adjustment reaction with disturbance emotion and conduct |
| 47809 | E261200 | Psychogenic hiccough |
| 48561 | E260000 | Psychogenic paralysis |
| 48588 | E292y00 | Adjustment reaction with mixed disturbance of emotion |
| 48671 | Eu45z00 | [X]Somatoform disorder, unspecified |
| 48906 | Eu44z00 | [X]Dissociative [conversion] disorder, unspecified |
| 49628 | Eu46z00 | [X]Neurotic disorder, unspecified |
| 51497 | E211z00 | Affective personality disorder NOS |
| 53362 | E29y000 | Concentration camp syndrome |
| 53766 | E278000 | Psychogenic pain unspecified |
| 54373 | E278z00 | Psychalgia NOS |
| 54658 | E292200 | Early adult emancipation disorder |
| 55781 | E265300 | Psychogenic dysuria |
| 56141 | Eu44200 | [X]Dissociative stupor |
| 56800 | E260100 | Psychogenic torticollis |
| 56924 | E292400 | Adjustment reaction with anxious mood |
| 56966 | Eu44500 | [X]Dissociative convulsions |
| 57877 | Eu45100 | [X]Undifferentiated somatoform disorder |
| 58013 | E292500 | Culture shock |
| 62002 | Eu45y00 | [X]Other somatoform disorders |
| 62400 | E26y.00 | Other psychogenic malfunction |
| 64166 | Eu44y00 | [X]Other dissociative [conversion] disorders |
| 66398 | E293200 | Adjustment reaction with destructiveness |
| 67304 | E292311 | Specific academic or work inhibition |
| 68379 | E265.00 | Psychogenic genitourinary tract symptoms |
| 71437 | E264z00 | Psychogenic gastrointestinal tract symptom NOS |
| 72171 | E20y300 | Psychasthenic neurosis |
| 73547 | E265z00 | Psychogenic genitourinary tract symptom NOS |
| 88758 | Eu44700 | [X]Mixed dissociative [conversion] disorders |
| 89237 | E267.00 | Psychogenic symptom of special sense organ |
| 96391 | E26yz00 | Other psychogenic malfunction NOS |
| 99609 | ZS7C700 | Post-traumatic mutism |
| 104891 | E293z00 | Adjustment reaction with predominant disturbance conduct NOS |
| Self-harm | |  |
|  | CODEs | description |
|  | 14K1.00 | Intentional overdose of prescription only medication |
|  | SL...14 | Overdose of biological substance |
|  | SL...15 | Overdose of drug |
|  | SL90.00 | Antidepressant poisoning |
|  | SL90z00 | Anti-depressant poisoning NOS |
|  | SLHz.00 | Drug and medicament poisoning NOS |
|  | TK...00 | Suicide and selfinflicted injury |
|  | TK0..00 | Suicide + selfinflicted poisoning by solid/liquid substances |
|  | TK00.00 | Suicide + selfinflicted poisoning by analgesic/antipyretic |
|  | TK01.00 | Suicide + selfinflicted poisoning by barbiturates |
|  | TK01000 | Suicide and self inflicted injury by Amylobarbitone |
|  | TK01100 | Suicide and self inflicted injury by Barbitone |
|  | TK01400 | Suicide and self inflicted injury by Phenobarbitone |
|  | TK02.00 | Suicide + selfinflicted poisoning by oth sedatives/hypnotics |
|  | TK03.00 | Suicide + selfinflicted poisoning tranquilliser/psychotropic |
|  | TK04.00 | Suicide + selfinflicted poisoning by other drugs/medicines |
|  | TK05.00 | Suicide + selfinflicted poisoning by drug or medicine NOS |
|  | TK06.00 | Suicide + selfinflicted poisoning by agricultural chemical |
|  | TK07.00 | Suicide + selfinflicted poisoning by corrosive/caustic subst |
|  | TK0z.00 | Suicide + selfinflicted poisoning by solid/liquid subst NOS |
|  | TK1..00 | Suicide + selfinflicted poisoning by gases in domestic use |
|  | TK10.00 | Suicide + selfinflicted poisoning by gas via pipeline |
|  | TK...11 | Cause of overdose - deliberate |
|  | TK11.00 | Suicide + selfinflicted poisoning by liquified petrol gas |
|  | TK...12 | Injury - self-inflicted |
|  | TK...13 | Poisoning - self-inflicted |
|  | TK...14 | Suicide and self harm |
|  | TK...15 | Attempted suicide |
|  | TK...17 | Para-suicide |
|  | TK1y.00 | Suicide and selfinflicted poisoning by other utility gas |
|  | TK1z.00 | Suicide + selfinflicted poisoning by domestic gases NOS |
|  | TK2..00 | Suicide + selfinflicted poisoning by other gases and vapours |
|  | TK20.00 | Suicide + selfinflicted poisoning by motor veh exhaust gas |
|  | TK21.00 | Suicide and selfinflicted poisoning by other carbon monoxide |
|  | TK2z.00 | Suicide + selfinflicted poisoning by gases and vapours NOS |
|  | TK3..00 | Suicide + selfinflicted injury by hang/strangulate/suffocate |
|  | TK30.00 | Suicide and selfinflicted injury by hanging |
|  | TK31.00 | Suicide + selfinflicted injury by suffocation by plastic bag |
|  | TK3y.00 | Suicide + selfinflicted inj oth mean hang/strangle/suffocate |
|  | TK3z.00 | Suicide + selfinflicted inj by hang/strangle/suffocate NOS |
|  | TK4..00 | Suicide and selfinflicted injury by drowning |
|  | TK5..00 | Suicide and selfinflicted injury by firearms and explosives |
|  | TK51.00 | Suicide and selfinflicted injury by shotgun |
|  | TK52.00 | Suicide and selfinflicted injury by hunting rifle |
|  | TK54.00 | Suicide and selfinflicted injury by other firearm |
|  | TK5z.00 | Suicide and selfinflicted injury by firearms/explosives NOS |
|  | TK6..00 | Suicide and selfinflicted injury by cutting and stabbing |
|  | TK60.00 | Suicide and selfinflicted injury by cutting |
|  | TK60100 | Self inflicted lacerations to wrist |
|  | TK60111 | Slashed wrists self inflicted |
|  | TK61.00 | Suicide and selfinflicted injury by stabbing |
|  | TK6z.00 | Suicide and selfinflicted injury by cutting and stabbing NOS |
|  | TK7..00 | Suicide and selfinflicted injury by jumping from high place |
|  | TK70.00 | Suicide+selfinflicted injury-jump from residential premises |
|  | TK71.00 | Suicide+selfinflicted injury-jump from oth manmade structure |
|  | TK72.00 | Suicide+selfinflicted injury-jump from natural sites |
|  | TK7z.00 | Suicide+selfinflicted injury-jump from high place NOS |
|  | TKx..00 | Suicide and selfinflicted injury by other means |
|  | TKx0.00 | Suicide + selfinflicted injury-jump/lie before moving object |
|  | TKx0000 | Suicide + selfinflicted injury-jumping before moving object |
|  | TKx1.00 | Suicide and selfinflicted injury by burns or fire |
|  | TKx2.00 | Suicide and selfinflicted injury by scald |
|  | TKx3.00 | Suicide and selfinflicted injury by extremes of cold |
|  | TKx4.00 | Suicide and selfinflicted injury by electrocution |
|  | TKx5.00 | Suicide and selfinflicted injury by crashing motor vehicle |
|  | TKx6.00 | Suicide and selfinflicted injury by crashing of aircraft |
|  | TKx7.00 | Suicide and selfinflicted injury caustic subst; excl poison |
|  | TKxy.00 | Suicide and selfinflicted injury by other specified means |
|  | TKxz.00 | Suicide and selfinflicted injury by other means NOS |
|  | TKy..00 | Late effects of selfinflicted injury |
|  | TKz..00 | Suicide and selfinflicted injury NOS |
|  | U2...00 | [X]Intentional self-harm |
|  | U20..00 | [X]Intentional self poisoning/exposure to noxious substances |
|  | U200.00 | [X]Intent self poison/exposure to nonopioid analgesic |
|  | U200000 | [X]Int self poison/exposure to nonopioid analgesic at home |
|  | U200100 | [X]Intent self poison nonopioid analgesic at res institut |
|  | U200.11 | [X]Overdose - paracetamol |
|  | U200.12 | [X]Overdose - ibuprofen |
|  | U200.13 | [X]Overdose - aspirin |
|  | U200400 | [X]Intent self pois nonopioid analgesic in street/highway |
|  | U200500 | [X]Intent self pois nonopioid analgesic trade/service area |
|  | U200y00 | [X]Int self poison nonopioid analgesic other spec place |
|  | U200z00 | [X]Intent self poison nonopioid analgesic unspecif place |
|  | U201.00 | [X]Intent self poison/exposure to antiepileptic |
|  | U201000 | [X]Int self poison/exposure to antiepileptic at home |
|  | U20..11 | [X]Deliberate drug overdose / other poisoning |
|  | U201z00 | [X]Intent self poison antiepileptic unspecif place |
|  | U202.00 | [X]Intent self poison/exposure to sedative hypnotic |
|  | U202000 | [X]Int self poison/exposure to sedative hypnotic at home |
|  | U202.11 | [X]Overdose - sleeping tabs |
|  | U202.12 | [X]Overdose - diazepam |
|  | U202.13 | [X]Overdose - temazepam |
|  | U202.15 | [X]Overdose - nitrazepam |
|  | U202.16 | [X]Overdose - benzodiazepine |
|  | U202.17 | [X]Overdose - barbiturate |
|  | U202.18 | [X]Overdose - amobarbital |
|  | U202400 | [X]Intent self pois sedative hypnotic in street/highway |
|  | U202y00 | [X]Int self poison sedative hypnotic other spec place |
|  | U202z00 | [X]Intent self poison sedative hypnotic unspecif place |
|  | U204.00 | [X]Intent self poison/exposure to psychotropic drug |
|  | U204000 | [X]Int self poison/exposure to psychotropic drug at home |
|  | U204100 | [X]Intent self poison psychotropic drug at res institut |
|  | U204.11 | [X]Overdose - antidepressant |
|  | U204.12 | [X]Overdose - amitriptyline |
|  | U204.13 | [X]Overdose - SSRI |
|  | U204y00 | [X]Int self poison psychotropic drug other spec place |
|  | U204z00 | [X]Intent self poison psychotropic drug unspecif place |
|  | U205.00 | [X]Intent self poison/exposure to narcotic drug |
|  | U205000 | [X]Int self poison/exposure to narcotic drug at home |
|  | U205y00 | [X]Int self poison narcotic drug other spec place |
|  | U205z00 | [X]Intent self poison narcotic drug unspecif place |
|  | U206.00 | [X]Intent self poison/exposure to hallucinogen |
|  | U206400 | [X]Intent self pois hallucinogen in street/highway |
|  | U207.00 | [X]Intent self poison/exposure to oth autonomic drug |
|  | U207000 | [X]Int self poison/exposure to oth autonomic drug at home |
|  | U207z00 | [X]Intent self poison oth autonomic drug unspecif place |
|  | U208.00 | [X]Int self poison/exposure to other/unspec drug/medicament |
|  | U208000 | [X]Int self poison/exposure to oth/unsp drug/medicam home |
|  | U208400 | [X]Intent self pois oth/unsp drug/medic in street/highway |
|  | U208y00 | [X]Int self poison oth/unsp drug/medic other spec place |
|  | U208z00 | [X]Intent self poison oth/unsp drug/medic unspecif place |
|  | U209.00 | [X]Intent self poison/exposure to alcohol |
|  | U209y00 | [X]Int self poison alcohol other spec place |
|  | U209z00 | [X]Intent self poison alcohol unspecif place |
|  | U20A.00 | [X]Intentional self poison organ solvent;halogen hydrocarb |
|  | U20A000 | [X]Intent self pois organ solvent;halogen hydrocarb; home |
|  | U20A.11 | [X]Self poisoning from glue solvent |
|  | U20A400 | [X]Int self poison org solvent;halogen hydrocarb;in highway |
|  | U20Az00 | [X]Int self pois org solv;halogen hydrocarb; unspec place |
|  | U20B.00 | [X]Intent self poison/exposure to other gas/vapour |
|  | U20B000 | [X]Int self poison/exposure to other gas/vapour at home |
|  | U20B.11 | [X]Self carbon monoxide poisoning |
|  | U20B200 | [X]Int self poison other gas/vapour school/pub admin area |
|  | U20By00 | [X]Int self poison other gas/vapour other spec place |
|  | U20Bz00 | [X]Intent self poison other gas/vapour unspecif place |
|  | U20C.00 | [X]Intent self poison/exposure to pesticide |
|  | U20C000 | [X]Int self poison/exposure to pesticide at home |
|  | U20C.11 | [X]Self poisoning with weedkiller |
|  | U20C.12 | [X]Self poisoning with paraquat |
|  | U20Cy00 | [X]Int self poison pesticide other spec place |
|  | U20y.00 | [X]Intent self poison/exposure to unspecif chemical |
|  | U20y000 | [X]Int self poison/exposure to unspecif chemical at home |
|  | U20y200 | [X]Int self poison unspecif chemical school/pub admin area |
|  | U20yz00 | [X]Intent self poison unspecif chemical unspecif place |
|  | U21..00 | [X]Intent self harm by hanging strangulation / suffocation |
|  | U210.00 | [X]Intent self harm by hanging strangulat/suffocat occ home |
|  | U2...11 | [X]Self inflicted injury |
|  | U211.00 | [X]Intent self harm by hangng strangult/suffoct resid instit |
|  | U2...12 | [X]Injury - self-inflicted |
|  | U212.00 | [X]Inten slf harm hang strang/suffc sch oth ins/pub adm area |
|  | U2...13 | [X]Suicide |
|  | U2...14 | [X]Attempted suicide |
|  | U2...15 | [X]Para-suicide |
|  | U216.00 | [X]Intent self harm by hang strangl/suffc indust/constr area |
|  | U21y.00 | [X]Intent self harm by hangng strangul/suffoct oth spec plce |
|  | U21z.00 | [X]Intent self harm by hangng strangul/suffoct unspecif plce |
|  | U22..00 | [X]Intentional self harm by drowning and submersion |
|  | U220.00 | [X]Intent self harm by drowning/submersion occurrn at home |
|  | U221.00 | [X]Intent self harm by drowning/submersn occ resid instit'n |
|  | U22y.00 | [X]Intent self harm by drown/submersn occ oth specif place |
|  | U22z.00 | [X]Intent self harm by drown/submersn occ unspecified place |
|  | U24..00 | [X]Intent self harm by rifle shotgun/larger firearm disch |
|  | U241.00 | [X]Int self harm rifl s'gun/lrg frarm disch occ resid instit |
|  | U242.00 | [X]Int slf hrm rifl s'gun/lrg frarm dis sch/ins/pub adm area |
|  | U25..00 | [X]Intent self harm by other/unspecified firearm discharge |
|  | U250.00 | [X]Intent self harm oth/unspecif firearm disch occ at home |
|  | U26..00 | [X]Intentional self harm by explosive material |
|  | U27..00 | [X]Intentional self harm by smoke; fire and flames |
|  | U270.00 | [X]Intention self harm by smoke fire/flames occurrn at home |
|  | U274.00 | [X]Intent self harm by smoke fire/flame occ street/highway |
|  | U27z.00 | [X]Intent self harm by smoke fire/flames occ unspecif place |
|  | U28..00 | [X]Intentional self harm by steam hot vapours / hot objects |
|  | U280.00 | [X]Intent self harm by steam hot vapour/hot obj occ at home |
|  | U28z.00 | [X]Intent self harm by steam hot vapour/obj occ unspec place |
|  | U29..00 | [X]Intentional self harm by sharp object |
|  | U290.00 | [X]Intentional self harm by sharp object occurrence at home |
|  | U291.00 | [X]Intent self harm by sharp object occ resident instit'n |
|  | U294.00 | [X]Intention self harm by sharp object occ street/highway |
|  | U29y.00 | [X]Intention self harm by sharp object occ oth specif place |
|  | U29z.00 | [X]Intentional self harm by sharp object occ unspecif place |
|  | U2A..00 | [X]Intentional self harm by blunt object |
|  | U2A0.00 | [X]Intentional self harm by blunt object occurrence at home |
|  | U2A1.00 | [X]Intent self harm by blunt object occ resident instit'n |
|  | U2A3.00 | [X]Intent self harm by blunt object occ sports/athlet area |
|  | U2B..00 | [X]Intentional self harm by jumping from a high place |
|  | U2B0.00 | [X]Intent self harm by jumping from high place occ at home |
|  | U2B4.00 | [X]Intent self harm by jump from high place occ street/h'way |
|  | U2B6.00 | [X]Int self harm by jump from high place indust/constr area |
|  | U2By.00 | [X]Int self harm by jump from high place occ oth specif plce |
|  | U2Bz.00 | [X]Int self harm by jump from high place occ unspecif place |
|  | U2C..00 | [X]Intent self harm by jumping / lying before moving object |
|  | U2C1.00 | [X]Int self harm jump/lying befr mov obje occ resid instit'n |
|  | U2C4.00 | [X]Int self harm jump/lying befr mov obje occ street/highway |
|  | U2Cy.00 | [X]Int self harm jump/lying bef mov obje occ oth specif plce |
|  | U2D..00 | [X]Intentional self harm by crashing of motor vehicle |
|  | U2D0.00 | [X]Intent self harm by crash of motor vehicl occurrn at home |
|  | U2D4.00 | [X]Intent self harm by crash motor vehicl occ street/highway |
|  | U2D6.00 | [X]Intent self harm crash motor vehic occ indust/constr area |
|  | U2E..00 | [X]Self mutilation |
|  | U2y..00 | [X]Intentional self harm by other specified means |
|  | U2y0.00 | [X]Intentionl self harm by oth specif means occurrn at home |
|  | U2y1.00 | [X]Intent self harm by oth specif means occ resid instit'n |
|  | U2yz.00 | [X]Intent self harm by oth specif means occ unspecif place |
|  | U2z..00 | [X]Intentional self harm by unspecified means |
|  | U2z0.00 | [X]Intentional self harm by unspecif means occurrn at home |
|  | U2z2.00 | [X]Intent self harm by unspec mean occ sch/ins/pub adm area |
|  | U2zy.00 | [X]Intent self harm by unspecif means occ oth specif place |
|  | U2zz.00 | [X]Intent self harm by unspecif means occ at unspecif place |
|  | U30..11 | [X]Deliberate drug poisoning |
|  | U41..00 | [X]Hanging strangulation + suffocation undetermined intent |
|  | U44..00 | [X]Rifle shotgun+larger firearm discharge undetermin intent |
|  | U45..00 | [X]Other+unspecified firearm discharge undetermined intent |
|  | U4B..00 | [X]Falling jumping/pushed from high place undeterm intent |
|  | U4Bz.00 | [X]Fall jump/push frm high plce undt intnt occ unspecif plce |
|  | U72..00 | [X]Sequel intentn self-harm assault+event of undeterm intent |
|  | U720.00 | [X]Sequelae of intentional self-harm |
|  | ZRLfC12 | HoNOS item 2 - non-accidental self injury |
|  | ZRn3.00 | Suicide intent score subscale - attempt circumstances |
|  | ZX...00 | Self-harm |
|  | ZX1..00 | Self-injurious behaviour |
|  | ZX...11 | Self-damage |
|  | ZX11.00 | Biting self |
|  | ZX11.11 | Bites self |
|  | ZX1..12 | SIB - Self-injurious behaviour |
|  | ZX1..13 | Deliberate self-harm |
|  | ZX12.00 | Burning self |
|  | ZX13.00 | Cutting self |
|  | ZX13100 | Cutting own wrists |
|  | ZX13.11 | Cuts self |
|  | ZX15.00 | Drowning self |
|  | ZX18.00 | Hanging self |
|  | ZX19.00 | Hitting self |
|  | ZX19100 | Punching self |
|  | ZX19200 | Slapping self |
|  | ZX1B.00 | Jumping from height |
|  | ZX1B100 | Jumping from building |
|  | ZX1B200 | Jumping from bridge |
|  | ZX1B300 | Jumping from cliff |
|  | ZX1C.00 | Nipping self |
|  | ZX1E.00 | Pinching self |
|  | ZX1G.00 | Scratches self |
|  | ZX1H.00 | Self-asphyxiation |
|  | ZX1H100 | Self-strangulation |
|  | ZX1H200 | Self-suffocation |
|  | ZX1I.00 | Self-scalding |
|  | ZX1J.00 | Self-electrocution |
|  | ZX1K.00 | Self-incineration |
|  | ZX1K.11 | Setting fire to self |
|  | ZX1K.12 | Setting self alight |
|  | ZX1L.00 | Self-mutilation |
|  | ZX1L100 | Self-mutilation of hands |
|  | ZX1L200 | Self-mutilation of genitalia |
|  | ZX1L300 | Self-mutilation of penis |
|  | ZX1L600 | Self-mutilation of ears |
|  | ZX1LD00 | [X]Self mutilation |
|  | ZX1M.00 | Shooting self |
|  | ZX1N.00 | Stabbing self |
|  | ZX1Q.00 | Throwing self in front of train |
|  | ZX1Q.11 | Jumping under train |
|  | ZX1R.00 | Throwing self in front of vehicle |
|  | ZX1S.00 | Throwing self onto floor |

Depression, anxiety and self-harm product codes

| Product code | Product name | gemscriptcode |
| --- | --- | --- |
| depression | |  |
| 22 | Fluoxetine 20mg capsules | 69654020 |
| 49 | Amitriptyline 25mg tablets | 58950020 |
| 50 | Paroxetine 20mg tablets | 72932020 |
| 67 | Citalopram 20mg tablets | 79462020 |
| 74 | Dosulepin 75mg tablets | 62834020 |
| 83 | Amitriptyline 10mg tablets | 58949020 |
| 84 | Dosulepin 25mg capsules | 62833020 |
| 114 | Lofepramine 70mg tablets | 64034020 |
| 182 | Tryptizol 10mg/ml Injection (Merck Sharp & Dohme Ltd) | 55445020 |
| 252 | Prozac 20mg/5ml liquid (Eli Lilly and Company Ltd) | 69507020 |
| 301 | Venlafaxine 37.5mg tablets | 54686020 |
| 418 | Prozac 20mg capsules (Eli Lilly and Company Ltd) | 69506020 |
| 470 | Venlafaxine 75mg modified-release capsules | 85108020 |
| 476 | Citalopram 10mg tablets | 79463020 |
| 487 | Amitriptyline 25mg modified-release capsules | 60222020 |
| 488 | Sertraline 50mg tablets | 74050020 |
| 513 | Citalopram 40mg/ml oral drops sugar free | 77306020 |
| 527 | Paroxetine 10mg/5ml oral suspension sugar free | 72934020 |
| 595 | Amitriptyline 25mg / Perphenazine 2mg tablets | 68729020 |
| 603 | Escitalopram 10mg tablets | 78768020 |
| 623 | Efexor 37.5mg tablets (Wyeth Pharmaceuticals) | 48376020 |
| 648 | Cipralex 10mg tablets (Lundbeck Ltd) | 86222020 |
| 727 | Sertraline 100mg tablets | 74051020 |
| 742 | Mirtazapine 30mg tablets | 85247020 |
| 785 | Cipralex 5mg tablets (Lundbeck Ltd) | 87526020 |
| 815 | Cipramil 40mg/ml drops (Lundbeck Ltd) | 77312020 |
| 841 | Seroxat 20mg tablets (GlaxoSmithKline UK Ltd) | 72937020 |
| 1169 | Prothiaden 25mg capsules (Teofarma) | 55456020 |
| 1208 | Triptafen tablets (AMCo) | 52046020 |
| 1222 | Venlafaxine 75mg tablets | 54687020 |
| 1310 | Imipramine 10mg tablets | 59429020 |
| 1397 | Paroxetine 30mg tablets | 72933020 |
| 1453 | Triptafen m 2mg+10mg Tablet (Goldshield Pharmaceuticals Ltd) | 52047020 |
| 1474 | Efexor XL 75mg capsules (Pfizer Ltd) | 85163020 |
| 1575 | Seroxat 30mg tablets (GlaxoSmithKline UK Ltd) | 72938020 |
| 1612 | Lustral 50mg tablets (Pfizer Ltd) | 74046020 |
| 1712 | Cipramil 20mg tablets (Lundbeck Ltd) | 79420020 |
| 1730 | Trazodone 100mg capsules | 67152020 |
| 1809 | Imipramine 25mg tablets | 59430020 |
| 1888 | Amitriptyline 50mg tablets | 58951020 |
| 1940 | Dothapax 25 capsules (Ashbourne Pharmaceuticals Ltd) | 56698020 |
| 2039 | Trimipramine 25mg tablets | 67281020 |
| 2093 | Gamanil 70mg tablets (Merck Serono Ltd) | 55432020 |
| 2290 | Fluvoxamine 100mg tablets | 62070020 |
| 2320 | Prothiaden 75mg tablets (Teofarma) | 55457020 |
| 2356 | Reboxetine 4mg tablets | 84955020 |
| 2408 | Cipramil 40mg tablets (Lundbeck Ltd) | 79422020 |
| 2525 | Amitriptyline 75mg modified-release capsules | 60224020 |
| 2531 | Surmontil 50mg capsules (Sanofi) | 55418020 |
| 2532 | Surmontil 25mg tablets (Sanofi) | 55417020 |
| 2533 | TRIMIPRAMINE 50 MG TAB | 1787007 |
| 2548 | Fluoxetine 20mg/5ml oral solution | 69655020 |
| 2579 | Tofranil 10mg Tablet (Novartis Pharmaceuticals UK Ltd) | 55402020 |
| 2617 | Venlafaxine 50mg tablets | 54688020 |
| 2654 | Venlafaxine 150mg modified-release capsules | 85109020 |
| 2880 | Fluvoxamine 50mg tablets | 62069020 |
| 2883 | Moclobemide 150mg tablets | 72031020 |
| 2897 | Faverin 50mg tablets (Mylan) | 62073020 |
| 3083 | Mianserin 10mg tablets | 64729020 |
| 3183 | Nortriptyline 10mg tablets | 65153020 |
| 3194 | Clomipramine 10mg capsules | 61463020 |
| 3196 | Trimipramine 50mg capsules | 67282020 |
| 3349 | Nardil 15mg tablets (Kyowa Kirin Ltd) | 50620020 |
| 3353 | L-TRYPTOPHAN 500 MG CAP | 62007 |
| 3355 | Trazodone 50mg capsules | 67151020 |
| 3356 | Parstelin Tablet (GlaxoSmithKline Consumer Healthcare) | 51013020 |
| 3490 | Amitriptyline 10mg / Perphenazine 2mg tablets | 68728020 |
| 3554 | Doxepin 25mg capsules | 62851020 |
| 3601 | Seroxat 20mg/10ml liquid (GlaxoSmithKline UK Ltd) | 72939020 |
| 3657 | Anafranil 25mg capsules (Novartis Pharmaceuticals UK Ltd) | 55387020 |
| 3668 | IMIPRAMINE 100 MG TAB | 3312007 |
| 3670 | Clomipramine 25mg capsules | 61464020 |
| 3777 | Amitriptyline 10mg/5ml sugar free oral solution | 75331020 |
| 3783 | Tranylcypromine 10mg tablets | 67145020 |
| 3842 | Doxepin 10mg capsules | 62850020 |
| 3861 | Cipramil 10mg tablets (Lundbeck Ltd) | 79421020 |
| 3903 | Nortriptyline 25mg tablets | 65154020 |
| 3925 | Clomipramine 50mg capsules | 61465020 |
| 3951 | Fluanxol 1mg tablets (Lundbeck Ltd) | 49503020 |
| 3953 | Fluanxol 500microgram tablets (Lundbeck Ltd) | 49502020 |
| 3954 | OPTIMAX TAB | 4647007 |
| 3955 | Tranylcypromine with trifluoperazine Tablet | 67148020 |
| 4003 | Molipaxin 150mg tablets (Zentiva) | 54110020 |
| 4020 | Trazodone 150mg tablets | 67153020 |
| 4075 | Fluoxetine 60mg capsules | 69656020 |
| 4118 | Nortriptyline 10mg Capsule | 65157020 |
| 4194 | Molipaxin 100mg capsules (Zentiva) | 54109020 |
| 4218 | Lofepramine 70mg/5ml oral suspension sugar free | 64035020 |
| 4310 | Trimipramine 10mg tablets | 67280020 |
| 4321 | Phenelzine 15mg tablets | 65635020 |
| 4329 | Mianserin 20mg tablets | 64730020 |
| 4352 | Lustral 100mg tablets (Pfizer Ltd) | 74047020 |
| 4422 | Tryptophan 500mg tablets | 67310020 |
| 4682 | Amitriptyline 50mg modified-release capsules | 60223020 |
| 4690 | Amitriptyline 50mg/5ml oral solution sugar free | 75329020 |
| 4726 | Zispin 30mg tablets (Organon Laboratories Ltd) | 85251020 |
| 4770 | Citalopram 40mg tablets | 79464020 |
| 4874 | Molipaxin 50mg capsules (Zentiva) | 54108020 |
| 4907 | Prozac 60mg capsules (Eli Lilly and Company Ltd) | 69508020 |
| 5073 | Doxepin 50mg capsules | 62852020 |
| 5187 | Moclobemide 300mg tablets | 72032020 |
| 5611 | Optimax 500mg tablets (Intrapharm Laboratories Ltd) | 50868020 |
| 5710 | Efexor XL 150mg capsules (Pfizer Ltd) | 85164020 |
| 5832 | Manerix 300mg tablets (Meda Pharmaceuticals Ltd) | 71995020 |
| 6054 | Dosulepin 25mg/5ml oral solution sugar free | 62835020 |
| 6218 | Escitalopram 20mg tablets | 53823020 |
| 6255 | Mianserin 30mg tablets | 64731020 |
| 6274 | Efexor 50mg tablets (Wyeth Pharmaceuticals) | 48378020 |
| 6312 | Amitriptyline 25mg/5ml oral solution sugar free | 75330020 |
| 6360 | Cipralex 20mg tablets (Lundbeck Ltd) | 55602020 |
| 6405 | Escitalopram 5mg tablets | 87524020 |
| 6421 | Mirtazapine 15mg orodispersible tablets | 87476020 |
| 6442 | Trazodone 50mg/5ml oral solution sugar free | 67156020 |
| 6481 | Mirtazapine 45mg orodispersible tablets | 87478020 |
| 6488 | Mirtazapine 30mg orodispersible tablets | 86969020 |
| 6795 | Mirtazapine 15mg tablets | 88879020 |
| 6846 | Zispin SolTab 15mg orodispersible tablets (Merck Sharp & Dohme Ltd) | 87480020 |
| 6854 | Mirtazapine 45mg tablets | 88881020 |
| 6894 | Perphenazine 2mg with Amitriptyline 25mg tablet | 65593020 |
| 6895 | Duloxetine 60mg gastro-resistant capsules | 88847020 |
| 7059 | Doxepin 75mg capsules | 62857020 |
| 7122 | Duloxetine 30mg gastro-resistant capsules | 88845020 |
| 7328 | Sertraline 50mg/5ml oral suspension | 90497020 |
| 7468 | Bolvidon 10mg Tablet (Organon Laboratories Ltd) | 48430020 |
| 7515 | Anafranil 10mg capsules (Novartis Pharmaceuticals UK Ltd) | 55386020 |
| 7573 | IMIPRAMINE 25 MG CAP | 4844007 |
| 7677 | Allegron 10mg tablets (King Pharmaceuticals Ltd) | 55355020 |
| 7678 | Nortriptyline 25mg Capsule | 65158020 |
| 7693 | Anafranil 50mg capsules (Novartis Pharmaceuticals UK Ltd) | 55388020 |
| 7751 | Tryptizol 25mg Tablet (Merck Sharp & Dohme Ltd) | 55441020 |
| 7780 | Nortriptyline 10mg / Fluphenazine 500microgram tablets | 68989020 |
| 7784 | IMIPRAMINE 50 MG TAB | 4843007 |
| 7894 | Anafranil SR 75mg tablets (Novartis Pharmaceuticals UK Ltd) | 55396020 |
| 7910 | Tofranil 25mg tablets (Novartis Pharmaceuticals UK Ltd) | 55403020 |
| 8055 | Imipramine 25mg/5ml oral solution | 63537020 |
| 8144 | Bolvidon 20mg Tablet (Organon Laboratories Ltd) | 48431020 |
| 8174 | Molipaxin 50mg/5ml oral liquid (Sanofi) | 54779020 |
| 8332 | Tryptizol 50mg Tablet (Merck Sharp & Dohme Ltd) | 55442020 |
| 8585 | Bolvidon 30mg Tablet (Organon Laboratories Ltd) | 48432020 |
| 8640 | Allegron 25mg tablets (King Pharmaceuticals Ltd) | 55356020 |
| 8661 | Clomipramine 75mg modified-release tablets | 61473020 |
| 8719 | Anafranil 25mg/5ml syrup (Novartis Pharmaceuticals UK Ltd) | 55392020 |
| 8720 | Clomipramine 25mg/5ml oral solution | 61469020 |
| 8726 | Tryptizol 10mg Tablet (Merck Sharp & Dohme Ltd) | 55440020 |
| 8831 | Tryptizol mr 75mg Modified-release capsule (Merck Sharp & Dohme Ltd) | 55446020 |
| 8844 | Tryptophan with ascorbic acid and pyridoxine powder | 67314020 |
| 8845 | OPTIMAX 6 GM POW | 5487007 |
| 8878 | Tryptizol 10mg/5ml sugar free Oral solution (Merck Sharp and Dohme Ltd) | 55450020 |
| 8928 | Surmontil 10mg tablets (Sanofi) | 55416020 |
| 9182 | Efexor 75mg tablets (Wyeth Pharmaceuticals) | 48377020 |
| 9206 | Manerix 150mg tablets (Meda Pharmaceuticals Ltd) | 71994020 |
| 10083 | Zispin SolTab 30mg orodispersible tablets (Merck Sharp & Dohme Ltd) | 86971020 |
| 10413 | Sinequan 10mg capsules (Pfizer Ltd) | 55464020 |
| 10649 | IMIPRAMINE 75 MG TAB | 4845007 |
| 10787 | Parnate 10mg Tablet (Goldshield Pharmaceuticals Ltd) | 51010020 |
| 10948 | Dosulepin 75mg/5ml oral solution sugar free | 54727020 |
| 11956 | Norval 20mg Tablet (Bencard) | 50775020 |
| 11963 | Limbitrol 10 Capsule (Roche Products Ltd) | 54655020 |
| 12123 | Faverin 100mg tablets (Mylan) | 62074020 |
| 12125 | Sinequan 50mg capsules (Pfizer Ltd) | 55466020 |
| 12129 | Sinequan 25mg capsules (Pfizer Ltd) | 55465020 |
| 12192 | Norval 30mg Tablet (Bencard) | 50776020 |
| 12194 | NORTRIPTYLINE 10 MG ELI | 5750007 |
| 12207 | Isocarboxazid 10mg tablets | 63638020 |
| 12221 | Pacitron 500mg Tablet (Rorer Pharmaceuticals Ltd) | 50957020 |
| 12368 | Norval 10mg Tablet (Bencard) | 50774020 |
| 12503 | Marplan 10mg Tablet (Cambridge Laboratories Ltd) | 50286020 |
| 12710 | Trazodone 150mg modified-release tablets | 67157020 |
| 13237 | Venlafaxine 37.5mg/5ml oral suspension | 89954020 |
| 13621 | Molipaxin CR 150mg tablets (Aventis Pharma) | 54780020 |
| 14519 | Sinequan 75mg capsules (Pfizer Ltd) | 55469020 |
| 14521 | SINEQUAN 15 MG TAB | 1972007 |
| 14534 | Limbitrol 5 Capsule (Roche Products Ltd) | 50198020 |
| 14578 | Nortriptyline 30mg / Fluphenazine 1.5mg tablets | 68990020 |
| 14740 | Oxactin 20mg capsules (Discovery Pharmaceuticals) | 80679020 |
| 15163 | Edronax 4mg tablets (Pfizer Ltd) | 84961020 |
| 15268 | Zispin SolTab 45mg orodispersible tablets (Merck Sharp & Dohme Ltd) | 87482020 |
| 15632 | Dothapax 75 tablets (Ashbourne Pharmaceuticals Ltd) | 56699020 |
| 16154 | Mirtazapine 15mg/ml oral solution sugar free | 87990020 |
| 16323 | Perphenazine 2mg with Amitriptyline 10mg tablet | 65592020 |
| 18342 | Amitriptyline 25mg / Chlordiazepoxide 10mg capsules | 68725020 |
| 19168 | Dosulepin 25mg/5ml mixture | 54728020 |
| 19181 | Trazodone 100mg capsules (Mylan) | 62912020 |
| 19183 | Fluoxetine 20mg capsules (A A H Pharmaceuticals Ltd) | 61084020 |
| 19186 | Dosulepin 75mg tablets (Actavis UK Ltd) | 56943020 |
| 19470 | Fluoxetine 20mg capsules (Ranbaxy (UK) Ltd) | 63016020 |
| 19779 | Amitriptyline 10mg/ml injection | 60229020 |
| 20026 | Domical 25mg Tablet (Berk Pharmaceuticals Ltd) | 55363020 |
| 20152 | Escitalopram 10mg/ml oral drops sugar free | 90871020 |
| 20504 | Optimax wv Tablet (E. Merck) | 54978020 |
| 20571 | Fluphenazine with nortriptyline 500microgramswith10mg Tablet | 62045020 |
| 20715 | LIMBITROL 5 | !4097101 |
| 21081 | Amitriptyline 12.5mg / Chlordiazepoxide 5mg capsules | 68724020 |
| 21157 | Thaden 75mg tablets (Opus Pharmaceuticals Ltd) | 84817020 |
| 21819 | Prepadine 75mg tablets (Teva UK Ltd) | 57111020 |
| 21820 | Prepadine 25mg capsules (Teva UK Ltd) | 57110020 |
| 22070 | Amitriptyline 10mg/5ml Oral solution (Rosemont Pharmaceuticals Ltd) | 55688020 |
| 23334 | FAVERIN | !2497701 |
| 23426 | Dosulepin 25mg capsules (A A H Pharmaceuticals Ltd) | 49588020 |
| 24134 | Amitriptyline 25mg tablets (Kent Pharmaceuticals Ltd) | 48517020 |
| 24141 | Amitriptyline 10mg tablets (Actavis UK Ltd) | 48507020 |
| 24145 | Amitriptyline 25mg tablets (Actavis UK Ltd) | 48508020 |
| 24147 | Amitriptyline 25mg tablets (Teva UK Ltd) | 48480020 |
| 24152 | Amitriptyline 10mg tablets (Teva UK Ltd) | 48479020 |
| 24680 | Elavil 10mg Tablet (DDSA Pharmaceuticals Ltd) | 59132020 |
| 24890 | Trifluoperazine with tranylcypromine 1mg + 10mg Tablet | 69003020 |
| 25045 | TRIMIPRAMINE | !7275101 |
| 25085 | TRIMIPRAMINE | !7275103 |
| 25444 | Lomont 70mg/5ml oral suspension (Rosemont Pharmaceuticals Ltd) | 84171020 |
| 26016 | Citalopram 20mg tablets (Sandoz Ltd) | 65128020 |
| 26056 | Cipralex 10mg/ml oral drops (Lundbeck Ltd) | 90873020 |
| 26213 | Domical 10mg Tablet (Berk Pharmaceuticals Ltd) | 55362020 |
| 27008 | Domical 50mg Tablet (Berk Pharmaceuticals Ltd) | 55364020 |
| 27565 | FLUANXOL | !2619101 |
| 29339 | Trazodone 50mg capsules (Mylan) | 62911020 |
| 29756 | Paxoran 20mg Tablet (Ranbaxy (UK) Ltd) | 88348020 |
| 29786 | Ranflutin 20mg capsules (Ranbaxy (UK) Ltd) | 82055020 |
| 29857 | Trazodone 150mg tablets (Teva UK Ltd) | 62363020 |
| 29875 | Dosulepin 25mg capsules (Mylan) | 60464020 |
| 30258 | Fluoxetine 20mg/5ml oral solution (Teva UK Ltd) | 64692020 |
| 30375 | Anafranil 25mg/2ml solution for injection ampoules (Novartis Pharmaceuticals UK Ltd) | 61477020 |
| 30376 | Thaden 25mg capsules (Opus Pharmaceuticals Ltd) | 84816020 |
| 30983 | Trazodone 150mg tablets (Mylan) | 62913020 |
| 31168 | MIRTAZAPINE | !8502180 |
| 31824 | Dosulepin 25mg capsules (IVAX Pharmaceuticals UK Ltd) | 53818020 |
| 31826 | Dosulepin 75mg tablets (IVAX Pharmaceuticals UK Ltd) | 53817020 |
| 32121 | Dosulepin 75mg tablets (A A H Pharmaceuticals Ltd) | 49589020 |
| 32401 | Sertraline 50mg tablets (A A H Pharmaceuticals Ltd) | 71725020 |
| 32439 | Amitriptyline 25mg Tablet (Sussex Pharmaceutical Ltd) | 48534020 |
| 32546 | Paxoran 10mg Tablet (Ranbaxy (UK) Ltd) | 88346020 |
| 32848 | Citalopram 10mg tablets (Actavis UK Ltd) | 67890020 |
| 32863 | Imipramine 10mg tablets (Teva UK Ltd) | 68803020 |
| 32899 | Paroxetine 20mg tablets (Actavis UK Ltd) | 65578020 |
| 33071 | Felicium 20mg capsules (Opus Pharmaceuticals Ltd) | 49680020 |
| 33074 | Praminil 10mg Tablet (DDSA Pharmaceuticals Ltd) | 59528020 |
| 33090 | Amitriptyline 10mg tablets (A A H Pharmaceuticals Ltd) | 48496020 |
| 33164 | Dosulepin 25mg capsules (Sandoz Ltd) | 66765020 |
| 33337 | Mirtazapine 45mg tablets (A A H Pharmaceuticals Ltd) | 69832020 |
| 33410 | Fluoxetine 20mg capsules (Zentiva) | 61616020 |
| 33624 | Amitriptyline 50mg tablets (Teva UK Ltd) | 48481020 |
| 33720 | Citalopram 10mg tablets (IVAX Pharmaceuticals UK Ltd) | 71225020 |
| 33779 | Prozit 20mg/5ml oral solution (Pinewood Healthcare) | 80777020 |
| 33978 | Paroxetine 20mg tablets (Mylan) | 66399020 |
| 34003 | Trazodone 50mg capsules (A A H Pharmaceuticals Ltd) | 61096020 |
| 34046 | Lofepramine 70mg tablets (A A H Pharmaceuticals Ltd) | 59084020 |
| 34058 | Dosulepin 75mg tablets (Teva UK Ltd) | 54620020 |
| 34107 | Amitriptyline 50mg tablets (Wockhardt UK Ltd) | 48486020 |
| 34129 | Amitriptyline 25mg tablets (Wockhardt UK Ltd) | 48485020 |
| 34182 | Amitriptyline 50mg tablets (Kent Pharmaceuticals Ltd) | 48518020 |
| 34197 | Amitriptyline 25mg Tablet (Berk Pharmaceuticals Ltd) | 48492020 |
| 34202 | Fluoxetine 20mg capsules (Genus Pharmaceuticals Ltd) | 61455020 |
| 34216 | Fluoxetine 20mg/5ml oral solution (A A H Pharmaceuticals Ltd) | 64717020 |
| 34222 | Imipramine 10mg tablets (Actavis UK Ltd) | 49841020 |
| 34223 | Dosulepin 25mg capsules (Teva UK Ltd) | 54621020 |
| 34224 | Amitriptyline 25mg/5ml oral solution sugar free (Rosemont Pharmaceuticals Ltd) | 55686020 |
| 34245 | Clomipramine 25mg capsules (A A H Pharmaceuticals Ltd) | 50947020 |
| 34251 | Amitriptyline 50mg/5ml oral solution sugar free (Rosemont Pharmaceuticals Ltd) | 55687020 |
| 34274 | Amitriptyline 50mg tablets (A A H Pharmaceuticals Ltd) | 48498020 |
| 34288 | Fluoxetine 20mg capsules (Mylan) | 61409020 |
| 34294 | Fluoxetine 20mg capsules (IVAX Pharmaceuticals UK Ltd) | 61167020 |
| 34351 | Paroxetine 20mg tablets (IVAX Pharmaceuticals UK Ltd) | 63703020 |
| 34355 | Imipramine 25mg tablets (Actavis UK Ltd) | 49842020 |
| 34356 | Citalopram 20mg tablets (A A H Pharmaceuticals Ltd) | 65381020 |
| 34401 | Amitriptyline 10mg tablets (Wockhardt UK Ltd) | 48484020 |
| 34413 | Citalopram 10mg tablets (Zentiva) | 66149020 |
| 34415 | Citalopram 20mg tablets (Mylan) | 66454020 |
| 34419 | Paroxetine 20mg tablets (A A H Pharmaceuticals Ltd) | 66464020 |
| 34421 | Trazodone 50mg capsules (Zentiva) | 62274020 |
| 34436 | Citalopram 10mg tablets (Mylan) | 66451020 |
| 34456 | Fluoxetine 20mg capsules (Teva UK Ltd) | 61319020 |
| 34466 | Citalopram 40mg tablets (Sandoz Ltd) | 65131020 |
| 34470 | Trazodone 150mg tablets (Zentiva) | 62276020 |
| 34474 | Amitriptyline 25mg Tablet (Regent Laboratories Ltd) | 48528020 |
| 34498 | Citalopram 10mg Tablet (Neo Laboratories Ltd) | 65254020 |
| 34499 | Citalopram 10mg tablets (Sandoz Ltd) | 65125020 |
| 34503 | Amitriptyline 25mg tablets (IVAX Pharmaceuticals UK Ltd) | 48523020 |
| 34525 | Dosulepin 75mg tablets (Mylan) | 60465020 |
| 34578 | Lofepramine 70mg tablets (IVAX Pharmaceuticals UK Ltd) | 56517020 |
| 34580 | Trazodone 100mg capsules (A A H Pharmaceuticals Ltd) | 61097020 |
| 34586 | Citalopram 10mg tablets (A A H Pharmaceuticals Ltd) | 65376020 |
| 34587 | Paroxetine 30mg tablets (A A H Pharmaceuticals Ltd) | 67650020 |
| 34603 | Citalopram 40mg tablets (Mylan) | 66459020 |
| 34634 | Amitriptyline 50mg tablets (Actavis UK Ltd) | 48509020 |
| 34641 | Dosulepin 25mg capsules (Sovereign Medical Ltd) | 62179020 |
| 34643 | Dosulepin 25mg capsules (Almus Pharmaceuticals Ltd) | 68380020 |
| 34672 | Lofepramine 70mg tablets (Sterwin Medicines) | 60055020 |
| 34722 | Citalopram 20mg Tablet (Neo Laboratories Ltd) | 65257020 |
| 34731 | Amitriptyline 10mg tablets (Kent Pharmaceuticals Ltd) | 48516020 |
| 34745 | Dosulepin 25mg capsules (Actavis UK Ltd) | 56942020 |
| 34782 | Amitriptyline 25mg tablets (A A H Pharmaceuticals Ltd) | 48497020 |
| 34813 | Imipramine 25mg tablets (A A H Pharmaceuticals Ltd) | 49846020 |
| 34822 | Citalopram 20mg tablets (Zentiva) | 66152020 |
| 34849 | Fluoxetine 20mg capsules (Tillomed Laboratories Ltd) | 66273020 |
| 34856 | Fluoxetine 60mg capsules (Mylan) | 65464020 |
| 34866 | Clomipramine 10mg capsules (A A H Pharmaceuticals Ltd) | 50946020 |
| 34871 | Citalopram 20mg tablets (Actavis UK Ltd) | 67893020 |
| 34872 | Imipramine 25mg Tablet (C P Pharmaceuticals Ltd) | 49838020 |
| 34916 | Amitriptyline 10mg Tablet (Berk Pharmaceuticals Ltd) | 48490020 |
| 34950 | Lofepramine 70mg tablets (Accord Healthcare Ltd) | 57022020 |
| 34966 | Citalopram 20mg tablets (Teva UK Ltd) | 68045020 |
| 34970 | Citalopram 20mg tablets (Niche Generics Ltd) | 66685020 |
| 35021 | Paroxetine 10mg tablets | 92043020 |
| 35112 | Seroxat 10mg tablets (GlaxoSmithKline UK Ltd) | 93187020 |
| 35258 | Sinepin 25mg capsules (Marlborough Pharmaceuticals Ltd) | 92457020 |
| 35493 | Sinepin 50mg capsules (Marlborough Pharmaceuticals Ltd) | 92459020 |
| 36746 | Citalopram 40mg tablets (A A H Pharmaceuticals Ltd) | 65386020 |
| 36893 | Fluoxetine 20mg/5ml oral solution sugar free | 93984020 |
| 37256 | Prozep 20mg/5ml oral solution (Chemidex Pharma Ltd) | 93918020 |
| 38274 | Clomipramine 50mg/5ml oral suspension | 95021020 |
| 38827 | Triptafen-M tablets (Mercury Pharma Group Ltd) | 95533020 |
| 38890 | Fluoxetine 20mg Capsule (Milpharm Ltd) | 74425020 |
| 39145 | Nortriptyline 10mg/5ml Liquid | 65159020 |
| 39359 | Venlafaxine 75mg modified-release tablets | 96445020 |
| 39360 | Venlafaxine 150mg modified-release tablets | 96447020 |
| 39770 | Tifaxin XL 75mg capsules (Genus Pharmaceuticals Ltd) | 96465020 |
| 39809 | Tifaxin XL 150mg capsules (Genus Pharmaceuticals Ltd) | 96467020 |
| 40048 | ViePax XL 75mg tablets (Dexcel-Pharma Ltd) | 96449020 |
| 40049 | ViePax XL 150mg tablets (Dexcel-Pharma Ltd) | 96451020 |
| 40054 | Venlafaxine 225mg modified-release tablets | 96838020 |
| 40059 | Venlalic XL 75mg tablets (Ethypharm UK Ltd) | 96834020 |
| 40062 | Venlalic XL 150mg tablets (Ethypharm UK Ltd) | 96836020 |
| 40092 | Vensir XL 150mg capsules (Morningside Healthcare Ltd) | 96357020 |
| 40160 | Mirtazapine 30mg tablets (Actavis UK Ltd) | 68213020 |
| 40165 | Paroxetine 30mg tablets (Actavis UK Ltd) | 67575020 |
| 40277 | Vensir XL 75mg capsules (Morningside Healthcare Ltd) | 96355020 |
| 40295 | Valdoxan 25mg tablets (Servier Laboratories Ltd) | 97030020 |
| 40396 | Amitriptyline 50mg Tablet (Berk Pharmaceuticals Ltd) | 48491020 |
| 40407 | Venlalic XL 225mg tablets (Ethypharm UK Ltd) | 96841020 |
| 40494 | Agomelatine 25mg tablets | 97028020 |
| 40514 | Venaxx XL 150mg capsules (AMCo) | 97005020 |
| 40515 | Venaxx XL 75mg capsules (AMCo) | 97003020 |
| 40517 | Vexarin XL 150mg capsules (Mylan) | 96536020 |
| 40726 | Escitalopram 20mg/ml oral drops sugar free | 97169020 |
| 40764 | ViePax 37.5mg tablets (Dexcel-Pharma Ltd) | 96439020 |
| 40777 | Doxepin 25mg/5ml oral suspension | 96353020 |
| 40815 | Tardcaps XL 75mg capsules (IXL Pharma Ltd) | 96329020 |
| 40817 | Tardcaps XL 150mg capsules (IXL Pharma Ltd) | 96331020 |
| 40892 | Paroxetine 20mg tablets (Genus Pharmaceuticals Ltd) | 69754020 |
| 40917 | ViePax 75mg tablets (Dexcel-Pharma Ltd) | 96441020 |
| 41033 | Rodomel XL 75mg capsules (Teva UK Ltd) | 96239020 |
| 41062 | Cipralex 20mg/ml oral drops (Lundbeck Ltd) | 97171020 |
| 41299 | Politid XL 75mg capsules (Actavis UK Ltd) | 96473020 |
| 41314 | Rodomel XL 150mg capsules (Teva UK Ltd) | 96241020 |
| 41408 | Imipramine 25mg tablets (Teva UK Ltd) | 55369020 |
| 41528 | Citalopram 10mg tablets (Teva UK Ltd) | 68042020 |
| 41563 | Clomipramine 25mg capsules (IVAX Pharmaceuticals UK Ltd) | 57825020 |
| 41597 | Clomipramine 50mg capsules (IVAX Pharmaceuticals UK Ltd) | 57826020 |
| 41609 | Trazodone 50mg capsules (Teva UK Ltd) | 62361020 |
| 41627 | Lofepramine 70mg Tablet (Teva UK Ltd) | 59282020 |
| 41628 | Clomipramine 10mg capsules (IVAX Pharmaceuticals UK Ltd) | 57824020 |
| 41654 | Tranylcypromine 10mg tablets (AMCo) | 65266020 |
| 41681 | Imipramine 10mg tablets (A A H Pharmaceuticals Ltd) | 49845020 |
| 41709 | Trazodone 100mg capsules (Teva UK Ltd) | 62362020 |
| 41710 | Trazodone 100mg capsules (Zentiva) | 62275020 |
| 41729 | Amitriptyline 25mg Tablet (Celltech Pharma Europe Ltd) | 48502020 |
| 41731 | Isocarboxazid 10mg Tablet (Cambridge Laboratories Ltd) | 59180020 |
| 41747 | Moclobemide 150mg tablets (Teva UK Ltd) | 63795020 |
| 42078 | Amitriptyline 25mg tablets (Almus Pharmaceuticals Ltd) | 70948020 |
| 42107 | Fluoxetine 20mg capsules (Niche Generics Ltd) | 61420020 |
| 42228 | Trimipramine 10mg tablets (A A H Pharmaceuticals Ltd) | 71733020 |
| 42247 | Imipramine 25mg/5ml oral solution sugar free | 97872020 |
| 42387 | Sertraline 50mg tablets (Actavis UK Ltd) | 72034020 |
| 42394 | Amitriptyline 25mg Tablet (Crosspharma Ltd) | 48513020 |
| 42499 | Fluoxetine 10mg tablets | 98001020 |
| 42600 | Vexarin XL 75mg capsules (Mylan) | 96534020 |
| 42660 | Citalopram 10mg tablets (Almus Pharmaceuticals Ltd) | 71373020 |
| 42734 | Dosulepin 75mg tablets (Almus Pharmaceuticals Ltd) | 68383020 |
| 42803 | Fluoxetine 20mg/5ml oral solution (IVAX Pharmaceuticals UK Ltd) | 66129020 |
| 43024 | Dosulepin 100mg/5ml oral solution | 97461020 |
| 43203 | Venlafaxine 75mg modified-release capsules (Sandoz Ltd) | 96614020 |
| 43234 | Mirtazapine 45mg orodispersible tablets (Teva UK Ltd) | 74033020 |
| 43235 | Mirtazapine 45mg orodispersible tablets (A A H Pharmaceuticals Ltd) | 74708020 |
| 43236 | Mirtazapine 45mg orodispersible tablets (Accord Healthcare Ltd) | 74668020 |
| 43237 | Mirtazapine 15mg orodispersible tablets (Teva UK Ltd) | 74026020 |
| 43239 | Mirtazapine 15mg tablets (A A H Pharmaceuticals Ltd) | 69827020 |
| 43241 | Mirtazapine 15mg orodispersible tablets (Aurobindo Pharma Ltd) | 76512020 |
| 43242 | Mirtazapine 15mg tablets (Genus Pharmaceuticals Ltd) | 71074020 |
| 43246 | Mirtazapine 15mg orodispersible tablets (Genus Pharmaceuticals Ltd) | 74967020 |
| 43247 | Mirtazapine 45mg orodispersible tablets (Genus Pharmaceuticals Ltd) | 74977020 |
| 43248 | Mirtazapine 15mg orodispersible tablets (Focus Pharmaceuticals Ltd) | 74684020 |
| 43250 | Mirtazapine 30mg orodispersible tablets (A A H Pharmaceuticals Ltd) | 74705020 |
| 43253 | Mirtazapine 15mg orodispersible tablets (A A H Pharmaceuticals Ltd) | 74702020 |
| 43256 | Mirtazapine 45mg orodispersible tablets (Focus Pharmaceuticals Ltd) | 74689020 |
| 43257 | Mirtazapine 15mg tablets (Teva UK Ltd) | 75309020 |
| 43334 | Venlafaxine 150mg modified-release capsules (Sandoz Ltd) | 96616020 |
| 43518 | Fluvoxamine 100mg tablets (IVAX Pharmaceuticals UK Ltd) | 60735020 |
| 43519 | Citalopram 40mg Tablet (Neo Laboratories Ltd) | 65262020 |
| 43534 | Lofepramine 70mg/5ml Oral suspension (Rosemont Pharmaceuticals Ltd) | 55646020 |
| 43561 | Clomipramine 10mg capsules (Teva UK Ltd) | 54669020 |
| 43673 | Politid XL 150mg capsules (Actavis UK Ltd) | 96475020 |
| 43968 | Foraven XL 75mg capsules (Forum Products Ltd) | 97351020 |
| 44853 | Dosulepin 25mg capsules (Kent Pharmaceuticals Ltd) | 63443020 |
| 44861 | Fluvoxamine 100mg tablets (Actavis UK Ltd) | 62693020 |
| 44936 | Venlaneo XL 150mg capsules (Kent Pharmaceuticals Ltd) | 99210020 |
| 44937 | Venlaneo XL 75mg capsules (Kent Pharmaceuticals Ltd) | 99208020 |
| 44944 | Sertraline 100mg tablets (Teva UK Ltd) | 72095020 |
| 45223 | Citalopram 40mg tablets (Niche Generics Ltd) | 66688020 |
| 45224 | Fluoxetine 20mg capsules (Sandoz Ltd) | 61387020 |
| 45226 | Trimipramine 25mg tablets (A A H Pharmaceuticals Ltd) | 71736020 |
| 45233 | Amitriptyline 10mg tablets (IVAX Pharmaceuticals UK Ltd) | 48522020 |
| 45242 | Amitriptyline 10mg Tablet (Sussex Pharmaceutical Ltd) | 48533020 |
| 45247 | Fluoxetine 20mg capsules (Fannin UK Ltd) | 62975020 |
| 45286 | Citalopram 10mg tablets (Niche Generics Ltd) | 66680020 |
| 45304 | Citalopram 40mg tablets (Teva UK Ltd) | 68049020 |
| 45316 | Fluoxetine 20mg capsules (Wockhardt UK Ltd) | 61439020 |
| 45318 | Clomipramine 50mg capsules (A A H Pharmaceuticals Ltd) | 50948020 |
| 45329 | Fluoxetine 20mg capsules (Actavis UK Ltd) | 61452020 |
| 45350 | Clomipramine 25mg capsules (Teva UK Ltd) | 54670020 |
| 45664 | Depefex XL 150mg capsules (Chiesi Ltd) | 98863020 |
| 45737 | Dosulepin 25mg/5ml Oral solution (Rosemont Pharmaceuticals Ltd) | 55642020 |
| 45806 | Venlafaxine 37.5mg modified-release tablets | 99679020 |
| 45818 | Venlalic XL 37.5mg tablets (Ethypharm UK Ltd) | 99681020 |
| 45915 | Sertraline 50mg tablets (Almus Pharmaceuticals Ltd) | 75620020 |
| 45959 | Depefex XL 75mg capsules (Chiesi Ltd) | 98861020 |
| 46668 | Mirtazapine 15mg tablets (Arrow Generics Ltd) | 70353020 |
| 46801 | Amitriptyline 10mg/5ml oral solution | 503021 |
| 46818 | Amitriptyline 10mg/5ml oral suspension | 505021 |
| 46926 | Citalopram 40mg tablets (Zentiva) | 66160020 |
| 46970 | Amitriptyline 50mg tablets (IVAX Pharmaceuticals UK Ltd) | 48524020 |
| 46977 | Citalopram 40mg tablets (Actavis UK Ltd) | 67898020 |
| 47363 | Mianserin 20mg Tablet (Berk Pharmaceuticals Ltd) | 50103020 |
| 47945 | Mirtazapine 30mg tablets (A A H Pharmaceuticals Ltd) | 68484020 |
| 47966 | Mirtazapine 15mg/ml oral solution sugar free (Rosemont Pharmaceuticals Ltd) | 68128020 |
| 48026 | Citalopram 20mg tablets (Almus Pharmaceuticals Ltd) | 71376020 |
| 48045 | Fluvoxamine 100mg tablets (A A H Pharmaceuticals Ltd) | 66244020 |
| 48065 | Amitriptyline oral solution | 92481020 |
| 48185 | Mirtazapine 30mg orodispersible tablets (Almus Pharmaceuticals Ltd) | 78577020 |
| 48199 | Ranfaxine XL 75mg capsules (Ranbaxy (UK) Ltd) | 97656020 |
| 48216 | Nortriptyline 25mg tablets (A A H Pharmaceuticals Ltd) | 77778020 |
| 48220 | Prozac 20mg capsules (Lexon (UK) Ltd) | 3837020 |
| 48698 | Mirtazapine 15mg orodispersible tablets sugar free | 41252020 |
| 49165 | Citalopram 10mg tablets (Alliance Healthcare (Distribution) Ltd) | 4004020 |
| 49511 | Venlablue XL 75mg capsules (Creo Pharma Ltd) | 3965020 |
| 49519 | Sertraline 100mg/5ml oral suspension | 20738020 |
| 49820 | Mirtazapine 45mg orodispersible tablets sugar free | 41254020 |
| 50081 | Venlablue XL 150mg capsules (Creo Pharma Ltd) | 3976020 |
| 50592 | Fluanxol 1mg tablets (Sigma Pharmaceuticals Plc) | 3818020 |
| 50722 | Dosulepin 25mg/5ml oral solution | 19725020 |
| 50892 | Zispin SolTab 15mg orodispersible tablets (Necessity Supplies Ltd) | 9892020 |
| 50934 | Venlafaxine 150mg/5ml oral solution | 35355020 |
| 51280 | Efexor XL 150mg capsules (Waymade Healthcare Plc) | 3970020 |
| 51361 | Venlafaxine 37.5mg tablets (Ranbaxy (UK) Ltd) | 3942020 |
| 51383 | Duloxetine 60mg gastro-resistant capsules (Sigma Pharmaceuticals Plc) | 10967020 |
| 51699 | Venlafaxine 37.5mg/5ml oral solution | 35357020 |
| 51758 | Prothiaden 25mg capsules (Stephar (U.K.) Ltd) | 3717020 |
| 52074 | Alventa XL 75mg capsules (Consilient Health Ltd) | 3966020 |
| 52100 | Citalopram 10mg tablets (Arrow Generics Ltd) | 4007020 |
| 52354 | Citalopram 20mg tablets (DE Pharmaceuticals) | 38526020 |
| 52408 | Citalopram 10mg tablets (Kent Pharmaceuticals Ltd) | 4003020 |
| 52516 | Alventa XL 150mg capsules (Consilient Health Ltd) | 3977020 |
| 52607 | Citalopram 20mg tablets (Bristol Laboratories Ltd) | 3997020 |
| 52716 | Tonpular XL 75mg capsules (Wockhardt UK Ltd) | 98348020 |
| 52824 | Citalopram 10mg tablets (PLIVA Pharma Ltd) | 71214020 |
| 52867 | Amitriptyline 10mg tablets (Accord Healthcare Ltd) | 3674020 |
| 53161 | Clomipramine 50mg/5ml oral solution | 19451020 |
| 53187 | Clomipramine 50mg capsules (Kent Pharmaceuticals Ltd) | 3701020 |
| 53321 | Mirtazapine 15mg/ml oral solution sugar free (A A H Pharmaceuticals Ltd) | 21206020 |
| 53326 | Venlafaxine 75mg/5ml oral solution | 35359020 |
| 53394 | Citalopram 20mg tablets (Alliance Healthcare (Distribution) Ltd) | 3985020 |
| 53543 | Zispin SolTab 30mg orodispersible tablets (Necessity Supplies Ltd) | 9905020 |
| 53648 | Mirtazapine 30mg orodispersible tablets (Accord Healthcare Ltd) | 74665020 |
| 53699 | Mirtazapine 15mg tablets (Actavis UK Ltd) | 76425020 |
| 53787 | Citalopram 10mg tablets (Bristol Laboratories Ltd) | 4011020 |
| 53808 | Trimipramine 10mg tablets (Phoenix Healthcare Distribution Ltd) | 3784020 |
| 54012 | Mirtazapine 15mg orodispersible tablets sugar free (Sandoz Ltd) | 41253020 |
| 54081 | Sertraline 25mg/5ml oral suspension | 33816020 |
| 54342 | Mirtazapine 15mg tablets (Medreich Plc) | 38595020 |
| 54644 | Mirtazapine 15mg tablets (Pfizer Ltd) | 9876020 |
| 54686 | Tryptophan 500mg capsules | 47014020 |
| 54747 | Optimax 500mg capsules (Intrapharm Laboratories Ltd) | 47015020 |
| 54792 | Mirtazapine 30mg tablets (Alliance Healthcare (Distribution) Ltd) | 4053020 |
| 54826 | Sertraline 150mg/5ml oral suspension | 33812020 |
| 54827 | Citalopram 10mg/5ml oral suspension | 30393020 |
| 54877 | Amitriptyline 25mg tablets (Accord Healthcare Ltd) | 3679020 |
| 54933 | Sertraline 100mg tablets (PLIVA Pharma Ltd) | 72071020 |
| 55023 | Paroxetine 20mg tablets (Medreich Plc) | 45165020 |
| 55033 | Citalopram 40mg tablets (DE Pharmaceuticals) | 38528020 |
| 55137 | Trazodone 150mg/5ml oral suspension | 34736020 |
| 55138 | Trazodone 250mg/5ml oral solution | 34738020 |
| 55139 | Amitriptyline 25mg tablets (Alliance Healthcare (Distribution) Ltd) | 3675020 |
| 55146 | Sertraline 100mg tablets (A A H Pharmaceuticals Ltd) | 71728020 |
| 55424 | Venlafaxine | 95179020 |
| 55482 | Mirtazapine 15mg orodispersible tablets (Mylan) | 77222020 |
| 55488 | Sertraline 50mg tablets (Teva UK Ltd) | 72091020 |
| 55491 | Amitriptyline 10mg tablets (Almus Pharmaceuticals Ltd) | 70945020 |
| 55501 | Venlafaxine 150mg Modified-release capsule (Hillcross Pharmaceuticals Ltd) | 76126020 |
| 55537 | Seroxat 30mg tablets (Lexon (UK) Ltd) | 3921020 |
| 55970 | Nortriptyline 10mg tablets (King Pharmaceuticals Ltd) | 3751020 |
| 56009 | Citalopram 20mg tablets (Arrow Generics Ltd) | 3990020 |
| 56209 | Mirtazapine 30mg tablets (Phoenix Healthcare Distribution Ltd) | 4060020 |
| 56229 | Lofepramine 70mg/5ml oral solution | 32936020 |
| 56292 | Citalopram 40mg/ml oral drops sugar free (Actavis UK Ltd) | 4037020 |
| 56355 | Citalopram 10mg tablets (Waymade Healthcare Plc) | 47301020 |
| 56457 | Venlafaxine 75mg tablets (Teva UK Ltd) | 76076020 |
| 56501 | Tofranil 25mg tablets (Lexon (UK) Ltd) | 3734020 |
| 56662 | Venlafaxine 37.5mg tablets (A A H Pharmaceuticals Ltd) | 76115020 |
| 56703 | Lofepramine 70mg tablets (Sandoz Ltd) | 60806020 |
| 57107 | Amitriptyline 10mg tablets (Phoenix Healthcare Distribution Ltd) | 3673020 |
| 57226 | Trazodone 25mg/5ml oral suspension | 34744020 |
| 57532 | Prozac 20mg capsules (Waymade Healthcare Plc) | 3831020 |
| 57751 | Tonpular XL 150mg capsules (Wockhardt UK Ltd) | 98350020 |
| 57926 | Dosulepin 75mg/5ml oral solution | 19721020 |
| 57936 | Citalopram 40mg/ml oral drops sugar free (A A H Pharmaceuticals Ltd) | 76354020 |
| 57972 | Amitriptyline 10mg tablets (Alliance Healthcare (Distribution) Ltd) | 3671020 |
| 57978 | Trimipramine 25mg tablets (Waymade Healthcare Plc) | 47285020 |
| 58291 | Mirtazapine 15mg orodispersible tablets (Pfizer Ltd) | 9888020 |
| 58476 | Citalopram 20mg tablets (Aurobindo Pharma Ltd) | 4002020 |
| 58625 | Mirtazapine 45mg tablets (Actavis UK Ltd) | 76428020 |
| 58664 | Sertraline 50mg tablets (Mylan) | 72232020 |
| 58723 | Sertraline 50mg tablets (Accord Healthcare Ltd) | 39811020 |
| 58837 | Venlafaxine 37.5mg modified-release capsules | 20695021 |
| 59035 | Venlablue XL 37.5mg capsules (Creo Pharma Ltd) | 20696021 |
| 59161 | Amitriptyline 10mg tablets (Waymade Healthcare Plc) | 47269020 |
| 59193 | Citalopram 10mg tablets (Ranbaxy (UK) Ltd) | 69079020 |
| 59288 | Paroxetine 10mg tablets (Actavis UK Ltd) | 45504020 |
| 59358 | Fluoxetine 20mg capsules (Milpharm Ltd) | 3843020 |
| 59563 | Venlafaxine 75mg modified-release capsules (Kent Pharmaceuticals Ltd) | 3963020 |
| 59593 | Fluanxol 500microgram tablets (Lexon (UK) Ltd) | 3813020 |
| 59600 | Sertraline 100mg tablets (Almus Pharmaceuticals Ltd) | 75616020 |
| 59650 | Citalopram 10mg tablets (Aurobindo Pharma Ltd) | 4014020 |
| 59694 | Mirtazapine 30mg orodispersible tablets (Phoenix Healthcare Distribution Ltd) | 9903020 |
| 59820 | Amitriptyline 50mg/5ml oral solution sugar free (Wockhardt UK Ltd) | 77899020 |
| 59923 | Venlafaxine 37.5mg tablets (Bristol Laboratories Ltd) | 43833020 |
| 59931 | Trazodone 50mg/5ml oral solution sugar free (A A H Pharmaceuticals Ltd) | 78020020 |
| 59953 | Mirtazapine 15mg tablets (Almus Pharmaceuticals Ltd) | 9878020 |
| 59954 | Mirtazapine 45mg tablets (Almus Pharmaceuticals Ltd) | 9912020 |
| 60138 | Fluoxetine 20mg orodispersible tablets sugar free | 23601021 |
| 60355 | Amitriptyline 25mg tablets (Phoenix Healthcare Distribution Ltd) | 3678020 |
| 60370 | Zispin SolTab 15mg orodispersible tablets (Mawdsley-Brooks & Company Ltd) | 9889020 |
| 60410 | Amitriptyline 25mg/5ml oral solution sugar free (Wockhardt UK Ltd) | 77894020 |
| 60449 | Venlafaxine 75mg tablets (A A H Pharmaceuticals Ltd) | 76118020 |
| 60534 | Fluoxetine 20mg dispersible tablets sugar free | 24094021 |
| 60538 | Mirtazapine 30mg tablets (DE Pharmaceuticals) | 23710021 |
| 60549 | Venlafaxine 150mg modified-release capsules (Kent Pharmaceuticals Ltd) | 3975020 |
| 60568 | Citalopram 20mg tablets (Waymade Healthcare Plc) | 47300020 |
| 60591 | Lofepramine 70mg tablets (Teva UK Ltd) | 75291020 |
| 60619 | Fluoxetine 20mg/5ml oral solution (Kent Pharmaceuticals Ltd) | 3844020 |
| 60839 | Citalopram 40mg tablets (Almus Pharmaceuticals Ltd) | 71379020 |
| 60888 | Citalopram 10mg tablets (Sigma Pharmaceuticals Plc) | 4010020 |
| 60895 | Venlafaxine 37.5mg tablets (Teva UK Ltd) | 76073020 |
| 60962 | Fluoxetine 20mg capsules (Alliance Healthcare (Distribution) Ltd) | 3830020 |
| 61335 | Prozac 20mg capsules (Mawdsley-Brooks & Company Ltd) | 3838020 |
| 61503 | Sertraline 100mg tablets (Actavis UK Ltd) | 72038020 |
| 61547 | Mirtazapine 15mg/ml oral solution sugar free (DE Pharmaceuticals) | 24001021 |
| 61657 | Trazodone 75mg/5ml oral solution | 34750020 |
| 61835 | Amitriptyline 10mg tablets (DE Pharmaceuticals) | 16423021 |
| 61842 | Trazodone 50mg/5ml oral solution | 34746020 |
| 61856 | Mirtazapine 15mg orodispersible tablets (Consilient Health Ltd) | 9884020 |
| 62155 | Fluoxetine 20mg capsules (Phoenix Healthcare Distribution Ltd) | 3839020 |
| 62620 | Clomipramine 10mg capsules (Mylan) | 60329020 |
| 62681 | Dosulepin 75mg tablets (Sandoz Ltd) | 66768020 |
| 62688 | Duloxetine 30mg gastro-resistant capsules (Sigma Pharmaceuticals Plc) | 10980020 |
| 62692 | Sertraline 100mg tablets (Bristol Laboratories Ltd) | 3894020 |
| 62693 | Sertraline 50mg tablets (Bristol Laboratories Ltd) | 3875020 |
| 62734 | Venlafaxine 150mg/5ml oral suspension | 35023020 |
| 62819 | Sertraline 12.5mg/5ml oral suspension | 33810020 |
| 62927 | Sertraline 50mg tablets (Wockhardt UK Ltd) | 72107020 |
| 62950 | Sertraline 100mg tablets (Accord Healthcare Ltd) | 39812020 |
| 63268 | Venlafaxine 75mg/5ml oral suspension | 35361020 |
| 63276 | Nortriptyline 25mg tablets (Alliance Healthcare (Distribution) Ltd) | 3761020 |
| 63370 | Duloxetine 30mg gastro-resistant capsules (Mawdsley-Brooks & Company Ltd) | 10981020 |
| 63403 | Mirtazapine 30mg tablets (Teva UK Ltd) | 69054020 |
| 63441 | Citalopram 10mg tablets (Rivopharm (UK) Ltd) | 44246021 |
| 63481 | Sertraline 50mg tablets (Milpharm Ltd) | 3878020 |
| 63763 | Duloxetine 60mg gastro-resistant capsules (A A H Pharmaceuticals Ltd) | 48890021 |
| 63859 | Venlafaxine 75mg tablets (Waymade Healthcare Plc) | 47297020 |
| 63916 | Escitalopram 10mg tablets (Actavis UK Ltd) | 26791021 |
| 63953 | Cipramil 20mg tablets (DE Pharmaceuticals) | 3994020 |
| 64101 | Mirtazapine 15mg orodispersible tablets (Accord Healthcare Ltd) | 74662020 |
| 64139 | Mirtazapine 45mg orodispersible tablets (Mylan) | 77228020 |
| 64141 | Amitriptyline 5mg/5ml oral solution | 18973020 |
| 64223 | Mirtazapine 45mg tablets (Teva UK Ltd) | 75313020 |
| 64330 | Amitriptyline 50mg tablets (Almus Pharmaceuticals Ltd) | 70951020 |
| 64423 | Citalopram 10mg tablets (Accord Healthcare Ltd) | 4013020 |
| 64442 | Duloxetine 60mg gastro-resistant capsules (Teva UK Ltd) | 52348021 |
| 64458 | Clomipramine 25mg/5ml oral suspension | 19449020 |
| 64647 | Amitriptyline 25mg tablets (DE Pharmaceuticals) | 16424021 |
| 64785 | Paroxetine 30mg tablets (Alliance Healthcare (Distribution) Ltd) | 3913020 |
| 65152 | Trazodone 100mg/5ml oral solution | 34726020 |
| 65213 | Trimipramine 50mg/5ml oral solution | 34986020 |
| 65237 | Nortriptyline 10mg tablets (A A H Pharmaceuticals Ltd) | 77775020 |
| 65439 | Amitriptyline 25mg tablets (Sandoz Ltd) | 68492020 |
| 65445 | Trimipramine 50mg capsules (A A H Pharmaceuticals Ltd) | 71739020 |
| 65555 | Mirtazapine 15mg orodispersible tablets (Sigma Pharmaceuticals Plc) | 9886020 |
| 65618 | Duloxetine 30mg gastro-resistant capsules (A A H Pharmaceuticals Ltd) | 48892021 |
| 65666 | Venlafaxine 225mg modified-release capsules | 60861021 |
| 65738 | Efexor 37.5mg tablets (Sigma Pharmaceuticals Plc) | 3939020 |
| 65762 | Clomipramine 25mg capsules (Waymade Healthcare Plc) | 47275020 |
| 65771 | Sertraline 200mg/5ml oral suspension (Special Order) | 52635021 |
| 65804 | Clomipramine 50mg capsules (Teva UK Ltd) | 54671020 |
| 65809 | Duloxetine 30mg gastro-resistant capsules (Actavis UK Ltd) | 48893021 |
| 65879 | Amitriptyline 10mg tablets (Sigma Pharmaceuticals Plc) | 52215021 |
| 65888 | Duloxetine 60mg gastro-resistant capsules (Actavis UK Ltd) | 48889021 |
| 65892 | Duloxetine 60mg gastro-resistant capsules (Mawdsley-Brooks & Company Ltd) | 53770021 |
| 65899 | Efexor XL 225mg capsules (Pfizer Ltd) | 60862021 |
| 65987 | Amitriptyline 25mg tablets (Crescent Pharma Ltd) | 48019021 |
| anxiety |  |  |
| 20 | Temazepam 10mg tablets | 68342020 |
| 35 | Nitrazepam 5mg tablets | 58928020 |
| 66 | Zopiclone 7.5mg tablets | 72416020 |
| 563 | Clomethiazole 192mg capsules | 61174020 |
| 664 | Lorazepam 4mg/1ml solution for injection ampoules | 64057020 |
| 721 | Zopiclone 3.75mg tablets | 72417020 |
| 780 | Temazepam 10mg/5ml oral solution sugar free | 68341020 |
| 921 | Temazepam 10mg capsules | 66877020 |
| 1088 | Lorazepam 1mg tablets | 58993020 |
| 1729 | Temazepam 20mg tablets | 68343020 |
| 1730 | Trazodone 100mg capsules | 67152020 |
| 2017 | Zolpidem 5mg tablets | 75692020 |
| 2091 | Lorazepam 2.5mg tablets | 58994020 |
| 2394 | Buspar 5mg tablets (IXL Pharma Ltd) | 68298020 |
| 2403 | Temazepam 20mg capsules | 66878020 |
| 2407 | Nitrazepam 5mg Capsule | 65063020 |
| 2828 | Meprobamate 400mg tablets | 64406020 |
| 3126 | Stilnoct 5mg tablets (Sanofi) | 75696020 |
| 3205 | Diazepam 5mg | 62418020 |
| 3320 | Zimovane 7.5mg tablets (Sanofi) | 72408020 |
| 3354 | Lormetazepam 1mg tablets | 64061020 |
| 3355 | Trazodone 50mg capsules | 67151020 |
| 3357 | Lormetazepam 1mg Capsule | 64062020 |
| 3491 | Heminevrin 192mg capsules (AstraZeneca UK Ltd) | 49790020 |
| 3524 | Mogadon 5mg Capsule (Roche Products Ltd) | 58776020 |
| 3574 | Buspirone 5mg tablets | 68294020 |
| 3686 | Nitrazepam 10mg Tablet | 65066020 |
| 3687 | Lormetazepam 500microgram tablets | 64060020 |
| 3741 | Stilnoct 10mg tablets (Sanofi) | 75697020 |
| 3870 | Diazepam 2mg capsules | 62417020 |
| 4003 | Molipaxin 150mg tablets (Zentiva) | 54110020 |
| 4020 | Trazodone 150mg tablets | 67153020 |
| 4140 | Oxazepam 30mg Capsule | 65255020 |
| 4187 | Zimovane LS 3.75mg tablets (Sanofi) | 83069020 |
| 4194 | Molipaxin 100mg capsules (Zentiva) | 54109020 |
| 4338 | Valium 5mg Tablet (Roche Products Ltd) | 52130020 |
| 4874 | Molipaxin 50mg capsules (Zentiva) | 54108020 |
| 5058 | Zileze 3.75 tablets (Opus Pharmaceuticals Ltd) | 85821020 |
| 5306 | Sonata 10mg capsules (Meda Pharmaceuticals Ltd) | 86244020 |
| 5352 | Zaleplon 10mg capsules | 86241020 |
| 5385 | Buspirone 10mg tablets | 68295020 |
| 5459 | Zolpidem 10mg tablets | 75693020 |
| 5916 | Zaleplon 5mg capsules | 86240020 |
| 6442 | Trazodone 50mg/5ml oral solution sugar free | 67156020 |
| 7444 | Ativan 4mg/1ml solution for injection ampoules (Pfizer Ltd) | 48240020 |
| 7567 | Temazepam planpak Capsule (Manufacturer unknown) | 70449020 |
| 7569 | Temazepam 15mg capsules | 66879020 |
| 7786 | Mogadon 5mg Tablet (ICN Pharmaceuticals France S.A.) | 50487020 |
| 7924 | Nitrazepam 2.5mg/5ml oral suspension | 65067020 |
| 8174 | Molipaxin 50mg/5ml oral liquid (Sanofi) | 54779020 |
| 8721 | Oxazepam 30mg Tablet | 59842020 |
| 8798 | Temazepam gelthix 10mg Capsule (Pharmacia Ltd) | 73176020 |
| 8913 | Librium 10mg Capsule (ICN Pharmaceuticals France S.A.) | 50188020 |
| 9008 | Buspar 10mg tablets (IXL Pharma Ltd) | 68299020 |
| 9048 | Librium 5mg Tablet (ICN Pharmaceuticals France S.A.) | 50189020 |
| 9111 | Diazepam 10mg capsules | 79693020 |
| 9598 | Sonata 5mg capsules (Meda Pharmaceuticals Ltd) | 86243020 |
| 9696 | Alprazolam 250microgram tablets | 67486020 |
| 9814 | Nitrazepam 5mg/5ml oral suspension | 65068020 |
| 10402 | Valium 10mg Tablet (Roche Products Ltd) | 52131020 |
| 10430 | Temazepam 30mg capsules | 72955020 |
| 10802 | Xanax 250microgram tablets (Pfizer Ltd) | 56351020 |
| 10954 | Ativan 1mg Tablet (Wyeth Pharmaceuticals) | 48244020 |
| 11486 | Alprazolam 500microgram tablets | 67487020 |
| 12293 | Normison 10mg Capsule (Wyeth Pharmaceuticals) | 50769020 |
| 12462 | Temazepam gelthix 20mg Capsule (Pharmacia Ltd) | 73178020 |
| 12477 | Librium 5mg Capsule (ICN Pharmaceuticals France S.A.) | 50187020 |
| 12598 | Xanax 500microgram tablets (Pfizer Ltd) | 56352020 |
| 12710 | Trazodone 150mg modified-release tablets | 67157020 |
| 13279 | Lorazepam 500micrograms/5ml oral suspension | 89956020 |
| 13621 | Molipaxin CR 150mg tablets (Aventis Pharma) | 54780020 |
| 14365 | Zopiclone 3.75mg/5ml oral suspension | 91009020 |
| 15110 | Temazepam 10mg/5ml Oral solution (Generics (UK) Ltd) | 58226020 |
| 15492 | Nitrados 5mg Tablet (Rorer Pharmaceuticals Ltd) | 50709020 |
| 15852 | Zileze 7.5 tablets (Opus Pharmaceuticals Ltd) | 85822020 |
| 16169 | Librium 100mg Injection (Roche Products Ltd) | 53992020 |
| 17294 | Librium 10mg Tablet (ICN Pharmaceuticals France S.A.) | 50193020 |
| 17830 | Ativan 2.5mg Tablet (Wyeth Pharmaceuticals) | 48245020 |
| 18291 | Noctamid 1mg Tablet (Schering Health Care Ltd) | 50737020 |
| 19181 | Trazodone 100mg capsules (Mylan) | 62912020 |
| 19299 | Valium 5mg Capsule (Roche Products Ltd) | 52122020 |
| 19450 | Mogadon 5mg tablets (Meda Pharmaceuticals Ltd) | 88270020 |
| 20164 | Valium 2mg Capsule (Roche Products Ltd) | 52121020 |
| 20245 | Temazepam 10mg gel-fill capsules | 73169020 |
| 20801 | Temazepam gelthix 30mg Capsule (Pharmacia Ltd) | 73180020 |
| 21437 | Loramet 1mg Capsule (Wyeth Pharmaceuticals) | 52944020 |
| 21454 | Normison 20mg Capsule (Wyeth Pharmaceuticals) | 50770020 |
| 23120 | Temazepam gelthix 15mg Capsule (Pharmacia Ltd) | 73177020 |
| 23874 | Somnite 2.5mg/5ml oral suspension (Norgine Pharmaceuticals Ltd) | 51614020 |
| 24135 | Zopiclone 7.5mg tablets (Actavis UK Ltd) | 59209020 |
| 25273 | Oxanid 10mg Tablet (M A Steinhard Ltd) | 65258020 |
| 27367 | Temazepam 10mg/5ml oral solution sugar free (Rosemont Pharmaceuticals Ltd) | 56161020 |
| 27847 | Surem 5mg Capsule (Galen Ltd) | 59025020 |
| 28703 | Evacalm 5mg Tablet (Unimed Pharmaceuticals Ltd) | 59073020 |
| 28880 | Buspirone 5mg Tablet (Galen Ltd) | 59277020 |
| 29219 | Zopiclone 3.75mg tablets (Actavis UK Ltd) | 59210020 |
| 29339 | Trazodone 50mg capsules (Mylan) | 62911020 |
| 29441 | Euhypnos forte 20mg Capsule (Pharmacia Ltd) | 52633020 |
| 29857 | Trazodone 150mg tablets (Teva UK Ltd) | 62363020 |
| 29869 | Zolpidem 5mg Tablet (Winthrop Pharmaceuticals Ltd) | 63881020 |
| 30056 | Zopiclone 3.75mg tablets (IVAX Pharmaceuticals UK Ltd) | 59236020 |
| 30321 | Valium 2mg/5ml Oral solution (Roche Products Ltd) | 52125020 |
| 30377 | Zopiclone 3.75mg tablets (Mylan) | 59785020 |
| 30779 | Temazepam 20mg Tablet (Wyeth Pharmaceuticals) | 50701020 |
| 30981 | Zolpidem 10mg tablets (A A H Pharmaceuticals Ltd) | 64738020 |
| 30983 | Trazodone 150mg tablets (Mylan) | 62913020 |
| 30985 | Temazepam 20mg tablets (IVAX Pharmaceuticals UK Ltd) | 54008020 |
| 31710 | Zolpidem 5mg tablets (A A H Pharmaceuticals Ltd) | 64733020 |
| 32231 | Librium 25mg Tablet (ICN Pharmaceuticals France S.A.) | 50194020 |
| 32320 | Temazepam 20mg gel-fill capsules | 73171020 |
| 32847 | Temazepam 10mg/5ml oral solution sugar free (A A H Pharmaceuticals Ltd) | 51936020 |
| 33045 | Zopiclone 7.5mg tablets (IVAX Pharmaceuticals UK Ltd) | 59237020 |
| 33070 | Solis 5mg Capsule (Galen Ltd) | 59082020 |
| 33086 | Lorazepam 1mg tablets (Teva UK Ltd) | 50645020 |
| 33648 | Temazepam 10mg Tablet (Wyeth Pharmaceuticals) | 50702020 |
| 33663 | Zopiclone 7.5mg tablets (Mylan) | 59784020 |
| 33841 | Zolpidem 10mg tablets (Mylan) | 66946020 |
| 34002 | Temazepam 10mg tablets (IVAX Pharmaceuticals UK Ltd) | 54007020 |
| 34003 | Trazodone 50mg capsules (A A H Pharmaceuticals Ltd) | 61096020 |
| 34292 | Lormetazepam 500microgram tablets (A A H Pharmaceuticals Ltd) | 51447020 |
| 34331 | Temazepam 10mg tablets (A A H Pharmaceuticals Ltd) | 50859020 |
| 34361 | Lormetazepam 500microgram tablets (Thornton & Ross Ltd) | 60685020 |
| 34372 | Zopiclone 7.5mg tablets (PLIVA Pharma Ltd) | 63765020 |
| 34406 | Temazepam 10mg tablets (Teva UK Ltd) | 53646020 |
| 34408 | Nitrazepam 5mg tablets (A A H Pharmaceuticals Ltd) | 49130020 |
| 34421 | Trazodone 50mg capsules (Zentiva) | 62274020 |
| 34470 | Trazodone 150mg tablets (Zentiva) | 62276020 |
| 34508 | Temazepam 10mg tablets (Mylan) | 59161020 |
| 34516 | Lormetazepam 1mg tablets (Mylan) | 60533020 |
| 34534 | Lormetazepam 1mg Tablet (Wyeth Pharmaceuticals) | 49955020 |
| 34555 | Nitrazepam 5mg tablets (Wockhardt UK Ltd) | 49124020 |
| 34561 | Diazepam 2mg Tablet (Regent Laboratories Ltd) | 62837020 |
| 34572 | Temazepam 20mg tablets (Mylan) | 59162020 |
| 34580 | Trazodone 100mg capsules (A A H Pharmaceuticals Ltd) | 61097020 |
| 34612 | Zopiclone 3.75mg tablets (A A H Pharmaceuticals Ltd) | 60509020 |
| 34642 | Lormetazepam 500microgram tablets (Mylan) | 60532020 |
| 34681 | Diazepam 5mg Tablet (Crosspharma Ltd) | 49516020 |
| 34686 | Nitrazepam 5mg tablets (Teva UK Ltd) | 53356020 |
| 34692 | Lormetazepam 1mg tablets (Thornton & Ross Ltd) | 60686020 |
| 34770 | Nitrazepam 5mg Tablet (DDSA Pharmaceuticals Ltd) | 51544020 |
| 34777 | Zopiclone 3.75mg tablets (Teva UK Ltd) | 59593020 |
| 34806 | Nitrazepam 5mg tablets (Mylan) | 64053020 |
| 34823 | Zopiclone 7.5mg tablets (Teva UK Ltd) | 59594020 |
| 34874 | Zopiclone 7.5mg tablets (Kent Pharmaceuticals Ltd) | 63436020 |
| 34876 | Diazepam 2mg Tablet (Berk Pharmaceuticals Ltd) | 49456020 |
| 34892 | Diazepam 5mg Tablet (Berk Pharmaceuticals Ltd) | 49457020 |
| 34897 | Zopiclone 3.75mg tablets (Kent Pharmaceuticals Ltd) | 65546020 |
| 34964 | Nitrazepam 5mg Tablet (Berk Pharmaceuticals Ltd) | 49121020 |
| 35932 | Lorazepam 2.5mg tablets (Teva UK Ltd) | 50646020 |
| 36200 | Lorazepam 1mg tablets (Mylan) | 71875020 |
| 36581 | Atensine 10mg Tablet (Rorer Pharmaceuticals Ltd) | 48237020 |
| 36602 | Temazepam 20mg Tablet (Pharmacia Ltd) | 61571020 |
| 36611 | Euhypnos 10mg/5ml Oral solution (Pharmacia Ltd) | 49359020 |
| 37325 | Remnos 10mg Tablet (DDSA Pharmaceuticals Ltd) | 65074020 |
| 37566 | Lorazepam 1mg/5ml oral suspension | 94513020 |
| 37745 | Lorazepam 1mg/5ml oral solution | 94516020 |
| 38418 | Temazepam 10mg Capsule (Berk Pharmaceuticals Ltd) | 50687020 |
| 38424 | Temazepam 20mg Capsule (Berk Pharmaceuticals Ltd) | 50688020 |
| 39284 | Lorazepam 1mg tablets (Genus Pharmaceuticals Ltd) | 51443020 |
| 40153 | Buspirone 10mg Tablet (Galen Ltd) | 59278020 |
| 41385 | Nitrazepam 5mg tablets (Actavis UK Ltd) | 54170020 |
| 41391 | Lorazepam 1mg tablets (Arrow Generics Ltd) | 66962020 |
| 41516 | Temazepam 10mg Tablet (IVAX Pharmaceuticals UK Ltd) | 55860020 |
| 41539 | Zolpidem 10mg tablets (IVAX Pharmaceuticals UK Ltd) | 65835020 |
| 41542 | Oxazepam 10mg Tablet (IVAX Pharmaceuticals UK Ltd) | 57821020 |
| 41562 | Temazepam 10mg Tablet (Pharmacia Ltd) | 53651020 |
| 41601 | Oxazepam 15mg Tablet (IVAX Pharmaceuticals UK Ltd) | 57820020 |
| 41609 | Trazodone 50mg capsules (Teva UK Ltd) | 62361020 |
| 41653 | Temazepam 20mg Capsule (Hillcross Pharmaceuticals Ltd) | 50696020 |
| 41696 | Zolpidem 5mg tablets (Teva UK Ltd) | 64484020 |
| 41697 | Zolpidem 5mg tablets (IVAX Pharmaceuticals UK Ltd) | 65832020 |
| 41709 | Trazodone 100mg capsules (Teva UK Ltd) | 62362020 |
| 41710 | Trazodone 100mg capsules (Zentiva) | 62275020 |
| 41717 | Temazepam 20mg tablets (A A H Pharmaceuticals Ltd) | 50860020 |
| 41718 | Temazepam 10mg Capsule (Hillcross Pharmaceuticals Ltd) | 50695020 |
| 42089 | Zolpidem 10mg Tablet (Winthrop Pharmaceuticals Ltd) | 63882020 |
| 42814 | Lorazepam 2.5mg tablets (Genus Pharmaceuticals Ltd) | 51444020 |
| 43240 | Buspirone 5mg tablets (Mylan) | 77191020 |
| 43445 | Zopiclone 7.5mg tablets (A A H Pharmaceuticals Ltd) | 60510020 |
| 43560 | Zolpidem 10mg tablets (Teva UK Ltd) | 64487020 |
| 45135 | Diazepam 2mg Tablet (M & A Pharmachem Ltd) | 63587020 |
| 45254 | Temazepam 10mg tablets (Actavis UK Ltd) | 57123020 |
| 45275 | Buspirone 5mg tablets (Actavis UK Ltd) | 61689020 |
| 45283 | Temazepam 10mg tablets (Genus Pharmaceuticals Ltd) | 61199020 |
| 45353 | Zopiclone 7.5mg tablets (Sandoz Ltd) | 62715020 |
| 45829 | Lorazepam 1mg tablets (Sandoz Ltd) | 74808020 |
| 46078 | Temazepam 20mg tablets (Genus Pharmaceuticals Ltd) | 61200020 |
| 46799 | Zopiclone 3.75mg/5ml oral solution | 479021 |
| 46847 | Buspirone 10mg tablets (Actavis UK Ltd) | 61690020 |
| 46896 | Lorazepam 500micrograms/5ml oral solution | 445021 |
| 46939 | Temazepam 20mg tablets (Teva UK Ltd) | 56303020 |
| 46953 | Nitrazepam 5mg tablets (Ranbaxy (UK) Ltd) | 60189020 |
| 46964 | Temazepam 20mg tablets (Actavis UK Ltd) | 57124020 |
| 48517 | Lormetazepam 1mg/5ml oral suspension | 20436020 |
| 49504 | Buspar 10mg tablets (Lexon (UK) Ltd) | 3295020 |
| 49589 | Temazepam 10mg tablets (Sandoz Ltd) | 66824020 |
| 52022 | Zimovane 7.5mg tablets (Lexon (UK) Ltd) | 39801020 |
| 54695 | Diazepam 10mg Tablet (M & A Pharmachem Ltd) | 63597020 |
| 55137 | Trazodone 150mg/5ml oral suspension | 34736020 |
| 55836 | Temazepam 10mg/5ml oral solution sugar free (Focus Pharmaceuticals Ltd) | 74527020 |
| 56551 | Lorazepam 5mg/5ml oral solution | 20432020 |
| 56811 | Temazepam 10mg tablets (F.Maltby & Sons Ltd) | 14501021 |
| 56927 | Temazepam 10mg tablets (Ethigen Ltd) | 46781020 |
| 57226 | Trazodone 25mg/5ml oral suspension | 34744020 |
| 57268 | Lorazepam 2.5mg tablets (Sandoz Ltd) | 74813020 |
| 57937 | Zopiclone 3.75mg tablets (Almus Pharmaceuticals Ltd) | 71655020 |
| 59095 | Buspirone 5mg tablets (A A H Pharmaceuticals Ltd) | 65857020 |
| 59170 | Clomethiazole 192mg capsules (A A H Pharmaceuticals Ltd) | 3126020 |
| 59640 | Zopiclone 7.5mg/5ml oral suspension | 20950020 |
| 59931 | Trazodone 50mg/5ml oral solution sugar free (A A H Pharmaceuticals Ltd) | 78020020 |
| 60825 | Temazepam 20mg tablets (Sandoz Ltd) | 66828020 |
| 61443 | Buspirone 10mg tablets (A A H Pharmaceuticals Ltd) | 65862020 |
| 61450 | Lorazepam 1mg tablets (A A H Pharmaceuticals Ltd) | 61626020 |
| 61657 | Trazodone 75mg/5ml oral solution | 34750020 |
| 61886 | Lorazepam 5mg/5ml oral suspension | 20434020 |
| 62645 | Temazepam 20mg Tablet (Lagap) | 57271020 |
| 63592 | Zopiclone 7.5mg tablets (Sigma Pharmaceuticals Plc) | 3170020 |
| 63665 | Noctamid 0.5mg Tablet (Schering Health Care Ltd) | 50738020 |
| 63674 | Temazepam 30mg gel-fill capsules | 73174020 |
| 64729 | Lorazepam 1mg tablets (Genesis Pharmaceuticals Ltd) | 40522020 |
| 64775 | Nitrazepam 10mg/5ml oral suspension | 31061020 |
| 64876 | Lorazepam 2mg/5ml oral suspension | 20428020 |
| 65152 | Trazodone 100mg/5ml oral solution | 34726020 |
| 65190 | Zolpidem 10mg tablets (Zentiva) | 42169020 |
| 65637 | Zopiclone 7.5mg tablets (Phoenix Healthcare Distribution Ltd) | 3171020 |

Sexual dysfunction

| Medical code | | Read code | | Read term |
| --- | --- | --- | --- | --- |
| 365 | 15D..00 | | Dyspareunia | |
| 2708 | K580.00 | | Dyspareunia due to non psychogenic cause in the female | |
| 15264 | 1595 | | H/O: dyspareunia | |
| 2259 | E227100 | | Inhibited sexual desire | |
| 25196 | 1ABZ.00 | | Sexual activity NOS | |
| 9485 | Eu52.00 | | [X]Sex dysfunction not caused by organic disorder or disease | |
| 103745 | 1P7A.00 | | Duration of sexual abstinence | |
| 28283 | Eu52000 | | [X]Lack or loss of sexual desire | |
| 10811 | 1598.11 | | H/O:sexual problem - female | |
| 31196 | ZG43200 | | Advice for sexual dysfunction | |
| 94316 | 1ABG.00 | | Sexual intercourse difficult | |
| 97690 | 9hK..00 | | Exception reporting: sexual health quality indicators | |
| 29694 | 1598 | | H/O:sexual dysfunction problem | |
| 27693 | 14E3.00 | | H/O: sexual function problem | |
| 21089 | Eu52600 | | [X]Nonorganic dyspareunia | |
| 60716 | Eu52211 | | [X]Female sexual arousal disorder | |
| 56603 | Eu52012 | | [X]Hypoactive sexual desire disorder | |
| 42056 | Eu52100 | | [X]Sexual aversion and lack of sexual enjoyment | |

Antibiotics

| Product code | Product name | **Antibiotic type** |
| --- | --- | --- |
| 21500 | AMIKIN 500 MG INJ | aminoglycosides |
| 14475 | Cidomycin 80mg/ml Injection (Hoechst Marion Roussel) | aminoglycosides |
| 73488 | Gentamicin 1g/vial Powder for injection | aminoglycosides |
| 15584 | Gentamicin 1mg/ml Intrathecal injection | aminoglycosides |
| 4116 | Gentamicin 40mg/ml Injection | aminoglycosides |
| 3236 | Gentamicin 60mg/ml Injection | aminoglycosides |
| 55840 | Gentamicin 80mg/2ml Injection (Hospira UK Ltd) | aminoglycosides |
| 4289 | Gentamicin 80mg/ml Injection | aminoglycosides |
| 52458 | Gentamicin Oral solution | aminoglycosides |
| 29588 | Genticin 1mg/ml Intrathecal injection (Roche Products Ltd) | aminoglycosides |
| 25590 | Kanamycin 1g/vial Powder | aminoglycosides |
| 29363 | Kanamycin 250mg/ml Solution | aminoglycosides |
| 159 | NEOMYCIN 500 MG CAP | aminoglycosides |
| 31854 | NEOMYCIN SULPHATE 500 MG SYR | aminoglycosides |
| 16806 | Netilmicin 100mg/ml Injection | aminoglycosides |
| 335 | STREPTOMYCIN 250 MG CAP | aminoglycosides |
| 144 | STREPTOMYCIN 500 MG CAP | aminoglycosides |
| 17677 | Tobramycin 10mg/ml Injection | aminoglycosides |
| 65446 | Tobramycin 40mg/ml Injection (Actavis UK Ltd) | aminoglycosides |
| 28571 | GENTICIN 25 MG INJ | aminoglycosides |
| 18705 | GENTICIN 80 MG INJ | aminoglycosides |
| 27191 | KANAMYCIN 250 MG CAP | aminoglycosides |
| 28491 | NETILLIN 150 MG INJ | aminoglycosides |
| 32581 | NETILMICIN SULPHATE 150 MG INJ | aminoglycosides |
| 40409 | Bramitob 300mg/4ml nebuliser solution 4ml ampoules (Chiesi Ltd) | aminoglycosides |
| 16884 | Tobi 300mg/5ml nebuliser solution 5ml ampoules (Mylan) | aminoglycosides |
| 47546 | Tobi Podhaler 28mg inhalation powder capsules with device (Mylan) | aminoglycosides |
| 47750 | Tobramycin 28mg inhalation powder capsules with device | aminoglycosides |
| 40046 | Tobramycin 300mg/4ml nebuliser liquid ampoules | aminoglycosides |
| 17628 | Tobramycin 300mg/5ml nebuliser liquid ampoules | aminoglycosides |
| 69812 | Tymbrineb 300mg/5ml nebuliser solution 5ml ampoules (Teva UK Ltd) | aminoglycosides |
| 42459 | Cidomycin Intrathecal 5mg/1ml solution for injection ampoules (Aventis Pharma) | aminoglycosides |
| 21016 | Gentamicin 5mg/1ml solution for injection ampoules | aminoglycosides |
| 59020 | Gentamicin Intrathecal 5mg/1ml solution for injection ampoules (Zentiva) | aminoglycosides |
| 54970 | Gentamicin 225mg implant | aminoglycosides |
| 52337 | Gentamicin 28mg implant | aminoglycosides |
| 45009 | Gentamicin 75mg implant | aminoglycosides |
| 79073 | Septopal Chain (10 beads) 75mg implant (Zimmer Biomet) | aminoglycosides |
| 72837 | Septopal Chain (30 beads) 225mg implant (Zimmer Biomet) | aminoglycosides |
| 409 | Streptomycin 1g powder for solution for injection vials | aminoglycosides |
| 48089 | Streptomycin 1g powder for solution for injection vials (Focus Pharmaceuticals Ltd) | aminoglycosides |
| 71077 | Streptomycin 1g powder for solution for injection vials (Special Order) | aminoglycosides |
| 28915 | Amikacin 100mg/2ml solution for injection vials | aminoglycosides |
| 50313 | Cidomycin 80mg/2ml solution for injection ampoules (Sanofi) | aminoglycosides |
| 49564 | Gentamicin 80mg/2ml solution for injection ampoules | aminoglycosides |
| 66671 | Gentamicin 80mg/2ml solution for injection ampoules (A A H Pharmaceuticals Ltd) | aminoglycosides |
| 62004 | Gentamicin 80mg/2ml solution for injection ampoules (Advanz Pharma) | aminoglycosides |
| 66295 | Gentamicin 80mg/2ml solution for injection ampoules (Wockhardt UK Ltd) | aminoglycosides |
| 20295 | Genticin Injectable 80mg/2ml solution for injection ampoules (Advanz Pharma) | aminoglycosides |
| 7448 | Netilmicin 15mg/1.5ml solution for injection ampoules | aminoglycosides |
| 41052 | Tobramycin 240mg/6ml solution for injection vials | aminoglycosides |
| 78522 | Tobramycin 240mg/6ml solution for injection vials (Pfizer Ltd) | aminoglycosides |
| 51800 | Tobramycin 40mg/1ml solution for injection vials | aminoglycosides |
| 76549 | Tobramycin 40mg/1ml solution for injection vials (A A H Pharmaceuticals Ltd) | aminoglycosides |
| 58875 | Tobramycin 40mg/1ml solution for injection vials (Teva UK Ltd) | aminoglycosides |
| 37360 | Netillin 15mg/1.5ml solution for injection ampoules (Schering-Plough Ltd) | aminoglycosides |
| 64812 | Gentamicin 20mg/2ml solution for injection ampoules | aminoglycosides |
| 66754 | Gentamicin 20mg/2ml solution for injection ampoules (A A H Pharmaceuticals Ltd) | aminoglycosides |
| 77891 | Gentamicin 20mg/2ml solution for injection ampoules (Wockhardt UK Ltd) | aminoglycosides |
| 64774 | Gentamicin 240mg/80ml infusion bags | aminoglycosides |
| 65039 | Gentamicin 360mg/120ml infusion bags | aminoglycosides |
| 64529 | Gentamicin 80mg/80ml infusion bags | aminoglycosides |
| 72811 | Gentamicin 80mg/80ml infusion bags (A A H Pharmaceuticals Ltd) | aminoglycosides |
| 73204 | Gentamicin 80mg/80ml infusion bags (B.Braun Medical Ltd) | aminoglycosides |
| 26813 | Cidomycin 40mg/ml Injection (Aventis Pharma) | aminoglycosides |
| 28377 | Cidomycin 40mg/ml Injection (Beacon Pharmaceuticals Ltd) | aminoglycosides |
| 32714 | Cidomycin 80mg/2ml Injection (Sanofi) | aminoglycosides |
| 40903 | Gentamicin 40mg/ml Injection (Mayne Pharma Plc 1) | aminoglycosides |
| 40588 | Nebcin 80mg/2ml Injection (Flynn Pharma Ltd) | aminoglycosides |
| 32383 | Netillin 100mg/ml Injection (Schering-Plough Ltd) | aminoglycosides |
| 35283 | Tobramycin 40mg/ml Injection | aminoglycosides |
| 8688 | Tobramycin 40mg/ml Injection | aminoglycosides |
| 33746 | Tobramycin 40mg/ml Injection (Mayne Pharma Plc 1) | aminoglycosides |
| 35512 | Tobramycin 80mg/2ml Injection | aminoglycosides |
| 21498 | Amikacin 500mg/2ml solution for injection vials | aminoglycosides |
| 26770 | Amikin 500mg/2ml solution for injection vials (Bristol-Myers Squibb Pharmaceuticals Ltd) | aminoglycosides |
| 51225 | Cidomycin 80mg/2ml solution for injection vials (Sanofi) | aminoglycosides |
| 7302 | Gentamicin 20mg/2ml solution for injection vials | aminoglycosides |
| 28487 | Gentamicin 40mg/1ml solution for injection ampoules | aminoglycosides |
| 49015 | Gentamicin 80mg/2ml solution for injection vials | aminoglycosides |
| 63009 | Gentamicin 80mg/2ml solution for injection vials (A A H Pharmaceuticals Ltd) | aminoglycosides |
| 56669 | Gentamicin 80mg/2ml solution for injection vials (Pfizer Ltd) | aminoglycosides |
| 47747 | Gentamicin Paediatric 20mg/2ml solution for injection vials (Zentiva) | aminoglycosides |
| 15791 | Nebcin 80mg/2ml solution for injection vials (Flynn Pharma Ltd) | aminoglycosides |
| 31332 | Netilmicin 50mg/1ml solution for injection ampoules | aminoglycosides |
| 51900 | Tobramycin 80mg/2ml solution for injection vials | aminoglycosides |
| 72558 | Tobramycin 80mg/2ml solution for injection vials (Pfizer Ltd) | aminoglycosides |
| 29265 | Netillin 50mg/1ml solution for injection ampoules (Schering-Plough Ltd) | aminoglycosides |
| 18229 | Gentamicin 80mg/2ml Injection | aminoglycosides |
| 13566 | Tobramycin 3mg/ml eye drops | aminoglycosides |
| 45746 | Tobravisc 3mg/ml eye drops (Alcon Eye Care Ltd) | aminoglycosides |
| 16591 | Framycetin 250mg Tablet | aminoglycosides |
| 71842 | Gentamicin 62.5mg/5ml oral solution | aminoglycosides |
| 73995 | Neomycin 100mg/5ml oral solution | aminoglycosides |
| 10487 | Neomycin 500mg tablets | aminoglycosides |
| 68406 | Neomycin 500mg tablets (Advanz Pharma) | aminoglycosides |
| 16168 | Neomycin sulphate Oral solution | aminoglycosides |
| 27855 | Nivemycin 100mg/5ml Oral solution (Sovereign Medical Ltd) | aminoglycosides |
| 18696 | Nivemycin 500mg tablets (Advanz Pharma) | aminoglycosides |
| 50239 | Paromomycin 125mg/5ml oral solution | aminoglycosides |
| 75857 | Paromomycin 250mg tablets | aminoglycosides |
| 22059 | Soframycin 250mg Tablet (Hoechst Marion Roussel) | aminoglycosides |
| 78089 | Tobramycin 20mg/5ml oral solution | aminoglycosides |
| 59642 | Tobramycin 400mg/5ml oral solution | aminoglycosides |
| 79513 | Tobramycin 40mg/5ml oral solution | aminoglycosides |
| 18428 | Neomycin sulfate powder | aminoglycosides |
| 325 | Cefalotin 1g/vial injection | cephalosporins |
| 7506 | Cefodizime 1g/vial injection | cephalosporins |
| 12554 | Cefsulodin 1g/vial sterile powder | cephalosporins |
| 14749 | Ceftazidime 2g/vial powder for injection solution | cephalosporins |
| 18313 | CEFTAZIDIME INTERMATE inf DEVICE | cephalosporins |
| 30530 | Ceftazidime with Saline 2g infusion | cephalosporins |
| 67606 | Ceftizoxime 1000mg/vial injection | cephalosporins |
| 7503 | Cefuroxime (as sodium salt) 1.5g/vial infusion | cephalosporins |
| 25666 | Cefuroxime with Saline 750mg infusion | cephalosporins |
| 30471 | Ceporin 1g Injection (Glaxo Laboratories Ltd) | cephalosporins |
| 17676 | Imipenem with cilastatin 250mg+250mg Injection | cephalosporins |
| 25953 | Kefadol 500mg/vial Injection (Dista Products Ltd) | cephalosporins |
| 31333 | Meropenem 250mg Injection | cephalosporins |
| 32750 | Moxalactam 1g/vial Injection (Eli Lilly and Company Ltd) | cephalosporins |
| 43245 | CEFSULODIN SODIUM & LIGNOCAINE 0.5% 1 GM INJ | cephalosporins |
| 21956 | DISTACLOR | cephalosporins |
| 14420 | KEFLEX 500 MG INJ | cephalosporins |
| 20526 | KEFLEX-C 125 MG TAB | cephalosporins |
| 21902 | KEFLEX | cephalosporins |
| 21877 | KEFLEX | cephalosporins |
| 20515 | KEFLEX-C 250 MG TAB | cephalosporins |
| 26581 | PRIMAXIN INJ | cephalosporins |
| 14553 | VELOSEF 125 MG SYR | cephalosporins |
| 17687 | Imipenem 500mg / Cilastatin 500mg powder for suspension for injection vials | cephalosporins |
| 29640 | Primaxin IM 500mg powder for suspension for injection vials (Merck Sharp & Dohme Ltd) | cephalosporins |
| 7501 | Cefotaxime 1g powder for solution for injection vials | cephalosporins |
| 72784 | Cefotaxime 1g powder for solution for injection vials (Wockhardt UK Ltd) | cephalosporins |
| 40155 | Cefotaxime 1g/vial Injection (C P Pharmaceuticals Ltd) | cephalosporins |
| 16983 | Cefradine 1g powder for solution for injection vials | cephalosporins |
| 14995 | Cefradine 500mg powder for solution for injection vials | cephalosporins |
| 2651 | Ceftazidime 1g powder for solution for injection vials | cephalosporins |
| 55897 | Ceftazidime 1g powder for solution for injection vials (Mylan) | cephalosporins |
| 67797 | Ceftazidime 1g powder for solution for injection vials (Wockhardt UK Ltd) | cephalosporins |
| 16595 | Ceftazidime 250mg powder for solution for injection vials | cephalosporins |
| 7502 | Ceftazidime 2g powder for solution for injection vials | cephalosporins |
| 66852 | Ceftazidime 2g powder for solution for injection vials (A A H Pharmaceuticals Ltd) | cephalosporins |
| 55877 | Ceftazidime 2g powder for solution for injection vials (Genus Pharmaceuticals Ltd) | cephalosporins |
| 71271 | Ceftazidime 2g powder for solution for injection vials (Stravencon Ltd) | cephalosporins |
| 20003 | Ceftazidime 3g powder for solution for injection vials | cephalosporins |
| 24046 | Ceftazidime 500mg powder for solution for injection vials | cephalosporins |
| 61881 | Ceftazidime 500mg powder for solution for injection vials (A A H Pharmaceuticals Ltd) | cephalosporins |
| 77241 | Ceftazidime 500mg powder for solution for injection vials (Bowmed Ibisqus Ltd) | cephalosporins |
| 58806 | Ceftazidime 500mg powder for solution for injection vials (Genus Pharmaceuticals Ltd) | cephalosporins |
| 3704 | Ceftriaxone 250mg powder for solution for injection vials | cephalosporins |
| 56444 | Ceftriaxone 250mg powder for solution for injection vials (A A H Pharmaceuticals Ltd) | cephalosporins |
| 58525 | Ceftriaxone 2g powder for solution for infusion vials | cephalosporins |
| 58963 | Ceftriaxone 2g powder for solution for infusion vials (Kent Pharmaceuticals Ltd) | cephalosporins |
| 19838 | Ceftriaxone 2g powder for solution for injection vials | cephalosporins |
| 78007 | Ceftriaxone 2g powder for solution for injection vials (Stravencon Ltd) | cephalosporins |
| 290 | Cefuroxime 1.5g powder for injection vials | cephalosporins |
| 63596 | Cefuroxime 1.5g powder for injection vials (A A H Pharmaceuticals Ltd) | cephalosporins |
| 7587 | Cefuroxime 250mg powder for injection vials | cephalosporins |
| 74202 | Cefuroxime 250mg powder for injection vials (Bowmed Ibisqus Ltd) | cephalosporins |
| 20569 | Claforan 1g powder for solution for injection vials (Sanofi) | cephalosporins |
| 17873 | Fortum 1g powder for solution for injection vials (GlaxoSmithKline UK Ltd) | cephalosporins |
| 32730 | Fortum 250mg powder for solution for injection vials (GlaxoSmithKline UK Ltd) | cephalosporins |
| 68444 | Fortum 2g powder for solution for injection vials (GlaxoSmithKline UK Ltd) | cephalosporins |
| 41982 | Fortum 2g/vial Powder for solution for injection (Glaxo Laboratories Ltd) | cephalosporins |
| 17871 | Fortum 500mg powder for solution for injection vials (GlaxoSmithKline UK Ltd) | cephalosporins |
| 31000 | Kefadim 1g Injection (Eli Lilly and Company Ltd) | cephalosporins |
| 42070 | Kefadim 1g powder for solution for injection vials (Flynn Pharma Ltd) | cephalosporins |
| 32221 | Kefadim 2g Injection (Eli Lilly and Company Ltd) | cephalosporins |
| 23510 | Kefadim 500mg powder for solution for injection vials (Flynn Pharma Ltd) | cephalosporins |
| 26042 | Rocephin 250mg powder for solution for injection vials (Roche Products Ltd) | cephalosporins |
| 28685 | Rocephin 2g powder for solution for injection vials (Roche Products Ltd) | cephalosporins |
| 23779 | Velosef 1g powder for solution for injection vials (Bristol-Myers Squibb Pharmaceuticals Ltd) | cephalosporins |
| 24817 | Velosef 500mg powder for solution for injection vials (Bristol-Myers Squibb Pharmaceuticals Ltd) | cephalosporins |
| 17123 | Zinacef 1.5g powder for injection vials (GlaxoSmithKline UK Ltd) | cephalosporins |
| 28054 | Zinacef 250mg powder for injection vials (GlaxoSmithKline UK Ltd) | cephalosporins |
| 7507 | Cefpirome 1g powder for solution for injection vials | cephalosporins |
| 61470 | Ceftaroline fosamil 600mg powder for solution for infusion vials | cephalosporins |
| 70174 | Ceftolozane 1g / Tazobactam 500mg powder for solution for infusion vials | cephalosporins |
| 78016 | Ertapenem 1g powder for concentrate for solution for infusion vials (Fresenius Kabi Ltd) | cephalosporins |
| 22770 | Ertapenem 1g powder for solution for infusion vials | cephalosporins |
| 14996 | Imipenem 500mg / Cilastatin 500mg powder for solution for infusion vials | cephalosporins |
| 70173 | Imipenem 500mg / Cilastatin 500mg powder for solution for infusion vials (A A H Pharmaceuticals Ltd) | cephalosporins |
| 67944 | Imipenem 500mg / Cilastatin 500mg powder for solution for infusion vials (Kent Pharmaceuticals Ltd) | cephalosporins |
| 47562 | Invanz 1g powder for solution for infusion vials (Merck Sharp & Dohme Ltd) | cephalosporins |
| 27510 | Meronem 1g powder for solution for injection vials (Pfizer Ltd) | cephalosporins |
| 42058 | Meronem 500mg powder for solution for injection vials (Pfizer Ltd) | cephalosporins |
| 16804 | Meropenem 1g powder for solution for injection vials | cephalosporins |
| 71502 | Meropenem 1g powder for solution for injection vials (Alliance Healthcare (Distribution) Ltd) | cephalosporins |
| 72954 | Meropenem 1g powder for solution for injection vials (Pfizer Ltd) | cephalosporins |
| 17114 | Meropenem 500mg powder for solution for injection vials | cephalosporins |
| 70232 | Meropenem 500mg powder for solution for injection vials (Fresenius Kabi Ltd) | cephalosporins |
| 62736 | Meropenem 500mg powder for solution for injection vials (Kent Pharmaceuticals Ltd) | cephalosporins |
| 33200 | Primaxin 500mg+500mg Powder for solution for intravenous infusion (Merck Sharp & Dohme Ltd) | cephalosporins |
| 37857 | Cefuroxime 1.5g with 500mg infusion | cephalosporins |
| 29183 | Cefuroxime 750mg with metronidazole 500mg infusion | cephalosporins |
| 36667 | Kefadim 2g Infusion (Eli Lilly and Company Ltd) | cephalosporins |
| 25667 | Zinacef 1.5g/vial Infusion (Glaxo Laboratories Ltd) | cephalosporins |
| 25614 | Cefotaxime 2g powder for solution for injection vials | cephalosporins |
| 43274 | Cefotaxime 2g/vial Injection (C P Pharmaceuticals Ltd) | cephalosporins |
| 232 | Cefotaxime 500mg powder for solution for injection vials | cephalosporins |
| 57842 | Cefotaxime 500mg powder for solution for injection vials (Genus Pharmaceuticals Ltd) | cephalosporins |
| 41394 | Cefotaxime 500mg/vial Injection (C P Pharmaceuticals Ltd) | cephalosporins |
| 7508 | Cefoxitin 1g powder for solution for injection vials | cephalosporins |
| 684 | Ceftriaxone 1g powder for solution for injection vials | cephalosporins |
| 58239 | Ceftriaxone 1g powder for solution for injection vials (A A H Pharmaceuticals Ltd) | cephalosporins |
| 59708 | Ceftriaxone 1g powder for solution for injection vials (Genus Pharmaceuticals Ltd) | cephalosporins |
| 55895 | Ceftriaxone 1g powder for solution for injection vials (PLIVA Pharma Ltd) | cephalosporins |
| 64903 | Ceftriaxone 1g powder for solution for injection vials (Wockhardt UK Ltd) | cephalosporins |
| 19909 | Cefuroxime 750mg powder for injection vials | cephalosporins |
| 79098 | Cefuroxime 750mg powder for injection vials (Flynn Pharma Ltd) | cephalosporins |
| 32441 | Claforan 2g powder for solution for injection vials (Sanofi) | cephalosporins |
| 30046 | Claforan 500mg powder for solution for injection vials (Sanofi) | cephalosporins |
| 44263 | Mefoxin 1g powder for solution for injection vials (Merck Sharp & Dohme Ltd) | cephalosporins |
| 16202 | Rocephin 1g powder for solution for injection vials (Roche Products Ltd) | cephalosporins |
| 17292 | Zinacef 750mg powder for injection vials (GlaxoSmithKline UK Ltd) | cephalosporins |
| 24661 | Kefadol 1g powder for solution for injection vials (Dista Products Ltd) | cephalosporins |
| 68106 | Ceftazidime 5% eye drops preservative free | cephalosporins |
| 26289 | Bacticlor MR 375mg tablets (Ranbaxy (UK) Ltd) | cephalosporins |
| 12268 | Baxan 125mg/5ml oral suspension (Bristol-Myers Squibb Pharmaceuticals Ltd) | cephalosporins |
| 12496 | Baxan 250mg/5ml oral suspension (Bristol-Myers Squibb Pharmaceuticals Ltd) | cephalosporins |
| 11992 | Baxan 500mg capsules (Bristol-Myers Squibb Pharmaceuticals Ltd) | cephalosporins |
| 17782 | Baxan 500mg/5ml oral suspension (Bristol-Myers Squibb Pharmaceuticals Ltd) | cephalosporins |
| 20549 | Cedax 18mg/ml Liquid (Schering-Plough Ltd) | cephalosporins |
| 21842 | Cedax 36mg/ml Liquid (Schering-Plough Ltd) | cephalosporins |
| 8127 | Cedax 400mg Capsule (Schering-Plough Ltd) | cephalosporins |
| 13910 | Cefaclor 125mg/5ml Liquid (Generics (UK) Ltd) | cephalosporins |
| 14607 | Cefaclor 125mg/5ml Liquid (Lagap) | cephalosporins |
| 1038 | Cefaclor 125mg/5ml oral suspension | cephalosporins |
| 39703 | Cefaclor 125mg/5ml oral suspension (A A H Pharmaceuticals Ltd) | cephalosporins |
| 34913 | Cefaclor 125mg/5ml Oral suspension (Genus Pharmaceuticals Ltd) | cephalosporins |
| 71071 | Cefaclor 125mg/5ml oral suspension (Phoenix Healthcare Distribution Ltd) | cephalosporins |
| 32235 | Cefaclor 125mg/5ml oral suspension (Ranbaxy (UK) Ltd) | cephalosporins |
| 7526 | Cefaclor 125mg/5ml oral suspension sugar free | cephalosporins |
| 56610 | Cefaclor 125mg/5ml oral suspension sugar free (Phoenix Healthcare Distribution Ltd) | cephalosporins |
| 73144 | Cefaclor 125mg/5ml oral suspension sugar free (Teva UK Ltd) | cephalosporins |
| 9520 | Cefaclor 250mg Capsule (Lagap) | cephalosporins |
| 366 | Cefaclor 250mg capsules | cephalosporins |
| 67770 | Cefaclor 250mg capsules (Focus Pharmaceuticals Ltd) | cephalosporins |
| 71038 | Cefaclor 250mg capsules (Phoenix Healthcare Distribution Ltd) | cephalosporins |
| 30772 | Cefaclor 250mg capsules (Ranbaxy (UK) Ltd) | cephalosporins |
| 62640 | Cefaclor 250mg capsules (Teva UK Ltd) | cephalosporins |
| 20420 | Cefaclor 250mg/5ml Liquid (Generics (UK) Ltd) | cephalosporins |
| 20409 | Cefaclor 250mg/5ml Liquid (Lagap) | cephalosporins |
| 3737 | Cefaclor 250mg/5ml oral suspension | cephalosporins |
| 46973 | Cefaclor 250mg/5ml Oral suspension (Genus Pharmaceuticals Ltd) | cephalosporins |
| 67792 | Cefaclor 250mg/5ml oral suspension (Mylan) | cephalosporins |
| 48025 | Cefaclor 250mg/5ml oral suspension (Ranbaxy (UK) Ltd) | cephalosporins |
| 9293 | Cefaclor 250mg/5ml oral suspension sugar free | cephalosporins |
| 65990 | Cefaclor 250mg/5ml oral suspension sugar free (Teva UK Ltd) | cephalosporins |
| 3180 | Cefaclor 375mg modified-release tablets | cephalosporins |
| 34838 | Cefaclor 375mg modified-release tablets (A A H Pharmaceuticals Ltd) | cephalosporins |
| 20881 | Cefaclor 375mg modified-release tablets (Ranbaxy (UK) Ltd) | cephalosporins |
| 4689 | Cefaclor 500mg Capsule (Lagap) | cephalosporins |
| 2976 | Cefaclor 500mg capsules | cephalosporins |
| 43425 | Cefaclor 500mg capsules (A A H Pharmaceuticals Ltd) | cephalosporins |
| 55211 | Cefaclor 500mg capsules (Kent Pharmaceuticals Ltd) | cephalosporins |
| 78824 | Cefaclor 500mg capsules (Phoenix Healthcare Distribution Ltd) | cephalosporins |
| 30771 | Cefaclor 500mg capsules (Ranbaxy (UK) Ltd) | cephalosporins |
| 8051 | Cefaclor 500mg modified-release tablets | cephalosporins |
| 8675 | Cefadroxil 125mg/5ml oral suspension | cephalosporins |
| 5444 | Cefadroxil 250mg/5ml oral suspension | cephalosporins |
| 1942 | Cefadroxil 500mg capsules | cephalosporins |
| 52179 | Cefadroxil 500mg capsules (Phoenix Healthcare Distribution Ltd) | cephalosporins |
| 69614 | Cefadroxil 500mg capsules (Ranbaxy (UK) Ltd) | cephalosporins |
| 56250 | Cefadroxil 500mg capsules (Sandoz Ltd) | cephalosporins |
| 61418 | Cefadroxil 500mg capsules (Sigma Pharmaceuticals Plc) | cephalosporins |
| 66477 | Cefadroxil 500mg capsules (Waymade Healthcare Plc) | cephalosporins |
| 14540 | Cefadroxil 500mg/5ml oral suspension | cephalosporins |
| 12248 | Cefalexin 125mg/1.25ml paediatric drops | cephalosporins |
| 1693 | Cefalexin 125mg/5ml oral suspension | cephalosporins |
| 29748 | Cefalexin 125mg/5ml oral suspension (A A H Pharmaceuticals Ltd) | cephalosporins |
| 32181 | Cefalexin 125mg/5ml oral suspension (Actavis UK Ltd) | cephalosporins |
| 53945 | Cefalexin 125mg/5ml oral suspension (Alliance Healthcare (Distribution) Ltd) | cephalosporins |
| 68521 | Cefalexin 125mg/5ml oral suspension (Arrow Generics Ltd) | cephalosporins |
| 74060 | Cefalexin 125mg/5ml oral suspension (Dowelhurst Ltd) | cephalosporins |
| 32642 | Cefalexin 125mg/5ml oral suspension (Kent Pharmaceuticals Ltd) | cephalosporins |
| 39417 | Cefalexin 125mg/5ml oral suspension (Mylan) | cephalosporins |
| 77752 | Cefalexin 125mg/5ml oral suspension (Phoenix Healthcare Distribution Ltd) | cephalosporins |
| 36578 | Cefalexin 125mg/5ml oral suspension (Ranbaxy (UK) Ltd) | cephalosporins |
| 33329 | Cefalexin 125mg/5ml oral suspension (Teva UK Ltd) | cephalosporins |
| 61661 | Cefalexin 125mg/5ml oral suspension (Waymade Healthcare Plc) | cephalosporins |
| 6651 | Cefalexin 125mg/5ml oral suspension sugar free | cephalosporins |
| 74893 | Cefalexin 125mg/5ml oral suspension sugar free (Alliance Healthcare (Distribution) Ltd) | cephalosporins |
| 72224 | Cefalexin 125mg/5ml oral suspension sugar free (Milpharm Ltd) | cephalosporins |
| 62898 | Cefalexin 125mg/5ml oral suspension sugar free (PLIVA Pharma Ltd) | cephalosporins |
| 19144 | Cefalexin 125mg/5ml oral suspension sugar free (Teva UK Ltd) | cephalosporins |
| 1384 | Cefalexin 125mg/5ml suspension | cephalosporins |
| 18451 | Cefalexin 1g tablets | cephalosporins |
| 33802 | Cefalexin 250mg Capsule (Berk Pharmaceuticals Ltd) | cephalosporins |
| 155 | Cefalexin 250mg capsules | cephalosporins |
| 34253 | Cefalexin 250mg capsules (A A H Pharmaceuticals Ltd) | cephalosporins |
| 19152 | Cefalexin 250mg capsules (Actavis UK Ltd) | cephalosporins |
| 54864 | Cefalexin 250mg capsules (Alliance Healthcare (Distribution) Ltd) | cephalosporins |
| 52283 | Cefalexin 250mg capsules (Arrow Generics Ltd) | cephalosporins |
| 59269 | Cefalexin 250mg capsules (DE Pharmaceuticals) | cephalosporins |
| 19133 | Cefalexin 250mg capsules (IVAX Pharmaceuticals UK Ltd) | cephalosporins |
| 41736 | Cefalexin 250mg capsules (Kent Pharmaceuticals Ltd) | cephalosporins |
| 75783 | Cefalexin 250mg capsules (Lupin Healthcare (UK) Ltd) | cephalosporins |
| 65953 | Cefalexin 250mg capsules (Mawdsley-Brooks & Company Ltd) | cephalosporins |
| 52282 | Cefalexin 250mg capsules (Milpharm Ltd) | cephalosporins |
| 19160 | Cefalexin 250mg capsules (Mylan) | cephalosporins |
| 62188 | Cefalexin 250mg capsules (Phoenix Healthcare Distribution Ltd) | cephalosporins |
| 24090 | Cefalexin 250mg capsules (PLIVA Pharma Ltd) | cephalosporins |
| 36599 | Cefalexin 250mg capsules (Ranbaxy (UK) Ltd) | cephalosporins |
| 69107 | Cefalexin 250mg capsules (Sigma Pharmaceuticals Plc) | cephalosporins |
| 9690 | Cefalexin 250mg capsules (Teva UK Ltd) | cephalosporins |
| 60039 | Cefalexin 250mg capsules (Waymade Healthcare Plc) | cephalosporins |
| 40747 | Cefalexin 250mg chewable tablets | cephalosporins |
| 1146 | Cefalexin 250mg tablets | cephalosporins |
| 33334 | Cefalexin 250mg tablets (A A H Pharmaceuticals Ltd) | cephalosporins |
| 36330 | Cefalexin 250mg tablets (Actavis UK Ltd) | cephalosporins |
| 47163 | Cefalexin 250mg tablets (Arrow Generics Ltd) | cephalosporins |
| 31825 | Cefalexin 250mg tablets (IVAX Pharmaceuticals UK Ltd) | cephalosporins |
| 36701 | Cefalexin 250mg tablets (Mylan) | cephalosporins |
| 59069 | Cefalexin 250mg tablets (Phoenix Healthcare Distribution Ltd) | cephalosporins |
| 67749 | Cefalexin 250mg tablets (PLIVA Pharma Ltd) | cephalosporins |
| 74637 | Cefalexin 250mg tablets (Ranbaxy (UK) Ltd) | cephalosporins |
| 9698 | Cefalexin 250mg tablets (Teva UK Ltd) | cephalosporins |
| 41825 | Cefalexin 250mg/5ml Oral solution (C P Pharmaceuticals Ltd) | cephalosporins |
| 75579 | Cefalexin 250mg/5ml Oral solution (Lagap) | cephalosporins |
| 1860 | Cefalexin 250mg/5ml oral suspension | cephalosporins |
| 42008 | Cefalexin 250mg/5ml oral suspension (A A H Pharmaceuticals Ltd) | cephalosporins |
| 45221 | Cefalexin 250mg/5ml oral suspension (Actavis UK Ltd) | cephalosporins |
| 67794 | Cefalexin 250mg/5ml oral suspension (Alliance Healthcare (Distribution) Ltd) | cephalosporins |
| 60202 | Cefalexin 250mg/5ml oral suspension (Kent Pharmaceuticals Ltd) | cephalosporins |
| 65072 | Cefalexin 250mg/5ml oral suspension (Mawdsley-Brooks & Company Ltd) | cephalosporins |
| 29464 | Cefalexin 250mg/5ml oral suspension (Mylan) | cephalosporins |
| 41192 | Cefalexin 250mg/5ml oral suspension (Ranbaxy (UK) Ltd) | cephalosporins |
| 41968 | Cefalexin 250mg/5ml oral suspension (Teva UK Ltd) | cephalosporins |
| 6671 | Cefalexin 250mg/5ml oral suspension sugar free | cephalosporins |
| 75386 | Cefalexin 250mg/5ml oral suspension sugar free (Alliance Healthcare (Distribution) Ltd) | cephalosporins |
| 34133 | Cefalexin 250mg/5ml oral suspension sugar free (Teva UK Ltd) | cephalosporins |
| 1713 | Cefalexin 250mg/5ml suspension | cephalosporins |
| 44755 | Cefalexin 500mg Capsule (Berk Pharmaceuticals Ltd) | cephalosporins |
| 400 | Cefalexin 500mg capsules | cephalosporins |
| 32643 | Cefalexin 500mg capsules (A A H Pharmaceuticals Ltd) | cephalosporins |
| 19138 | Cefalexin 500mg capsules (Actavis UK Ltd) | cephalosporins |
| 52851 | Cefalexin 500mg capsules (Alliance Healthcare (Distribution) Ltd) | cephalosporins |
| 77916 | Cefalexin 500mg capsules (DE Pharmaceuticals) | cephalosporins |
| 9664 | Cefalexin 500mg capsules (IVAX Pharmaceuticals UK Ltd) | cephalosporins |
| 36569 | Cefalexin 500mg capsules (Kent Pharmaceuticals Ltd) | cephalosporins |
| 69472 | Cefalexin 500mg capsules (Lupin Healthcare (UK) Ltd) | cephalosporins |
| 70402 | Cefalexin 500mg capsules (Mawdsley-Brooks & Company Ltd) | cephalosporins |
| 54955 | Cefalexin 500mg capsules (Milpharm Ltd) | cephalosporins |
| 19184 | Cefalexin 500mg capsules (Mylan) | cephalosporins |
| 67828 | Cefalexin 500mg capsules (Phoenix Healthcare Distribution Ltd) | cephalosporins |
| 19161 | Cefalexin 500mg capsules (Ranbaxy (UK) Ltd) | cephalosporins |
| 74505 | Cefalexin 500mg capsules (Sigma Pharmaceuticals Plc) | cephalosporins |
| 29281 | Cefalexin 500mg capsules (Teva UK Ltd) | cephalosporins |
| 66179 | Cefalexin 500mg capsules (Waymade Healthcare Plc) | cephalosporins |
| 77168 | Cefalexin 500mg Tablet (Berk Pharmaceuticals Ltd) | cephalosporins |
| 865 | Cefalexin 500mg tablets | cephalosporins |
| 29202 | Cefalexin 500mg tablets (A A H Pharmaceuticals Ltd) | cephalosporins |
| 31827 | Cefalexin 500mg tablets (IVAX Pharmaceuticals UK Ltd) | cephalosporins |
| 22321 | Cefalexin 500mg tablets (Mylan) | cephalosporins |
| 67746 | Cefalexin 500mg tablets (PLIVA Pharma Ltd) | cephalosporins |
| 9689 | Cefalexin 500mg tablets (Teva UK Ltd) | cephalosporins |
| 59406 | Cefalexin 500mg tablets (Waymade Healthcare Plc) | cephalosporins |
| 2227 | Cefalexin 500mg/5ml oral suspension | cephalosporins |
| 66349 | Cefalexin 500mg/5ml oral suspension (Waymade Healthcare Plc) | cephalosporins |
| 1110 | Cefixime 100mg/5ml oral suspension | cephalosporins |
| 843 | Cefixime 200mg tablets | cephalosporins |
| 70888 | Cefixime 400mg tablets | cephalosporins |
| 12065 | Cefpodoxime 100mg tablets | cephalosporins |
| 7510 | Cefpodoxime 40mg/5ml oral suspension | cephalosporins |
| 565 | Cefradine 250mg capsules | cephalosporins |
| 34543 | Cefradine 250mg capsules (A A H Pharmaceuticals Ltd) | cephalosporins |
| 72356 | Cefradine 250mg capsules (Alliance Healthcare (Distribution) Ltd) | cephalosporins |
| 30769 | Cefradine 250mg capsules (IVAX Pharmaceuticals UK Ltd) | cephalosporins |
| 44299 | Cefradine 250mg capsules (Kent Pharmaceuticals Ltd) | cephalosporins |
| 30789 | Cefradine 250mg capsules (Mylan) | cephalosporins |
| 71055 | Cefradine 250mg capsules (Phoenix Healthcare Distribution Ltd) | cephalosporins |
| 43433 | Cefradine 250mg capsules (Teva UK Ltd) | cephalosporins |
| 77524 | Cefradine 250mg capsules (Waymade Healthcare Plc) | cephalosporins |
| 5410 | Cefradine 250mg/5ml oral solution | cephalosporins |
| 4454 | Cefradine 500mg capsules | cephalosporins |
| 34965 | Cefradine 500mg capsules (A A H Pharmaceuticals Ltd) | cephalosporins |
| 34844 | Cefradine 500mg capsules (IVAX Pharmaceuticals UK Ltd) | cephalosporins |
| 67743 | Cefradine 500mg capsules (Kent Pharmaceuticals Ltd) | cephalosporins |
| 34629 | Cefradine 500mg capsules (Mylan) | cephalosporins |
| 34654 | Cefradine 500mg capsules (Teva UK Ltd) | cephalosporins |
| 17881 | Ceftibuten 180mg/5ml suspension | cephalosporins |
| 7509 | Ceftibuten 400mg capsules | cephalosporins |
| 17889 | Ceftibuten 90mg/5ml suspension | cephalosporins |
| 47993 | Cefuroxime (as axetil) 500mg tablets | cephalosporins |
| 55387 | Cefuroxime 125mg granules sachets | cephalosporins |
| 12850 | Cefuroxime 125mg tablets | cephalosporins |
| 725 | Cefuroxime 125mg/5ml oral suspension | cephalosporins |
| 3678 | Cefuroxime 250mg tablets | cephalosporins |
| 47983 | Cefuroxime 250mg tablets (A A H Pharmaceuticals Ltd) | cephalosporins |
| 58623 | Cefuroxime 250mg tablets (Alliance Healthcare (Distribution) Ltd) | cephalosporins |
| 77667 | Cefuroxime 250mg tablets (DE Pharmaceuticals) | cephalosporins |
| 58463 | Cefuroxime 250mg tablets (Sandoz Ltd) | cephalosporins |
| 59068 | Cefuroxime 250mg tablets (Teva UK Ltd) | cephalosporins |
| 58920 | Cefuroxime 250mg tablets (Tillomed Laboratories Ltd) | cephalosporins |
| 61268 | Cefuroxime 250mg tablets (Waymade Healthcare Plc) | cephalosporins |
| 7560 | Ceporex 125mg/5ml Liquid (Galen Ltd) | cephalosporins |
| 3609 | Ceporex 125mg/5ml Oral solution (Galen Ltd) | cephalosporins |
| 41106 | Ceporex 125mg/5ml syrup (Strides Pharma UK Ltd) | cephalosporins |
| 12235 | Ceporex 1g Tablet (Galen Ltd) | cephalosporins |
| 192 | Ceporex 250mg Capsule (Galen Ltd) | cephalosporins |
| 40884 | Ceporex 250mg capsules (Strides Pharma UK Ltd) | cephalosporins |
| 8019 | Ceporex 250mg Tablet (Galen Ltd) | cephalosporins |
| 41049 | Ceporex 250mg tablets (Strides Pharma UK Ltd) | cephalosporins |
| 8625 | Ceporex 250mg/5ml Liquid (Galen Ltd) | cephalosporins |
| 8008 | Ceporex 250mg/5ml Oral solution (Galen Ltd) | cephalosporins |
| 40945 | Ceporex 250mg/5ml syrup (Strides Pharma UK Ltd) | cephalosporins |
| 2661 | Ceporex 500mg Capsule (Galen Ltd) | cephalosporins |
| 40915 | Ceporex 500mg capsules (Strides Pharma UK Ltd) | cephalosporins |
| 8085 | Ceporex 500mg Tablet (Galen Ltd) | cephalosporins |
| 40914 | Ceporex 500mg tablets (Strides Pharma UK Ltd) | cephalosporins |
| 5859 | Ceporex 500mg/5ml Oral solution (Galen Ltd) | cephalosporins |
| 41230 | Ceporex 500mg/5ml syrup (Strides Pharma UK Ltd) | cephalosporins |
| 2428 | Distaclor 125mg/5ml Liquid (Dista Products Ltd) | cephalosporins |
| 25384 | Distaclor 125mg/5ml oral suspension (Flynn Pharma Ltd) | cephalosporins |
| 4576 | Distaclor 250mg Capsule (Dista Products Ltd) | cephalosporins |
| 9219 | Distaclor 250mg/5ml Liquid (Dista Products Ltd) | cephalosporins |
| 22042 | Distaclor 250mg/5ml oral suspension (Flynn Pharma Ltd) | cephalosporins |
| 7889 | Distaclor 375mg Modified-release tablet (Dista Products Ltd) | cephalosporins |
| 319 | Distaclor 500mg Capsule (Dista Products Ltd) | cephalosporins |
| 18243 | Distaclor 500mg capsules (Flynn Pharma Ltd) | cephalosporins |
| 3523 | Distaclor 500mg Modified-release tablet (Dista Products Ltd) | cephalosporins |
| 20992 | Distaclor MR 375mg tablets (Flynn Pharma Ltd) | cephalosporins |
| 7485 | Keflex 125mg/5ml Liquid (Eli Lilly and Company Ltd) | cephalosporins |
| 27072 | Keflex 125mg/5ml oral suspension (Flynn Pharma Ltd) | cephalosporins |
| 7430 | Keflex 250mg Capsule (Eli Lilly and Company Ltd) | cephalosporins |
| 11989 | Keflex 250mg capsules (Flynn Pharma Ltd) | cephalosporins |
| 9157 | Keflex 250mg Tablet (Eli Lilly and Company Ltd) | cephalosporins |
| 830 | Keflex 250mg tablets (Flynn Pharma Ltd) | cephalosporins |
| 10455 | Keflex 250mg/5ml Liquid (Eli Lilly and Company Ltd) | cephalosporins |
| 28722 | Keflex 250mg/5ml oral suspension (Flynn Pharma Ltd) | cephalosporins |
| 12276 | Keflex 500mg Capsule (Eli Lilly and Company Ltd) | cephalosporins |
| 24618 | Keflex 500mg capsules (Flynn Pharma Ltd) | cephalosporins |
| 9603 | Keflex 500mg Tablet (Eli Lilly and Company Ltd) | cephalosporins |
| 31110 | Keflex 500mg tablets (Flynn Pharma Ltd) | cephalosporins |
| 26233 | Keftid 125mg/5ml oral suspension (Strides Pharma UK Ltd) | cephalosporins |
| 26207 | Keftid 250mg capsules (Strides Pharma UK Ltd) | cephalosporins |
| 41853 | Keftid 250mg/5ml oral suspension (Strides Pharma UK Ltd) | cephalosporins |
| 26236 | Keftid 500mg capsules (Strides Pharma UK Ltd) | cephalosporins |
| 26989 | Kiflone 125mg/5ml Oral solution (Berk Pharmaceuticals Ltd) | cephalosporins |
| 21835 | Kiflone 250mg Capsule (Berk Pharmaceuticals Ltd) | cephalosporins |
| 21979 | Kiflone 250mg/5ml Oral solution (Berk Pharmaceuticals Ltd) | cephalosporins |
| 27017 | Kiflone 500mg Capsule (Berk Pharmaceuticals Ltd) | cephalosporins |
| 26992 | Kiflone 500mg Tablet (Berk Pharmaceuticals Ltd) | cephalosporins |
| 30610 | Nicef 250mg capsules (Strides Pharma UK Ltd) | cephalosporins |
| 33605 | Nicef 500mg capsules (Strides Pharma UK Ltd) | cephalosporins |
| 18702 | Orelox 100mg tablets (Sanofi) | cephalosporins |
| 17129 | Orelox 40mg/5ml oral suspension paediatric (Sanofi) | cephalosporins |
| 1306 | Suprax 200mg tablets (Sanofi) | cephalosporins |
| 3831 | Suprax Paediatric 100mg/5ml oral suspension (Sanofi) | cephalosporins |
| 27254 | Tenkorex 500mg Capsule (OPD Pharm) | cephalosporins |
| 7624 | Velosef 250mg capsules (Bristol-Myers Squibb Pharmaceuticals Ltd) | cephalosporins |
| 9647 | Velosef 250mg/5ml syrup (Bristol-Myers Squibb Pharmaceuticals Ltd) | cephalosporins |
| 7909 | Velosef 500mg capsules (Bristol-Myers Squibb Pharmaceuticals Ltd) | cephalosporins |
| 10794 | Zinnat 125mg tablets (GlaxoSmithKline UK Ltd) | cephalosporins |
| 13412 | Zinnat 125mg/5ml oral suspension (GlaxoSmithKline UK Ltd) | cephalosporins |
| 3846 | Zinnat 250mg tablets (GlaxoSmithKline UK Ltd) | cephalosporins |
| 73065 | Zinnat 250mg tablets (Lexon (UK) Ltd) | cephalosporins |
| 79740 | Clindamycin 100mg/5ml oral suspension | clindamycin |
| 64734 | Clindamycin 120mg/5ml oral solution | clindamycin |
| 615 | Clindamycin 150mg capsules | clindamycin |
| 32843 | Clindamycin 150mg capsules (A A H Pharmaceuticals Ltd) | clindamycin |
| 71047 | Clindamycin 150mg capsules (Creo Pharma Ltd) | clindamycin |
| 76667 | Clindamycin 150mg capsules (Mylan) | clindamycin |
| 72647 | Clindamycin 150mg capsules (Phoenix Healthcare Distribution Ltd) | clindamycin |
| 34940 | Clindamycin 150mg capsules (Sandoz Ltd) | clindamycin |
| 73278 | Clindamycin 150mg capsules (Waymade Healthcare Plc) | clindamycin |
| 50778 | Clindamycin 150mg/5ml oral solution | clindamycin |
| 76887 | Clindamycin 200mg/5ml oral suspension | clindamycin |
| 78821 | Clindamycin 20mg/5ml oral solution | clindamycin |
| 54447 | Clindamycin 300mg capsules | clindamycin |
| 76862 | Clindamycin 300mg capsules (Actavis UK Ltd) | clindamycin |
| 65415 | Clindamycin 300mg capsules (Creo Pharma Ltd) | clindamycin |
| 73457 | Clindamycin 300mg capsules (DE Pharmaceuticals) | clindamycin |
| 74262 | Clindamycin 300mg capsules (Rivopharm (UK) Ltd) | clindamycin |
| 67532 | Clindamycin 300mg capsules (Sigma Pharmaceuticals Plc) | clindamycin |
| 78853 | Clindamycin 300mg capsules (Teva UK Ltd) | clindamycin |
| 35538 | Clindamycin 300mg/2ml solution for injection ampoules | clindamycin |
| 62293 | Clindamycin 300mg/2ml solution for injection ampoules (A A H Pharmaceuticals Ltd) | clindamycin |
| 60754 | Clindamycin 300mg/5ml oral solution | clindamycin |
| 38397 | Clindamycin 600mg/4ml solution for injection ampoules | clindamycin |
| 59813 | Clindamycin 60mg/5ml oral solution | clindamycin |
| 207 | Clindamycin 75mg capsules | clindamycin |
| 77833 | Clindamycin 75mg capsules (Alliance Healthcare (Distribution) Ltd) | clindamycin |
| 50779 | Clindamycin 75mg/5ml oral solution | clindamycin |
| 60265 | Clindamycin 75mg/5ml oral suspension | clindamycin |
| 23518 | Clindamycin HCl 150mg/ml injection | clindamycin |
| 10987 | Clindamycin HCl 75mg/5ml granules for suspension | clindamycin |
| 45222 | Clindamycin oral liquid | clindamycin |
| 677 | Clindamycin phosphate 150mg/ml injection | clindamycin |
| 59579 | Dalacin C 150mg capsules (DE Pharmaceuticals) | clindamycin |
| 78781 | Dalacin C 150mg capsules (Mawdsley-Brooks & Company Ltd) | clindamycin |
| 4137 | Dalacin C 150mg capsules (Pfizer Ltd) | clindamycin |
| 79448 | Dalacin C 150mg capsules (Sigma Pharmaceuticals Plc) | clindamycin |
| 4509 | Dalacin C 75mg capsules (Pfizer Ltd) | clindamycin |
| 10462 | Dalacin c 75mg/5ml Granules (Pharmacia Ltd) | clindamycin |
| 9165 | Dalacin c phosphate 150mg/ml Injection (Pharmacia Ltd) | clindamycin |
| 43041 | Dalacin C Phosphate 600mg/4ml solution for injection ampoules (Pfizer Ltd) | clindamycin |
| 32705 | LINCOCIN | clindamycin |
| 44407 | Lincocin 300mg/ml Injection (Pharmacia Ltd) | clindamycin |
| 27722 | Lincocin 500mg Capsule (Pharmacia Ltd) | clindamycin |
| 28002 | Lincomycin hcl 250mg/5ml Oral solution | clindamycin |
| 32428 | Lincomycin hcl 300mg/ml Injection | clindamycin |
| 18235 | Lincomycin hcl 500mg Capsule | clindamycin |
| 10326 | Clarithromycin 125mg granules straws | macrolides |
| 26059 | Clarithromycin 187.5mg granules straws | macrolides |
| 17645 | Clarithromycin 250mg granules straws | macrolides |
| 54882 | Clarithromycin 250mg tablets (Almus Pharmaceuticals Ltd) | macrolides |
| 53715 | Clarithromycin 500mg tablets (Almus Pharmaceuticals Ltd) | macrolides |
| 28349 | Clarosip 125mg granules for oral suspension straws (Grunenthal Ltd) | macrolides |
| 31689 | Clarosip 187.5mg granules for oral suspension straws (Grunenthal Ltd) | macrolides |
| 31690 | Clarosip 250mg granules for oral suspension straws (Grunenthal Ltd) | macrolides |
| 18703 | ERYCEN 125 MG SUS | macrolides |
| 18652 | ERYCEN 250 MG SUS | macrolides |
| 1376 | ERYTHROMYCIN 100 MG SYR | macrolides |
| 7792 | ERYTHROMYCIN 12 MG SYR | macrolides |
| 1969 | ERYTHROMYCIN 250 MG MIX | macrolides |
| 103 | Erythromycin 250mg gastro-resistant capsules | macrolides |
| 251 | ERYTHROMYCIN 50 MG INJ | macrolides |
| 3408 | ERYTHROMYCIN 500 MG CAP | macrolides |
| 1037 | ERYTHROMYCIN ETHYLSUCCINATE SF 125 MG/5ML SUS | macrolides |
| 16605 | ERYTHROMYCIN I/V 1 GM INJ | macrolides |
| 25943 | ERYTHROMYCIN I/V 300 MG INJ | macrolides |
| 3907 | ERYTHROMYCIN SF sach 250 MG | macrolides |
| 26669 | ERYMAX | macrolides |
| 10782 | ERYMAX 500 MG CAP | macrolides |
| 12252 | ERYMAX 125 MG SYR | macrolides |
| 14557 | ERYMIN 250 MG/5ML SUS | macrolides |
| 27955 | ERYTHROCIN 300 MG INJ | macrolides |
| 22653 | ERYTHROCIN 100 MG SYR | macrolides |
| 17798 | ERYTHROCIN A 1 GM TAB | macrolides |
| 2304 | ERYTHROCIN 250 250 MG TAB | macrolides |
| 22632 | ERYTHROCIN B-PACK 10 FILMTABS 500 MG TAB | macrolides |
| 19692 | ERYTHROCIN 250 | macrolides |
| 2019 | ERYTHROCIN 125 MG SYR | macrolides |
| 19615 | ERYTHROPED FORTE 500mg/5ml | macrolides |
| 23185 | ERYTHROPED SUGAR FREE SACHET | macrolides |
| 2313 | ERYTHROPED 250 MG TAB | macrolides |
| 19652 | ERYTHROPED | macrolides |
| 23243 | ERYTHROPED P.I. | macrolides |
| 32427 | KLARICID IV 500MG VIAL DRY 500 MG INJ | macrolides |
| 28730 | ROVAMYCIN 250 MG TAB | macrolides |
| 21658 | SPIRAMYCIN 250 MG TAB | macrolides |
| 32669 | ZITHROMAX | macrolides |
| 5662 | Amoxicillin 500mg / Clarithromycin 500mg / Lansoprazole 30mg triple pack | macrolides |
| 11433 | Clarithromycin 500mg with lansoprazole 30mg and amoxicillin 500mg triple pack | macrolides |
| 15290 | Lansoprazole with amoxicillin and clarithromycin 30mg + 500mg + 500mg Triple pack | macrolides |
| 6497 | Clarithromycin 500mg with metronidazole 400mg with lansoprazole 30mg triple pack | macrolides |
| 27495 | Arpimycin 125mg/5ml Liquid (Rosemont Pharmaceuticals Ltd) | macrolides |
| 36544 | Arpimycin 125mg/5ml Oral suspension (Rosemont Pharmaceuticals Ltd) | macrolides |
| 24220 | Arpimycin 250mg/5ml Liquid (Rosemont Pharmaceuticals Ltd) | macrolides |
| 36514 | Arpimycin 250mg/5ml Oral suspension (Rosemont Pharmaceuticals Ltd) | macrolides |
| 37022 | Arpimycin 500mg/5ml Liquid (Rosemont Pharmaceuticals Ltd) | macrolides |
| 5057 | Azithromycin 200mg/5ml oral suspension | macrolides |
| 53850 | Azithromycin 200mg/5ml oral suspension (A A H Pharmaceuticals Ltd) | macrolides |
| 65486 | Azithromycin 200mg/5ml oral suspension (Alliance Healthcare (Distribution) Ltd) | macrolides |
| 49530 | Azithromycin 200mg/5ml oral suspension (Sandoz Ltd) | macrolides |
| 76263 | Azithromycin 200mg/5ml oral suspension (Waymade Healthcare Plc) | macrolides |
| 65855 | Azithromycin 200mg/5ml oral suspension sugar free | macrolides |
| 5116 | Azithromycin 250mg capsules | macrolides |
| 62785 | Azithromycin 250mg capsules (A A H Pharmaceuticals Ltd) | macrolides |
| 79270 | Azithromycin 250mg capsules (Creo Pharma Ltd) | macrolides |
| 60382 | Azithromycin 250mg capsules (DE Pharmaceuticals) | macrolides |
| 69613 | Azithromycin 250mg capsules (Teva UK Ltd) | macrolides |
| 33888 | Azithromycin 250mg tablets | macrolides |
| 74293 | Azithromycin 250mg tablets (Alliance Healthcare (Distribution) Ltd) | macrolides |
| 66034 | Azithromycin 250mg tablets (Sandoz Ltd) | macrolides |
| 58206 | Azithromycin 250mg tablets (Teva UK Ltd) | macrolides |
| 76155 | Azithromycin 500mg powder for solution for infusion vials | macrolides |
| 46695 | Azithromycin 500mg Tablet (Hillcross Pharmaceuticals Ltd) | macrolides |
| 743 | Azithromycin 500mg tablets | macrolides |
| 61810 | Azithromycin 500mg tablets (A A H Pharmaceuticals Ltd) | macrolides |
| 74523 | Azithromycin 500mg tablets (Aspire Pharma Ltd) | macrolides |
| 60814 | Azithromycin 500mg tablets (DE Pharmaceuticals) | macrolides |
| 70783 | Azithromycin 500mg tablets (Kent Pharmaceuticals Ltd) | macrolides |
| 58426 | Azithromycin 500mg tablets (Sandoz Ltd) | macrolides |
| 40218 | Azithromycin 500mg tablets (Teva UK Ltd) | macrolides |
| 43400 | Clamelle 500mg tablets (Actavis UK Ltd) | macrolides |
| 45591 | Clarie XL 500mg tablets (Teva UK Ltd) | macrolides |
| 331 | Clarithromycin 125mg/5ml oral suspension | macrolides |
| 45795 | Clarithromycin 125mg/5ml oral suspension (A A H Pharmaceuticals Ltd) | macrolides |
| 79562 | Clarithromycin 125mg/5ml oral suspension (Actavis UK Ltd) | macrolides |
| 54903 | Clarithromycin 125mg/5ml oral suspension (Alliance Healthcare (Distribution) Ltd) | macrolides |
| 72126 | Clarithromycin 125mg/5ml oral suspension (Almus Pharmaceuticals Ltd) | macrolides |
| 68723 | Clarithromycin 125mg/5ml oral suspension (DE Pharmaceuticals) | macrolides |
| 61001 | Clarithromycin 125mg/5ml oral suspension (Kent Pharmaceuticals Ltd) | macrolides |
| 51831 | Clarithromycin 125mg/5ml oral suspension (Phoenix Healthcare Distribution Ltd) | macrolides |
| 41453 | Clarithromycin 125mg/5ml oral suspension (Ranbaxy (UK) Ltd) | macrolides |
| 53168 | Clarithromycin 125mg/5ml oral suspension (Sandoz Ltd) | macrolides |
| 61830 | Clarithromycin 125mg/5ml oral suspension (Sigma Pharmaceuticals Plc) | macrolides |
| 57267 | Clarithromycin 125mg/5ml oral suspension (Waymade Healthcare Plc) | macrolides |
| 765 | Clarithromycin 250mg granules sachets | macrolides |
| 537 | Clarithromycin 250mg tablets | macrolides |
| 34650 | Clarithromycin 250mg tablets (A A H Pharmaceuticals Ltd) | macrolides |
| 54472 | Clarithromycin 250mg tablets (Accord Healthcare Ltd) | macrolides |
| 48163 | Clarithromycin 250mg tablets (Actavis UK Ltd) | macrolides |
| 52158 | Clarithromycin 250mg tablets (Alliance Healthcare (Distribution) Ltd) | macrolides |
| 57660 | Clarithromycin 250mg tablets (Almus Pharmaceuticals Ltd) | macrolides |
| 52719 | Clarithromycin 250mg tablets (Apotex UK Ltd) | macrolides |
| 53086 | Clarithromycin 250mg tablets (DE Pharmaceuticals) | macrolides |
| 51154 | Clarithromycin 250mg tablets (Kent Pharmaceuticals Ltd) | macrolides |
| 74616 | Clarithromycin 250mg tablets (Milpharm Ltd) | macrolides |
| 34394 | Clarithromycin 250mg tablets (Mylan) | macrolides |
| 53153 | Clarithromycin 250mg tablets (Phoenix Healthcare Distribution Ltd) | macrolides |
| 53688 | Clarithromycin 250mg tablets (Ranbaxy (UK) Ltd) | macrolides |
| 47582 | Clarithromycin 250mg tablets (Sandoz Ltd) | macrolides |
| 50946 | Clarithromycin 250mg tablets (Sigma Pharmaceuticals Plc) | macrolides |
| 54269 | Clarithromycin 250mg tablets (Somex Pharma) | macrolides |
| 34533 | Clarithromycin 250mg tablets (Teva UK Ltd) | macrolides |
| 54897 | Clarithromycin 250mg tablets (Tillomed Laboratories Ltd) | macrolides |
| 78328 | Clarithromycin 250mg tablets (Torrent Pharma (UK) Ltd) | macrolides |
| 63033 | Clarithromycin 250mg tablets (Waymade Healthcare Plc) | macrolides |
| 53144 | Clarithromycin 250mg tablets (Wockhardt UK Ltd) | macrolides |
| 5357 | Clarithromycin 250mg/5ml oral suspension | macrolides |
| 54241 | Clarithromycin 250mg/5ml oral suspension (A A H Pharmaceuticals Ltd) | macrolides |
| 55148 | Clarithromycin 250mg/5ml oral suspension (Alliance Healthcare (Distribution) Ltd) | macrolides |
| 67560 | Clarithromycin 250mg/5ml oral suspension (Almus Pharmaceuticals Ltd) | macrolides |
| 71680 | Clarithromycin 250mg/5ml oral suspension (DE Pharmaceuticals) | macrolides |
| 62897 | Clarithromycin 250mg/5ml oral suspension (Phoenix Healthcare Distribution Ltd) | macrolides |
| 34811 | Clarithromycin 250mg/5ml oral suspension (Ranbaxy (UK) Ltd) | macrolides |
| 53179 | Clarithromycin 250mg/5ml oral suspension (Sandoz Ltd) | macrolides |
| 54208 | Clarithromycin 250mg/5ml oral suspension (Sigma Pharmaceuticals Plc) | macrolides |
| 55428 | Clarithromycin 250mg/5ml oral suspension (Waymade Healthcare Plc) | macrolides |
| 54529 | Clarithromycin 500mg Modified-release tablet (Hillcross Pharmaceuticals Ltd) | macrolides |
| 6803 | Clarithromycin 500mg modified-release tablets | macrolides |
| 77727 | Clarithromycin 500mg powder for concentrate for solution for infusion vials (Alliance Healthcare (Distribution) Ltd) | macrolides |
| 75871 | Clarithromycin 500mg powder for concentrate for solution for infusion vials (Hameln Pharmaceuticals Ltd) | macrolides |
| 54953 | Clarithromycin 500mg powder for concentrate for solution for infusion vials (Martindale Pharmaceuticals Ltd) | macrolides |
| 53398 | Clarithromycin 500mg powder for concentrate for solution for infusion vials (Mercury Pharma Group Ltd) | macrolides |
| 13323 | Clarithromycin 500mg powder for solution for infusion vials | macrolides |
| 66092 | Clarithromycin 500mg powder for solution for infusion vials (A A H Pharmaceuticals Ltd) | macrolides |
| 72108 | Clarithromycin 500mg powder for solution for infusion vials (Bowmed Ibisqus Ltd) | macrolides |
| 78226 | Clarithromycin 500mg powder for solution for infusion vials (Teva UK Ltd) | macrolides |
| 681 | Clarithromycin 500mg tablets | macrolides |
| 38163 | Clarithromycin 500mg tablets (A A H Pharmaceuticals Ltd) | macrolides |
| 51426 | Clarithromycin 500mg tablets (Accord Healthcare Ltd) | macrolides |
| 48023 | Clarithromycin 500mg tablets (Actavis UK Ltd) | macrolides |
| 49939 | Clarithromycin 500mg tablets (Alliance Healthcare (Distribution) Ltd) | macrolides |
| 58037 | Clarithromycin 500mg tablets (Almus Pharmaceuticals Ltd) | macrolides |
| 53776 | Clarithromycin 500mg tablets (DE Pharmaceuticals) | macrolides |
| 53703 | Clarithromycin 500mg tablets (Kent Pharmaceuticals Ltd) | macrolides |
| 72318 | Clarithromycin 500mg tablets (Milpharm Ltd) | macrolides |
| 34608 | Clarithromycin 500mg tablets (Mylan) | macrolides |
| 58902 | Clarithromycin 500mg tablets (Phoenix Healthcare Distribution Ltd) | macrolides |
| 46488 | Clarithromycin 500mg tablets (Ranbaxy (UK) Ltd) | macrolides |
| 40784 | Clarithromycin 500mg tablets (Sandoz Ltd) | macrolides |
| 68943 | Clarithromycin 500mg tablets (Sigma Pharmaceuticals Plc) | macrolides |
| 53109 | Clarithromycin 500mg tablets (Somex Pharma) | macrolides |
| 34974 | Clarithromycin 500mg tablets (Teva UK Ltd) | macrolides |
| 53875 | Clarithromycin 500mg tablets (Tillomed Laboratories Ltd) | macrolides |
| 60805 | Clarithromycin 500mg tablets (Waymade Healthcare Plc) | macrolides |
| 58175 | Clarithromycin 500mg tablets (Wockhardt UK Ltd) | macrolides |
| 70218 | Dificlir 200mg tablets (Astellas Pharma Ltd) | macrolides |
| 14511 | Erymax sprinkle 125mg Capsule (Elan Pharma) | macrolides |
| 9434 | Erymin 250mg/5ml Oral suspension (Elan Pharma) | macrolides |
| 48017 | Erythoden 125mg/5ml Liquid (Stevenden Healthcare) | macrolides |
| 41389 | Erythoden 250mg/5ml Liquid (Stevenden Healthcare) | macrolides |
| 201 | Erythrocin 1g/vial Injection (Abbott Laboratories Ltd) | macrolides |
| 39616 | Erythrocin 250 tablets (Advanz Pharma) | macrolides |
| 480 | Erythrocin 250mg Tablet (Abbott Laboratories Ltd) | macrolides |
| 1072 | Erythrocin 500 500mg Tablet (Abbott Laboratories Ltd) | macrolides |
| 39613 | Erythrocin 500 tablets (Advanz Pharma) | macrolides |
| 71363 | Erythrocin 500 tablets (Dowelhurst Ltd) | macrolides |
| 53449 | Erythrocin 500 tablets (Lexon (UK) Ltd) | macrolides |
| 51984 | Erythrocin 500 tablets (Mawdsley-Brooks & Company Ltd) | macrolides |
| 53004 | Erythrocin 500 tablets (Necessity Supplies Ltd) | macrolides |
| 50693 | Erythrocin 500 tablets (Sigma Pharmaceuticals Plc) | macrolides |
| 50223 | Erythrocin 500 tablets (Stephar (U.K.) Ltd) | macrolides |
| 66819 | Erythrocin 500 tablets (Waymade Healthcare Plc) | macrolides |
| 37681 | Erythrocin IV lactobionate 1g powder for solution for infusion vials (Advanz Pharma) | macrolides |
| 27768 | Erythrolar 250mg Tablet (Lagap) | macrolides |
| 50205 | Erythrolar 250mg tablets (Ennogen Pharma Ltd) | macrolides |
| 4153 | Erythrolar 250mg/5ml Liquid (Lagap) | macrolides |
| 23954 | Erythrolar 500mg Tablet (Lagap) | macrolides |
| 49301 | Erythrolar 500mg tablets (Ennogen Pharma Ltd) | macrolides |
| 14429 | Erythromycin 125mg sprinkle capsules | macrolides |
| 34231 | Erythromycin 125mg/5ml Liquid (Berk Pharmaceuticals Ltd) | macrolides |
| 33248 | Erythromycin 125mg/5ml Liquid (IVAX Pharmaceuticals UK Ltd) | macrolides |
| 397 | Erythromycin 125mg/5ml oral suspension | macrolides |
| 26189 | Erythromycin 1g powder for solution for infusion vials | macrolides |
| 77276 | Erythromycin 1g powder for solution for infusion vials (Advanz Pharma) | macrolides |
| 553 | Erythromycin 250mg.5ml oral suspension | macrolides |
| 75595 | Erythromycin 250mg/5ml Liquid (Berk Pharmaceuticals Ltd) | macrolides |
| 47242 | Erythromycin 250mg/5ml Liquid (C P Pharmaceuticals Ltd) | macrolides |
| 41584 | Erythromycin 250mg/5ml Liquid (IVAX Pharmaceuticals UK Ltd) | macrolides |
| 69330 | Erythromycin 250mg/5ml Liquid (Rosemont Pharmaceuticals Ltd) | macrolides |
| 47676 | Erythromycin 500mg/5ml Liquid (C P Pharmaceuticals Ltd) | macrolides |
| 2326 | Erythromycin 500mg/5ml oral suspension | macrolides |
| 2429 | Erythromycin ethyl succinate 125mg/5ml oral suspension | macrolides |
| 13167 | Erythromycin ethyl succinate 125mg/5ml oral suspension (A A H Pharmaceuticals Ltd) | macrolides |
| 60308 | Erythromycin ethyl succinate 125mg/5ml oral suspension (Alliance Healthcare (Distribution) Ltd) | macrolides |
| 59441 | Erythromycin ethyl succinate 125mg/5ml oral suspension (DE Pharmaceuticals) | macrolides |
| 49978 | Erythromycin ethyl succinate 125mg/5ml oral suspension (Focus Pharmaceuticals Ltd) | macrolides |
| 59126 | Erythromycin ethyl succinate 125mg/5ml oral suspension (Kent Pharmaceuticals Ltd) | macrolides |
| 50948 | Erythromycin ethyl succinate 125mg/5ml oral suspension (Phoenix Healthcare Distribution Ltd) | macrolides |
| 47126 | Erythromycin ethyl succinate 125mg/5ml oral suspension (Pinewood Healthcare) | macrolides |
| 34779 | Erythromycin ethyl succinate 125mg/5ml oral suspension (Sandoz Ltd) | macrolides |
| 65956 | Erythromycin ethyl succinate 125mg/5ml oral suspension (Sigma Pharmaceuticals Plc) | macrolides |
| 58756 | Erythromycin ethyl succinate 125mg/5ml oral suspension (Waymade Healthcare Plc) | macrolides |
| 4672 | Erythromycin ethyl succinate 125mg/5ml oral suspension sugar free | macrolides |
| 33697 | Erythromycin ethyl succinate 125mg/5ml oral suspension sugar free (A A H Pharmaceuticals Ltd) | macrolides |
| 42659 | Erythromycin ethyl succinate 125mg/5ml oral suspension sugar free (Abbott Laboratories Ltd) | macrolides |
| 55589 | Erythromycin ethyl succinate 125mg/5ml oral suspension sugar free (Alliance Healthcare (Distribution) Ltd) | macrolides |
| 62143 | Erythromycin ethyl succinate 125mg/5ml oral suspension sugar free (DE Pharmaceuticals) | macrolides |
| 48101 | Erythromycin ethyl succinate 125mg/5ml oral suspension sugar free (Focus Pharmaceuticals Ltd) | macrolides |
| 34795 | Erythromycin ethyl succinate 125mg/5ml oral suspension sugar free (IVAX Pharmaceuticals UK Ltd) | macrolides |
| 58841 | Erythromycin ethyl succinate 125mg/5ml oral suspension sugar free (Kent Pharmaceuticals Ltd) | macrolides |
| 33695 | Erythromycin ethyl succinate 125mg/5ml oral suspension sugar free (Mylan) | macrolides |
| 64732 | Erythromycin ethyl succinate 125mg/5ml oral suspension sugar free (Phoenix Healthcare Distribution Ltd) | macrolides |
| 45870 | Erythromycin ethyl succinate 125mg/5ml oral suspension sugar free (Pinewood Healthcare) | macrolides |
| 33705 | Erythromycin ethyl succinate 125mg/5ml oral suspension sugar free (Teva UK Ltd) | macrolides |
| 64615 | Erythromycin ethyl succinate 125mg/5ml oral suspension sugar free (Waymade Healthcare Plc) | macrolides |
| 2376 | Erythromycin ethyl succinate 250mg/5ml oral suspension | macrolides |
| 13120 | Erythromycin ethyl succinate 250mg/5ml oral suspension (A A H Pharmaceuticals Ltd) | macrolides |
| 74651 | Erythromycin ethyl succinate 250mg/5ml oral suspension (Actavis UK Ltd) | macrolides |
| 70311 | Erythromycin ethyl succinate 250mg/5ml oral suspension (Alliance Healthcare (Distribution) Ltd) | macrolides |
| 63032 | Erythromycin ethyl succinate 250mg/5ml oral suspension (DE Pharmaceuticals) | macrolides |
| 76071 | Erythromycin ethyl succinate 250mg/5ml oral suspension (Focus Pharmaceuticals Ltd) | macrolides |
| 32902 | Erythromycin ethyl succinate 250mg/5ml oral suspension (Kent Pharmaceuticals Ltd) | macrolides |
| 64312 | Erythromycin ethyl succinate 250mg/5ml oral suspension (Pinewood Healthcare) | macrolides |
| 46696 | Erythromycin ethyl succinate 250mg/5ml oral suspension (Sandoz Ltd) | macrolides |
| 68226 | Erythromycin ethyl succinate 250mg/5ml oral suspension (Sigma Pharmaceuticals Plc) | macrolides |
| 61561 | Erythromycin ethyl succinate 250mg/5ml oral suspension (Waymade Healthcare Plc) | macrolides |
| 2225 | Erythromycin ethyl succinate 250mg/5ml oral suspension sugar free | macrolides |
| 32898 | Erythromycin ethyl succinate 250mg/5ml oral suspension sugar free (A A H Pharmaceuticals Ltd) | macrolides |
| 46154 | Erythromycin ethyl succinate 250mg/5ml oral suspension sugar free (Abbott Laboratories Ltd) | macrolides |
| 52860 | Erythromycin ethyl succinate 250mg/5ml oral suspension sugar free (Alliance Healthcare (Distribution) Ltd) | macrolides |
| 71423 | Erythromycin ethyl succinate 250mg/5ml oral suspension sugar free (Focus Pharmaceuticals Ltd) | macrolides |
| 30177 | Erythromycin ethyl succinate 250mg/5ml oral suspension sugar free (IVAX Pharmaceuticals UK Ltd) | macrolides |
| 73461 | Erythromycin ethyl succinate 250mg/5ml oral suspension sugar free (Kent Pharmaceuticals Ltd) | macrolides |
| 75358 | Erythromycin ethyl succinate 250mg/5ml oral suspension sugar free (Mawdsley-Brooks & Company Ltd) | macrolides |
| 33694 | Erythromycin ethyl succinate 250mg/5ml oral suspension sugar free (Mylan) | macrolides |
| 63458 | Erythromycin ethyl succinate 250mg/5ml oral suspension sugar free (Phoenix Healthcare Distribution Ltd) | macrolides |
| 59036 | Erythromycin ethyl succinate 250mg/5ml oral suspension sugar free (Pinewood Healthcare) | macrolides |
| 74382 | Erythromycin ethyl succinate 250mg/5ml oral suspension sugar free (Sigma Pharmaceuticals Plc) | macrolides |
| 34853 | Erythromycin ethyl succinate 250mg/5ml oral suspension sugar free (Teva UK Ltd) | macrolides |
| 61612 | Erythromycin ethyl succinate 250mg/5ml oral suspension sugar free (Waymade Healthcare Plc) | macrolides |
| 733 | Erythromycin ethyl succinate 500mg tablets | macrolides |
| 75422 | Erythromycin ethyl succinate 500mg tablets (Alliance Healthcare (Distribution) Ltd) | macrolides |
| 68367 | Erythromycin ethyl succinate 500mg tablets (Dawa Ltd) | macrolides |
| 71066 | Erythromycin ethyl succinate 500mg tablets (Sigma Pharmaceuticals Plc) | macrolides |
| 2226 | Erythromycin ethyl succinate 500mg/5ml oral suspension | macrolides |
| 62995 | Erythromycin ethyl succinate 500mg/5ml oral suspension (A A H Pharmaceuticals Ltd) | macrolides |
| 30980 | Erythromycin ethyl succinate 500mg/5ml oral suspension (Kent Pharmaceuticals Ltd) | macrolides |
| 73663 | Erythromycin ethyl succinate 500mg/5ml oral suspension (Pinewood Healthcare) | macrolides |
| 78497 | Erythromycin ethyl succinate 500mg/5ml oral suspension (Sandoz Ltd) | macrolides |
| 14171 | Erythromycin ethyl succinate 500mg/5ml oral suspension sugar free | macrolides |
| 62466 | Erythromycin ethyl succinate 500mg/5ml oral suspension sugar free (A A H Pharmaceuticals Ltd) | macrolides |
| 31514 | Erythromycin ethyl succinate 500mg/5ml oral suspension sugar free (Abbott Laboratories Ltd) | macrolides |
| 60263 | Erythromycin ethyl succinate 500mg/5ml oral suspension sugar free (Alliance Healthcare (Distribution) Ltd) | macrolides |
| 71435 | Erythromycin ethyl succinate 500mg/5ml oral suspension sugar free (Focus Pharmaceuticals Ltd) | macrolides |
| 25595 | Erythromycin ethyl succinate 500mg/5ml oral suspension sugar free (IVAX Pharmaceuticals UK Ltd) | macrolides |
| 72439 | Erythromycin ethyl succinate 500mg/5ml oral suspension sugar free (Pinewood Healthcare) | macrolides |
| 27203 | Erythromycin ethyl succinate 500mg/5ml oral suspension sugar free (Teva UK Ltd) | macrolides |
| 25751 | Erythromycin ethylsuccinate (coated) 250mg/5ml oral suspension sugar free | macrolides |
| 30234 | Erythromycin ethylsuccinate 125mg sachets | macrolides |
| 12330 | Erythromycin ethylsuccinate 1g sachets | macrolides |
| 13635 | Erythromycin ethylsuccinate 250mg sachets | macrolides |
| 15713 | Erythromycin ethylsuccinate 500mg sachets | macrolides |
| 438 | Erythromycin stearate 250mg tablets | macrolides |
| 2350 | Erythromycin stearate 500mg tablets | macrolides |
| 3572 | Erythroped 250mg Powder (Abbott Laboratories Ltd) | macrolides |
| 16747 | Erythroped 250mg Sachets (Abbott Laboratories Ltd) | macrolides |
| 105 | Erythroped 250mg/5ml Liquid (Abbott Laboratories Ltd) | macrolides |
| 532 | Erythroped 250mg/5ml Oral suspension (Abbott Laboratories Ltd) | macrolides |
| 4596 | Erythroped a 1g Sachets (Abbott Laboratories Ltd) | macrolides |
| 327 | Erythroped a 500mg Tablet (Abbott Laboratories Ltd) | macrolides |
| 39632 | Erythroped A 500mg tablets (Advanz Pharma) | macrolides |
| 54098 | Erythroped A 500mg tablets (Lexon (UK) Ltd) | macrolides |
| 65667 | Erythroped A 500mg tablets (Mawdsley-Brooks & Company Ltd) | macrolides |
| 58760 | Erythroped A 500mg tablets (Necessity Supplies Ltd) | macrolides |
| 56203 | Erythroped A 500mg tablets (Sigma Pharmaceuticals Plc) | macrolides |
| 4372 | Erythroped forte 500mg Sachets (Abbott Laboratories Ltd) | macrolides |
| 993 | Erythroped forte 500mg/5ml Liquid (Abbott Laboratories Ltd) | macrolides |
| 4610 | Erythroped forte 500mg/5ml Oral suspension (Abbott Laboratories Ltd) | macrolides |
| 39642 | Erythroped Forte SF 500mg/5ml oral suspension (Advanz Pharma) | macrolides |
| 3042 | Erythroped pi 125mg Sachets (Abbott Laboratories Ltd) | macrolides |
| 997 | Erythroped pi 125mg/5ml Liquid (Abbott Laboratories Ltd) | macrolides |
| 825 | Erythroped pi 125mg/5ml Oral suspension (Abbott Laboratories Ltd) | macrolides |
| 39623 | Erythroped PI SF 125mg/5ml oral suspension (Advanz Pharma) | macrolides |
| 39669 | Erythroped SF 250mg/5ml oral suspension (Advanz Pharma) | macrolides |
| 52502 | Fidaxomicin 200mg tablets | macrolides |
| 31379 | Ketek 400mg tablets (Sanofi) | macrolides |
| 3736 | Klaricid 125mg/5ml Oral suspension (Abbott Laboratories Ltd) | macrolides |
| 2719 | Klaricid 250mg tablets (Abbott Laboratories Ltd) | macrolides |
| 52411 | Klaricid 250mg tablets (Necessity Supplies Ltd) | macrolides |
| 9583 | Klaricid 250mg/5ml Oral suspension (Abbott Laboratories Ltd) | macrolides |
| 6623 | Klaricid 500 tablets (Abbott Laboratories Ltd) | macrolides |
| 63236 | Klaricid 500 tablets (Necessity Supplies Ltd) | macrolides |
| 65259 | Klaricid 500 tablets (Sigma Pharmaceuticals Plc) | macrolides |
| 14816 | Klaricid Adult 250mg granules sachets (Mylan) | macrolides |
| 28289 | Klaricid IV 500mg powder for solution for infusion vials (Mylan) | macrolides |
| 71154 | Klaricid Paediatric 125mg/5ml oral suspension (Dowelhurst Ltd) | macrolides |
| 38997 | Klaricid Paediatric 125mg/5ml oral suspension (Mylan) | macrolides |
| 79303 | Klaricid Paediatric 250mg/5ml oral suspension (Dowelhurst Ltd) | macrolides |
| 39010 | Klaricid Paediatric 250mg/5ml oral suspension (Mylan) | macrolides |
| 75316 | Klaricid XL 500mg tablets (Dowelhurst Ltd) | macrolides |
| 6121 | Klaricid XL 500mg tablets (Mylan) | macrolides |
| 30520 | Primacine 125mg/5ml Liquid (Pinewood Healthcare) | macrolides |
| 39118 | Primacine 250mg/5ml Liquid (Pinewood Healthcare) | macrolides |
| 27504 | Primacine 500mg/5ml Liquid (Pinewood Healthcare) | macrolides |
| 21808 | Rommix 125mg/5ml Oral suspension sugar free (Ashbourne Pharmaceuticals Ltd) | macrolides |
| 31420 | Rovamycin 500mg Tablet (Rhone-Poulenc Rorer Ltd) | macrolides |
| 66957 | Roxithromycin 150mg tablets | macrolides |
| 25900 | Spiramycin 500mg tablet | macrolides |
| 22964 | Telithromycin 400mg tablets | macrolides |
| 14514 | Zithromax 200mg/5ml oral suspension (Pfizer Ltd) | macrolides |
| 69438 | Zithromax 200mg/5ml oral suspension (Waymade Healthcare Plc) | macrolides |
| 52029 | Zithromax 250mg capsules (Mawdsley-Brooks & Company Ltd) | macrolides |
| 4165 | Zithromax 250mg capsules (Pfizer Ltd) | macrolides |
| 5335 | Zithromax 500mg tablets (Pfizer Ltd) | macrolides |
| 4489 | Erycen 250mg Tablet (Berk Pharmaceuticals Ltd) | macrolides |
| 23017 | Erycen 500mg Tablet (Berk Pharmaceuticals Ltd) | macrolides |
| 318 | Erymax 250mg Capsule (Elan Pharma) | macrolides |
| 10190 | Erymax 250mg gastro-resistant capsules (Teva UK Ltd) | macrolides |
| 3209 | Erythromid 250mg Tablet (Abbott Laboratories Ltd) | macrolides |
| 9148 | Erythromid ds 500mg Tablet (Abbott Laboratories Ltd) | macrolides |
| 29154 | Erythromycin 250mg Capsule (Actavis UK Ltd) | macrolides |
| 33686 | Erythromycin 250mg gastro-resistant capsules (A A H Pharmaceuticals Ltd) | macrolides |
| 50580 | Erythromycin 250mg gastro-resistant capsules (Actavis UK Ltd) | macrolides |
| 50694 | Erythromycin 250mg gastro-resistant capsules (Alliance Healthcare (Distribution) Ltd) | macrolides |
| 79164 | Erythromycin 250mg gastro-resistant capsules (Arrow Generics Ltd) | macrolides |
| 55133 | Erythromycin 250mg gastro-resistant capsules (Kent Pharmaceuticals Ltd) | macrolides |
| 49952 | Erythromycin 250mg gastro-resistant capsules (Phoenix Healthcare Distribution Ltd) | macrolides |
| 34512 | Erythromycin 250mg gastro-resistant capsules (Teva UK Ltd) | macrolides |
| 55397 | Erythromycin 250mg gastro-resistant capsules (Waymade Healthcare Plc) | macrolides |
| 34837 | Erythromycin 250mg Gastro-resistant tablet (Co-Pharma Ltd) | macrolides |
| 63 | Erythromycin 250mg gastro-resistant tablets | macrolides |
| 24127 | Erythromycin 250mg gastro-resistant tablets (A A H Pharmaceuticals Ltd) | macrolides |
| 33703 | Erythromycin 250mg gastro-resistant tablets (Abbott Laboratories Ltd) | macrolides |
| 29344 | Erythromycin 250mg gastro-resistant tablets (Actavis UK Ltd) | macrolides |
| 52906 | Erythromycin 250mg gastro-resistant tablets (Alliance Healthcare (Distribution) Ltd) | macrolides |
| 42661 | Erythromycin 250mg gastro-resistant tablets (Almus Pharmaceuticals Ltd) | macrolides |
| 60828 | Erythromycin 250mg gastro-resistant tablets (Bristol Laboratories Ltd) | macrolides |
| 61264 | Erythromycin 250mg gastro-resistant tablets (DE Pharmaceuticals) | macrolides |
| 42296 | Erythromycin 250mg gastro-resistant tablets (Dr Reddy's Laboratories (UK) Ltd) | macrolides |
| 69403 | Erythromycin 250mg gastro-resistant tablets (Genesis Pharmaceuticals Ltd) | macrolides |
| 24129 | Erythromycin 250mg gastro-resistant tablets (IVAX Pharmaceuticals UK Ltd) | macrolides |
| 58824 | Erythromycin 250mg gastro-resistant tablets (Kent Pharmaceuticals Ltd) | macrolides |
| 69480 | Erythromycin 250mg gastro-resistant tablets (Mawdsley-Brooks & Company Ltd) | macrolides |
| 53986 | Erythromycin 250mg gastro-resistant tablets (Medreich Plc) | macrolides |
| 55483 | Erythromycin 250mg gastro-resistant tablets (Milpharm Ltd) | macrolides |
| 34334 | Erythromycin 250mg gastro-resistant tablets (Mylan) | macrolides |
| 52428 | Erythromycin 250mg gastro-resistant tablets (Phoenix Healthcare Distribution Ltd) | macrolides |
| 31530 | Erythromycin 250mg gastro-resistant tablets (Ranbaxy (UK) Ltd) | macrolides |
| 74355 | Erythromycin 250mg gastro-resistant tablets (Sigma Pharmaceuticals Plc) | macrolides |
| 34479 | Erythromycin 250mg gastro-resistant tablets (Sovereign Medical Ltd) | macrolides |
| 52952 | Erythromycin 250mg gastro-resistant tablets (Strides Pharma UK Ltd) | macrolides |
| 33685 | Erythromycin 250mg gastro-resistant tablets (Teva UK Ltd) | macrolides |
| 59100 | Erythromycin 250mg gastro-resistant tablets (Waymade Healthcare Plc) | macrolides |
| 60190 | Erythromycin 250mg gastro-resistant tablets (Waymade Healthcare Plc) | macrolides |
| 34873 | Erythromycin 250mg Tablet (Berk Pharmaceuticals Ltd) | macrolides |
| 34189 | Erythromycin 250mg Tablet (C P Pharmaceuticals Ltd) | macrolides |
| 401 | Erythromycin 500mg ec gastro-resistant tablets | macrolides |
| 34869 | Erythromycin 500mg Tablet (C P Pharmaceuticals Ltd) | macrolides |
| 41604 | Erythromycin 500mg Tablet (Hillcross Pharmaceuticals Ltd) | macrolides |
| 26365 | Erythromycin 500mg Tablet (IVAX Pharmaceuticals UK Ltd) | macrolides |
| 55300 | Erythromycin 500mg Tablet (Teva UK Ltd) | macrolides |
| 37796 | Erythromycin estolate 125mg/5ml suspension | macrolides |
| 9903 | Erythromycin estolate 250mg capsules | macrolides |
| 40073 | Erythromycin estolate 250mg/5ml suspension | macrolides |
| 37694 | Erythromycin estolate 500mg tablets | macrolides |
| 18682 | Ilosone 125mg/5ml Liquid (Dista Products Ltd) | macrolides |
| 17207 | Ilosone 250mg Capsule (Dista Products Ltd) | macrolides |
| 19330 | Ilosone 250mg/5ml Liquid (Dista Products Ltd) | macrolides |
| 18643 | Ilosone 500mg Tablet (Dista Products Ltd) | macrolides |
| 23244 | Ilotycin 250mg Tablet (Eli Lilly and Company Ltd) | macrolides |
| 33304 | Kerymax 250mg gastro-resistant capsules (Kent Pharmaceuticals Ltd) | macrolides |
| 31428 | Retcin 250mg Tablet (DDSA Pharmaceuticals Ltd) | macrolides |
| 11611 | Rommix 250 EC tablets (Ashbourne Pharmaceuticals Ltd) | macrolides |
| 25278 | Rommix 500mg Tablet (Ashbourne Pharmaceuticals Ltd) | macrolides |
| 25280 | Tiloryth 250mg gastro-resistant capsules (Tillomed Laboratories Ltd) | macrolides |
| 27016 | CIPROFLOXACIN | metronidazole |
| 48031 | Ciprofloxacin 100mg tablets (Almus Pharmaceuticals Ltd) | metronidazole |
| 73645 | Ciprofloxacin 750mg tablets (Almus Pharmaceuticals Ltd) | metronidazole |
| 76303 | Generic Mictral granules 7g sachets | metronidazole |
| 27584 | METRONIDAZOLE | metronidazole |
| 26834 | METRONIDAZOLE | metronidazole |
| 12441 | METRONIDAZOLE .5 % SOL | metronidazole |
| 21059 | METRONIDAZOLE .9 % SOL | metronidazole |
| 27210 | Metronidazole 100mg/20ml solution for infusion ampoules | metronidazole |
| 1681 | METRONIDAZOLE 200 MG MIX | metronidazole |
| 12199 | METRONIDAZOLE POW | metronidazole |
| 7674 | Mictral granules 7g sachets (Sanofi-Synthelabo Ltd) | metronidazole |
| 19659 | MICTRAL SACHETS | metronidazole |
| 10306 | Nalidixic acid 300mg/5ml oral suspension sugar free | metronidazole |
| 4064 | Nalidixic acid 500mg tablets | metronidazole |
| 7414 | Negram 300mg/5ml oral suspension (Sanofi-Synthelabo Ltd) | metronidazole |
| 7513 | Negram 500mg tablets (Sanofi) | metronidazole |
| 25078 | NITROFURANTOIN | metronidazole |
| 7495 | NITROFURANTOIN 25 MG TAB | metronidazole |
| 19008 | PHENAZOPYRIDINE HCl 100 MG TAB | metronidazole |
| 2973 | PYRIDIUM .1 GM TAB | metronidazole |
| 6497 | Clarithromycin 500mg with metronidazole 400mg with lansoprazole 30mg triple pack | metronidazole |
| 21060 | Metrolyl 200mg Tablet (Sandoz Ltd) | metronidazole |
| 4465 | Metrolyl 400mg Tablet (Sandoz Ltd) | metronidazole |
| 75009 | Metronidazole 200mg Tablet (Regent Laboratories Ltd) | metronidazole |
| 66324 | Metronidazole Oral solution | metronidazole |
| 24696 | Vaginyl 200mg Tablet (DDSA Pharmaceuticals Ltd) | metronidazole |
| 9652 | Fasigyn 500mg tablets (Pfizer Ltd) | metronidazole |
| 702 | Tinidazole 500mg tablets | metronidazole |
| 28245 | Elyzol 1g Suppository (C P Pharmaceuticals Ltd) | metronidazole |
| 41669 | Metronidazole 1g Suppository (C P Pharmaceuticals Ltd) | metronidazole |
| 46933 | Metronidazole 1g Suppository (Sandoz Ltd) | metronidazole |
| 29590 | Zadstat 500mg Suppository (Wyeth Pharmaceuticals) | metronidazole |
| 37857 | Cefuroxime 1.5g with 500mg infusion | metronidazole |
| 29183 | Cefuroxime 750mg with metronidazole 500mg infusion | metronidazole |
| 7106 | Flagyl 1g suppositories (Sanofi) | metronidazole |
| 449 | Flagyl 200mg tablets (Sanofi) | metronidazole |
| 3499 | Flagyl 400mg tablets (Sanofi) | metronidazole |
| 17240 | Flagyl 500mg suppositories (Sanofi) | metronidazole |
| 15372 | Flagyl 500mg/100ml infusion 100ml bags (Baxter Healthcare Ltd) | metronidazole |
| 151 | Flagyl 5mg/ml Infusion (Aventis Pharma) | metronidazole |
| 46188 | Flagyl minibag plus 500mg/100ml Infusion (Baxter Healthcare Ltd) | metronidazole |
| 3702 | Flagyl-S 200mg/5ml oral suspension (Zentiva) | metronidazole |
| 15216 | Metrolyl 1g suppositories (Sandoz Ltd) | metronidazole |
| 22420 | Metrolyl 500mg suppositories (Sandoz Ltd) | metronidazole |
| 57554 | Metronidazole 100mg/5ml oral solution | metronidazole |
| 50234 | Metronidazole 100mg/5ml oral suspension | metronidazole |
| 5836 | Metronidazole 1g suppositories | metronidazole |
| 75734 | Metronidazole 200mg suppositories | metronidazole |
| 305 | Metronidazole 200mg tablets | metronidazole |
| 45260 | Metronidazole 200mg tablets (A A H Pharmaceuticals Ltd) | metronidazole |
| 41692 | Metronidazole 200mg tablets (Actavis UK Ltd) | metronidazole |
| 50808 | Metronidazole 200mg tablets (Alliance Healthcare (Distribution) Ltd) | metronidazole |
| 40994 | Metronidazole 200mg tablets (Almus Pharmaceuticals Ltd) | metronidazole |
| 51910 | Metronidazole 200mg tablets (Crescent Pharma Ltd) | metronidazole |
| 62878 | Metronidazole 200mg tablets (DE Pharmaceuticals) | metronidazole |
| 34596 | Metronidazole 200mg tablets (IVAX Pharmaceuticals UK Ltd) | metronidazole |
| 52636 | Metronidazole 200mg tablets (Kent Pharmaceuticals Ltd) | metronidazole |
| 67979 | Metronidazole 200mg tablets (Mawdsley-Brooks & Company Ltd) | metronidazole |
| 49531 | Metronidazole 200mg tablets (Milpharm Ltd) | metronidazole |
| 36757 | Metronidazole 200mg tablets (Mylan) | metronidazole |
| 53959 | Metronidazole 200mg tablets (Phoenix Healthcare Distribution Ltd) | metronidazole |
| 38544 | Metronidazole 200mg tablets (Sandoz Ltd) | metronidazole |
| 59056 | Metronidazole 200mg tablets (Sigma Pharmaceuticals Plc) | metronidazole |
| 32068 | Metronidazole 200mg tablets (Teva UK Ltd) | metronidazole |
| 61619 | Metronidazole 200mg tablets (Waymade Healthcare Plc) | metronidazole |
| 63765 | Metronidazole 200mg/5ml oral solution | metronidazole |
| 4933 | Metronidazole 200mg/5ml oral suspension | metronidazole |
| 31531 | Metronidazole 200mg/5ml oral suspension (A A H Pharmaceuticals Ltd) | metronidazole |
| 56049 | Metronidazole 200mg/5ml oral suspension (Alliance Healthcare (Distribution) Ltd) | metronidazole |
| 76909 | Metronidazole 200mg/5ml oral suspension (Dawa Ltd) | metronidazole |
| 76849 | Metronidazole 200mg/5ml oral suspension (DE Pharmaceuticals) | metronidazole |
| 29336 | Metronidazole 200mg/5ml oral suspension (Rosemont Pharmaceuticals Ltd) | metronidazole |
| 78326 | Metronidazole 200mg/5ml oral suspension (Sigma Pharmaceuticals Plc) | metronidazole |
| 61832 | Metronidazole 200mg/5ml oral suspension (Waymade Healthcare Plc) | metronidazole |
| 57159 | Metronidazole 200mg/5ml oral suspension (Zentiva) | metronidazole |
| 431 | Metronidazole 400mg tablets | metronidazole |
| 19205 | Metronidazole 400mg tablets (A A H Pharmaceuticals Ltd) | metronidazole |
| 29327 | Metronidazole 400mg tablets (Actavis UK Ltd) | metronidazole |
| 55666 | Metronidazole 400mg tablets (Alliance Healthcare (Distribution) Ltd) | metronidazole |
| 47990 | Metronidazole 400mg tablets (Almus Pharmaceuticals Ltd) | metronidazole |
| 54262 | Metronidazole 400mg tablets (Crescent Pharma Ltd) | metronidazole |
| 61434 | Metronidazole 400mg tablets (DE Pharmaceuticals) | metronidazole |
| 34405 | Metronidazole 400mg tablets (IVAX Pharmaceuticals UK Ltd) | metronidazole |
| 52481 | Metronidazole 400mg tablets (Kent Pharmaceuticals Ltd) | metronidazole |
| 57216 | Metronidazole 400mg tablets (Milpharm Ltd) | metronidazole |
| 34306 | Metronidazole 400mg tablets (Mylan) | metronidazole |
| 58219 | Metronidazole 400mg tablets (Phoenix Healthcare Distribution Ltd) | metronidazole |
| 40090 | Metronidazole 400mg tablets (Sandoz Ltd) | metronidazole |
| 67218 | Metronidazole 400mg tablets (Sigma Pharmaceuticals Plc) | metronidazole |
| 33896 | Metronidazole 400mg tablets (Teva UK Ltd) | metronidazole |
| 59906 | Metronidazole 400mg tablets (Waymade Healthcare Plc) | metronidazole |
| 5461 | Metronidazole 500mg suppositories | metronidazole |
| 1311 | Metronidazole 500mg tablets | metronidazole |
| 22246 | Metronidazole 500mg tablets (Accord Healthcare Ltd) | metronidazole |
| 59724 | Metronidazole 500mg tablets (Alliance Healthcare (Distribution) Ltd) | metronidazole |
| 61034 | Metronidazole 500mg tablets (Sigma Pharmaceuticals Plc) | metronidazole |
| 15375 | Metronidazole 500mg/100ml infusion 100ml bags | metronidazole |
| 63589 | Metronidazole 500mg/100ml infusion 100ml bags (A A H Pharmaceuticals Ltd) | metronidazole |
| 56448 | Metronidazole 500mg/100ml infusion 100ml Macoflex bags (Maco Pharma (UK) Ltd) | metronidazole |
| 72801 | Metronidazole 500mg/100ml solution for infusion bottles (Peckforton Pharmaceuticals Ltd) | metronidazole |
| 67676 | Metronidazole 500mg/5ml oral suspension | metronidazole |
| 65099 | Metronidazole 50mg/5ml oral solution | metronidazole |
| 16179 | Metronidazole 5mg/ml Infusion | metronidazole |
| 406 | Metronidazole Treatment pack | metronidazole |
| 28452 | Metrozol 500mg/100ml Infusion (Parkfields) | metronidazole |
| 30089 | Norzol 200mg/5ml oral suspension (Rosemont Pharmaceuticals Ltd) | metronidazole |
| 32683 | Tinidazole 2mg/ml Infusion | metronidazole |
| 25127 | Avelox 400mg tablets (Bayer Plc) | metronidazole |
| 47995 | Avelox 400mg/250ml solution for infusion bottles (Bayer Plc) | metronidazole |
| 10567 | Cinobac 500mg Capsule (Eli Lilly and Company Ltd) | metronidazole |
| 12277 | Cinoxacin 500mg capsules | metronidazole |
| 498 | Ciprofloxacin 100mg tablets | metronidazole |
| 42507 | Ciprofloxacin 100mg tablets (A A H Pharmaceuticals Ltd) | metronidazole |
| 58323 | Ciprofloxacin 100mg tablets (Alliance Healthcare (Distribution) Ltd) | metronidazole |
| 61302 | Ciprofloxacin 100mg tablets (Almus Pharmaceuticals Ltd) | metronidazole |
| 58608 | Ciprofloxacin 100mg tablets (Bristol Laboratories Ltd) | metronidazole |
| 54555 | Ciprofloxacin 100mg tablets (DE Pharmaceuticals) | metronidazole |
| 58021 | Ciprofloxacin 100mg tablets (Dr Reddy's Laboratories (UK) Ltd) | metronidazole |
| 54674 | Ciprofloxacin 100mg tablets (Phoenix Healthcare Distribution Ltd) | metronidazole |
| 39913 | Ciprofloxacin 100mg tablets (Sandoz Ltd) | metronidazole |
| 52309 | Ciprofloxacin 100mg tablets (Sigma Pharmaceuticals Plc) | metronidazole |
| 26840 | Ciprofloxacin 100mg/50ml solution for infusion bottles | metronidazole |
| 11883 | Ciprofloxacin 100mg/50ml solution for infusion vials | metronidazole |
| 58955 | Ciprofloxacin 100mg/50ml solution for infusion vials (A A H Pharmaceuticals Ltd) | metronidazole |
| 70998 | Ciprofloxacin 100mg/5ml oral suspension (Special Order) | metronidazole |
| 77778 | Ciprofloxacin 150mg/5ml oral suspension | metronidazole |
| 66483 | Ciprofloxacin 170mg/5ml oral suspension | metronidazole |
| 33215 | Ciprofloxacin 200mg/100ml in sodium chloride 0.9% infusion | metronidazole |
| 38171 | Ciprofloxacin 200mg/100ml infusion bags | metronidazole |
| 57703 | Ciprofloxacin 200mg/100ml solution for infusion bottles | metronidazole |
| 71572 | Ciprofloxacin 200mg/100ml solution for infusion bottles (Kent Pharmaceuticals Ltd) | metronidazole |
| 52945 | Ciprofloxacin 200mg/100ml solution for infusion vials | metronidazole |
| 56439 | Ciprofloxacin 200mg/100ml solution for infusion vials (A A H Pharmaceuticals Ltd) | metronidazole |
| 34647 | Ciprofloxacin 250mg Tablet (Neo Laboratories Ltd) | metronidazole |
| 281 | Ciprofloxacin 250mg tablets | metronidazole |
| 29343 | Ciprofloxacin 250mg tablets (A A H Pharmaceuticals Ltd) | metronidazole |
| 50601 | Ciprofloxacin 250mg tablets (Accord Healthcare Ltd) | metronidazole |
| 34308 | Ciprofloxacin 250mg tablets (Actavis UK Ltd) | metronidazole |
| 51537 | Ciprofloxacin 250mg tablets (Alliance Healthcare (Distribution) Ltd) | metronidazole |
| 60436 | Ciprofloxacin 250mg tablets (Almus Pharmaceuticals Ltd) | metronidazole |
| 71011 | Ciprofloxacin 250mg tablets (APC Pharmaceuticals & Chemicals (Europe) Ltd) | metronidazole |
| 54393 | Ciprofloxacin 250mg tablets (Arrow Generics Ltd) | metronidazole |
| 54701 | Ciprofloxacin 250mg tablets (Bristol Laboratories Ltd) | metronidazole |
| 58235 | Ciprofloxacin 250mg tablets (DE Pharmaceuticals) | metronidazole |
| 43814 | Ciprofloxacin 250mg tablets (Dr Reddy's Laboratories (UK) Ltd) | metronidazole |
| 41561 | Ciprofloxacin 250mg tablets (IVAX Pharmaceuticals UK Ltd) | metronidazole |
| 57118 | Ciprofloxacin 250mg tablets (Kent Pharmaceuticals Ltd) | metronidazole |
| 54302 | Ciprofloxacin 250mg tablets (Medreich Plc) | metronidazole |
| 33989 | Ciprofloxacin 250mg tablets (Mylan) | metronidazole |
| 34448 | Ciprofloxacin 250mg tablets (Niche Generics Ltd) | metronidazole |
| 73301 | Ciprofloxacin 250mg tablets (Phoenix Healthcare Distribution Ltd) | metronidazole |
| 34694 | Ciprofloxacin 250mg tablets (PLIVA Pharma Ltd) | metronidazole |
| 66214 | Ciprofloxacin 250mg tablets (Ranbaxy (UK) Ltd) | metronidazole |
| 34559 | Ciprofloxacin 250mg tablets (Sandoz Ltd) | metronidazole |
| 78359 | Ciprofloxacin 250mg tablets (Sigma Pharmaceuticals Plc) | metronidazole |
| 56381 | Ciprofloxacin 250mg tablets (Strides Pharma UK Ltd) | metronidazole |
| 34478 | Ciprofloxacin 250mg tablets (Teva UK Ltd) | metronidazole |
| 64446 | Ciprofloxacin 250mg tablets (Tillomed Laboratories Ltd) | metronidazole |
| 61783 | Ciprofloxacin 250mg tablets (Waymade Healthcare Plc) | metronidazole |
| 34655 | Ciprofloxacin 250mg tablets (Wockhardt UK Ltd) | metronidazole |
| 4091 | Ciprofloxacin 250mg/5ml oral suspension | metronidazole |
| 10304 | Ciprofloxacin 2mg/ml infusion | metronidazole |
| 29507 | Ciprofloxacin 400mg/200ml in sodium chloride 0.9% infusion | metronidazole |
| 47785 | Ciprofloxacin 400mg/200ml infusion bags | metronidazole |
| 73388 | Ciprofloxacin 400mg/200ml infusion bags (A A H Pharmaceuticals Ltd) | metronidazole |
| 75275 | Ciprofloxacin 400mg/200ml infusion bags (Bowmed Ibisqus Ltd) | metronidazole |
| 58246 | Ciprofloxacin 400mg/200ml infusion bags (Pfizer Ltd) | metronidazole |
| 58074 | Ciprofloxacin 400mg/200ml solution for infusion bottles | metronidazole |
| 69893 | Ciprofloxacin 400mg/200ml solution for infusion bottles (Kent Pharmaceuticals Ltd) | metronidazole |
| 54993 | Ciprofloxacin 400mg/200ml solution for infusion vials | metronidazole |
| 66971 | Ciprofloxacin 400mg/200ml solution for infusion vials (A A H Pharmaceuticals Ltd) | metronidazole |
| 64814 | Ciprofloxacin 400mg/200ml solution for infusion vials (Genus Pharmaceuticals Ltd) | metronidazole |
| 73933 | Ciprofloxacin 400mg/200ml solution for infusion vials (Intrapharm Laboratories Ltd) | metronidazole |
| 45341 | Ciprofloxacin 500mg Tablet (Neo Laboratories Ltd) | metronidazole |
| 34322 | Ciprofloxacin 500mg Tablet (Niche Generics Ltd) | metronidazole |
| 583 | Ciprofloxacin 500mg tablets | metronidazole |
| 29458 | Ciprofloxacin 500mg tablets (A A H Pharmaceuticals Ltd) | metronidazole |
| 52501 | Ciprofloxacin 500mg tablets (Accord Healthcare Ltd) | metronidazole |
| 34605 | Ciprofloxacin 500mg tablets (Actavis UK Ltd) | metronidazole |
| 72884 | Ciprofloxacin 500mg tablets (Alliance Healthcare (Distribution) Ltd) | metronidazole |
| 49445 | Ciprofloxacin 500mg tablets (Almus Pharmaceuticals Ltd) | metronidazole |
| 56789 | Ciprofloxacin 500mg tablets (APC Pharmaceuticals & Chemicals (Europe) Ltd) | metronidazole |
| 52616 | Ciprofloxacin 500mg tablets (Arrow Generics Ltd) | metronidazole |
| 67656 | Ciprofloxacin 500mg tablets (Bristol Laboratories Ltd) | metronidazole |
| 50055 | Ciprofloxacin 500mg tablets (DE Pharmaceuticals) | metronidazole |
| 53088 | Ciprofloxacin 500mg tablets (Dr Reddy's Laboratories (UK) Ltd) | metronidazole |
| 42174 | Ciprofloxacin 500mg tablets (IVAX Pharmaceuticals UK Ltd) | metronidazole |
| 64301 | Ciprofloxacin 500mg tablets (Kent Pharmaceuticals Ltd) | metronidazole |
| 55917 | Ciprofloxacin 500mg tablets (Medreich Plc) | metronidazole |
| 30707 | Ciprofloxacin 500mg tablets (Mylan) | metronidazole |
| 79288 | Ciprofloxacin 500mg tablets (Nexcape Pharmaceuticals Ltd) | metronidazole |
| 43557 | Ciprofloxacin 500mg tablets (PLIVA Pharma Ltd) | metronidazole |
| 53878 | Ciprofloxacin 500mg tablets (Ranbaxy (UK) Ltd) | metronidazole |
| 43797 | Ciprofloxacin 500mg tablets (Sandoz Ltd) | metronidazole |
| 59572 | Ciprofloxacin 500mg tablets (Sigma Pharmaceuticals Plc) | metronidazole |
| 53641 | Ciprofloxacin 500mg tablets (Strides Pharma UK Ltd) | metronidazole |
| 45285 | Ciprofloxacin 500mg tablets (Teva UK Ltd) | metronidazole |
| 57960 | Ciprofloxacin 500mg tablets (Tillomed Laboratories Ltd) | metronidazole |
| 34494 | Ciprofloxacin 500mg tablets (Wockhardt UK Ltd) | metronidazole |
| 66727 | Ciprofloxacin 500mg/5ml oral suspension | metronidazole |
| 34973 | Ciprofloxacin 750mg Tablet (Niche Generics Ltd) | metronidazole |
| 1837 | Ciprofloxacin 750mg tablets | metronidazole |
| 29472 | Ciprofloxacin 750mg tablets (A A H Pharmaceuticals Ltd) | metronidazole |
| 59937 | Ciprofloxacin 750mg tablets (Accord Healthcare Ltd) | metronidazole |
| 43517 | Ciprofloxacin 750mg tablets (Actavis UK Ltd) | metronidazole |
| 76430 | Ciprofloxacin 750mg tablets (APC Pharmaceuticals & Chemicals (Europe) Ltd) | metronidazole |
| 52099 | Ciprofloxacin 750mg tablets (Bristol Laboratories Ltd) | metronidazole |
| 63501 | Ciprofloxacin 750mg tablets (Medreich Plc) | metronidazole |
| 68409 | Ciprofloxacin 750mg tablets (Phoenix Healthcare Distribution Ltd) | metronidazole |
| 56856 | Ciprofloxacin 750mg tablets (Ranbaxy (UK) Ltd) | metronidazole |
| 74658 | Ciprofloxacin 750mg tablets (Sandoz Ltd) | metronidazole |
| 28544 | Ciprofloxaxin 400mg/200ml in glucose 5% infusion | metronidazole |
| 9154 | Ciproxin 100mg tablets (Bayer Plc) | metronidazole |
| 32388 | Ciproxin 200mg/100ml Infusion (Bayer Plc) | metronidazole |
| 1202 | Ciproxin 250mg tablets (Bayer Plc) | metronidazole |
| 52353 | Ciproxin 250mg tablets (DE Pharmaceuticals) | metronidazole |
| 53519 | Ciproxin 250mg tablets (Lexon (UK) Ltd) | metronidazole |
| 65896 | Ciproxin 250mg tablets (Waymade Healthcare Plc) | metronidazole |
| 163 | Ciproxin 250mg/5ml oral suspension (Bayer Plc) | metronidazole |
| 71582 | Ciproxin 250mg/5ml oral suspension (Lexon (UK) Ltd) | metronidazole |
| 61869 | Ciproxin 250mg/5ml oral suspension (Waymade Healthcare Plc) | metronidazole |
| 14376 | Ciproxin 2mg/ml Infusion (Bayer Plc) | metronidazole |
| 38006 | Ciproxin 400mg/200ml Infusion (Bayer Plc) | metronidazole |
| 728 | Ciproxin 500mg tablets (Bayer Plc) | metronidazole |
| 68274 | Ciproxin 500mg tablets (DE Pharmaceuticals) | metronidazole |
| 52807 | Ciproxin 500mg tablets (Mawdsley-Brooks & Company Ltd) | metronidazole |
| 52177 | Ciproxin 500mg tablets (Sigma Pharmaceuticals Plc) | metronidazole |
| 49839 | Ciproxin 500mg tablets (Waymade Healthcare Plc) | metronidazole |
| 7752 | Ciproxin 750mg tablets (Bayer Plc) | metronidazole |
| 21812 | Ciproxin Infusion 100mg/50ml solution for infusion bottles (Bayer Plc) | metronidazole |
| 54663 | Ciproxin Infusion 200mg/100ml solution for infusion bottles (Bayer Plc) | metronidazole |
| 59653 | Ciproxin Infusion 400mg/200ml solution for infusion bottles (Bayer Plc) | metronidazole |
| 32530 | Ciproxin iv flexibag 400mg/200ml Infusion (Bayer Plc) | metronidazole |
| 14389 | Comprecin 200mg Tablet (Parke-davis Research Laboratories) | metronidazole |
| 12428 | Enoxacin 200mg tablets | metronidazole |
| 17890 | Eradacin 150mg Capsule (Sanofi-Synthelabo Ltd) | metronidazole |
| 48200 | Grepafloxacin 400mg Tablet | metronidazole |
| 23666 | Grepafloxacin 600mg Tablet | metronidazole |
| 73859 | Levofloxacin 100mg/ml nebuliser liquid ampoules | metronidazole |
| 6295 | Levofloxacin 250mg tablets | metronidazole |
| 58940 | Levofloxacin 250mg tablets (A A H Pharmaceuticals Ltd) | metronidazole |
| 67572 | Levofloxacin 250mg tablets (Accord Healthcare Ltd) | metronidazole |
| 55708 | Levofloxacin 250mg tablets (Actavis UK Ltd) | metronidazole |
| 72157 | Levofloxacin 250mg tablets (DE Pharmaceuticals) | metronidazole |
| 56012 | Levofloxacin 250mg tablets (Dr Reddy's Laboratories (UK) Ltd) | metronidazole |
| 75298 | Levofloxacin 250mg tablets (Macleods Pharma UK Ltd) | metronidazole |
| 58345 | Levofloxacin 250mg tablets (Mylan) | metronidazole |
| 73335 | Levofloxacin 250mg tablets (Teva UK Ltd) | metronidazole |
| 5238 | Levofloxacin 500mg tablets | metronidazole |
| 61850 | Levofloxacin 500mg tablets (A A H Pharmaceuticals Ltd) | metronidazole |
| 64991 | Levofloxacin 500mg tablets (Accord Healthcare Ltd) | metronidazole |
| 60817 | Levofloxacin 500mg tablets (Actavis UK Ltd) | metronidazole |
| 78938 | Levofloxacin 500mg tablets (Alliance Healthcare (Distribution) Ltd) | metronidazole |
| 73431 | Levofloxacin 500mg tablets (DE Pharmaceuticals) | metronidazole |
| 73464 | Levofloxacin 500mg tablets (Mylan) | metronidazole |
| 77918 | Levofloxacin 500mg tablets (Teva UK Ltd) | metronidazole |
| 65885 | Levofloxacin 500mg tablets (Waymade Healthcare Plc) | metronidazole |
| 53673 | Levofloxacin 500mg/100ml infusion bags | metronidazole |
| 10319 | Levofloxacin 500mg/100ml Intravenous infusion | metronidazole |
| 66211 | Levofloxacin 500mg/100ml solution for infusion bottles | metronidazole |
| 56075 | Levofloxacin 500mg/100ml solution for infusion vials | metronidazole |
| 72099 | Moxifloxacin 0.3mg/0.3ml solution for injection pre-filled syringes | metronidazole |
| 6306 | Moxifloxacin 400mg tablets | metronidazole |
| 43123 | Moxifloxacin 400mg/250ml solution for infusion bottles | metronidazole |
| 77184 | Nalidixic 500mg Tablet (IVAX Pharmaceuticals UK Ltd) | metronidazole |
| 372 | Nalidixic acid 300mg/5ml oral suspension | metronidazole |
| 79657 | Nalidixic acid 300mg/5ml oral suspension (Alliance Healthcare (Distribution) Ltd) | metronidazole |
| 9073 | Nalidixic acid with sodium citrate 660mg + 3750mg Sachets | metronidazole |
| 7519 | Norfloxacin 400mg tablets | metronidazole |
| 79522 | Norfloxacin 400mg tablets (A A H Pharmaceuticals Ltd) | metronidazole |
| 32112 | Norfloxacin 400mg tablets (Genus Pharmaceuticals Ltd) | metronidazole |
| 70756 | Ofloxacin 100mg/5ml oral solution | metronidazole |
| 561 | Ofloxacin 200mg tablets | metronidazole |
| 76099 | Ofloxacin 200mg tablets (A A H Pharmaceuticals Ltd) | metronidazole |
| 34523 | Ofloxacin 200mg tablets (Sandoz Ltd) | metronidazole |
| 34541 | Ofloxacin 200mg tablets (Teva UK Ltd) | metronidazole |
| 40252 | Ofloxacin 200mg/100ml solution for infusion bottles | metronidazole |
| 29280 | Ofloxacin 2mg/ml Infusion | metronidazole |
| 566 | Ofloxacin 400mg tablets | metronidazole |
| 45263 | Ofloxacin 400mg tablets (A A H Pharmaceuticals Ltd) | metronidazole |
| 34819 | Ofloxacin 400mg tablets (Mylan) | metronidazole |
| 34391 | Ofloxacin 400mg tablets (Sandoz Ltd) | metronidazole |
| 33707 | Ofloxacin 400mg tablets (Teva UK Ltd) | metronidazole |
| 78601 | Quinsair 240mg nebuliser solution ampoules (Chiesi Ltd) | metronidazole |
| 35777 | Rosoxacin 150mg capsule | metronidazole |
| 25901 | Sparfloxacin 200mg tablet | metronidazole |
| 4513 | Tarivid 200mg tablets (Sanofi) | metronidazole |
| 66317 | Tarivid 200mg/100ml solution for infusion bottles (Sanofi) | metronidazole |
| 30877 | Tarivid 2mg/ml Infusion (Aventis Pharma) | metronidazole |
| 2726 | Tarivid 400mg tablets (Sanofi) | metronidazole |
| 17693 | Tavanic 250mg tablets (Sanofi) | metronidazole |
| 6206 | Tavanic 500mg tablets (Sanofi) | metronidazole |
| 24373 | Tavanic 500mg/100ml solution for infusion vials (Sanofi) | metronidazole |
| 17272 | Teflox 300mg Tablet (Abbott Laboratories Ltd) | metronidazole |
| 17749 | Teflox 400mg Tablet (Abbott Laboratories Ltd) | metronidazole |
| 18661 | Temafloxacin 400mg tablets | metronidazole |
| 21147 | Uriben 300mg/5ml oral suspension (Rosemont Pharmaceuticals Ltd) | metronidazole |
| 2253 | Utinor 400mg tablets (Merck Sharp & Dohme Ltd) | metronidazole |
| 78256 | Fosfomycin 250mg/5ml oral suspension | metronidazole |
| 26113 | Fosfomycin 2g Sachets | metronidazole |
| 21487 | Fosfomycin 3g granules sachets | metronidazole |
| 71169 | Fosfomycin 3g granules sachets (Advanz Pharma) | metronidazole |
| 55112 | Fosfomycin 500mg capsules | metronidazole |
| 2541 | Furadantin 100mg tablets (Advanz Pharma) | metronidazole |
| 272 | Furadantin 25mg/5ml oral suspension (Mercury Pharma Group Ltd) | metronidazole |
| 2023 | Furadantin 50mg tablets (Advanz Pharma) | metronidazole |
| 67981 | Genfura 100mg tablets (Genesis Pharmaceuticals Ltd) | metronidazole |
| 69434 | Genfura 50mg tablets (Genesis Pharmaceuticals Ltd) | metronidazole |
| 4282 | Hiprex 1g tablets (Mylan) | metronidazole |
| 7525 | Macrobid 100mg modified-release capsules (Advanz Pharma) | metronidazole |
| 65803 | Macrobid 100mg modified-release capsules (Waymade Healthcare Plc) | metronidazole |
| 2036 | Macrodantin 100mg capsules (Advanz Pharma) | metronidazole |
| 1825 | Macrodantin 50mg capsules (Advanz Pharma) | metronidazole |
| 65251 | Macrodantin 50mg capsules (Waymade Healthcare Plc) | metronidazole |
| 4290 | Methenamine hippurate 1g tablets | metronidazole |
| 29795 | Methenamine hippurate 300mg Tablet | metronidazole |
| 27986 | Monuril 2g Paediatric sachet (Pharmax Ltd) | metronidazole |
| 64501 | Monuril 3g granules sachets (Lexon (UK) Ltd) | metronidazole |
| 68216 | Monuril 3g granules sachets (Profile Pharma Ltd) | metronidazole |
| 12379 | Monuril 3g Sachets (Pharmax Ltd) | metronidazole |
| 466 | Nitrofurantoin 100mg capsules | metronidazole |
| 75133 | Nitrofurantoin 100mg capsules (A A H Pharmaceuticals Ltd) | metronidazole |
| 60713 | Nitrofurantoin 100mg capsules (Advanz Pharma) | metronidazole |
| 61642 | Nitrofurantoin 100mg capsules (Alliance Healthcare (Distribution) Ltd) | metronidazole |
| 76738 | Nitrofurantoin 100mg capsules (Morningside Healthcare Ltd) | metronidazole |
| 79573 | Nitrofurantoin 100mg capsules (Tillomed Laboratories Ltd) | metronidazole |
| 6370 | Nitrofurantoin 100mg modified-release capsules | metronidazole |
| 2887 | Nitrofurantoin 100mg tablets | metronidazole |
| 35850 | Nitrofurantoin 100mg tablets (A A H Pharmaceuticals Ltd) | metronidazole |
| 41397 | Nitrofurantoin 100mg tablets (Actavis UK Ltd) | metronidazole |
| 71315 | Nitrofurantoin 100mg tablets (Alliance Healthcare (Distribution) Ltd) | metronidazole |
| 76427 | Nitrofurantoin 100mg tablets (Genesis Pharmaceuticals Ltd) | metronidazole |
| 70380 | Nitrofurantoin 100mg tablets (Mawdsley-Brooks & Company Ltd) | metronidazole |
| 67762 | Nitrofurantoin 100mg tablets (Mylan) | metronidazole |
| 74317 | Nitrofurantoin 100mg tablets (Phoenix Healthcare Distribution Ltd) | metronidazole |
| 74369 | Nitrofurantoin 100mg tablets (Sigma Pharmaceuticals Plc) | metronidazole |
| 53638 | Nitrofurantoin 100mg tablets (Teva UK Ltd) | metronidazole |
| 75209 | Nitrofurantoin 100mg tablets (Waymade Healthcare Plc) | metronidazole |
| 64690 | Nitrofurantoin 100mg/5ml oral solution | metronidazole |
| 71845 | Nitrofurantoin 100mg/5ml oral suspension | metronidazole |
| 73593 | Nitrofurantoin 10mg/5ml oral solution | metronidazole |
| 75366 | Nitrofurantoin 10mg/5ml oral suspension | metronidazole |
| 75365 | Nitrofurantoin 12.5mg/5ml oral suspension | metronidazole |
| 79614 | Nitrofurantoin 18mg/5ml oral solution | metronidazole |
| 75257 | Nitrofurantoin 1mg/5ml oral solution | metronidazole |
| 65207 | Nitrofurantoin 24mg/5ml oral suspension | metronidazole |
| 56621 | Nitrofurantoin 25mg/5ml oral solution | metronidazole |
| 2198 | Nitrofurantoin 25mg/5ml Oral suspension | metronidazole |
| 53659 | Nitrofurantoin 25mg/5ml oral suspension | metronidazole |
| 48353 | Nitrofurantoin 25mg/5ml oral suspension sugar free | metronidazole |
| 35673 | Nitrofurantoin 25mg/5ml oral suspension sugar free (Advanz Pharma) | metronidazole |
| 69479 | Nitrofurantoin 25mg/5ml oral suspension sugar free (Alliance Healthcare (Distribution) Ltd) | metronidazole |
| 64389 | Nitrofurantoin 30mg/5ml oral solution | metronidazole |
| 68651 | Nitrofurantoin 30mg/5ml oral suspension | metronidazole |
| 60795 | Nitrofurantoin 35mg/5ml oral solution | metronidazole |
| 51726 | Nitrofurantoin 40mg/5ml oral suspension | metronidazole |
| 79415 | Nitrofurantoin 5.5mg/5ml oral suspension | metronidazole |
| 210 | Nitrofurantoin 50mg capsules | metronidazole |
| 63588 | Nitrofurantoin 50mg capsules (A A H Pharmaceuticals Ltd) | metronidazole |
| 79598 | Nitrofurantoin 50mg capsules (Actavis UK Ltd) | metronidazole |
| 60252 | Nitrofurantoin 50mg capsules (Advanz Pharma) | metronidazole |
| 61907 | Nitrofurantoin 50mg capsules (Alliance Healthcare (Distribution) Ltd) | metronidazole |
| 62647 | Nitrofurantoin 50mg Tablet (Biorex Laboratories Ltd) | metronidazole |
| 778 | Nitrofurantoin 50mg tablets | metronidazole |
| 53094 | Nitrofurantoin 50mg tablets (A A H Pharmaceuticals Ltd) | metronidazole |
| 40164 | Nitrofurantoin 50mg tablets (Actavis UK Ltd) | metronidazole |
| 51959 | Nitrofurantoin 50mg tablets (Alliance Healthcare (Distribution) Ltd) | metronidazole |
| 66013 | Nitrofurantoin 50mg tablets (Almus Pharmaceuticals Ltd) | metronidazole |
| 53171 | Nitrofurantoin 50mg tablets (Dr Reddy's Laboratories (UK) Ltd) | metronidazole |
| 57669 | Nitrofurantoin 50mg tablets (Genesis Pharmaceuticals Ltd) | metronidazole |
| 67759 | Nitrofurantoin 50mg tablets (Mylan) | metronidazole |
| 54325 | Nitrofurantoin 50mg tablets (Phoenix Healthcare Distribution Ltd) | metronidazole |
| 70796 | Nitrofurantoin 50mg tablets (Teva UK Ltd) | metronidazole |
| 57779 | Nitrofurantoin 50mg tablets (Waymade Healthcare Plc) | metronidazole |
| 75997 | Nitrofurantoin 50mg/5ml oral solution | metronidazole |
| 73590 | Nitrofurantoin 50mg/5ml oral suspension | metronidazole |
| 58469 | Nitrofurantoin 5mg/5ml oral solution | metronidazole |
| 57248 | Nitrofurantoin 5mg/5ml oral suspension | metronidazole |
| 78391 | Nitrofurantoin 75mg/5ml oral suspension | metronidazole |
| 59497 | Nitrofurantoin 7mg/5ml oral suspension | metronidazole |
| 73557 | Nitrofurantoin 8mg/5ml oral solution | metronidazole |
| 67095 | Nitrofurantoin 9mg/5ml oral suspension | metronidazole |
| 12893 | Phenazopyridine 100mg tablet | metronidazole |
| 30619 | Pyridium 100mg Tablet (Pfizer Consumer Healthcare Ltd) | metronidazole |
| 16284 | Urantoin 100mg tablets (Dr Reddy's Laboratories (UK) Ltd) | metronidazole |
| 78209 | Urantoin 50mg tablets (Dr Reddy's Laboratories (UK) Ltd) | metronidazole |
| 22016 | Almodan 125mg/5ml Oral solution (Berk Pharmaceuticals Ltd) | penicillins |
| 17282 | Almodan 125mg/5ml syrup (Teva UK Ltd) | penicillins |
| 21799 | Almodan 250mg Capsule (Berk Pharmaceuticals Ltd) | penicillins |
| 21963 | Almodan 250mg/5ml Oral solution (Berk Pharmaceuticals Ltd) | penicillins |
| 21845 | Almodan 250mg/5ml Oral solution (Berk Pharmaceuticals Ltd) | penicillins |
| 21827 | Almodan 500mg Capsule (Berk Pharmaceuticals Ltd) | penicillins |
| 12489 | Ambaxin 400mg Tablet (Pharmacia Ltd) | penicillins |
| 32347 | Amfipen 125mg/5ml Oral solution (Yamanouchi Pharma Ltd) | penicillins |
| 20531 | Amfipen 250mg Capsule (Yamanouchi Pharma Ltd) | penicillins |
| 26356 | Amfipen 500mg Capsule (Yamanouchi Pharma Ltd) | penicillins |
| 21926 | Amfipen 500mg/vial Injection (Yamanouchi Pharma Ltd) | penicillins |
| 32148 | Amfipen forte 250mg/5ml Oral solution (Yamanouchi Pharma Ltd) | penicillins |
| 22029 | Amiclav 250mg/125mg tablets (Ashbourne Pharmaceuticals Ltd) | penicillins |
| 11634 | Amix 125 oral suspension (Ashbourne Pharmaceuticals Ltd) | penicillins |
| 11613 | Amix 250 capsules (Ashbourne Pharmaceuticals Ltd) | penicillins |
| 21844 | Amix 250 oral suspension (Ashbourne Pharmaceuticals Ltd) | penicillins |
| 18786 | Amix 500 capsules (Ashbourne Pharmaceuticals Ltd) | penicillins |
| 29697 | Amopen 125mg/5ml Liquid (Yorkshire Pharmaceuticals Ltd) | penicillins |
| 30498 | Amopen 250mg Capsule (Yorkshire Pharmaceuticals Ltd) | penicillins |
| 31423 | Amopen 250mg/5ml Liquid (Yorkshire Pharmaceuticals Ltd) | penicillins |
| 17711 | Amopen 500mg Capsule (Yorkshire Pharmaceuticals Ltd) | penicillins |
| 12378 | Amoram 125mg/5ml oral suspension (LPC Medical (UK) Ltd) | penicillins |
| 9243 | Amoram 250mg capsules (LPC Medical (UK) Ltd) | penicillins |
| 22438 | Amoram 250mg/5ml oral suspension (LPC Medical (UK) Ltd) | penicillins |
| 22415 | Amoram 500mg capsules (LPC Medical (UK) Ltd) | penicillins |
| 29474 | Amoxicillin 1000mg with clavulanic acid 100mg/vial injection | penicillins |
| 13285 | Amoxicillin 125mg / Clavulanic acid 31mg/5ml oral suspension | penicillins |
| 8906 | Amoxicillin 125mg / Clavulanic acid 31mg/5ml oral suspension | penicillins |
| 53942 | Amoxicillin 125mg / Clavulanic acid 62.5mg/5ml oral suspension | penicillins |
| 41835 | Amoxicillin 125mg Powder (IVAX Pharmaceuticals UK Ltd) | penicillins |
| 3742 | Amoxicillin 125mg sugar free chewable tablets | penicillins |
| 13848 | Amoxicillin 125mg sugar free powder | penicillins |
| 485 | Amoxicillin 125mg/1.25ml oral suspension paediatric | penicillins |
| 42822 | Amoxicillin 125mg/5ml Mixture (Celltech Pharma Europe Ltd) | penicillins |
| 28872 | Amoxicillin 125mg/5ml Mixture (Crosspharma Ltd) | penicillins |
| 69340 | Amoxicillin 125mg/5ml Mixture (Mepra-Pharm) | penicillins |
| 41818 | Amoxicillin 125mg/5ml Oral solution (Berk Pharmaceuticals Ltd) | penicillins |
| 42240 | Amoxicillin 125mg/5ml Oral solution (Co-Pharma Ltd) | penicillins |
| 29337 | Amoxicillin 125mg/5ml Oral solution (Neo Laboratories Ltd) | penicillins |
| 62 | Amoxicillin 125mg/5ml oral suspension | penicillins |
| 33690 | Amoxicillin 125mg/5ml oral suspension (A A H Pharmaceuticals Ltd) | penicillins |
| 34857 | Amoxicillin 125mg/5ml oral suspension (Actavis UK Ltd) | penicillins |
| 61207 | Amoxicillin 125mg/5ml oral suspension (Alliance Healthcare (Distribution) Ltd) | penicillins |
| 42545 | Amoxicillin 125mg/5ml oral suspension (Almus Pharmaceuticals Ltd) | penicillins |
| 50002 | Amoxicillin 125mg/5ml oral suspension (Bristol Laboratories Ltd) | penicillins |
| 63582 | Amoxicillin 125mg/5ml oral suspension (Crescent Pharma Ltd) | penicillins |
| 59391 | Amoxicillin 125mg/5ml oral suspension (DE Pharmaceuticals) | penicillins |
| 23238 | Amoxicillin 125mg/5ml oral suspension (IVAX Pharmaceuticals UK Ltd) | penicillins |
| 48038 | Amoxicillin 125mg/5ml oral suspension (Kent Pharmaceuticals Ltd) | penicillins |
| 32622 | Amoxicillin 125mg/5ml oral suspension (Mylan) | penicillins |
| 52685 | Amoxicillin 125mg/5ml oral suspension (Phoenix Healthcare Distribution Ltd) | penicillins |
| 28875 | Amoxicillin 125mg/5ml oral suspension (Ranbaxy (UK) Ltd) | penicillins |
| 43229 | Amoxicillin 125mg/5ml Oral suspension (Sandoz Ltd) | penicillins |
| 55047 | Amoxicillin 125mg/5ml oral suspension (Sandoz Ltd) | penicillins |
| 64355 | Amoxicillin 125mg/5ml oral suspension (Sigma Pharmaceuticals Plc) | penicillins |
| 28870 | Amoxicillin 125mg/5ml oral suspension (Teva UK Ltd) | penicillins |
| 56561 | Amoxicillin 125mg/5ml oral suspension (Waymade Healthcare Plc) | penicillins |
| 503 | Amoxicillin 125mg/5ml oral suspension sugar free | penicillins |
| 33696 | Amoxicillin 125mg/5ml oral suspension sugar free (A A H Pharmaceuticals Ltd) | penicillins |
| 34679 | Amoxicillin 125mg/5ml oral suspension sugar free (Actavis UK Ltd) | penicillins |
| 53078 | Amoxicillin 125mg/5ml oral suspension sugar free (Alliance Healthcare (Distribution) Ltd) | penicillins |
| 36054 | Amoxicillin 125mg/5ml oral suspension sugar free (Almus Pharmaceuticals Ltd) | penicillins |
| 69901 | Amoxicillin 125mg/5ml oral suspension sugar free (Arrow Generics Ltd) | penicillins |
| 52122 | Amoxicillin 125mg/5ml oral suspension sugar free (Bristol Laboratories Ltd) | penicillins |
| 59112 | Amoxicillin 125mg/5ml oral suspension sugar free (DE Pharmaceuticals) | penicillins |
| 74365 | Amoxicillin 125mg/5ml oral suspension sugar free (Dowelhurst Ltd) | penicillins |
| 24150 | Amoxicillin 125mg/5ml oral suspension sugar free (IVAX Pharmaceuticals UK Ltd) | penicillins |
| 34384 | Amoxicillin 125mg/5ml oral suspension sugar free (Kent Pharmaceuticals Ltd) | penicillins |
| 66062 | Amoxicillin 125mg/5ml oral suspension sugar free (Mawdsley-Brooks & Company Ltd) | penicillins |
| 75961 | Amoxicillin 125mg/5ml oral suspension sugar free (Medreich Plc) | penicillins |
| 31014 | Amoxicillin 125mg/5ml oral suspension sugar free (Mylan) | penicillins |
| 52857 | Amoxicillin 125mg/5ml oral suspension sugar free (Phoenix Healthcare Distribution Ltd) | penicillins |
| 29858 | Amoxicillin 125mg/5ml oral suspension sugar free (Sandoz Ltd) | penicillins |
| 74550 | Amoxicillin 125mg/5ml oral suspension sugar free (Sigma Pharmaceuticals Plc) | penicillins |
| 34638 | Amoxicillin 125mg/5ml oral suspension sugar free (Teva UK Ltd) | penicillins |
| 55626 | Amoxicillin 125mg/5ml oral suspension sugar free (Waymade Healthcare Plc) | penicillins |
| 17509 | Amoxicillin 1g powder for solution for injection vials | penicillins |
| 68545 | Amoxicillin 1g powder for solution for injection vials (A A H Pharmaceuticals Ltd) | penicillins |
| 34238 | Amoxicillin 1g powder for solution for injection vials (Wockhardt UK Ltd) | penicillins |
| 1391 | Amoxicillin 250mg / Clavulanic acid 125mg tablets | penicillins |
| 7636 | Amoxicillin 250mg / Clavulanic acid 62mg/5ml oral suspension | penicillins |
| 13262 | Amoxicillin 250mg / Clavulanic acid 62mg/5ml oral suspension | penicillins |
| 42809 | Amoxicillin 250mg Capsule (C P Pharmaceuticals Ltd) | penicillins |
| 79359 | Amoxicillin 250mg Capsule (Celltech Pharma Europe Ltd) | penicillins |
| 31661 | Amoxicillin 250mg Capsule (Co-Pharma Ltd) | penicillins |
| 28882 | Amoxicillin 250mg Capsule (Crosspharma Ltd) | penicillins |
| 34435 | Amoxicillin 250mg Capsule (DDSA Pharmaceuticals Ltd) | penicillins |
| 33222 | Amoxicillin 250mg Capsule (Lagap) | penicillins |
| 32872 | Amoxicillin 250mg Capsule (Mepra-Pharm) | penicillins |
| 34714 | Amoxicillin 250mg Capsule (Neo Laboratories Ltd) | penicillins |
| 45267 | Amoxicillin 250mg Capsule (Regent Laboratories Ltd) | penicillins |
| 9 | Amoxicillin 250mg capsules | penicillins |
| 25484 | Amoxicillin 250mg capsules (A A H Pharmaceuticals Ltd) | penicillins |
| 59432 | Amoxicillin 250mg capsules (Accord Healthcare Ltd) | penicillins |
| 33343 | Amoxicillin 250mg capsules (Actavis UK Ltd) | penicillins |
| 59042 | Amoxicillin 250mg capsules (Alliance Healthcare (Distribution) Ltd) | penicillins |
| 63911 | Amoxicillin 250mg capsules (Almus Pharmaceuticals Ltd) | penicillins |
| 54796 | Amoxicillin 250mg capsules (Boston Healthcare Ltd) | penicillins |
| 54491 | Amoxicillin 250mg capsules (Bristol Laboratories Ltd) | penicillins |
| 69711 | Amoxicillin 250mg capsules (Brown & Burk UK Ltd) | penicillins |
| 58771 | Amoxicillin 250mg capsules (DE Pharmaceuticals) | penicillins |
| 77924 | Amoxicillin 250mg capsules (Flamingo Pharma (UK) Ltd) | penicillins |
| 34042 | Amoxicillin 250mg capsules (IVAX Pharmaceuticals UK Ltd) | penicillins |
| 30528 | Amoxicillin 250mg capsules (Kent Pharmaceuticals Ltd) | penicillins |
| 54271 | Amoxicillin 250mg capsules (Mawdsley-Brooks & Company Ltd) | penicillins |
| 71609 | Amoxicillin 250mg capsules (Mawdsley-Brooks & Company Ltd) | penicillins |
| 57966 | Amoxicillin 250mg capsules (Medreich Plc) | penicillins |
| 51536 | Amoxicillin 250mg capsules (Milpharm Ltd) | penicillins |
| 30745 | Amoxicillin 250mg capsules (Mylan) | penicillins |
| 59481 | Amoxicillin 250mg capsules (Phoenix Healthcare Distribution Ltd) | penicillins |
| 30743 | Amoxicillin 250mg capsules (Ranbaxy (UK) Ltd) | penicillins |
| 79767 | Amoxicillin 250mg capsules (RX Farma) | penicillins |
| 48006 | Amoxicillin 250mg capsules (Sandoz Ltd) | penicillins |
| 64794 | Amoxicillin 250mg capsules (Sigma Pharmaceuticals Plc) | penicillins |
| 23967 | Amoxicillin 250mg capsules (Teva UK Ltd) | penicillins |
| 59153 | Amoxicillin 250mg capsules (Waymade Healthcare Plc) | penicillins |
| 54185 | Amoxicillin 250mg capsules (Wockhardt UK Ltd) | penicillins |
| 598 | Amoxicillin 250mg powder for solution for injection vials | penicillins |
| 76434 | Amoxicillin 250mg powder for solution for injection vials (A A H Pharmaceuticals Ltd) | penicillins |
| 62786 | Amoxicillin 250mg powder for solution for injection vials (Wockhardt UK Ltd) | penicillins |
| 870 | Amoxicillin 250mg sugar free chewable tablets | penicillins |
| 42815 | Amoxicillin 250mg/5ml Mixture (Celltech Pharma Europe Ltd) | penicillins |
| 33570 | Amoxicillin 250mg/5ml Mixture (Crosspharma Ltd) | penicillins |
| 40238 | Amoxicillin 250mg/5ml Mixture (Mepra-Pharm) | penicillins |
| 45317 | Amoxicillin 250mg/5ml Oral solution (Neo Laboratories Ltd) | penicillins |
| 427 | Amoxicillin 250mg/5ml oral suspension | penicillins |
| 33165 | Amoxicillin 250mg/5ml oral suspension (A A H Pharmaceuticals Ltd) | penicillins |
| 34760 | Amoxicillin 250mg/5ml oral suspension (Actavis UK Ltd) | penicillins |
| 62074 | Amoxicillin 250mg/5ml oral suspension (Alliance Healthcare (Distribution) Ltd) | penicillins |
| 41090 | Amoxicillin 250mg/5ml oral suspension (Almus Pharmaceuticals Ltd) | penicillins |
| 55018 | Amoxicillin 250mg/5ml oral suspension (Bristol Laboratories Ltd) | penicillins |
| 65031 | Amoxicillin 250mg/5ml oral suspension (Crescent Pharma Ltd) | penicillins |
| 60267 | Amoxicillin 250mg/5ml oral suspension (DE Pharmaceuticals) | penicillins |
| 32640 | Amoxicillin 250mg/5ml oral suspension (IVAX Pharmaceuticals UK Ltd) | penicillins |
| 62762 | Amoxicillin 250mg/5ml oral suspension (Kent Pharmaceuticals Ltd) | penicillins |
| 33689 | Amoxicillin 250mg/5ml oral suspension (Mylan) | penicillins |
| 51382 | Amoxicillin 250mg/5ml oral suspension (Phoenix Healthcare Distribution Ltd) | penicillins |
| 55499 | Amoxicillin 250mg/5ml oral suspension (Ranbaxy (UK) Ltd) | penicillins |
| 37755 | Amoxicillin 250mg/5ml Oral suspension (Sandoz Ltd) | penicillins |
| 56223 | Amoxicillin 250mg/5ml oral suspension (Sandoz Ltd) | penicillins |
| 53924 | Amoxicillin 250mg/5ml oral suspension (Sigma Pharmaceuticals Plc) | penicillins |
| 27725 | Amoxicillin 250mg/5ml oral suspension (Teva UK Ltd) | penicillins |
| 62102 | Amoxicillin 250mg/5ml oral suspension (Waymade Healthcare Plc) | penicillins |
| 585 | Amoxicillin 250mg/5ml oral suspension sugar free | penicillins |
| 34232 | Amoxicillin 250mg/5ml oral suspension sugar free (A A H Pharmaceuticals Ltd) | penicillins |
| 40243 | Amoxicillin 250mg/5ml oral suspension sugar free (Actavis UK Ltd) | penicillins |
| 54222 | Amoxicillin 250mg/5ml oral suspension sugar free (Alliance Healthcare (Distribution) Ltd) | penicillins |
| 42732 | Amoxicillin 250mg/5ml oral suspension sugar free (Almus Pharmaceuticals Ltd) | penicillins |
| 49065 | Amoxicillin 250mg/5ml oral suspension sugar free (Bristol Laboratories Ltd) | penicillins |
| 60027 | Amoxicillin 250mg/5ml oral suspension sugar free (DE Pharmaceuticals) | penicillins |
| 74309 | Amoxicillin 250mg/5ml oral suspension sugar free (Dowelhurst Ltd) | penicillins |
| 33699 | Amoxicillin 250mg/5ml oral suspension sugar free (IVAX Pharmaceuticals UK Ltd) | penicillins |
| 34855 | Amoxicillin 250mg/5ml oral suspension sugar free (Kent Pharmaceuticals Ltd) | penicillins |
| 78701 | Amoxicillin 250mg/5ml oral suspension sugar free (Medreich Plc) | penicillins |
| 31535 | Amoxicillin 250mg/5ml oral suspension sugar free (Mylan) | penicillins |
| 58053 | Amoxicillin 250mg/5ml oral suspension sugar free (Phoenix Healthcare Distribution Ltd) | penicillins |
| 73979 | Amoxicillin 250mg/5ml oral suspension sugar free (RX Farma) | penicillins |
| 58057 | Amoxicillin 250mg/5ml oral suspension sugar free (Sandoz Ltd) | penicillins |
| 65095 | Amoxicillin 250mg/5ml oral suspension sugar free (Sigma Pharmaceuticals Plc) | penicillins |
| 34775 | Amoxicillin 250mg/5ml oral suspension sugar free (Teva UK Ltd) | penicillins |
| 62442 | Amoxicillin 250mg/5ml oral suspension sugar free (Waymade Healthcare Plc) | penicillins |
| 17746 | Amoxicillin 375mg soluble tablets | penicillins |
| 1140 | Amoxicillin 3g oral powder sachets sugar free | penicillins |
| 33383 | Amoxicillin 3g oral powder sachets sugar free (A A H Pharmaceuticals Ltd) | penicillins |
| 70275 | Amoxicillin 3g oral powder sachets sugar free (Brown & Burk UK Ltd) | penicillins |
| 40168 | Amoxicillin 3g oral powder sachets sugar free (Kent Pharmaceuticals Ltd) | penicillins |
| 57178 | Amoxicillin 3g oral powder sachets sugar free (Mawdsley-Brooks & Company Ltd) | penicillins |
| 28130 | Amoxicillin 3g oral powder sachets sugar free (Teva UK Ltd) | penicillins |
| 74937 | Amoxicillin 3g oral powder sachets sugar free (Waymade Healthcare Plc) | penicillins |
| 41734 | Amoxicillin 3g Powder (Actavis UK Ltd) | penicillins |
| 15192 | Amoxicillin 400mg / Clavulanic acid 57mg/5ml sugar free oral suspension | penicillins |
| 5662 | Amoxicillin 500mg / Clarithromycin 500mg / Lansoprazole 30mg triple pack | penicillins |
| 13216 | Amoxicillin 500mg / Clavulanic acid 125mg tablets | penicillins |
| 38684 | Amoxicillin 500mg Capsule (C P Pharmaceuticals Ltd) | penicillins |
| 35570 | Amoxicillin 500mg Capsule (Crosspharma Ltd) | penicillins |
| 34885 | Amoxicillin 500mg Capsule (DDSA Pharmaceuticals Ltd) | penicillins |
| 44854 | Amoxicillin 500mg Capsule (Lagap) | penicillins |
| 34912 | Amoxicillin 500mg Capsule (Neo Laboratories Ltd) | penicillins |
| 48 | Amoxicillin 500mg capsules | penicillins |
| 33692 | Amoxicillin 500mg capsules (A A H Pharmaceuticals Ltd) | penicillins |
| 53627 | Amoxicillin 500mg capsules (Accord Healthcare Ltd) | penicillins |
| 26157 | Amoxicillin 500mg capsules (Actavis UK Ltd) | penicillins |
| 52820 | Amoxicillin 500mg capsules (Alliance Healthcare (Distribution) Ltd) | penicillins |
| 47640 | Amoxicillin 500mg capsules (Almus Pharmaceuticals Ltd) | penicillins |
| 55527 | Amoxicillin 500mg capsules (Boston Healthcare Ltd) | penicillins |
| 52771 | Amoxicillin 500mg capsules (Bristol Laboratories Ltd) | penicillins |
| 69118 | Amoxicillin 500mg capsules (Brown & Burk UK Ltd) | penicillins |
| 59879 | Amoxicillin 500mg capsules (DE Pharmaceuticals) | penicillins |
| 79240 | Amoxicillin 500mg capsules (Flamingo Pharma (UK) Ltd) | penicillins |
| 29463 | Amoxicillin 500mg capsules (IVAX Pharmaceuticals UK Ltd) | penicillins |
| 33706 | Amoxicillin 500mg capsules (Kent Pharmaceuticals Ltd) | penicillins |
| 76347 | Amoxicillin 500mg capsules (Mawdsley-Brooks & Company Ltd) | penicillins |
| 61906 | Amoxicillin 500mg capsules (Mawdsley-Brooks & Company Ltd) | penicillins |
| 52058 | Amoxicillin 500mg capsules (Medreich Plc) | penicillins |
| 54725 | Amoxicillin 500mg capsules (Milpharm Ltd) | penicillins |
| 23740 | Amoxicillin 500mg capsules (Mylan) | penicillins |
| 78078 | Amoxicillin 500mg capsules (Noumed Life Sciences Ltd) | penicillins |
| 59592 | Amoxicillin 500mg capsules (Pfizer Ltd) | penicillins |
| 68416 | Amoxicillin 500mg capsules (Phoenix Healthcare Distribution Ltd) | penicillins |
| 34852 | Amoxicillin 500mg capsules (Ranbaxy (UK) Ltd) | penicillins |
| 76233 | Amoxicillin 500mg capsules (RX Farma) | penicillins |
| 31801 | Amoxicillin 500mg capsules (Sandoz Ltd) | penicillins |
| 65958 | Amoxicillin 500mg capsules (Sigma Pharmaceuticals Plc) | penicillins |
| 34001 | Amoxicillin 500mg capsules (Teva UK Ltd) | penicillins |
| 64357 | Amoxicillin 500mg capsules (Waymade Healthcare Plc) | penicillins |
| 55394 | Amoxicillin 500mg capsules (Wockhardt UK Ltd) | penicillins |
| 1722 | Amoxicillin 500mg dispersible tablets | penicillins |
| 1746 | Amoxicillin 500mg powder for solution for injection vials | penicillins |
| 58205 | Amoxicillin 500mg powder for solution for injection vials (A A H Pharmaceuticals Ltd) | penicillins |
| 33840 | Amoxicillin 500mg powder for solution for injection vials (Wockhardt UK Ltd) | penicillins |
| 2281 | Amoxicillin 500mg sugar free chewable tablets | penicillins |
| 28592 | Amoxicillin 500mg with clavulanic acid 100mg/vial injection | penicillins |
| 73510 | Amoxicillin 500mg/50ml infusion bags | penicillins |
| 4582 | Amoxicillin 750mg soluble tablets | penicillins |
| 9343 | Amoxicillin 750mg sugar free powder | penicillins |
| 72664 | Amoxicillin 875mg / Clavulanic acid 125mg tablets | penicillins |
| 439 | Amoxicillin with Clavulanic acid dispersible tablets | penicillins |
| 24288 | AMOXIL | penicillins |
| 26519 | AMOXIL | penicillins |
| 2171 | Amoxil 125mg/1.25ml paediatric oral suspension (GlaxoSmithKline UK Ltd) | penicillins |
| 2153 | Amoxil 125mg/5ml syrup sucrose free (GlaxoSmithKline UK Ltd) | penicillins |
| 17099 | Amoxil 1g powder for solution for injection vials (GlaxoSmithKline UK Ltd) | penicillins |
| 133 | Amoxil 250mg capsules (GlaxoSmithKline UK Ltd) | penicillins |
| 10771 | Amoxil 250mg powder for solution for injection vials (GlaxoSmithKline UK Ltd) | penicillins |
| 1812 | Amoxil 250mg/5ml syrup sucrose free (GlaxoSmithKline UK Ltd) | penicillins |
| 2174 | Amoxil 3g oral powder sachets sucrose free (GlaxoSmithKline UK Ltd) | penicillins |
| 69532 | Amoxil 500mg capsules (DE Pharmaceuticals) | penicillins |
| 847 | Amoxil 500mg capsules (GlaxoSmithKline UK Ltd) | penicillins |
| 49590 | Amoxil 500mg capsules (Lexon (UK) Ltd) | penicillins |
| 51436 | Amoxil 500mg capsules (Mawdsley-Brooks & Company Ltd) | penicillins |
| 56700 | Amoxil 500mg capsules (Necessity Supplies Ltd) | penicillins |
| 68476 | Amoxil 500mg capsules (Sigma Pharmaceuticals Plc) | penicillins |
| 57886 | Amoxil 500mg capsules (Stephar (U.K.) Ltd) | penicillins |
| 57833 | Amoxil 500mg capsules (Waymade Healthcare Plc) | penicillins |
| 15148 | Amoxil 500mg Dispersible tablet (SmithKline Beecham Plc) | penicillins |
| 24819 | Amoxil 500mg powder for solution for injection vials (GlaxoSmithKline UK Ltd) | penicillins |
| 4010 | Amoxil 750mg Sachets (GlaxoSmithKline UK Ltd) | penicillins |
| 4154 | Amoxil fiztab 125mg Tablet (Bencard) | penicillins |
| 1637 | Amoxil fiztab 250mg Tablet (Bencard) | penicillins |
| 7737 | Amoxil fiztab 500mg Tablet (Bencard) | penicillins |
| 19593 | AMOXIL PAED 125MG IN 1.25ML | penicillins |
| 29061 | AMOXIL SF | penicillins |
| 966 | AMOXIL SF 125 MG/5ML SYR | penicillins |
| 22469 | AMOXYCILLIN 125mg/31mg CLAVULANIC ACID | penicillins |
| 25034 | AMOXYCILLIN 125mg/62mg CLAVULANIC ACID | penicillins |
| 7581 | AMOXYCILLIN 125MG/62MG CLAVULANIC ACID SYR | penicillins |
| 27886 | AMOXYCILLIN 250/CLAVULANIC ACID 125 DISP | penicillins |
| 19795 | AMOXYCILLIN 250MG/CLAVULANIC ACID 125MG | penicillins |
| 31286 | Amoxymed 125mg/5ml Oral solution (Medipharma Ltd) | penicillins |
| 3669 | Amoxymed 250mg Capsule (Medipharma Ltd) | penicillins |
| 10755 | Ampiciilin 60mg / Cloxacillin 30mg/0.6ml sugar free oral suspension | penicillins |
| 22482 | AMPICILLIN | penicillins |
| 14484 | Ampicillin / Cloxacillin 500mg capsules | penicillins |
| 108 | AMPICILLIN 125 MG CAP | penicillins |
| 343 | AMPICILLIN 125 MG TAB | penicillins |
| 10369 | Ampicillin 125mg / Flucloxacillin 125mg oral solution | penicillins |
| 55846 | Ampicillin 125mg/5ml Liquid (C P Pharmaceuticals Ltd) | penicillins |
| 857 | Ampicillin 125mg/5ml oral suspension | penicillins |
| 41744 | Ampicillin 125mg/5ml oral suspension (A A H Pharmaceuticals Ltd) | penicillins |
| 46175 | Ampicillin 125mg/5ml Oral suspension (Hillcross Pharmaceuticals Ltd) | penicillins |
| 71092 | Ampicillin 125mg/5ml oral suspension (Phoenix Healthcare Distribution Ltd) | penicillins |
| 8209 | Ampicillin 125mg/5ml paediatric oral suspension | penicillins |
| 900 | Ampicillin 125mg/5ml sugar free suspension | penicillins |
| 22544 | AMPICILLIN 125MG/FLUCLOXACILLIN 125MG | penicillins |
| 28919 | Ampicillin 250mg / Cloxacillin 250mg/vial injection | penicillins |
| 1450 | Ampicillin 250mg / Flucloxacillin 250mg capsules | penicillins |
| 26510 | Ampicillin 250mg / Flucloxacillin 250mg injection | penicillins |
| 41646 | Ampicillin 250mg Capsule (Berk Pharmaceuticals Ltd) | penicillins |
| 115 | Ampicillin 250mg capsules | penicillins |
| 34228 | Ampicillin 250mg capsules (A A H Pharmaceuticals Ltd) | penicillins |
| 58520 | Ampicillin 250mg capsules (Alliance Healthcare (Distribution) Ltd) | penicillins |
| 54471 | Ampicillin 250mg capsules (Kent Pharmaceuticals Ltd) | penicillins |
| 71001 | Ampicillin 250mg capsules (Phoenix Healthcare Distribution Ltd) | penicillins |
| 57997 | Ampicillin 250mg capsules (Waymade Healthcare Plc) | penicillins |
| 10685 | Ampicillin 250mg injection | penicillins |
| 106 | Ampicillin 250mg/5ml oral suspension | penicillins |
| 77655 | Ampicillin 250mg/5ml oral suspension (Alliance Healthcare (Distribution) Ltd) | penicillins |
| 71820 | Ampicillin 250mg/5ml oral suspension (Kent Pharmaceuticals Ltd) | penicillins |
| 67787 | Ampicillin 250mg/5ml oral suspension (Sigma Pharmaceuticals Plc) | penicillins |
| 16167 | Ampicillin 250mg/5ml sugar free suspension | penicillins |
| 19585 | AMPICILLIN 250MG/FLUCLOXACILLIN 250MG | penicillins |
| 24847 | Ampicillin 500mg / Flucloxacillin 500mg injection | penicillins |
| 78153 | Ampicillin 500mg Capsule (IVAX Pharmaceuticals UK Ltd) | penicillins |
| 926 | Ampicillin 500mg capsules | penicillins |
| 26174 | Ampicillin 500mg capsules (A A H Pharmaceuticals Ltd) | penicillins |
| 41647 | Ampicillin 500mg capsules (Actavis UK Ltd) | penicillins |
| 14485 | Ampicillin 500mg powder for solution for injection vials | penicillins |
| 28701 | Ampicillin 50mg / Cloxacillin 25mg/vial injection | penicillins |
| 10795 | Ampiclox 250mg/5ml Oral solution (Beecham Research Laboratories) | penicillins |
| 12382 | Ampiclox 500mg Capsule (Beecham Research Laboratories) | penicillins |
| 14988 | AMPICLOX MG INJ | penicillins |
| 10538 | Ampiclox neonatal 75mg/vial Injection (Beecham Research Laboratories) | penicillins |
| 12083 | Ampiclox neonatal 90mg/0.6ml Oral suspension (Beecham Research Laboratories) | penicillins |
| 32760 | Ampitrin 125mg/5ml Liquid (OPD Pharm) | penicillins |
| 31156 | Ampitrin 250mg Capsule (OPD Pharm) | penicillins |
| 31154 | Ampitrin 500mg Capsule (OPD Pharm) | penicillins |
| 33109 | Amrit 125mg/5ml Liquid (BHR Pharmaceuticals Ltd) | penicillins |
| 27714 | Amrit 250mg Capsule (BHR Pharmaceuticals Ltd) | penicillins |
| 33110 | Amrit 250mg/5ml Liquid (BHR Pharmaceuticals Ltd) | penicillins |
| 33112 | Amrit 500mg Capsule (BHR Pharmaceuticals Ltd) | penicillins |
| 21457 | APSIN VK 125 MG SYR | penicillins |
| 36524 | Apsin vk 125mg/5ml Oral solution (Approved Prescription Services Ltd) | penicillins |
| 25293 | Apsin vk 250mg Tablet (Approved Prescription Services Ltd) | penicillins |
| 22010 | Apsin vk 250mg/5ml Oral solution (Approved Prescription Services Ltd) | penicillins |
| 415 | Augmentin 125/31 SF oral suspension (GlaxoSmithKline UK Ltd) | penicillins |
| 50595 | Augmentin 125/31 SF oral suspension (Mawdsley-Brooks & Company Ltd) | penicillins |
| 51164 | Augmentin 125/31 SF oral suspension (Waymade Healthcare Plc) | penicillins |
| 569 | Augmentin 250/62 SF oral suspension (GlaxoSmithKline UK Ltd) | penicillins |
| 52666 | Augmentin 250/62 SF oral suspension (Sigma Pharmaceuticals Plc) | penicillins |
| 2507 | Augmentin 375mg dispersible tablets (GlaxoSmithKline UK Ltd) | penicillins |
| 49063 | Augmentin 375mg tablets (DE Pharmaceuticals) | penicillins |
| 73983 | Augmentin 375mg tablets (Dowelhurst Ltd) | penicillins |
| 399 | Augmentin 375mg tablets (GlaxoSmithKline UK Ltd) | penicillins |
| 48683 | Augmentin 375mg tablets (Lexon (UK) Ltd) | penicillins |
| 49374 | Augmentin 375mg tablets (Mawdsley-Brooks & Company Ltd) | penicillins |
| 49048 | Augmentin 375mg tablets (Waymade Healthcare Plc) | penicillins |
| 50279 | Augmentin 625mg tablets (DE Pharmaceuticals) | penicillins |
| 509 | Augmentin 625mg tablets (GlaxoSmithKline UK Ltd) | penicillins |
| 49656 | Augmentin 625mg tablets (Lexon (UK) Ltd) | penicillins |
| 52207 | Augmentin 625mg tablets (Mawdsley-Brooks & Company Ltd) | penicillins |
| 49321 | Augmentin 625mg tablets (Sigma Pharmaceuticals Plc) | penicillins |
| 49683 | Augmentin 625mg tablets (Waymade Healthcare Plc) | penicillins |
| 26658 | AUGMENTIN DISPERSIBLE 250/125 | penicillins |
| 244 | Augmentin Intravenous 1.2g powder for solution for injection vials (GlaxoSmithKline UK Ltd) | penicillins |
| 17852 | Augmentin Intravenous 600mg powder for solution for injection vials (GlaxoSmithKline UK Ltd) | penicillins |
| 76854 | Augmentin-Duo 400/57 oral suspension (Dowelhurst Ltd) | penicillins |
| 5341 | Augmentin-Duo 400/57 oral suspension (GlaxoSmithKline UK Ltd) | penicillins |
| 56591 | Augmentin-Duo 400/57 oral suspension (Lexon (UK) Ltd) | penicillins |
| 62597 | Augmentin-Duo 400/57 oral suspension (Mawdsley-Brooks & Company Ltd) | penicillins |
| 51194 | Augmentin-Duo 400/57 oral suspension (Sigma Pharmaceuticals Plc) | penicillins |
| 22453 | Azlocillin 1g/vial injection | penicillins |
| 25515 | Azlocillin 2g/vial injection | penicillins |
| 30423 | Azlocillin 5g/vial infusion | penicillins |
| 17375 | Azlocillin 5g/vial injection | penicillins |
| 21345 | Bacampicillin HCl 400mg tablets | penicillins |
| 8248 | BENETHAMINE PENICILLIN G /PEN.G SODIUM/ 475 MG INJ | penicillins |
| 77222 | Benethamine penicillin with procaine benzylpenicillin and benzylpenicillin sodium injection | penicillins |
| 78984 | Benzathine benzylpenicillin 2.4million unit powder and solvent for suspension for injection vials | penicillins |
| 29377 | Benzathine peniciilin 229mg/5ml suspension | penicillins |
| 17155 | Benzathine penicillin 115mg/ml drops | penicillins |
| 23000 | Benzathine penicillin 229mg/ml injection | penicillins |
| 21042 | Benzylpeniciilin 250mg tablets | penicillins |
| 25228 | Benzylpeniciilin 250mg/5ml oral solution | penicillins |
| 16100 | Benzylpenicillin 1.2g powder for solution for injection vials | penicillins |
| 64308 | Benzylpenicillin 1.2g powder for solution for injection vials (A A H Pharmaceuticals Ltd) | penicillins |
| 64596 | Benzylpenicillin 1.2g powder for solution for injection vials (Alliance Healthcare (Distribution) Ltd) | penicillins |
| 62945 | Benzylpenicillin 1.2g powder for solution for injection vials (Genus Pharmaceuticals Ltd) | penicillins |
| 16683 | Benzylpenicillin 125mg/5ml oral solution | penicillins |
| 55829 | Benzylpenicillin 12mg/vial intrathecal injection | penicillins |
| 12505 | Benzylpenicillin 300mg/vial injection | penicillins |
| 23776 | Benzylpenicillin 3g/vial injection | penicillins |
| 2545 | Benzylpenicillin 600mg powder for solution for injection vials | penicillins |
| 64049 | Benzylpenicillin 600mg powder for solution for injection vials (A A H Pharmaceuticals Ltd) | penicillins |
| 78989 | Benzylpenicillin 600mg powder for solution for injection vials (Alliance Healthcare (Distribution) Ltd) | penicillins |
| 62685 | Benzylpenicillin 600mg powder for solution for injection vials (Genus Pharmaceuticals Ltd) | penicillins |
| 76494 | Benzylpenicillin sodium with benemethamine penicillin with procaine benzylpenicillin injection | penicillins |
| 30099 | Benzylpenicillin sodium with procaine benzylpenicillin injection | penicillins |
| 29805 | Benzylpenicllin 6g/vial injection | penicillins |
| 19895 | Bicillin Injection (Yamanouchi Pharma Ltd) | penicillins |
| 22452 | Britcin 250mg Capsule (DDSA Pharmaceuticals Ltd) | penicillins |
| 21443 | Broxil 125mg/5ml Oral solution (Beecham Research Laboratories) | penicillins |
| 7427 | BROXIL 250 MG TAB | penicillins |
| 19286 | Broxil 250mg Capsule (Beecham Research Laboratories) | penicillins |
| 31101 | CALTHOR 125 MG SYR | penicillins |
| 18648 | Calthor 250mg Tablet (Wyeth Pharmaceuticals) | penicillins |
| 29545 | Calthor 500mg Tablet (Wyeth Pharmaceuticals) | penicillins |
| 289 | CARBENICILLIN 1 INJ | penicillins |
| 66222 | Carbenicillin 1g sterile powder | penicillins |
| 47648 | Carfecillin 500mg tablets | penicillins |
| 12553 | CARFECILLIN SODIUM 500 MG TAB | penicillins |
| 42100 | Ciclacillin 250mg tablets | penicillins |
| 11433 | Clarithromycin 500mg with lansoprazole 30mg and amoxicillin 500mg triple pack | penicillins |
| 17121 | Clavulanic acid 100mg with amoxicillin 500mg/vial injection | penicillins |
| 17095 | Clavulanic acid 100mg with Ticarcillin 1.5g infusion | penicillins |
| 9925 | Clavulanic acid 125mg with Amoxicillin 250mg tablets | penicillins |
| 13239 | Clavulanic acid 125mg with Amoxicillin 500mg tablets | penicillins |
| 24006 | Clavulanic acid 31mg with amoxcillin 125mg/5ml oral suspension | penicillins |
| 21775 | Clavulanic acid 31mg with amoxicillin 125mg/5ml sugar free oral suspension | penicillins |
| 20432 | Clavulanic acid 57mg with amoxicillin 400mg/5ml sugar free suspension | penicillins |
| 42485 | Clavulanic acid 62mg with amoxicillin 250mg/5ml oral suspension | penicillins |
| 16612 | Clavulanic acid 62mg with amoxicillin 250mg/5ml sugar free suspension | penicillins |
| 24093 | Clavulanic acid with amoxicillin dispersible tablets | penicillins |
| 8282 | Cloxacillin 125mg/5ml oral solution | penicillins |
| 7703 | Cloxacillin 250mg capsules | penicillins |
| 54907 | Cloxacillin 250mg with Ampicillin 250mg injection | penicillins |
| 52340 | Cloxacillin 30mg with Ampicillin 60mg/0.6ml suspension | penicillins |
| 10461 | Cloxacillin 500mg capsules | penicillins |
| 28618 | Cloxacillin 500mg/vial Im injection | penicillins |
| 30461 | CLOXACILLIN TAB | penicillins |
| 24005 | Co-amoxiclav 1000mg/200mg powder for solution for injection vials | penicillins |
| 75243 | Co-amoxiclav 1000mg/200mg powder for solution for injection vials (Peckforton Pharmaceuticals Ltd) | penicillins |
| 67771 | Co-amoxiclav 1000mg/200mg powder for solution for injection vials (PLIVA Pharma Ltd) | penicillins |
| 79672 | Co-amoxiclav 1000mg/200mg powder for solution for injection vials (Teva UK Ltd) | penicillins |
| 66905 | Co-amoxiclav 1000mg/200mg powder for solution for injection vials (Wockhardt UK Ltd) | penicillins |
| 10200 | Co-amoxiclav 125mg/31mg/5ml oral suspension | penicillins |
| 54052 | Co-amoxiclav 125mg/31mg/5ml oral suspension (A A H Pharmaceuticals Ltd) | penicillins |
| 60281 | Co-amoxiclav 125mg/31mg/5ml oral suspension (CST Pharma Ltd) | penicillins |
| 75782 | Co-amoxiclav 125mg/31mg/5ml oral suspension (DE Pharmaceuticals) | penicillins |
| 69920 | Co-amoxiclav 125mg/31mg/5ml oral suspension (Ennogen Healthcare Ltd) | penicillins |
| 61299 | Co-amoxiclav 125mg/31mg/5ml oral suspension (Mawdsley-Brooks & Company Ltd) | penicillins |
| 54732 | Co-amoxiclav 125mg/31mg/5ml oral suspension (Mylan) | penicillins |
| 62686 | Co-amoxiclav 125mg/31mg/5ml oral suspension (Pharma-z Ltd) | penicillins |
| 71541 | Co-amoxiclav 125mg/31mg/5ml oral suspension (Sigma Pharmaceuticals Plc) | penicillins |
| 59588 | Co-amoxiclav 125mg/31mg/5ml oral suspension (Waymade Healthcare Plc) | penicillins |
| 1638 | Co-amoxiclav 125mg/31mg/5ml oral suspension sugar free | penicillins |
| 43548 | Co-amoxiclav 125mg/31mg/5ml oral suspension sugar free (A A H Pharmaceuticals Ltd) | penicillins |
| 54324 | Co-amoxiclav 125mg/31mg/5ml oral suspension sugar free (Actavis UK Ltd) | penicillins |
| 54452 | Co-amoxiclav 125mg/31mg/5ml oral suspension sugar free (Alliance Healthcare (Distribution) Ltd) | penicillins |
| 54808 | Co-amoxiclav 125mg/31mg/5ml oral suspension sugar free (Almus Pharmaceuticals Ltd) | penicillins |
| 61407 | Co-amoxiclav 125mg/31mg/5ml oral suspension sugar free (Colorama Pharmaceuticals Ltd) | penicillins |
| 28874 | Co-amoxiclav 125mg/31mg/5ml oral suspension sugar free (IVAX Pharmaceuticals UK Ltd) | penicillins |
| 58097 | Co-amoxiclav 125mg/31mg/5ml oral suspension sugar free (Kent Pharmaceuticals Ltd) | penicillins |
| 56884 | Co-amoxiclav 125mg/31mg/5ml oral suspension sugar free (Phoenix Healthcare Distribution Ltd) | penicillins |
| 34680 | Co-amoxiclav 125mg/31mg/5ml oral suspension sugar free (Ranbaxy (UK) Ltd) | penicillins |
| 34972 | Co-amoxiclav 125mg/31mg/5ml oral suspension sugar free (Sandoz Ltd) | penicillins |
| 76188 | Co-amoxiclav 125mg/31mg/5ml oral suspension sugar free (Teva UK Ltd) | penicillins |
| 66650 | Co-amoxiclav 125mg/31mg/5ml oral suspension sugar free (Waymade Healthcare Plc) | penicillins |
| 829 | Co-amoxiclav 250mg/125mg dispersible tablets sugar free | penicillins |
| 545 | Co-amoxiclav 250mg/125mg tablets | penicillins |
| 30786 | Co-amoxiclav 250mg/125mg tablets (A A H Pharmaceuticals Ltd) | penicillins |
| 19209 | Co-amoxiclav 250mg/125mg tablets (Actavis UK Ltd) | penicillins |
| 51623 | Co-amoxiclav 250mg/125mg tablets (Alliance Healthcare (Distribution) Ltd) | penicillins |
| 48147 | Co-amoxiclav 250mg/125mg tablets (Almus Pharmaceuticals Ltd) | penicillins |
| 58803 | Co-amoxiclav 250mg/125mg tablets (APC Pharmaceuticals & Chemicals (Europe) Ltd) | penicillins |
| 75663 | Co-amoxiclav 250mg/125mg tablets (Bristol Laboratories Ltd) | penicillins |
| 66747 | Co-amoxiclav 250mg/125mg tablets (Brown & Burk UK Ltd) | penicillins |
| 60034 | Co-amoxiclav 250mg/125mg tablets (DE Pharmaceuticals) | penicillins |
| 28871 | Co-amoxiclav 250mg/125mg tablets (IVAX Pharmaceuticals UK Ltd) | penicillins |
| 33693 | Co-amoxiclav 250mg/125mg tablets (Kent Pharmaceuticals Ltd) | penicillins |
| 67694 | Co-amoxiclav 250mg/125mg tablets (Mawdsley-Brooks & Company Ltd) | penicillins |
| 34297 | Co-amoxiclav 250mg/125mg tablets (Mylan) | penicillins |
| 50446 | Co-amoxiclav 250mg/125mg tablets (Phoenix Healthcare Distribution Ltd) | penicillins |
| 30783 | Co-amoxiclav 250mg/125mg tablets (Ranbaxy (UK) Ltd) | penicillins |
| 75269 | Co-amoxiclav 250mg/125mg tablets (Rivopharm (UK) Ltd) | penicillins |
| 19414 | Co-amoxiclav 250mg/125mg tablets (Sandoz Ltd) | penicillins |
| 65215 | Co-amoxiclav 250mg/125mg tablets (Sigma Pharmaceuticals Plc) | penicillins |
| 34734 | Co-amoxiclav 250mg/125mg tablets (Teva UK Ltd) | penicillins |
| 55312 | Co-amoxiclav 250mg/125mg tablets (Waymade Healthcare Plc) | penicillins |
| 46915 | Co-amoxiclav 250mg/125mg tablets (Zentiva) | penicillins |
| 7364 | Co-amoxiclav 250mg/62mg/5ml oral suspension | penicillins |
| 54708 | Co-amoxiclav 250mg/62mg/5ml oral suspension (A A H Pharmaceuticals Ltd) | penicillins |
| 65533 | Co-amoxiclav 250mg/62mg/5ml oral suspension (CST Pharma Ltd) | penicillins |
| 63063 | Co-amoxiclav 250mg/62mg/5ml oral suspension (DE Pharmaceuticals) | penicillins |
| 75547 | Co-amoxiclav 250mg/62mg/5ml oral suspension (Ennogen Healthcare Ltd) | penicillins |
| 54780 | Co-amoxiclav 250mg/62mg/5ml oral suspension (Mylan) | penicillins |
| 79302 | Co-amoxiclav 250mg/62mg/5ml oral suspension (Sigma Pharmaceuticals Plc) | penicillins |
| 524 | Co-amoxiclav 250mg/62mg/5ml oral suspension sugar free | penicillins |
| 42227 | Co-amoxiclav 250mg/62mg/5ml oral suspension sugar free (A A H Pharmaceuticals Ltd) | penicillins |
| 71819 | Co-amoxiclav 250mg/62mg/5ml oral suspension sugar free (Alliance Healthcare (Distribution) Ltd) | penicillins |
| 51678 | Co-amoxiclav 250mg/62mg/5ml oral suspension sugar free (Almus Pharmaceuticals Ltd) | penicillins |
| 58494 | Co-amoxiclav 250mg/62mg/5ml oral suspension sugar free (Colorama Pharmaceuticals Ltd) | penicillins |
| 37304 | Co-amoxiclav 250mg/62mg/5ml oral suspension sugar free (IVAX Pharmaceuticals UK Ltd) | penicillins |
| 60134 | Co-amoxiclav 250mg/62mg/5ml oral suspension sugar free (Kent Pharmaceuticals Ltd) | penicillins |
| 69140 | Co-amoxiclav 250mg/62mg/5ml oral suspension sugar free (Mawdsley-Brooks & Company Ltd) | penicillins |
| 59740 | Co-amoxiclav 250mg/62mg/5ml oral suspension sugar free (Phoenix Healthcare Distribution Ltd) | penicillins |
| 40320 | Co-amoxiclav 250mg/62mg/5ml oral suspension sugar free (Ranbaxy (UK) Ltd) | penicillins |
| 46918 | Co-amoxiclav 250mg/62mg/5ml oral suspension sugar free (Sandoz Ltd) | penicillins |
| 34234 | Co-amoxiclav 250mg/62mg/5ml oral suspension sugar free (Teva UK Ltd) | penicillins |
| 56578 | Co-amoxiclav 250mg/62mg/5ml oral suspension sugar free (Waymade Healthcare Plc) | penicillins |
| 6687 | Co-amoxiclav 400mg/57mg/5ml oral suspension sugar free | penicillins |
| 51637 | Co-amoxiclav 400mg/57mg/5ml oral suspension sugar free (A A H Pharmaceuticals Ltd) | penicillins |
| 78021 | Co-amoxiclav 400mg/57mg/5ml oral suspension sugar free (Alliance Healthcare (Distribution) Ltd) | penicillins |
| 68408 | Co-amoxiclav 400mg/57mg/5ml oral suspension sugar free (Brown & Burk UK Ltd) | penicillins |
| 65056 | Co-amoxiclav 400mg/57mg/5ml oral suspension sugar free (Sandoz Ltd) | penicillins |
| 577 | Co-amoxiclav 500mg/100mg powder for solution for injection vials | penicillins |
| 64986 | Co-amoxiclav 500mg/100mg powder for solution for injection vials (A A H Pharmaceuticals Ltd) | penicillins |
| 73039 | Co-amoxiclav 500mg/100mg powder for solution for injection vials (Bowmed Ibisqus Ltd) | penicillins |
| 35191 | Co-amoxiclav 500mg/100mg powder for solution for injection vials (Teva UK Ltd) | penicillins |
| 47184 | Co-amoxiclav 500mg/100mg powder for solution for injection vials (Wockhardt UK Ltd) | penicillins |
| 641 | Co-amoxiclav 500mg/125mg tablets | penicillins |
| 33701 | Co-amoxiclav 500mg/125mg tablets (A A H Pharmaceuticals Ltd) | penicillins |
| 50742 | Co-amoxiclav 500mg/125mg tablets (Actavis UK Ltd) | penicillins |
| 50341 | Co-amoxiclav 500mg/125mg tablets (Alliance Healthcare (Distribution) Ltd) | penicillins |
| 71937 | Co-amoxiclav 500mg/125mg tablets (Almus Pharmaceuticals Ltd) | penicillins |
| 53609 | Co-amoxiclav 500mg/125mg tablets (APC Pharmaceuticals & Chemicals (Europe) Ltd) | penicillins |
| 53996 | Co-amoxiclav 500mg/125mg tablets (Aurobindo Pharma Ltd) | penicillins |
| 75331 | Co-amoxiclav 500mg/125mg tablets (Bristol Laboratories Ltd) | penicillins |
| 67466 | Co-amoxiclav 500mg/125mg tablets (Brown & Burk UK Ltd) | penicillins |
| 75317 | Co-amoxiclav 500mg/125mg tablets (Consilient Health Ltd) | penicillins |
| 62377 | Co-amoxiclav 500mg/125mg tablets (Creo Pharma Ltd) | penicillins |
| 59908 | Co-amoxiclav 500mg/125mg tablets (DE Pharmaceuticals) | penicillins |
| 29356 | Co-amoxiclav 500mg/125mg tablets (IVAX Pharmaceuticals UK Ltd) | penicillins |
| 40148 | Co-amoxiclav 500mg/125mg tablets (Kent Pharmaceuticals Ltd) | penicillins |
| 49610 | Co-amoxiclav 500mg/125mg tablets (Medreich Plc) | penicillins |
| 30705 | Co-amoxiclav 500mg/125mg tablets (Mylan) | penicillins |
| 54591 | Co-amoxiclav 500mg/125mg tablets (Phoenix Healthcare Distribution Ltd) | penicillins |
| 34493 | Co-amoxiclav 500mg/125mg tablets (Ranbaxy (UK) Ltd) | penicillins |
| 32910 | Co-amoxiclav 500mg/125mg tablets (Sandoz Ltd) | penicillins |
| 69292 | Co-amoxiclav 500mg/125mg tablets (Sigma Pharmaceuticals Plc) | penicillins |
| 29353 | Co-amoxiclav 500mg/125mg tablets (Teva UK Ltd) | penicillins |
| 57081 | Co-amoxiclav 500mg/125mg tablets (Waymade Healthcare Plc) | penicillins |
| 44154 | Co-amoxiclav 500mg/125mg tablets (Zentiva) | penicillins |
| 62332 | Co-amoxiclav 875mg/125mg tablets | penicillins |
| 63452 | Co-amoxiclav 875mg/125mg tablets (Creo Pharma Ltd) | penicillins |
| 76760 | Co-amoxiclav 875mg/125mg tablets (Mawdsley-Brooks & Company Ltd) | penicillins |
| 29995 | CO-CAPS PENICILLIN V-K 250 MG CAP | penicillins |
| 23719 | CO-FLUAMPICIL | penicillins |
| 9473 | Co-fluampicil 125mg/125mg/5ml oral suspension | penicillins |
| 75978 | Co-fluampicil 125mg/125mg/5ml oral suspension (A A H Pharmaceuticals Ltd) | penicillins |
| 71420 | Co-fluampicil 125mg/125mg/5ml oral suspension (Alliance Healthcare (Distribution) Ltd) | penicillins |
| 5454 | Co-fluampicil 250mg/250mg capsules | penicillins |
| 19648 | Co-fluampicil 250mg/250mg capsules (A A H Pharmaceuticals Ltd) | penicillins |
| 34380 | Co-fluampicil 250mg/250mg capsules (Actavis UK Ltd) | penicillins |
| 71086 | Co-fluampicil 250mg/250mg capsules (Almus Pharmaceuticals Ltd) | penicillins |
| 30764 | Co-fluampicil 250mg/250mg capsules (IVAX Pharmaceuticals UK Ltd) | penicillins |
| 41415 | Co-fluampicil 250mg/250mg capsules (Kent Pharmaceuticals Ltd) | penicillins |
| 34358 | Co-fluampicil 250mg/250mg capsules (Mylan) | penicillins |
| 25570 | Co-fluampicil 250mg/250mg capsules (Sandoz Ltd) | penicillins |
| 73125 | Co-fluampicil 250mg/250mg capsules (Waymade Healthcare Plc) | penicillins |
| 26329 | Co-fluampicil 250mg/250mg powder for solution for injection vials | penicillins |
| 45237 | Co-fluampicil 500mg with 500mg injection | penicillins |
| 24119 | CRYSTAPEN (SOD SALT) 3 GM INJ | penicillins |
| 12034 | CRYSTAPEN (SOD SALT) 300 GM INJ | penicillins |
| 12218 | CRYSTAPEN (SOD SALT) 6 GM INJ | penicillins |
| 46217 | Crystapen 1.2g powder for solution for injection vials (Thornton & Ross Ltd) | penicillins |
| 25532 | Crystapen 1200mg Powder for solution for injection (Britannia Pharmaceuticals Ltd) | penicillins |
| 32469 | CRYSTAPEN 1200MG/VIAL | penicillins |
| 43624 | Crystapen 600mg powder for solution for injection vials (Thornton & Ross Ltd) | penicillins |
| 2232 | Crystapen 600mg/vial Powder for solution for injection (Britannia Pharmaceuticals Ltd) | penicillins |
| 54455 | Crystapen 6g/vial Injection (Glaxo Laboratories Ltd) | penicillins |
| 32809 | CRYSTAPEN G 125 MG SYR | penicillins |
| 22179 | Crystapen g 250mg Tablet (Glaxo Laboratories Ltd) | penicillins |
| 31100 | CRYSTAPEN V 125 MG SUS | penicillins |
| 32819 | CRYSTAPEN V 125 MG SYR | penicillins |
| 26206 | CRYSTAPEN V 125 MG TAB | penicillins |
| 38110 | Crystapen v 125mg/5ml Oral solution (Glaxo Laboratories Ltd) | penicillins |
| 25628 | CRYSTAPEN V 250 MG TAB | penicillins |
| 18310 | DISTAQUAINE V-K (DQV-K) 125 MG TAB | penicillins |
| 32185 | Distaquaine v-k 125mg Tablet (Dista Products Ltd) | penicillins |
| 15014 | Distaquaine v-k 125mg/5ml Liquid (Dista Products Ltd) | penicillins |
| 18309 | Distaquaine v-k 250mg Tablet (Dista Products Ltd) | penicillins |
| 26659 | Distaquaine v-k 250mg/5ml Liquid (Dista Products Ltd) | penicillins |
| 18930 | Flemoxin 375mg Soluble tablet (Paines & Byrne Ltd) | penicillins |
| 24396 | Flemoxin 750mg Soluble tablet (Paines & Byrne Ltd) | penicillins |
| 8933 | Floxapen 125mg/5ml syrup (Actavis UK Ltd) | penicillins |
| 26558 | Floxapen 1g powder for solution for injection vials (GlaxoSmithKline UK Ltd) | penicillins |
| 3708 | Floxapen 250mg Capsule (Beecham Research Laboratories) | penicillins |
| 39989 | Floxapen 250mg capsules (Actavis UK Ltd) | penicillins |
| 26326 | Floxapen 250mg powder for solution for injection vials (GlaxoSmithKline UK Ltd) | penicillins |
| 2642 | Floxapen 250mg/5ml syrup (Actavis UK Ltd) | penicillins |
| 8468 | Floxapen 500mg Capsule (Beecham Research Laboratories) | penicillins |
| 40025 | Floxapen 500mg capsules (Actavis UK Ltd) | penicillins |
| 26350 | Floxapen 500mg powder for solution for injection vials (GlaxoSmithKline UK Ltd) | penicillins |
| 41983 | Floxapen iv 500mg/vial Intravenous injection (Beecham Research Laboratories) | penicillins |
| 23485 | Flu-amp 500mg Capsule (Generics (UK) Ltd) | penicillins |
| 2299 | Fluclomix 250 capsules (Ashbourne Pharmaceuticals Ltd) | penicillins |
| 4008 | Fluclomix 500 capsules (Ashbourne Pharmaceuticals Ltd) | penicillins |
| 23583 | FLUCLOXACILLIN | penicillins |
| 161 | FLUCLOXACILLIN 1 GM CAP | penicillins |
| 3877 | FLUCLOXACILLIN 125 MG CAP | penicillins |
| 1694 | FLUCLOXACILLIN 125 MG POW | penicillins |
| 45312 | Flucloxacillin 125mg/5ml Liquid (Lagap) | penicillins |
| 457 | Flucloxacillin 125mg/5ml oral solution | penicillins |
| 27057 | Flucloxacillin 125mg/5ml oral solution (A A H Pharmaceuticals Ltd) | penicillins |
| 43546 | Flucloxacillin 125mg/5ml oral solution (Actavis UK Ltd) | penicillins |
| 51262 | Flucloxacillin 125mg/5ml oral solution (Alliance Healthcare (Distribution) Ltd) | penicillins |
| 56869 | Flucloxacillin 125mg/5ml oral solution (Crescent Pharma Ltd) | penicillins |
| 65945 | Flucloxacillin 125mg/5ml oral solution (DE Pharmaceuticals) | penicillins |
| 34701 | Flucloxacillin 125mg/5ml oral solution (IVAX Pharmaceuticals UK Ltd) | penicillins |
| 34766 | Flucloxacillin 125mg/5ml oral solution (Kent Pharmaceuticals Ltd) | penicillins |
| 60593 | Flucloxacillin 125mg/5ml oral solution (Medreich Plc) | penicillins |
| 60032 | Flucloxacillin 125mg/5ml oral solution (Milpharm Ltd) | penicillins |
| 34683 | Flucloxacillin 125mg/5ml oral solution (Mylan) | penicillins |
| 57415 | Flucloxacillin 125mg/5ml oral solution (Phoenix Healthcare Distribution Ltd) | penicillins |
| 34259 | Flucloxacillin 125mg/5ml oral solution (Sandoz Ltd) | penicillins |
| 28284 | Flucloxacillin 125mg/5ml oral solution (Teva UK Ltd) | penicillins |
| 55754 | Flucloxacillin 125mg/5ml oral solution (Waymade Healthcare Plc) | penicillins |
| 41163 | Flucloxacillin 125mg/5ml oral solution sugar free | penicillins |
| 72219 | Flucloxacillin 125mg/5ml oral solution sugar free (A A H Pharmaceuticals Ltd) | penicillins |
| 47125 | Flucloxacillin 125mg/5ml oral solution sugar free (Accord Healthcare Ltd) | penicillins |
| 79172 | Flucloxacillin 125mg/5ml oral solution sugar free (Almus Pharmaceuticals Ltd) | penicillins |
| 49349 | Flucloxacillin 125mg/5ml oral solution sugar free (Kent Pharmaceuticals Ltd) | penicillins |
| 61896 | Flucloxacillin 125mg/5ml oral solution sugar free (Waymade Healthcare Plc) | penicillins |
| 1000 | Flucloxacillin 125mg/5ml oral suspension | penicillins |
| 16191 | Flucloxacillin 1g powder for solution for injection vials | penicillins |
| 60390 | Flucloxacillin 1g powder for solution for injection vials (A A H Pharmaceuticals Ltd) | penicillins |
| 60330 | Flucloxacillin 1g powder for solution for injection vials (Wockhardt UK Ltd) | penicillins |
| 34870 | Flucloxacillin 250mg Capsule (C P Pharmaceuticals Ltd) | penicillins |
| 34944 | Flucloxacillin 250mg Capsule (Co-Pharma Ltd) | penicillins |
| 60 | Flucloxacillin 250mg capsules | penicillins |
| 23240 | Flucloxacillin 250mg capsules (A A H Pharmaceuticals Ltd) | penicillins |
| 34057 | Flucloxacillin 250mg capsules (Actavis UK Ltd) | penicillins |
| 51037 | Flucloxacillin 250mg capsules (Alliance Healthcare (Distribution) Ltd) | penicillins |
| 40108 | Flucloxacillin 250mg capsules (Almus Pharmaceuticals Ltd) | penicillins |
| 58541 | Flucloxacillin 250mg capsules (Bristol Laboratories Ltd) | penicillins |
| 68696 | Flucloxacillin 250mg capsules (DE Pharmaceuticals) | penicillins |
| 34721 | Flucloxacillin 250mg capsules (IVAX Pharmaceuticals UK Ltd) | penicillins |
| 34375 | Flucloxacillin 250mg capsules (Kent Pharmaceuticals Ltd) | penicillins |
| 68001 | Flucloxacillin 250mg capsules (Mawdsley-Brooks & Company Ltd) | penicillins |
| 57106 | Flucloxacillin 250mg capsules (Medreich Plc) | penicillins |
| 57634 | Flucloxacillin 250mg capsules (Milpharm Ltd) | penicillins |
| 34330 | Flucloxacillin 250mg capsules (Mylan) | penicillins |
| 29944 | Flucloxacillin 250mg capsules (Ranbaxy (UK) Ltd) | penicillins |
| 34364 | Flucloxacillin 250mg capsules (Sandoz Ltd) | penicillins |
| 56027 | Flucloxacillin 250mg capsules (Sigma Pharmaceuticals Plc) | penicillins |
| 33993 | Flucloxacillin 250mg capsules (Teva UK Ltd) | penicillins |
| 72454 | Flucloxacillin 250mg capsules (Waymade Healthcare Plc) | penicillins |
| 15247 | Flucloxacillin 250mg powder for solution for injection vials | penicillins |
| 57331 | Flucloxacillin 250mg powder for solution for injection vials (A A H Pharmaceuticals Ltd) | penicillins |
| 53851 | Flucloxacillin 250mg powder for solution for injection vials (Wockhardt UK Ltd) | penicillins |
| 1001 | Flucloxacillin 250mg/5ml oral solution | penicillins |
| 31775 | Flucloxacillin 250mg/5ml oral solution (A A H Pharmaceuticals Ltd) | penicillins |
| 64375 | Flucloxacillin 250mg/5ml oral solution (Actavis UK Ltd) | penicillins |
| 73965 | Flucloxacillin 250mg/5ml oral solution (Alliance Healthcare (Distribution) Ltd) | penicillins |
| 57462 | Flucloxacillin 250mg/5ml oral solution (Crescent Pharma Ltd) | penicillins |
| 50729 | Flucloxacillin 250mg/5ml oral solution (DE Pharmaceuticals) | penicillins |
| 32901 | Flucloxacillin 250mg/5ml oral solution (Kent Pharmaceuticals Ltd) | penicillins |
| 31940 | Flucloxacillin 250mg/5ml Oral solution (Lagap) | penicillins |
| 52549 | Flucloxacillin 250mg/5ml oral solution (Phoenix Healthcare Distribution Ltd) | penicillins |
| 32065 | Flucloxacillin 250mg/5ml Oral solution (Teva UK Ltd) | penicillins |
| 73462 | Flucloxacillin 250mg/5ml oral solution (Waymade Healthcare Plc) | penicillins |
| 41172 | Flucloxacillin 250mg/5ml oral solution sugar free | penicillins |
| 56189 | Flucloxacillin 250mg/5ml oral solution sugar free (A A H Pharmaceuticals Ltd) | penicillins |
| 51830 | Flucloxacillin 250mg/5ml oral solution sugar free (Accord Healthcare Ltd) | penicillins |
| 69709 | Flucloxacillin 250mg/5ml oral solution sugar free (Alliance Healthcare (Distribution) Ltd) | penicillins |
| 79072 | Flucloxacillin 250mg/5ml oral solution sugar free (DE Pharmaceuticals) | penicillins |
| 55438 | Flucloxacillin 250mg/5ml oral solution sugar free (Kent Pharmaceuticals Ltd) | penicillins |
| 57292 | Flucloxacillin 250mg/5ml oral solution sugar free (Waymade Healthcare Plc) | penicillins |
| 5423 | Flucloxacillin 250mg/5ml oral suspension | penicillins |
| 75944 | Flucloxacillin 2g powder for solution for injection vials | penicillins |
| 34848 | Flucloxacillin 500mg Capsule (C P Pharmaceuticals Ltd) | penicillins |
| 45252 | Flucloxacillin 500mg Capsule (Co-Pharma Ltd) | penicillins |
| 579 | Flucloxacillin 500mg capsules | penicillins |
| 34313 | Flucloxacillin 500mg capsules (A A H Pharmaceuticals Ltd) | penicillins |
| 34464 | Flucloxacillin 500mg capsules (Actavis UK Ltd) | penicillins |
| 57485 | Flucloxacillin 500mg capsules (Alliance Healthcare (Distribution) Ltd) | penicillins |
| 52374 | Flucloxacillin 500mg capsules (Almus Pharmaceuticals Ltd) | penicillins |
| 56384 | Flucloxacillin 500mg capsules (Bristol Laboratories Ltd) | penicillins |
| 60260 | Flucloxacillin 500mg capsules (DE Pharmaceuticals) | penicillins |
| 34052 | Flucloxacillin 500mg capsules (IVAX Pharmaceuticals UK Ltd) | penicillins |
| 34226 | Flucloxacillin 500mg capsules (Kent Pharmaceuticals Ltd) | penicillins |
| 67710 | Flucloxacillin 500mg capsules (Mawdsley-Brooks & Company Ltd) | penicillins |
| 50820 | Flucloxacillin 500mg capsules (Medreich Plc) | penicillins |
| 52281 | Flucloxacillin 500mg capsules (Milpharm Ltd) | penicillins |
| 34776 | Flucloxacillin 500mg capsules (Mylan) | penicillins |
| 77771 | Flucloxacillin 500mg capsules (Noumed Life Sciences Ltd) | penicillins |
| 78246 | Flucloxacillin 500mg capsules (Phoenix Healthcare Distribution Ltd) | penicillins |
| 44855 | Flucloxacillin 500mg capsules (Ranbaxy (UK) Ltd) | penicillins |
| 34617 | Flucloxacillin 500mg capsules (Sandoz Ltd) | penicillins |
| 55799 | Flucloxacillin 500mg capsules (Sigma Pharmaceuticals Plc) | penicillins |
| 34007 | Flucloxacillin 500mg capsules (Teva UK Ltd) | penicillins |
| 63306 | Flucloxacillin 500mg capsules (Waymade Healthcare Plc) | penicillins |
| 10814 | Flucloxacillin 500mg powder for solution for injection vials | penicillins |
| 56717 | Flucloxacillin 500mg powder for solution for injection vials (A A H Pharmaceuticals Ltd) | penicillins |
| 59422 | Flucloxacillin 500mg powder for solution for injection vials (Wockhardt UK Ltd) | penicillins |
| 40142 | Flucloxacillin iv 250mg/vial Injection | penicillins |
| 29671 | Flucloxacillin iv 500mg/vial Injection | penicillins |
| 58126 | Flucloxacillin sodium 125mg/5ml oral suspension | penicillins |
| 951 | Flucloxacillin with ampicillin 125mg+125mg Liquid | penicillins |
| 9242 | Flucloxacillin with ampicillin 250mg+250mg Capsule | penicillins |
| 20869 | Flucloxacillin with ampicillin 250mg+250mg Injection | penicillins |
| 2735 | Flucloxin 125mg/5ml Liquid (OPD Pharm) | penicillins |
| 9800 | Flucloxin 250mg Capsule (OPD Pharm) | penicillins |
| 4074 | Flucloxin 500mg Capsule (OPD Pharm) | penicillins |
| 14386 | Galenamox 125mg/5ml oral suspension (Galen Ltd) | penicillins |
| 14371 | Galenamox 250mg capsules (Galen Ltd) | penicillins |
| 14407 | Galenamox 250mg/5ml oral suspension (Galen Ltd) | penicillins |
| 14396 | Galenamox 500mg capsules (Galen Ltd) | penicillins |
| 22422 | Galfloxin 250mg Capsule (Galen Ltd) | penicillins |
| 32665 | Galfloxin 500mg Capsule (Galen Ltd) | penicillins |
| 24747 | Ladropen 125mg/5ml Liquid (Berk Pharmaceuticals Ltd) | penicillins |
| 21806 | Ladropen 250mg capsules (Teva UK Ltd) | penicillins |
| 21858 | Ladropen 500mg capsules (Teva UK Ltd) | penicillins |
| 15290 | Lansoprazole with amoxicillin and clarithromycin 30mg + 500mg + 500mg Triple pack | penicillins |
| 22194 | MAGNAPEN | penicillins |
| 13438 | Magnapen 1g/vial Injection (C P Pharmaceuticals Ltd) | penicillins |
| 308 | Magnapen 250mg/250mg capsules (Wockhardt UK Ltd) | penicillins |
| 13531 | Magnapen 500mg powder for solution for injection vials (Wockhardt UK Ltd) | penicillins |
| 2874 | Magnapen syrup (Wockhardt UK Ltd) | penicillins |
| 30696 | Mecillinam 400mg/vial Injection | penicillins |
| 28896 | Mezlocillin 1g/vial Injection | penicillins |
| 28572 | Mezlocillin 50mg/vial Injection | penicillins |
| 21029 | Miraxid 450 Tablet (Rpr / Fisons) | penicillins |
| 20516 | Miraxid Liquid (Rpr / Fisons) | penicillins |
| 17161 | Miraxid Tablet (Rpr / Fisons) | penicillins |
| 36084 | Negaban 1g powder for solution for injection vials (Eumedica Pharmaceuticals) | penicillins |
| 25834 | Orbenin 125mg/5ml Oral solution (Goldshield Pharmaceuticals Ltd) | penicillins |
| 8634 | Orbenin 250mg Capsule (Goldshield Pharmaceuticals Ltd) | penicillins |
| 8633 | Orbenin 500mg Capsule (Goldshield Pharmaceuticals Ltd) | penicillins |
| 18934 | Penbritin 125mg/1.25ml Liquid (Beecham Research Laboratories) | penicillins |
| 10603 | Penbritin 125mg/5ml Oral solution (Beecham Research Laboratories) | penicillins |
| 37485 | Penbritin 125mg/5ml syrup (Chemidex Pharma Ltd) | penicillins |
| 204 | Penbritin 250mg Capsule (Beecham Research Laboratories) | penicillins |
| 24483 | Penbritin 250mg capsules (Chemidex Pharma Ltd) | penicillins |
| 4318 | Penbritin 250mg/5ml Oral solution (Beecham Research Laboratories) | penicillins |
| 30630 | Penbritin 250mg/vial Injection (Beecham Research Laboratories) | penicillins |
| 15039 | Penbritin 500mg Capsule (Beecham Research Laboratories) | penicillins |
| 31281 | Penbritin 500mg capsules (Chemidex Pharma Ltd) | penicillins |
| 7531 | Penbritin 500mg powder for solution for injection vials (Chemidex Pharma Ltd) | penicillins |
| 38091 | Penbritin Forte 250mg/5ml syrup (Chemidex Pharma Ltd) | penicillins |
| 1067 | PENICILLIN 500 MG TAB | penicillins |
| 2755 | PENICILLIN G 125 MG SYR | penicillins |
| 53682 | Penicillin G 12mg/vial intrathecal injection | penicillins |
| 2730 | PENICILLIN G 250 MG SYR | penicillins |
| 2289 | PENICILLIN G 250 MG TAB | penicillins |
| 38084 | Penicillin G 300mg/vial injection | penicillins |
| 42995 | Penicillin G 3g/vial injection | penicillins |
| 391 | Penicillin G 600mg/vial injection | penicillins |
| 44282 | Penicillin G 6g/vial injection | penicillins |
| 24669 | PENICILLIN V | penicillins |
| 1783 | Penicillin V 125mg tablet | penicillins |
| 127 | Penicillin V 125mg/5ml oral solution | penicillins |
| 46145 | Penicillin V 125mg/5ml Oral solution (C P Pharmaceuticals Ltd) | penicillins |
| 45527 | Penicillin V 125mg/5ml Oral solution (Celltech Pharma Europe Ltd) | penicillins |
| 34363 | Penicillin V 125mg/5ml Oral solution (Generics (UK) Ltd) | penicillins |
| 45022 | Penicillin V 125mg/5ml Oral solution (Sovereign Medical Ltd) | penicillins |
| 10095 | Penicillin V 125mg/5ml oral solution sugar free | penicillins |
| 100 | Penicillin V 250mg capsule | penicillins |
| 102 | Penicillin V 250mg tablet | penicillins |
| 34496 | Penicillin V 250mg Tablet (Generics (UK) Ltd) | penicillins |
| 45724 | Penicillin V 250mg Tablet (Genesis Medical Ltd) | penicillins |
| 31534 | Penicillin V 250mg Tablet (Sovereign Medical Ltd) | penicillins |
| 408 | Penicillin V 250mg/5ml oral solution | penicillins |
| 43555 | Penicillin V 250mg/5ml Oral solution (C P Pharmaceuticals Ltd) | penicillins |
| 34895 | Penicillin V 250mg/5ml Oral solution (Celltech Pharma Europe Ltd) | penicillins |
| 34645 | Penicillin V 250mg/5ml Oral solution (Generics (UK) Ltd) | penicillins |
| 10079 | Penicillin V 250mg/5ml oral solution sugar free | penicillins |
| 481 | Penicillin V 250mg/5ml syrup | penicillins |
| 150 | PENICILLIN V 500 MG CAP | penicillins |
| 7884 | PENICILLIN V 500 MG TAB | penicillins |
| 224 | Penicillin V 62.5mg/5ml syrup | penicillins |
| 34756 | Penicillin VK 125mg/5ml Oral solution (Generics (UK) Ltd) | penicillins |
| 34244 | Penicillin VK 125mg/5ml Oral solution (Lagap) | penicillins |
| 46902 | Penicillin VK 125mg/5ml Oral solution (Sandoz Ltd) | penicillins |
| 34113 | Penicillin VK 250mg Tablet (Berk Pharmaceuticals Ltd) | penicillins |
| 44692 | Penicillin VK 250mg Tablet (C P Pharmaceuticals Ltd) | penicillins |
| 58367 | Penicillin VK 250mg Tablet (Celltech Pharma Europe Ltd) | penicillins |
| 34388 | Penicillin VK 250mg Tablet (Generics (UK) Ltd) | penicillins |
| 25694 | Penicillin VK 250mg Tablet (Kent Pharmaceuticals Ltd) | penicillins |
| 43759 | Penicillin VK 250mg Tablet (Lagap) | penicillins |
| 57754 | Penicillin VK 250mg Tablet (Nucross) | penicillins |
| 34909 | Penicillin VK 250mg Tablet (Sovereign Medical Ltd) | penicillins |
| 34156 | Penicillin VK 250mg/5ml Oral solution (Lagap) | penicillins |
| 18935 | Penidural Drops (Wyeth Pharmaceuticals) | penicillins |
| 25209 | PENIDURAL LA INJ | penicillins |
| 17690 | Penidural Liquid (Wyeth Pharmaceuticals) | penicillins |
| 21445 | Phenethicillin 125mg/5ml oral solution | penicillins |
| 21433 | Phenethicillin 250mg capsule | penicillins |
| 1351 | PHENOXYMETHYLPENICILLIN 125 MG CAP | penicillins |
| 4013 | PHENOXYMETHYLPENICILLIN 125 MG SYR | penicillins |
| 34105 | Phenoxymethylpenicillin 125mg tablet | penicillins |
| 4835 | Phenoxymethylpenicillin 125mg/5ml oral solution | penicillins |
| 34254 | Phenoxymethylpenicillin 125mg/5ml oral solution (A A H Pharmaceuticals Ltd) | penicillins |
| 34211 | Phenoxymethylpenicillin 125mg/5ml oral solution (Actavis UK Ltd) | penicillins |
| 49467 | Phenoxymethylpenicillin 125mg/5ml oral solution (Alliance Healthcare (Distribution) Ltd) | penicillins |
| 47241 | Phenoxymethylpenicillin 125mg/5ml oral solution (Almus Pharmaceuticals Ltd) | penicillins |
| 54310 | Phenoxymethylpenicillin 125mg/5ml oral solution (Bristol Laboratories Ltd) | penicillins |
| 53329 | Phenoxymethylpenicillin 125mg/5ml oral solution (Crescent Pharma Ltd) | penicillins |
| 61102 | Phenoxymethylpenicillin 125mg/5ml oral solution (DE Pharmaceuticals) | penicillins |
| 54270 | Phenoxymethylpenicillin 125mg/5ml oral solution (Ennogen Healthcare Ltd) | penicillins |
| 30231 | Phenoxymethylpenicillin 125mg/5ml Oral solution (Generics (UK) Ltd) | penicillins |
| 34675 | Phenoxymethylpenicillin 125mg/5ml oral solution (IVAX Pharmaceuticals UK Ltd) | penicillins |
| 34956 | Phenoxymethylpenicillin 125mg/5ml oral solution (Kent Pharmaceuticals Ltd) | penicillins |
| 48598 | Phenoxymethylpenicillin 125mg/5ml oral solution (Medreich Plc) | penicillins |
| 69193 | Phenoxymethylpenicillin 125mg/5ml oral solution (Sandoz Ltd) | penicillins |
| 51432 | Phenoxymethylpenicillin 125mg/5ml oral solution (Sigma Pharmaceuticals Plc) | penicillins |
| 34267 | Phenoxymethylpenicillin 125mg/5ml oral solution (Teva UK Ltd) | penicillins |
| 18191 | Phenoxymethylpenicillin 125mg/5ml oral solution sugar free | penicillins |
| 43022 | Phenoxymethylpenicillin 125mg/5ml oral solution sugar free (A A H Pharmaceuticals Ltd) | penicillins |
| 65785 | Phenoxymethylpenicillin 125mg/5ml oral solution sugar free (Actavis UK Ltd) | penicillins |
| 51437 | Phenoxymethylpenicillin 125mg/5ml oral solution sugar free (Alliance Healthcare (Distribution) Ltd) | penicillins |
| 62382 | Phenoxymethylpenicillin 125mg/5ml oral solution sugar free (Almus Pharmaceuticals Ltd) | penicillins |
| 51038 | Phenoxymethylpenicillin 125mg/5ml oral solution sugar free (Bristol Laboratories Ltd) | penicillins |
| 68019 | Phenoxymethylpenicillin 125mg/5ml oral solution sugar free (Brown & Burk UK Ltd) | penicillins |
| 56251 | Phenoxymethylpenicillin 125mg/5ml oral solution sugar free (Cubic Pharmaceuticals Ltd) | penicillins |
| 45898 | Phenoxymethylpenicillin 125mg/5ml oral solution sugar free (Kent Pharmaceuticals Ltd) | penicillins |
| 49832 | Phenoxymethylpenicillin 125mg/5ml oral solution sugar free (Phoenix Healthcare Distribution Ltd) | penicillins |
| 40906 | Phenoxymethylpenicillin 125mg/5ml oral solution sugar free (Teva UK Ltd) | penicillins |
| 56142 | Phenoxymethylpenicillin 125mg/5ml oral solution sugar free (Waymade Healthcare Plc) | penicillins |
| 20198 | PHENOXYMETHYLPENICILLIN 150 MG SYR | penicillins |
| 34089 | Phenoxymethylpenicillin 250mg capsule | penicillins |
| 43 | Phenoxymethylpenicillin 250mg tablets | penicillins |
| 34255 | Phenoxymethylpenicillin 250mg tablets (A A H Pharmaceuticals Ltd) | penicillins |
| 34098 | Phenoxymethylpenicillin 250mg tablets (Actavis UK Ltd) | penicillins |
| 49166 | Phenoxymethylpenicillin 250mg tablets (Alliance Healthcare (Distribution) Ltd) | penicillins |
| 42211 | Phenoxymethylpenicillin 250mg tablets (Almus Pharmaceuticals Ltd) | penicillins |
| 49807 | Phenoxymethylpenicillin 250mg tablets (Bristol Laboratories Ltd) | penicillins |
| 67039 | Phenoxymethylpenicillin 250mg tablets (Brown & Burk UK Ltd) | penicillins |
| 63415 | Phenoxymethylpenicillin 250mg tablets (Crescent Pharma Ltd) | penicillins |
| 19155 | Phenoxymethylpenicillin 250mg tablets (IVAX Pharmaceuticals UK Ltd) | penicillins |
| 49236 | Phenoxymethylpenicillin 250mg tablets (Kent Pharmaceuticals Ltd) | penicillins |
| 65364 | Phenoxymethylpenicillin 250mg tablets (Mawdsley-Brooks & Company Ltd) | penicillins |
| 49064 | Phenoxymethylpenicillin 250mg tablets (Medreich Plc) | penicillins |
| 38241 | Phenoxymethylpenicillin 250mg tablets (Mylan) | penicillins |
| 49167 | Phenoxymethylpenicillin 250mg tablets (Phoenix Healthcare Distribution Ltd) | penicillins |
| 42671 | Phenoxymethylpenicillin 250mg tablets (Ranbaxy (UK) Ltd) | penicillins |
| 43758 | Phenoxymethylpenicillin 250mg tablets (Sandoz Ltd) | penicillins |
| 64805 | Phenoxymethylpenicillin 250mg tablets (Sigma Pharmaceuticals Plc) | penicillins |
| 34346 | Phenoxymethylpenicillin 250mg tablets (Teva UK Ltd) | penicillins |
| 56942 | Phenoxymethylpenicillin 250mg tablets (The Boots Company Plc) | penicillins |
| 55311 | Phenoxymethylpenicillin 250mg tablets (Waymade Healthcare Plc) | penicillins |
| 4925 | Phenoxymethylpenicillin 250mg/5ml oral solution | penicillins |
| 34112 | Phenoxymethylpenicillin 250mg/5ml oral solution (A A H Pharmaceuticals Ltd) | penicillins |
| 31392 | Phenoxymethylpenicillin 250mg/5ml oral solution (Actavis UK Ltd) | penicillins |
| 53652 | Phenoxymethylpenicillin 250mg/5ml oral solution (Alliance Healthcare (Distribution) Ltd) | penicillins |
| 46424 | Phenoxymethylpenicillin 250mg/5ml oral solution (Almus Pharmaceuticals Ltd) | penicillins |
| 54552 | Phenoxymethylpenicillin 250mg/5ml oral solution (Bristol Laboratories Ltd) | penicillins |
| 50001 | Phenoxymethylpenicillin 250mg/5ml oral solution (Crescent Pharma Ltd) | penicillins |
| 54727 | Phenoxymethylpenicillin 250mg/5ml oral solution (Ennogen Healthcare Ltd) | penicillins |
| 42512 | Phenoxymethylpenicillin 250mg/5ml Oral solution (Generics (UK) Ltd) | penicillins |
| 34646 | Phenoxymethylpenicillin 250mg/5ml oral solution (IVAX Pharmaceuticals UK Ltd) | penicillins |
| 34659 | Phenoxymethylpenicillin 250mg/5ml oral solution (Kent Pharmaceuticals Ltd) | penicillins |
| 66496 | Phenoxymethylpenicillin 250mg/5ml oral solution (Mawdsley-Brooks & Company Ltd) | penicillins |
| 30233 | Phenoxymethylpenicillin 250mg/5ml oral solution (Mylan) | penicillins |
| 45282 | Phenoxymethylpenicillin 250mg/5ml oral solution (Sandoz Ltd) | penicillins |
| 53467 | Phenoxymethylpenicillin 250mg/5ml oral solution (Sigma Pharmaceuticals Plc) | penicillins |
| 34424 | Phenoxymethylpenicillin 250mg/5ml oral solution (Teva UK Ltd) | penicillins |
| 21405 | Phenoxymethylpenicillin 250mg/5ml oral solution sugar free | penicillins |
| 38071 | Phenoxymethylpenicillin 250mg/5ml oral solution sugar free (A A H Pharmaceuticals Ltd) | penicillins |
| 68127 | Phenoxymethylpenicillin 250mg/5ml oral solution sugar free (Actavis UK Ltd) | penicillins |
| 53876 | Phenoxymethylpenicillin 250mg/5ml oral solution sugar free (Alliance Healthcare (Distribution) Ltd) | penicillins |
| 62684 | Phenoxymethylpenicillin 250mg/5ml oral solution sugar free (Almus Pharmaceuticals Ltd) | penicillins |
| 52738 | Phenoxymethylpenicillin 250mg/5ml oral solution sugar free (Bristol Laboratories Ltd) | penicillins |
| 68806 | Phenoxymethylpenicillin 250mg/5ml oral solution sugar free (Brown & Burk UK Ltd) | penicillins |
| 54821 | Phenoxymethylpenicillin 250mg/5ml oral solution sugar free (Cubic Pharmaceuticals Ltd) | penicillins |
| 40248 | Phenoxymethylpenicillin 250mg/5ml oral solution sugar free (Kent Pharmaceuticals Ltd) | penicillins |
| 65561 | Phenoxymethylpenicillin 250mg/5ml oral solution sugar free (Mawdsley-Brooks & Company Ltd) | penicillins |
| 42222 | Phenoxymethylpenicillin 250mg/5ml oral solution sugar free (Mylan) | penicillins |
| 50609 | Phenoxymethylpenicillin 250mg/5ml oral solution sugar free (Phoenix Healthcare Distribution Ltd) | penicillins |
| 44354 | Phenoxymethylpenicillin 250mg/5ml oral solution sugar free (Sandoz Ltd) | penicillins |
| 43693 | Phenoxymethylpenicillin 250mg/5ml oral solution sugar free (Teva UK Ltd) | penicillins |
| 56246 | Phenoxymethylpenicillin 250mg/5ml oral solution sugar free (Waymade Healthcare Plc) | penicillins |
| 48140 | Phenoxymethylpenicillin 250mg/5ml oral suspension | penicillins |
| 12516 | PHENOXYMETHYLPENICILLIN 300 MG TAB | penicillins |
| 48207 | Phenoxymethylpenicillin 333mg tablet | penicillins |
| 34183 | Phenoxymethylpenicillin 62.5mg/5ml oral solution | penicillins |
| 53888 | Phenoxymethylpenicillin 666mg tablet | penicillins |
| 16594 | Piperacillin 1g/vial injection | penicillins |
| 693 | Piperacillin 2g / Tazobactam 250mg powder for solution for infusion vials | penicillins |
| 71473 | Piperacillin 2g / Tazobactam 250mg powder for solution for infusion vials (Bowmed Ibisqus Ltd) | penicillins |
| 72041 | Piperacillin 2g / Tazobactam 250mg powder for solution for injection vials (A A H Pharmaceuticals Ltd) | penicillins |
| 21737 | Piperacillin 2g/vial injection | penicillins |
| 17323 | Piperacillin 4g / Tazobactam 500mg powder for solution for infusion vials | penicillins |
| 31174 | Piperacillin 4g/infusion bottle infusion | penicillins |
| 25832 | Pivampicillin 125mg with pivmecillinam100mg tablet | penicillins |
| 17181 | Pivampicillin 175mg sachet | penicillins |
| 22118 | PIVAMPICILLIN 175mg/5ml | penicillins |
| 8614 | Pivampicillin 175mg/5ml oral solution | penicillins |
| 12540 | Pivampicillin 250mg with pivmecillinam 200mg tablet | penicillins |
| 24974 | PIVAMPICILLIN 250MG/PIVMECILLINAM 200MG | penicillins |
| 7570 | Pivampicillin 500mg tablet | penicillins |
| 26101 | Pivmecillinam 100mg/sachet | penicillins |
| 12014 | Pivmecillinam 200mg tablets | penicillins |
| 75738 | Pivmecillinam 200mg with pivampicillin 250mg tablet | penicillins |
| 68786 | Pivmecillinam 200mg/5ml oral suspension | penicillins |
| 23586 | PONDOCILLIN | penicillins |
| 31669 | Pondocillin 120mg Sachets (LEO Pharma) | penicillins |
| 19555 | PONDOCILLIN 175mg/5ml | penicillins |
| 2377 | Pondocillin 175mg/5ml Oral suspension sugar free (LEO Pharma) | penicillins |
| 2246 | Pondocillin 500mg Tablet (LEO Pharma) | penicillins |
| 8960 | Pondocillin plus Tablet (Edwin Burgess Ltd) | penicillins |
| 21071 | PONDOCILLIN sach 175 MG | penicillins |
| 23607 | PONDOCILLIN SACHET | penicillins |
| 74288 | Procaine benzylpenicillin 1.5g/3.4ml suspension for injection pre-filled syringes | penicillins |
| 22808 | Procaine benzylpenicillin injection | penicillins |
| 15354 | Procaine benzylpenicillin with benzylpenicillin sodium injection | penicillins |
| 31778 | PROCAINE PENICILLIN 3 GM INJ | penicillins |
| 22670 | PROCAINE PENICILLIN/BENZYLPENICILLIN 3 GM INJ | penicillins |
| 18961 | Pyopen 1g sterile Powder (Link Pharmaceuticals Ltd) | penicillins |
| 27681 | Ranclav 125mg/31mg/5ml SF oral suspension (Ranbaxy (UK) Ltd) | penicillins |
| 25370 | Ranclav 375mg tablets (Ranbaxy (UK) Ltd) | penicillins |
| 22015 | Respillin 125mg/5ml Oral solution (OPD Pharm) | penicillins |
| 22017 | Respillin 125mg/5ml Oral solution (OPD Pharm) | penicillins |
| 24203 | Respillin 250mg Capsule (OPD Pharm) | penicillins |
| 69398 | Respillin 250mg capsules (Kent Pharmaceuticals Ltd) | penicillins |
| 24200 | Respillin 500mg Capsule (OPD Pharm) | penicillins |
| 32363 | Securopen 2g/vial Injection (Bayer Plc) | penicillins |
| 32318 | Securopen 5g/vial Infusion (Bayer Plc) | penicillins |
| 12015 | Selexid 100mg/sachet Liquid (Edwin Burgess Ltd) | penicillins |
| 9601 | Selexid 200mg tablets (Karo Pharma) | penicillins |
| 26215 | Stabilin V-K 250mg Tablet (Knoll Ltd) | penicillins |
| 26225 | Stabilin V-K 250mg/5ml Oral solution (Knoll Ltd) | penicillins |
| 33237 | Stabilin V-K 62.5mg/5ml Oral solution (Knoll Ltd) | penicillins |
| 32361 | Stafoxil 250mg Capsule (Yamanouchi Pharma Ltd) | penicillins |
| 16589 | Talampicillin 125mg/5ml syrup | penicillins |
| 20009 | TALAMPICILLIN 250 MG SYR | penicillins |
| 20007 | Talampicillin 250mg tablets | penicillins |
| 8680 | Talpen 125mg/5ml Oral solution (Beecham Research Laboratories) | penicillins |
| 14368 | TALPEN 250 MG SYR | penicillins |
| 11954 | Talpen 250mg Tablet (Beecham Research Laboratories) | penicillins |
| 19368 | Tazocin 2g/0.25g powder for solution for infusion vials (Pfizer Ltd) | penicillins |
| 694 | Tazocin 4.5g Powder for solution for injection (Pfizer Consumer Healthcare Ltd) | penicillins |
| 49724 | Tazocin 4g/0.5g powder for solution for infusion vials (Pfizer Ltd) | penicillins |
| 33420 | Temocillin 1g powder for solution for injection vials | penicillins |
| 25914 | Temopen 1g/vial Powder (Bencard) | penicillins |
| 29589 | Ticar 1g/vial Injection (Link Pharmaceuticals Ltd) | penicillins |
| 30615 | Ticar 5g/vial Injection (Link Pharmaceuticals Ltd) | penicillins |
| 18962 | Ticarcillin 1g/vial Injection | penicillins |
| 33421 | Ticarcillin 3g / Clavulanic acid 200mg powder for solution for infusion vials | penicillins |
| 23602 | Ticarcillin 3g/vial Injection | penicillins |
| 25938 | Ticarcillin 5g/vial Injection | penicillins |
| 23758 | Timentin 1.6g/vial Infusion (Beecham Research Laboratories) | penicillins |
| 24063 | Timentin 3.2g powder for solution for infusion vials (GlaxoSmithKline UK Ltd) | penicillins |
| 7942 | Triplopen Injection (Glaxo Laboratories Ltd) | penicillins |
| 8515 | Uticillin 500mg Tablet (Beecham Research Laboratories) | penicillins |
| 23245 | V-CIL-K | penicillins |
| 12365 | V-cil-k 125mg Tablet (Eli Lilly and Company Ltd) | penicillins |
| 10584 | V-cil-k 125mg/5ml Oral solution (Eli Lilly and Company Ltd) | penicillins |
| 13400 | V-cil-k 250mg Capsule (Eli Lilly and Company Ltd) | penicillins |
| 10647 | V-cil-k 250mg Tablet (Eli Lilly and Company Ltd) | penicillins |
| 10355 | V-cil-k 250mg/5ml Oral solution (Eli Lilly and Company Ltd) | penicillins |
| 23247 | V-CIL-K 500 MG PUL | penicillins |
| 41985 | V-cil-k 62.5mg/5ml Oral solution (Eli Lilly and Company Ltd) | penicillins |
| 21995 | V-CIL-K PAEDIATRIC | penicillins |
| 23186 | Vidopen 125mg/5ml Oral solution (Berk Pharmaceuticals Ltd) | penicillins |
| 21801 | Vidopen 250mg Capsule (Berk Pharmaceuticals Ltd) | penicillins |
| 31471 | Vidopen 250mg/5ml Oral solution (Berk Pharmaceuticals Ltd) | penicillins |
| 31473 | Vidopen 250mg/vial Injection (Berk Pharmaceuticals Ltd) | penicillins |
| 21967 | Vidopen 500mg Capsule (Berk Pharmaceuticals Ltd) | penicillins |
| 21850 | Zoxin 250mg Capsule (Opus Pharmaceuticals Ltd) | penicillins |
| 21829 | Zoxycil 250mg Capsule (Trinity Pharmaceuticals Ltd) | penicillins |
| 26262 | Zoxycil 500mg Capsule (Trinity Pharmaceuticals Ltd) | penicillins |
| 31744 | BACTRIM | sulfonamides |
| 21957 | BACTRIM ADULT SUSPENSION | sulfonamides |
| 21922 | BACTRIM DISPERSIBLE | sulfonamides |
| 23611 | BACTRIM DOUBLE STRENGTH | sulfonamides |
| 23248 | BACTRIM DRAPSULES | sulfonamides |
| 2725 | CO-TRIMOXAZOLE 100 MG TAB | sulfonamides |
| 31463 | Co-trimoxazole 160mg/800mg/10ml solution for infusion ampoules | sulfonamides |
| 4555 | CO-TRIMOXAZOLE 200 MG SUS | sulfonamides |
| 8394 | CO-TRIMOXAZOLE 80 MG SYR | sulfonamides |
| 8900 | CO-TRIMOXAZOLE 800 MG TAB | sulfonamides |
| 17738 | CO-TRIMOXAZOLE 96 MG INJ | sulfonamides |
| 10632 | CO-TRIMOXAZOLE F/C 480 MG TAB | sulfonamides |
| 3583 | CO-TRIMOXAZOLE PAED 120 MG TAB | sulfonamides |
| 10720 | CO-TRIMOXAZOLE PAED 200 MG SYR | sulfonamides |
| 20524 | GX CO-TRIMOXAZOLE 480 MG TAB | sulfonamides |
| 13306 | Monotrim 100mg/5ml solution for injection ampoules (Abbott Healthcare Products Ltd) | sulfonamides |
| 436 | Sulfadiazine 1g/4ml solution for injection ampoules | sulfonamides |
| 25074 | TRIMETHOPRIM 0.1%/POLYMIXIN B 10000U EYE | sulfonamides |
| 15062 | TRIMETHOPRIM 0.1%/POLYMIXIN B 10000U EYE DRO | sulfonamides |
| 226 | TRIMETHOPRIM 100 MG CAP | sulfonamides |
| 153 | Trimethoprim 100mg/5ml solution for injection ampoules | sulfonamides |
| 12466 | TRIMETHOPRIM 1MG/POLYMIXIN B 10000U EYE OIN | sulfonamides |
| 53793 | Trimethoprim 50mg/5ml oral suspension sugar free (Almus Pharmaceuticals Ltd) | sulfonamides |
| 21916 | COMIXCO 160 800 MG TAB | sulfonamides |
| 23660 | KELFIZINE W 2 GM SYR | sulfonamides |
| 27906 | SYRAPRIM ML INJ | sulfonamides |
| 1467 | Co-trimoxazole 160mg/800mg tablets | sulfonamides |
| 70023 | Co-trimoxazole 160mg/800mg tablets (A A H Pharmaceuticals Ltd) | sulfonamides |
| 68826 | Co-trimoxazole 160mg/800mg tablets (Alliance Healthcare (Distribution) Ltd) | sulfonamides |
| 73744 | Co-trimoxazole 160mg/800mg tablets (Aspen Pharma Trading Ltd) | sulfonamides |
| 79715 | Co-trimoxazole 160mg/800mg tablets (CST Pharma Ltd) | sulfonamides |
| 78965 | Co-trimoxazole 160mg/800mg tablets (Genesis Pharmaceuticals Ltd) | sulfonamides |
| 65343 | Co-trimoxazole 160mg/800mg tablets (Tillomed Laboratories Ltd) | sulfonamides |
| 1199 | Co-trimoxazole 40mg/200mg/5ml oral suspension sugar free | sulfonamides |
| 72595 | Co-trimoxazole 40mg/200mg/5ml oral suspension sugar free (A A H Pharmaceuticals Ltd) | sulfonamides |
| 73702 | Co-trimoxazole 40mg/200mg/5ml oral suspension sugar free (Alliance Healthcare (Distribution) Ltd) | sulfonamides |
| 68101 | Co-trimoxazole 40mg/200mg/5ml oral suspension sugar free (Aspen Pharma Trading Ltd) | sulfonamides |
| 606 | Co-trimoxazole 80mg/400mg tablets | sulfonamides |
| 41579 | Co-trimoxazole 80mg/400mg tablets (A A H Pharmaceuticals Ltd) | sulfonamides |
| 34727 | Co-trimoxazole 80mg/400mg tablets (Actavis UK Ltd) | sulfonamides |
| 59444 | Co-trimoxazole 80mg/400mg tablets (Alliance Healthcare (Distribution) Ltd) | sulfonamides |
| 69881 | Co-trimoxazole 80mg/400mg tablets (Aspen Pharma Trading Ltd) | sulfonamides |
| 69653 | Co-trimoxazole 80mg/400mg tablets (Crescent Pharma Ltd) | sulfonamides |
| 73695 | Co-trimoxazole 80mg/400mg tablets (DE Pharmaceuticals) | sulfonamides |
| 63733 | Co-trimoxazole 80mg/400mg tablets (Essential Generics Ltd) | sulfonamides |
| 72447 | Co-trimoxazole 80mg/400mg tablets (Genesis Pharmaceuticals Ltd) | sulfonamides |
| 54914 | Co-trimoxazole 80mg/400mg tablets (Kent Pharmaceuticals Ltd) | sulfonamides |
| 69493 | Co-trimoxazole 80mg/400mg tablets (Phoenix Healthcare Distribution Ltd) | sulfonamides |
| 68726 | Co-trimoxazole 80mg/400mg tablets (Sigma Pharmaceuticals Plc) | sulfonamides |
| 52198 | Co-trimoxazole 80mg/400mg tablets (Sigma Pharmaceuticals Plc) | sulfonamides |
| 79195 | Co-trimoxazole 80mg/400mg tablets (Tillomed Laboratories Ltd) | sulfonamides |
| 58282 | Co-trimoxazole 80mg/400mg tablets (Waymade Healthcare Plc) | sulfonamides |
| 2658 | Co-trimoxazole 80mg/400mg/5ml oral suspension | sulfonamides |
| 78909 | Co-trimoxazole 80mg/400mg/5ml oral suspension (Alliance Healthcare (Distribution) Ltd) | sulfonamides |
| 77838 | Co-trimoxazole 80mg/400mg/5ml oral suspension (Aspen Pharma Trading Ltd) | sulfonamides |
| 16620 | Co-trimoxazole 80mg/400mg/5ml solution for infusion ampoules | sulfonamides |
| 60448 | Co-trimoxazole 80mg/400mg/5ml solution for infusion ampoules (Alliance Healthcare (Distribution) Ltd) | sulfonamides |
| 69814 | Co-trimoxazole 80mg/400mg/5ml solution for infusion ampoules (Aspen Pharma Trading Ltd) | sulfonamides |
| 7962 | Kelfizine w 2g Tablet (Pharmacia Ltd) | sulfonamides |
| 13325 | Monotrim 100mg tablets (Abbott Healthcare Products Ltd) | sulfonamides |
| 8171 | Monotrim 200mg tablets (Abbott Healthcare Products Ltd) | sulfonamides |
| 36622 | Monotrim 50mg/5ml oral suspension (Chemidex Pharma Ltd) | sulfonamides |
| 109 | Septrin Adult 80mg/400mg/5ml oral suspension (Aspen Pharma Trading Ltd) | sulfonamides |
| 131 | Septrin for Infusion 80mg/400mg/5ml solution for infusion ampoules (Aspen Pharma Trading Ltd) | sulfonamides |
| 2460 | Septrin Forte 160mg/800mg tablets (Aspen Pharma Trading Ltd) | sulfonamides |
| 44286 | Septrin Paediatric 40mg/200mg/5ml oral suspension (Aspen Pharma Trading Ltd) | sulfonamides |
| 44075 | Septrin tablets (Aspen Pharma Trading Ltd) | sulfonamides |
| 76876 | Sulfadiazine 500mg tablets (Kent Pharmaceuticals Ltd) | sulfonamides |
| 54166 | Sulfadiazine oral solution | sulfonamides |
| 298 | Sulfadimidine 333mg/ml injection | sulfonamides |
| 17729 | Sulfadimidine 500mg tablet | sulfonamides |
| 29994 | Sulfaguanidine 500mg tablet | sulfonamides |
| 128 | Sulfametopyrazine 2g tablet | sulfonamides |
| 29357 | Sulphadimethoxine 500mg tablet | sulfonamides |
| 27417 | Sulphafurazole 500mg tablet | sulfonamides |
| 260 | Sulphafurazole 500mg/5ml oral solution | sulfonamides |
| 24856 | Sulphamezathine 333mg/ml Injection (AstraZeneca UK Ltd) | sulfonamides |
| 20523 | Thalazole 500mg Tablet (May and Baker) | sulfonamides |
| 340 | Trimethoprim 100mg tablets | sulfonamides |
| 32906 | Trimethoprim 100mg tablets (A A H Pharmaceuticals Ltd) | sulfonamides |
| 43545 | Trimethoprim 100mg tablets (Actavis UK Ltd) | sulfonamides |
| 58490 | Trimethoprim 100mg tablets (Alliance Healthcare (Distribution) Ltd) | sulfonamides |
| 53720 | Trimethoprim 100mg tablets (Almus Pharmaceuticals Ltd) | sulfonamides |
| 56267 | Trimethoprim 100mg tablets (Bristol Laboratories Ltd) | sulfonamides |
| 68225 | Trimethoprim 100mg tablets (Crescent Pharma Ltd) | sulfonamides |
| 67596 | Trimethoprim 100mg tablets (DE Pharmaceuticals) | sulfonamides |
| 34488 | Trimethoprim 100mg tablets (Kent Pharmaceuticals Ltd) | sulfonamides |
| 67147 | Trimethoprim 100mg tablets (Mawdsley-Brooks & Company Ltd) | sulfonamides |
| 49592 | Trimethoprim 100mg tablets (Phoenix Healthcare Distribution Ltd) | sulfonamides |
| 45246 | Trimethoprim 100mg tablets (Sandoz Ltd) | sulfonamides |
| 56259 | Trimethoprim 100mg tablets (Sigma Pharmaceuticals Plc) | sulfonamides |
| 34542 | Trimethoprim 100mg tablets (Teva UK Ltd) | sulfonamides |
| 57080 | Trimethoprim 100mg tablets (Waymade Healthcare Plc) | sulfonamides |
| 37 | Trimethoprim 200mg tablets | sulfonamides |
| 27255 | Trimethoprim 200mg tablets (A A H Pharmaceuticals Ltd) | sulfonamides |
| 50120 | Trimethoprim 200mg tablets (Accord Healthcare Ltd) | sulfonamides |
| 34392 | Trimethoprim 200mg tablets (Actavis UK Ltd) | sulfonamides |
| 50797 | Trimethoprim 200mg tablets (Alliance Healthcare (Distribution) Ltd) | sulfonamides |
| 39933 | Trimethoprim 200mg tablets (Almus Pharmaceuticals Ltd) | sulfonamides |
| 51510 | Trimethoprim 200mg tablets (Bristol Laboratories Ltd) | sulfonamides |
| 64028 | Trimethoprim 200mg tablets (Crescent Pharma Ltd) | sulfonamides |
| 65487 | Trimethoprim 200mg tablets (DE Pharmaceuticals) | sulfonamides |
| 33997 | Trimethoprim 200mg tablets (IVAX Pharmaceuticals UK Ltd) | sulfonamides |
| 34379 | Trimethoprim 200mg tablets (Kent Pharmaceuticals Ltd) | sulfonamides |
| 65497 | Trimethoprim 200mg tablets (Mawdsley-Brooks & Company Ltd) | sulfonamides |
| 53599 | Trimethoprim 200mg tablets (Phoenix Healthcare Distribution Ltd) | sulfonamides |
| 62630 | Trimethoprim 200mg tablets (Ranbaxy (UK) Ltd) | sulfonamides |
| 34633 | Trimethoprim 200mg tablets (Sandoz Ltd) | sulfonamides |
| 32908 | Trimethoprim 200mg tablets (Teva UK Ltd) | sulfonamides |
| 57981 | Trimethoprim 200mg tablets (Waymade Healthcare Plc) | sulfonamides |
| 52669 | Trimethoprim 200mg/5ml oral solution | sulfonamides |
| 55986 | Trimethoprim 200mg/5ml oral suspension | sulfonamides |
| 57642 | Trimethoprim 20mg/5ml oral solution | sulfonamides |
| 51725 | Trimethoprim 20mg/5ml oral suspension | sulfonamides |
| 75085 | Trimethoprim 240mg/5ml oral solution | sulfonamides |
| 477 | Trimethoprim 50mg/5ml oral suspension sugar free | sulfonamides |
| 34252 | Trimethoprim 50mg/5ml oral suspension sugar free (A A H Pharmaceuticals Ltd) | sulfonamides |
| 53828 | Trimethoprim 50mg/5ml oral suspension sugar free (Actavis UK Ltd) | sulfonamides |
| 53275 | Trimethoprim 50mg/5ml oral suspension sugar free (Alliance Healthcare (Distribution) Ltd) | sulfonamides |
| 60808 | Trimethoprim 50mg/5ml oral suspension sugar free (Almus Pharmaceuticals Ltd) | sulfonamides |
| 78888 | Trimethoprim 50mg/5ml oral suspension sugar free (DE Pharmaceuticals) | sulfonamides |
| 53276 | Trimethoprim 50mg/5ml oral suspension sugar free (Kent Pharmaceuticals Ltd) | sulfonamides |
| 67361 | Trimethoprim 50mg/5ml oral suspension sugar free (Phoenix Healthcare Distribution Ltd) | sulfonamides |
| 61714 | Trimethoprim 50mg/5ml oral suspension sugar free (Pinewood Healthcare) | sulfonamides |
| 53284 | Trimethoprim 50mg/5ml oral suspension sugar free (Sigma Pharmaceuticals Plc) | sulfonamides |
| 29351 | Trimethoprim 50mg/5ml oral suspension sugar free (Teva UK Ltd) | sulfonamides |
| 57116 | Trimethoprim 50mg/5ml oral suspension sugar free (Waymade Healthcare Plc) | sulfonamides |
| 73626 | Trimethoprim 7mg/5ml oral solution | sulfonamides |
| 21640 | Trimopan 100mg tablets (Teva UK Ltd) | sulfonamides |
| 10046 | Trimopan 200mg tablets (Teva UK Ltd) | sulfonamides |
| 21037 | Uromide Tablet (Consolidated Chemicals (UK) Ltd) | sulfonamides |
| 29800 | Phthalylsulfathiazole 500mg tablet | sulfonamides |
| 15081 | Ipral 100mg Tablet (E R Squibb and Sons Ltd) | sulfonamides |
| 14998 | Ipral 200mg Tablet (E R Squibb and Sons Ltd) | sulfonamides |
| 14367 | Ipral 50mg/5ml Liquid (E R Squibb and Sons Ltd) | sulfonamides |
| 280 | Monotrim 50mg/5ml Liquid (Solvay Healthcare) | sulfonamides |
| 25497 | Syraprim 100mg Tablet (Wellcome Medical Division) | sulfonamides |
| 27048 | Syraprim 300mg Tablet (Wellcome Medical Division) | sulfonamides |
| 34878 | Trimethoprim 100mg Tablet (C P Pharmaceuticals Ltd) | sulfonamides |
| 75766 | Trimethoprim 100mg Tablet (Celltech Pharma Europe Ltd) | sulfonamides |
| 41544 | Trimethoprim 100mg Tablet (IVAX Pharmaceuticals UK Ltd) | sulfonamides |
| 34455 | Trimethoprim 200mg Tablet (C P Pharmaceuticals Ltd) | sulfonamides |
| 43537 | Trimethoprim 200mg Tablet (Celltech Pharma Europe Ltd) | sulfonamides |
| 43505 | Trimethoprim 200mg Tablet (Numark Management Ltd) | sulfonamides |
| 31227 | Trimethoprim 200mg Tablet (Regent Laboratories Ltd) | sulfonamides |
| 8073 | Trimethoprim 300mg Tablet | sulfonamides |
| 78999 | Trimethoprim Oral solution | sulfonamides |
| 29532 | Trimogal 100mg Tablet (Lagap) | sulfonamides |
| 24324 | Trimogal 200mg Tablet (Lagap) | sulfonamides |
| 7616 | Trimopan 50mg/5ml Liquid (Berk Pharmaceuticals Ltd) | sulfonamides |
| 21805 | Triprimix 200 Tablet (Ashbourne Pharmaceuticals Ltd) | sulfonamides |
| 403 | Sulfadiazine 500mg tablets | sulfonamides |
| 55121 | Sulfadiazine 500mg tablets (A A H Pharmaceuticals Ltd) | sulfonamides |
| 53946 | Sulfadiazine 500mg tablets (Alliance Healthcare (Distribution) Ltd) | sulfonamides |
| 43509 | Sulfadiazine 500mg tablets (Wockhardt UK Ltd) | sulfonamides |
| 7420 | Bactrim 480mg Tablet (Roche Products Ltd) | sulfonamides |
| 60216 | Bactrim 96mg/ml Infusion (Roche Products Ltd) | sulfonamides |
| 7421 | Bactrim adult 480mg/5ml Liquid (Roche Products Ltd) | sulfonamides |
| 8741 | Bactrim Dispersible tablet (Roche Products Ltd) | sulfonamides |
| 10745 | Bactrim double strength 160mg+800mg Tablet (Roche Products Ltd) | sulfonamides |
| 25269 | Bactrim im 320mg/ml intramuscular injection (Roche Products Ltd) | sulfonamides |
| 8561 | Bactrim paediatric sugar free oral solution | sulfonamides |
| 46663 | Bactrim paediatric tablets (Roche Products Ltd) | sulfonamides |
| 22991 | Chemotrim Liquid (Rosemont Pharmaceuticals Ltd) | sulfonamides |
| 21809 | Comixco 80mg+400mg Tablet (Ashbourne Pharmaceuticals Ltd) | sulfonamides |
| 68990 | Comox forte Tablet (IVAX Pharmaceuticals UK Ltd) | sulfonamides |
| 29907 | Comox Tablet (IVAX Pharmaceuticals UK Ltd) | sulfonamides |
| 9100 | Co-trimoxazole (trimethoprim and sulfamethoxazole) 160mg+800mg dispersible tablets | sulfonamides |
| 28004 | Co-trimoxazole (trimethoprim and sulfamethoxazole) 20mg+100mg paediatric tablets | sulfonamides |
| 79048 | Co-trimoxazole (trimethoprim and sulfamethoxazole) 40mg+200mg paediatric tablets | sulfonamides |
| 8286 | Co-trimoxazole (trimethoprim and sulfamethoxazole) 80mg+400mg dispersible tablets | sulfonamides |
| 287 | Co-trimoxazole (trimethoprim with sulfamethoxazole) 320mg/ml IM injection | sulfonamides |
| 68027 | Co-trimoxazole 160mg+800mg Tablet (C P Pharmaceuticals Ltd) | sulfonamides |
| 303 | Co-trimoxazole 16mg with 80mg/ml concentrate solution for infusion | sulfonamides |
| 41978 | Co-trimoxazole 240mg/5ml Oral suspension (Approved Prescription Services Ltd) | sulfonamides |
| 43262 | Co-trimoxazole 240mg/5ml Oral suspension (C P Pharmaceuticals Ltd) | sulfonamides |
| 41967 | Co-trimoxazole 240mg/5ml Oral suspension (Hillcross Pharmaceuticals Ltd) | sulfonamides |
| 30201 | Co-trimoxazole 240mg/5ml Paediatric mixture (Lagap) | sulfonamides |
| 42517 | Co-trimoxazole 40mg+200mg Liquid (Celltech Pharma Europe Ltd) | sulfonamides |
| 38090 | Co-trimoxazole 480mg/5ml Adult Mixture (Lagap) | sulfonamides |
| 41991 | Co-trimoxazole 80mg+400mg Dispersible tablet (Approved Prescription Services Ltd) | sulfonamides |
| 67613 | Co-trimoxazole 80mg+400mg Dispersible tablet (IVAX Pharmaceuticals UK Ltd) | sulfonamides |
| 73468 | Co-trimoxazole 80mg+400mg Tablet (Approved Prescription Services Ltd) | sulfonamides |
| 79357 | Co-trimoxazole 80mg+400mg Tablet (C P Pharmaceuticals Ltd) | sulfonamides |
| 44241 | Fectrim Dispersible tablet (DDSA Pharmaceuticals Ltd) | sulfonamides |
| 31484 | Laratrim adult 480mg/5ml Liquid (Lagap) | sulfonamides |
| 27921 | Laratrim forte Tablet (Lagap) | sulfonamides |
| 31477 | Laratrim Liquid (Lagap) | sulfonamides |
| 3660 | Septrin Dispersible tablet (Wellcome Medical Division) | sulfonamides |
| 69362 | Septrin im 320mg/ml intramuscular injection (Wellcome Medical Division) | sulfonamides |
| 1634 | Septrin paediatric Dispersible tablet (Wellcome Medical Division) | sulfonamides |
| 1604 | Septrin paediatric Oral suspension sugar free (Wellcome Medical Division) | sulfonamides |
| 135 | Septrin Tablet (Wellcome Medical Division) | sulfonamides |
| 30614 | Sulfamethoxazole 200mg with trimethoprim 40mg/5ml oral suspension | sulfonamides |
| 10308 | Sulfamethoxazole 400mg with trimethoprim 80mg tablet | sulfonamides |
| 20368 | Sulfamethoxazole 400mg with trimethoprim 80mg/5ml concentrate solution for infusion | sulfonamides |
| 33794 | Sulfamethoxazole 400mg with trimethoprim 80mg/5ml oral suspension | sulfonamides |
| 25908 | Sulfamethoxazole 800mg with trimethoprim 160mg tablet | sulfonamides |
| 33987 | Sulfamethoxazole 800mg with trimethoprim 160mg/5ml concentrate solution for infusion | sulfonamides |
| 10318 | Sulfamethoxazole 80mg with trimethoprim 16mg/5ml concentrate solution for infusion | sulfonamides |
| 27445 | Trimethoprim with sulfamethoxazole 160mg + 800mg/10ml Concentrate for solution for infusion | sulfonamides |
| 20126 | Trimethoprim with sulfamethoxazole 160mg+800mg Tablet | sulfonamides |
| 45757 | Trimethoprim with sulfamethoxazole 16mg + 80mg/ml Concentrate for solution for infusion | sulfonamides |
| 27418 | Trimethoprim with sulfamethoxazole 40mg + 200mg/5ml Oral suspension | sulfonamides |
| 31905 | Trimethoprim with sulfamethoxazole 80mg + 400mg/5ml Concentrate for solution for infusion | sulfonamides |
| 20920 | Trimethoprim with sulfamethoxazole 80mg + 400mg/5ml Oral suspension | sulfonamides |
| 15988 | Trimethoprim with sulfamethoxazole 80mg+400mg Tablet | sulfonamides |
| 18685 | Achromycin 125mg/5ml Oral solution (Wyeth Pharmaceuticals) | tetracyclines |
| 4579 | Achromycin 250mg capsules (Wyeth Pharmaceuticals) | tetracyclines |
| 15513 | Achromycin 250mg Tablet (Wyeth Pharmaceuticals) | tetracyclines |
| 28573 | Achromycin im 100mg/vial intramuscular injection (Wyeth Pharmaceuticals) | tetracyclines |
| 31476 | Achromycin iv 250mg/vial Intravenous injection (Wyeth Pharmaceuticals) | tetracyclines |
| 31230 | Achromycin iv 500mg/vial Intravenous injection (Wyeth Pharmaceuticals) | tetracyclines |
| 32233 | Achromycin Powder (Wyeth Pharmaceuticals) | tetracyclines |
| 15407 | Achromycin v 250mg Capsule (Wyeth Pharmaceuticals) | tetracyclines |
| 54152 | Acnamino MR 100mg capsules (Almus Pharmaceuticals Ltd) | tetracyclines |
| 14984 | Acnamino MR 100mg capsules (Dexcel-Pharma Ltd) | tetracyclines |
| 77482 | Adjusan 36.4mg/260mg periodontal gel cartridge (Heraeus) | tetracyclines |
| 18728 | Aknemin 100mg capsules (Almirall Ltd) | tetracyclines |
| 18684 | Aknemin 50 capsules (Almirall Ltd) | tetracyclines |
| 2127 | Aureomycin 250mg Capsule (Wyeth Pharmaceuticals) | tetracyclines |
| 21802 | Berkmycen 250mg Tablet (Berk Pharmaceuticals Ltd) | tetracyclines |
| 17093 | Bisolvomycin Capsule (Boehringer Ingelheim Ltd) | tetracyclines |
| 21978 | Blemix 100mg tablets (Ashbourne Pharmaceuticals Ltd) | tetracyclines |
| 21865 | Blemix 50mg tablets (Ashbourne Pharmaceuticals Ltd) | tetracyclines |
| 7881 | Chlortetracycline 250mg capsules | tetracyclines |
| 17284 | CHLORTETRACYCLINE HYD./DEMECLOCYCLINE HY 115.4 MG TAB | tetracyclines |
| 738 | Chlortetracycline with demeclocycline with tetracycline tablets | tetracyclines |
| 12016 | Chymocyclar Capsule (Rorer Pharmaceuticals Ltd) | tetracyclines |
| 12504 | Clomocycline 170mg capsules | tetracyclines |
| 21860 | Cyclodox 100mg Capsule (Berk Pharmaceuticals Ltd) | tetracyclines |
| 21810 | CYCLODOX CAP 100 mg | tetracyclines |
| 24245 | Cyclomin 100mg Tablet (Berk Pharmaceuticals Ltd) | tetracyclines |
| 21837 | Cyclomin 50mg Tablet (Berk Pharmaceuticals Ltd) | tetracyclines |
| 9131 | Demeclocycline 150mg capsules | tetracyclines |
| 62533 | Demeclocycline 150mg tablets | tetracyclines |
| 78831 | Demeclocycline 150mg/5ml oral solution | tetracyclines |
| 78035 | Demeclocycline 300mg capsules | tetracyclines |
| 8694 | Demeclocycline 300mg tablets | tetracyclines |
| 50765 | Demeclocycline 300mg/5ml oral solution | tetracyclines |
| 24643 | Demeclocycline with chlortetracycline with tetracycline tablets | tetracyclines |
| 21878 | Demix 100 capsules (Ashbourne Pharmaceuticals Ltd) | tetracyclines |
| 21817 | DEMIX 100 MG CAP | tetracyclines |
| 21828 | Demix 50 capsules (Ashbourne Pharmaceuticals Ltd) | tetracyclines |
| 20674 | DETECLO | tetracyclines |
| 2256 | Deteclo 300mg Tablet (Wyeth Pharmaceuticals) | tetracyclines |
| 13327 | Deteclo 300mg tablets (Mercury Pharma Group Ltd) | tetracyclines |
| 14369 | DETECLO 75 MG SYR | tetracyclines |
| 21038 | Doxatet 100mg Tablet (Manufacturer unknown) | tetracyclines |
| 2884 | Doxycycline (as hyclate) 100mg dispersible tablets | tetracyclines |
| 970 | Doxycycline (as hyclate) 100mg tablets | tetracyclines |
| 12987 | Doxycycline (as hyclate) 50mg capsules with microgranules | tetracyclines |
| 23819 | Doxycycline (as hyclate) 50mg capsules with microgranules | tetracyclines |
| 8724 | Doxycycline (as hyclate) 50mg/5ml oral solution | tetracyclines |
| 41560 | Doxycycline 100mg Capsule (IVAX Pharmaceuticals UK Ltd) | tetracyclines |
| 34594 | Doxycycline 100mg Capsule (Neo Laboratories Ltd) | tetracyclines |
| 34423 | Doxycycline 100mg Capsule (PLIVA Pharma Ltd) | tetracyclines |
| 41605 | Doxycycline 100mg Capsule (Sandoz Ltd) | tetracyclines |
| 1046 | Doxycycline 100mg capsules | tetracyclines |
| 24149 | Doxycycline 100mg capsules (A A H Pharmaceuticals Ltd) | tetracyclines |
| 34300 | Doxycycline 100mg capsules (Actavis UK Ltd) | tetracyclines |
| 49737 | Doxycycline 100mg capsules (Alliance Healthcare (Distribution) Ltd) | tetracyclines |
| 46807 | Doxycycline 100mg capsules (Almus Pharmaceuticals Ltd) | tetracyclines |
| 60159 | Doxycycline 100mg capsules (DE Pharmaceuticals) | tetracyclines |
| 62025 | Doxycycline 100mg capsules (Healthcare Pharma Ltd) | tetracyclines |
| 24126 | Doxycycline 100mg capsules (IVAX Pharmaceuticals UK Ltd) | tetracyclines |
| 33671 | Doxycycline 100mg capsules (Kent Pharmaceuticals Ltd) | tetracyclines |
| 68690 | Doxycycline 100mg capsules (Mawdsley-Brooks & Company Ltd) | tetracyclines |
| 32066 | Doxycycline 100mg capsules (Mylan) | tetracyclines |
| 58988 | Doxycycline 100mg capsules (Phoenix Healthcare Distribution Ltd) | tetracyclines |
| 53310 | Doxycycline 100mg capsules (Sigma Pharmaceuticals Plc) | tetracyclines |
| 30739 | Doxycycline 100mg capsules (Teva UK Ltd) | tetracyclines |
| 55519 | Doxycycline 100mg capsules (Waymade Healthcare Plc) | tetracyclines |
| 6396 | Doxycycline 100mg dispersible tablets sugar free | tetracyclines |
| 26747 | Doxycycline 100mg Tablet (Neo Laboratories Ltd) | tetracyclines |
| 68110 | Doxycycline 100mg/5ml oral suspension | tetracyclines |
| 70869 | Doxycycline 36.4mg/260mg periodontal gel cartridge | tetracyclines |
| 40796 | Doxycycline 40mg modified-release capsules | tetracyclines |
| 79404 | Doxycycline 50mg Capsule (Merck Generics (UK) Ltd) | tetracyclines |
| 264 | Doxycycline 50mg capsules | tetracyclines |
| 34175 | Doxycycline 50mg capsules (A A H Pharmaceuticals Ltd) | tetracyclines |
| 48095 | Doxycycline 50mg capsules (Actavis UK Ltd) | tetracyclines |
| 53973 | Doxycycline 50mg capsules (Alliance Healthcare (Distribution) Ltd) | tetracyclines |
| 73070 | Doxycycline 50mg capsules (Almus Pharmaceuticals Ltd) | tetracyclines |
| 62008 | Doxycycline 50mg capsules (DE Pharmaceuticals) | tetracyclines |
| 61355 | Doxycycline 50mg capsules (Healthcare Pharma Ltd) | tetracyclines |
| 40391 | Doxycycline 50mg capsules (IVAX Pharmaceuticals UK Ltd) | tetracyclines |
| 63660 | Doxycycline 50mg capsules (Kent Pharmaceuticals Ltd) | tetracyclines |
| 34765 | Doxycycline 50mg capsules (Mylan) | tetracyclines |
| 65360 | Doxycycline 50mg capsules (Phoenix Healthcare Distribution Ltd) | tetracyclines |
| 73536 | Doxycycline 50mg capsules (Sigma Pharmaceuticals Plc) | tetracyclines |
| 32419 | Doxycycline 50mg capsules (Teva UK Ltd) | tetracyclines |
| 58326 | Doxycycline 50mg capsules (Waymade Healthcare Plc) | tetracyclines |
| 65189 | Doxycycline 50mg/5ml oral suspension | tetracyclines |
| 23405 | Doxylar 100mg capsules (Sandoz Ltd) | tetracyclines |
| 23432 | Doxylar 50mg capsules (Sandoz Ltd) | tetracyclines |
| 17226 | Economycin 250mg Capsule (DDSA Pharmaceuticals Ltd) | tetracyclines |
| 26111 | Economycin 250mg Tablet (DDSA Pharmaceuticals Ltd) | tetracyclines |
| 40980 | Efracea 40mg modified-release capsules (Galderma (UK) Ltd) | tetracyclines |
| 67782 | Generic Deteclo 300mg tablets | tetracyclines |
| 12541 | Imperacin 250mg Tablet (AstraZeneca UK Ltd) | tetracyclines |
| 7439 | Ledermycin 150mg Capsule (Wyeth Pharmaceuticals) | tetracyclines |
| 16613 | Ledermycin 150mg capsules (Mercury Pharma Group Ltd) | tetracyclines |
| 22076 | Ledermycin 300mg Tablet (Wyeth Pharmaceuticals) | tetracyclines |
| 30835 | LEDERMYCIN 75 MG SYR | tetracyclines |
| 20167 | LYMECYCLINE 204 MG CAP | tetracyclines |
| 453 | Lymecycline 408mg capsules | tetracyclines |
| 63806 | Lymecycline 408mg capsules (A A H Pharmaceuticals Ltd) | tetracyclines |
| 61202 | Lymecycline 408mg capsules (Teva UK Ltd) | tetracyclines |
| 19001 | Megaclor 170mg Capsule (Pharmax Ltd) | tetracyclines |
| 3413 | Minocin 100mg tablets (Wyeth Pharmaceuticals) | tetracyclines |
| 164 | Minocin 50mg tablets (Wyeth Pharmaceuticals) | tetracyclines |
| 1039 | Minocin MR 100mg capsules (Mylan) | tetracyclines |
| 9380 | Minocycline 100mg capsules | tetracyclines |
| 2578 | Minocycline 100mg modified-release capsules | tetracyclines |
| 34077 | Minocycline 100mg modified-release capsules (A A H Pharmaceuticals Ltd) | tetracyclines |
| 71056 | Minocycline 100mg modified-release capsules (Sigma Pharmaceuticals Plc) | tetracyclines |
| 46954 | Minocycline 100mg Tablet (Lagap) | tetracyclines |
| 1532 | Minocycline 100mg tablets | tetracyclines |
| 34926 | Minocycline 100mg tablets (A A H Pharmaceuticals Ltd) | tetracyclines |
| 40383 | Minocycline 100mg tablets (Actavis UK Ltd) | tetracyclines |
| 59039 | Minocycline 100mg tablets (Mylan) | tetracyclines |
| 59922 | Minocycline 100mg tablets (Teva UK Ltd) | tetracyclines |
| 67772 | Minocycline 100mg tablets (Tillomed Laboratories Ltd) | tetracyclines |
| 429 | Minocycline 50mg capsules | tetracyclines |
| 43700 | Minocycline 50mg Tablet (Lagap) | tetracyclines |
| 2999 | Minocycline 50mg tablets | tetracyclines |
| 79010 | Minocycline 50mg tablets (A A H Pharmaceuticals Ltd) | tetracyclines |
| 46947 | Minocycline 50mg tablets (Actavis UK Ltd) | tetracyclines |
| 71095 | Minocycline 50mg tablets (Phoenix Healthcare Distribution Ltd) | tetracyclines |
| 1013 | Mysteclin Capsule (Bristol-Myers Squibb Pharmaceuticals Ltd) | tetracyclines |
| 17222 | Mysteclin Oral solution (Bristol-Myers Squibb Pharmaceuticals Ltd) | tetracyclines |
| 1828 | Mysteclin Tablet (Bristol-Myers Squibb Pharmaceuticals Ltd) | tetracyclines |
| 15071 | Nordox 100mg Capsule (Sankyo Pharma UK Ltd) | tetracyclines |
| 8393 | NOVOBIOCIN/TETRACYCLINE 125 MG CAP | tetracyclines |
| 9361 | Oxymycin 250mg tablets (Dr Reddy's Laboratories (UK) Ltd) | tetracyclines |
| 2458 | OXYTETRACYCLINE 100 MG TAB | tetracyclines |
| 71393 | Oxytetracycline 125mg/5ml oral suspension | tetracyclines |
| 9034 | Oxytetracycline 125mg/5ml syrup | tetracyclines |
| 17209 | OXYTETRACYCLINE 250 MG INJ | tetracyclines |
| 8285 | OXYTETRACYCLINE 250 MG SYR | tetracyclines |
| 132 | Oxytetracycline 250mg capsules | tetracyclines |
| 34888 | Oxytetracycline 250mg Tablet (C P Pharmaceuticals Ltd) | tetracyclines |
| 77 | Oxytetracycline 250mg tablets | tetracyclines |
| 34044 | Oxytetracycline 250mg tablets (A A H Pharmaceuticals Ltd) | tetracyclines |
| 34040 | Oxytetracycline 250mg tablets (Actavis UK Ltd) | tetracyclines |
| 77008 | Oxytetracycline 250mg tablets (Almus Pharmaceuticals Ltd) | tetracyclines |
| 67752 | Oxytetracycline 250mg tablets (Dr Reddy's Laboratories (UK) Ltd) | tetracyclines |
| 71003 | Oxytetracycline 250mg tablets (Genesis Pharmaceuticals Ltd) | tetracyclines |
| 34336 | Oxytetracycline 250mg tablets (IVAX Pharmaceuticals UK Ltd) | tetracyclines |
| 40483 | Oxytetracycline 250mg tablets (Sandoz Ltd) | tetracyclines |
| 34141 | Oxytetracycline 250mg tablets (Teva UK Ltd) | tetracyclines |
| 67839 | Oxytetracycline 250mg/5ml oral suspension | tetracyclines |
| 17703 | Oxytetramix 250 tablets (Ashbourne Pharmaceuticals Ltd) | tetracyclines |
| 24097 | Rondomycin 150mg Capsule (Pfizer Ltd) | tetracyclines |
| 18109 | Sebomin MR 100mg capsules (Actavis UK Ltd) | tetracyclines |
| 37440 | Sebren MR 100mg capsules (Teva UK Ltd) | tetracyclines |
| 19693 | Sustamycin 250mg Capsule (Boehringer Mannheim UK Ltd) | tetracyclines |
| 7455 | Terramycin 250mg Capsule (Pfizer Ltd) | tetracyclines |
| 17467 | Terramycin 250mg tablets (Pfizer Ltd) | tetracyclines |
| 27195 | TERRAMYCIN SYR | tetracyclines |
| 9014 | Tetrabid-organon 250mg Capsule (Organon Laboratories Ltd) | tetracyclines |
| 8219 | Tetrachel 250mg Capsule (Berk Pharmaceuticals Ltd) | tetracyclines |
| 3816 | Tetrachel 250mg Tablet (Berk Pharmaceuticals Ltd) | tetracyclines |
| 25017 | TETRACYCLINE | tetracyclines |
| 56044 | Tetracycline 125mg/5ml oral solution | tetracyclines |
| 21804 | Tetracycline 125mg/5ml syrup | tetracyclines |
| 8284 | Tetracycline 125mg/5ml syrup | tetracyclines |
| 41547 | Tetracycline 250mg Capsule (Berk Pharmaceuticals Ltd) | tetracyclines |
| 121 | Tetracycline 250mg capsules | tetracyclines |
| 34011 | Tetracycline 250mg capsules | tetracyclines |
| 78552 | Tetracycline 250mg modified release capsules | tetracyclines |
| 56181 | Tetracycline 250mg Tablet (Celltech Pharma Europe Ltd) | tetracyclines |
| 45271 | Tetracycline 250mg Tablet (Numark Management Ltd) | tetracyclines |
| 386 | Tetracycline 250mg tablets | tetracyclines |
| 43538 | Tetracycline 250mg tablets (A A H Pharmaceuticals Ltd) | tetracyclines |
| 41636 | Tetracycline 250mg tablets (Actavis UK Ltd) | tetracyclines |
| 54214 | Tetracycline 250mg tablets (Alliance Healthcare (Distribution) Ltd) | tetracyclines |
| 60577 | Tetracycline 250mg tablets (Almus Pharmaceuticals Ltd) | tetracyclines |
| 53117 | Tetracycline 250mg tablets (Almus Pharmaceuticals Ltd) | tetracyclines |
| 63602 | Tetracycline 250mg tablets (Intrapharm Laboratories Ltd) | tetracyclines |
| 58076 | Tetracycline 250mg tablets (Kent Pharmaceuticals Ltd) | tetracyclines |
| 79117 | Tetracycline 250mg tablets (RX Farma) | tetracyclines |
| 48100 | Tetracycline 250mg tablets (Teva UK Ltd) | tetracyclines |
| 76159 | Tetracycline 250mg tablets (Waymade Healthcare Plc) | tetracyclines |
| 2922 | Tetracycline 250mg with nystatin 250000units tablets | tetracyclines |
| 21366 | Tetracycline 250mg/vial IV injection | tetracyclines |
| 2636 | TETRACYCLINE 500 MG CAP | tetracyclines |
| 3528 | TETRACYCLINE 500 MG TAB | tetracyclines |
| 25274 | Tetracycline 500mg/vial IV injection | tetracyclines |
| 31425 | TETRACYCLINE HCL/PANCREATIC CONCENTRATE CAP | tetracyclines |
| 28736 | TETRACYCLINE HYDROCHLORIDE/AMPHOTERICIN SYR | tetracyclines |
| 79062 | Tetracycline with amphoteracin syrup | tetracyclines |
| 15355 | Tetracycline with chlortetracycline & demeclocycline tablets | tetracyclines |
| 25071 | Tetracycline with nystatin capsules | tetracyclines |
| 24277 | TETRALYSAL 150 MG CAP | tetracyclines |
| 60452 | Tetralysal 300 capsules (DE Pharmaceuticals) | tetracyclines |
| 4951 | Tetralysal 300 capsules (Galderma (UK) Ltd) | tetracyclines |
| 20054 | Tetralysal 408mg Capsule (Pharmacia Ltd) | tetracyclines |
| 41763 | Tigecycline 50mg powder for solution for infusion vials | tetracyclines |
| 43896 | Tygacil 50mg powder for solution for infusion vials (Pfizer Ltd) | tetracyclines |
| 25016 | VIBRAMYCIN | tetracyclines |
| 268 | Vibramycin 100mg capsules (Pfizer Ltd) | tetracyclines |
| 3152 | Vibramycin 100mg Dispersible tablet (Pfizer Ltd) | tetracyclines |
| 2202 | Vibramycin 50 capsules (Pfizer Ltd) | tetracyclines |
| 10454 | Vibramycin 50mg/5ml Oral solution (Pfizer Ltd) | tetracyclines |
| 9267 | Vibramycin Acne Pack 50mg capsules (Pfizer Ltd) | tetracyclines |
| 56198 | Vibramycin-D 100mg dispersible tablets (Mawdsley-Brooks & Company Ltd) | tetracyclines |
| 14904 | Vibramycin-D 100mg dispersible tablets (Pfizer Ltd) | tetracyclines |
| 59542 | Vibramycin-D 100mg dispersible tablets (Sigma Pharmaceuticals Plc) | tetracyclines |
| 52967 | Vibramycin-D 100mg dispersible tablets (Stephar (U.K.) Ltd) | tetracyclines |
| 53135 | Vibramycin-D 100mg dispersible tablets (Waymade Healthcare Plc) | tetracyclines |
| 26392 | Vibrox 100mg capsules (Kent Pharmaceuticals Ltd) | tetracyclines |

Opioids

| Product code | gemscriptcode | Product name |
| --- | --- | --- |
| 36608 | 56226020 | Co-codamol Effervescent tablet (A A H Pharmaceuticals Ltd) |
| 40663 | 76604020 | Co-codamol 30mg/500mg effervescent tablets (Actavis UK Ltd) |
| 34264 | 66696020 | Co-codamol 30mg/500mg tablets (IVAX Pharmaceuticals UK Ltd) |
| 57353 | 16704021 | Co-codamol 30mg/500mg caplets (DE Pharmaceuticals) |
| 65904 | 52235021 | Co-codamol 8mg/500mg capsules (Sigma Pharmaceuticals Plc) |
| 34667 | 56463020 | Co-codamol 30mg/500mg tablets (A A H Pharmaceuticals Ltd) |
| 3029 | 6657007 | CO-CODAMOL EFF 30MG/500MG TAB |
| 66352 | 62276021 | Co-codamol 8mg/500mg caplets (Kent Pharmaceuticals Ltd) |
| 57865 | 17206021 | Co-codamol 30mg/500mg caplets (Actavis UK Ltd) |
| 37904 | 94749020 | Co-codamol 12.8mg/500mg tablets |
| 53702 | 72186020 | Co-codamol 30mg/500mg tablets (Actavis UK Ltd) |
| 34865 | 49324020 | Co-codamol 8mg+500mg Tablet (C P Pharmaceuticals Ltd) |
| 34815 | 63355020 | Co-codamol 8mg/500mg tablets (Kent Pharmaceuticals Ltd) |
| 59442 | 21373021 | Co-codamol 30mg/500mg caplets (Phoenix Healthcare Distribution Ltd) |
| 34495 | 59020020 | Co-codamol 8mg/500mg tablets (Mylan) |
| 43414 | 70680020 | Co-codamol 8mg/500mg effervescent tablets (Bayer Plc) |
| 51084 | 4340020 | Co-codamol 30mg/500mg capsules (AMCo) |
| 56006 | 4296020 | Co-codamol 8mg/500mg tablets (Bristol Laboratories Ltd) |
| 46633 | 74959020 | Co-codamol 8mg/500mg capsules (A A H Pharmaceuticals Ltd) |
| 33688 | 65467020 | Co-codamol 30mg/500mg effervescent tablets (Fannin UK Ltd) |
| 27785 | 49337020 | Co-codamol 30mg/500mg tablets (Zentiva) |
| 19 | 60971020 | Co-codamol 8mg/500mg tablets |
| 43244 | 70233020 | Co-codamol 30mg+500mg Effervescent tablet (Hillcross Pharmaceuticals Ltd) |
| 51819 | 4325020 | Co-codamol 8mg/500mg capsules (Phoenix Healthcare Distribution Ltd) |
| 2794 | 49325020 | Co-codamol 30mg/500mg tablets (Wockhardt UK Ltd) |
| 41276 | 76494020 | Co-codamol 8mg/500mg tablets (Almus Pharmaceuticals Ltd) |
| 36488 | 71799020 | Co-codamol 30mg/500mg capsules (Mylan) |
| 46729 | 469021 | Co-codamol 15mg/500mg capsules |
| 58855 | 47330020 | Co-codamol 30mg/500mg capsules (Waymade Healthcare Plc) |
| 29488 | 49335020 | Co-codamol 8mg/500mg effervescent tablets (Zentiva) |
| 625 | 60973020 | Co-codamol 8mg/500mg capsules |
| 30556 | 62877020 | Co-codamol 8mg/500mg effervescent tablets (Fannin UK Ltd) |
| 24209 | !5424102 | CO-CODAMOL EFFERVESCENT |
| 29342 | 63572020 | Co-codamol 8mg+500mg Tablet (M & A Pharmachem Ltd) |
| 47847 | 73826020 | Co-codamol 30mg/500mg capsules (Zentiva) |
| 57900 | 17205021 | Co-codamol 30mg/500mg caplets (A A H Pharmaceuticals Ltd) |
| 52085 | 4332020 | Co-codamol 30mg/500mg effervescent tablets (Zanza Laboratories Ltd) |
| 34840 | 67663020 | Co-codamol 30mg/500mg tablets (Almus Pharmaceuticals Ltd) |
| 43238 | 71802020 | Co-codamol 30mg/500mg effervescent tablets (Mylan) |
| 66538 | 4323020 | Co-codamol 8mg/500mg capsules (Alliance Healthcare (Distribution) Ltd) |
| 33643 | 60674020 | Co-codamol 8mg+500mg Tablet (Family Health) |
| 59479 | 4337020 | Co-codamol 30mg/500mg capsules (Actavis UK Ltd) |
| 56266 | 4311020 | Co-codamol 30mg/500mg tablets (Alliance Healthcare (Distribution) Ltd) |
| 70518 | 77572021 | Co-codamol 60mg/1000mg tablets |
| 46906 | 59611020 | Co-codamol 8mg+500mg Effervescent tablet (Numark Management Ltd) |
| 14602 | 84588020 | Co-codamol 60mg/1000mg effervescent powder sachets sugar free |
| 67106 | 66020021 | Co-codamol 30mg/500mg caplets (Wockhardt UK Ltd) |
| 56565 | 13497020 | Co-codamol 15mg/500mg tablets (A A H Pharmaceuticals Ltd) |
| 59986 | 21777021 | Co-codamol 30mg/500mg tablets (M & A Pharmachem Ltd) |
| 33653 | 49328020 | Co-codamol 8mg/500mg tablets (IVAX Pharmaceuticals UK Ltd) |
| 96 | 80055020 | Co-codamol 30mg/500mg tablets |
| 48775 | 41378020 | Co-codamol 30mg/500mg caplets (AMCo) |
| 42791 | 68530020 | Co-codamol 30mg/500mg tablets (Sandoz Ltd) |
| 41682 | 59950020 | Co-codamol 30mg+500mg Effervescent tablet (Roche Consumer Health) |
| 65440 | 71808020 | Co-codamol 30mg/500mg tablets (Mylan) |
| 46987 | 77582020 | Co-codamol 15mg+500mg Tablet (Hillcross Pharmaceuticals Ltd) |
| 7072 | 78380020 | Co-codamol 15mg/500mg tablets |
| 810 | 80054020 | Co-codamol 30mg/500mg effervescent tablets |
| 34968 | 55234020 | Co-codamol 8mg/500mg tablets (Teva UK Ltd) |
| 66904 | 61352021 | Co-codamol 8mg/500mg caplets (Wockhardt UK Ltd) |
| 36993 | 71989020 | Co-codamol 30mg/500mg capsules (Teva UK Ltd) |
| 56340 | 15644021 | Co-codamol 30mg/500mg caplets (J M McGill Ltd) |
| 41275 | 66420020 | Co-codamol 8mg+500mg Tablet (Nucare Plc) |
| 57839 | 14608021 | Co-codamol 15mg/500mg tablets (Waymade Healthcare Plc) |
| 58501 | 17422021 | Co-codamol 30mg/500mg caplets (AM Distributions (Yorkshire) Ltd) |
| 34518 | 49331020 | Co-codamol 8mg/500mg tablets (A A H Pharmaceuticals Ltd) |
| 62169 | 65245020 | Co-codamol 8mg/500mg tablets (Vantage) |
| 58636 | 69515020 | Co-codamol 8mg/500mg effervescent tablets (Teva UK Ltd) |
| 57 | 60972020 | Co-codamol 8mg/500mg effervescent tablets |
| 64387 | 16705021 | Co-codamol 30mg/500mg tablets (DE Pharmaceuticals) |
| 27784 | 53764020 | Co-codamol 8mg/500mg tablets (Actavis UK Ltd) |
| 57097 | 4335020 | Co-codamol 30mg/500mg capsules (Alliance Healthcare (Distribution) Ltd) |
| 39340 | 74574020 | Co-codamol 8mg/500mg caplets (Vantage) |
| 58288 | 16709021 | Co-codamol 30mg/500mg capsules (DE Pharmaceuticals) |
| 59705 | 16707021 | Co-codamol 8mg/500mg capsules (DE Pharmaceuticals) |
| 800 | 80053020 | Co-codamol 30mg/500mg capsules |
| 48311 | 4312020 | Co-codamol 30mg/500mg caplets (Kent Pharmaceuticals Ltd) |
| 34845 | 66914020 | Co-codamol 30mg/500mg effervescent tablets (Zentiva) |
| 42213 | 75096020 | Co-codamol 8mg/500mg capsules (Actavis UK Ltd) |
| 65092 | 53444021 | Co-codamol 30mg/500mg caplets (Mawdsley-Brooks & Company Ltd) |
| 63900 | 16703021 | Co-codamol 8mg/500mg tablets (DE Pharmaceuticals) |
| 57929 | 47329020 | Co-codamol 8mg/500mg capsules (Waymade Healthcare Plc) |
| 53679 | 4328020 | Co-codamol 30mg/500mg effervescent tablets (A A H Pharmaceuticals Ltd) |
| 34229 | 56031020 | Co-codamol 8mg+500mg Dispersible tablet (Rhone-Poulenc Rorer Ltd) |
| 34257 | 59021020 | Co-codamol 8mg/500mg effervescent tablets (Mylan) |
| 64545 | 68823020 | Co-codamol 8mg/500mg tablets (Aspar Pharmaceuticals Ltd) |
| 40662 | 76720020 | Co-codamol 8mg/500mg effervescent tablets (Actavis UK Ltd) |
| 41259 | 76276020 | Co-codamol 30mg/500mg effervescent tablets (Teva UK Ltd) |
| 64726 | 52898021 | Co-codamol 15mg/500mg tablets (Galen Ltd) |
| 66553 | 40547020 | Co-codamol 30mg/500mg effervescent tablets (AMCo) |
| 17808 | !8503906 | CO-CODAMOL 30MG/500MG |
| 61647 | 38870020 | Co-codamol 15mg/500mg tablets (Alliance Healthcare (Distribution) Ltd) |
| 40385 | 69578020 | Co-codamol 8mg/500mg effervescent tablets (Almus Pharmaceuticals Ltd) |
| 68252 | 43837020 | Co-codamol 30mg/500mg tablets (Bristol Laboratories Ltd) |
| 34784 | 59364020 | Co-codamol 8mg/500mg effervescent tablets (Sandoz Ltd) |
| 67753 | 78630020 | Co-codamol 8mg/500mg tablets (Wockhardt UK Ltd) |
| 65806 | 4289020 | Co-codamol 8mg/500mg tablets (Alliance Healthcare (Distribution) Ltd) |
| 22817 | !8504309 | CO-CODAMOL SF EFF POWDER |
| 34497 | 49320020 | Co-codamol 8mg+500mg Tablet (Berk Pharmaceuticals Ltd) |
| 53287 | 4326020 | Co-codamol 30mg/500mg effervescent tablets (Alliance Healthcare (Distribution) Ltd) |
| 57465 | 16702021 | Co-codamol 8mg/500mg caplets (DE Pharmaceuticals) |
| 59131 | 4324020 | Co-codamol 8mg/500mg capsules (Bayer Plc) |
| 1261 | 84587020 | Co-codamol 30mg/500mg effervescent powder sachets sugar free |
| 31577 | 56464020 | Co-codamol 30mg/500mg capsules (A A H Pharmaceuticals Ltd) |
| 44924 | 71985020 | Co-codamol 30mg/500mg tablets (Teva UK Ltd) |
| 63551 | 4321020 | Co-codamol 8mg/500mg effervescent tablets (Vantage) |
| 70497 | 77474021 | Co-codamol 15mg/500mg capsules (A A H Pharmaceuticals Ltd) |
| 60517 | 75431020 | Co-codamol 30mg/500mg effervescent tablets (Almus Pharmaceuticals Ltd) |
| 46511 | 243021 | Co-codamol 15mg/500mg effervescent tablets sugar free |
| 55465 | 4339020 | Co-codamol 30mg/500mg capsules (Sigma Pharmaceuticals Plc) |
| 56549 | 14519021 | Co-codamol 8mg/500mg effervescent tablets (Waymade Healthcare Plc) |
| 65314 | 57312021 | Co-codamol 8mg/500mg caplets (Actavis UK Ltd) |
| 33961 | 71396020 | Co-codamol 30mg/500mg capsules (IVAX Pharmaceuticals UK Ltd) |
| 32692 | 56462020 | Co-codamol 8mg/500mg effervescent tablets (A A H Pharmaceuticals Ltd) |
| 33679 | 49336020 | Co-codamol 8mg/500mg tablets (Zentiva) |
| 56461 | 4310020 | Co-codamol 30mg/500mg tablets (Kent Pharmaceuticals Ltd) |
| 56171 | 4334020 | Co-codamol 30mg/500mg capsules (Kent Pharmaceuticals Ltd) |
| 55044 | 47328020 | Co-codamol 30mg/500mg caplets (Waymade Healthcare Plc) |
| 69304 | 73914021 | Co-codamol 15mg/500mg tablets (Actavis UK Ltd) |
| 52856 | 29961020 | Co-codaprin 8mg/400mg tablets |
| 2986 | 55579020 | Co-codaprin 8mg/400mg dispersible tablets |
| 42218 | 74305020 | Co-codaprin 8mg/400mg dispersible tablets (A A H Pharmaceuticals Ltd) |
| 63658 | 53770020 | Co-codaprin 400/8 Tablet (Hillcross Pharmaceuticals Ltd) |
| 2047 | 55578020 | Co-codaprin 8mg with 400mg tablets |
| 23496 | !0375101 | CO-CODAPRIN DISPERSIBLE |
| 46925 | 53767020 | Co-codaprin 8mg/400mg dispersible tablets (Actavis UK Ltd) |
| 33340 | 70228020 | Co-dydramol 10mg+500mg/5ml Liquid (Rosemont Pharmaceuticals Ltd) |
| 62635 | 4302020 | Co-dydramol 10mg/500mg tablets (Alliance Healthcare (Distribution) Ltd) |
| 47071 | 547021 | Co-dydramol (dihydrocodeine and paracetamol) 10mg with 500mg/5ml oral suspension |
| 7063 | 89000020 | Co-dydramol 10mg/500mg/5ml oral suspension |
| 28780 | 49353020 | Co-dydramol 10mg/500mg tablets (A A H Pharmaceuticals Ltd) |
| 67779 | 76249020 | Co-dydramol 10mg/500mg tablets (Wockhardt UK Ltd) |
| 61372 | 4300020 | Co-dydramol 10mg/500mg tablets (Kent Pharmaceuticals Ltd) |
| 57197 | 14517021 | Co-dydramol 10mg/500mg tablets (Waymade Healthcare Plc) |
| 21927 | !1887101 | CO-DYDRAMOL |
| 36019 | 63577020 | Co-dydramol 10mg+500mg Tablet (M & A Pharmachem Ltd) |
| 32926 | 49347020 | Co-dydramol 10mg/500mg tablets (Actavis UK Ltd) |
| 30444 | 59024020 | Co-dydramol 10mg/500mg tablets (Mylan) |
| 11 | 62553020 | Co-dydramol 10mg/500mg tablets |
| 30165 | 62554020 | Co-dydramol (dihydrocodeine and paracetamol) 7.46mg with 500mg tablets |
| 61698 | 91554020 | Co-dydramol 10mg/500mg/5ml oral solution |
| 53079 | 4304020 | Co-dydramol 10mg/500mg tablets (Sigma Pharmaceuticals Plc) |
| 34920 | 49340020 | Co-dydramol 10mg+500mg Tablet (Berk Pharmaceuticals Ltd) |
| 71006 | 36797020 | Co-dydramol 10mg/500mg oral powder sachets (Special Order) |
| 34939 | 55837020 | Co-dydramol 10mg+500mg Tablet (Duncan Flockhart Ltd) |
| 65035 | 71382020 | Co-dydramol 10mg/500mg tablets (Almus Pharmaceuticals Ltd) |
| 71523 | 78735021 | Co-dydramol 20mg/500mg tablets (M & A Pharmachem Ltd) |
| 15198 | 49344020 | Co-dydramol 10mg+500mg Tablet (C P Pharmaceuticals Ltd) |
| 37291 | 60085020 | Co-dydramol 10mg/500mg tablets (Ranbaxy (UK) Ltd) |
| 69006 | 29394020 | Co-dydramol 10mg/500mg/5ml oral suspension sugar free |
| 41278 | 57801020 | Co-dydramol 10mg/500mg tablets (IVAX Pharmaceuticals UK Ltd) |
| 55530 | 4305020 | Co-dydramol 10mg/500mg tablets (Phoenix Healthcare Distribution Ltd) |
| 40422 | 49361020 | Co-dydramol 10mg/500mg tablets (Zentiva) |
| 64074 | 21372021 | Co-dydramol 10mg/500mg tablets (DE Pharmaceuticals) |
| 38430 | 58864020 | Co-dydramol 10mg+500mg Tablet (Merck Generics (UK) Ltd) |
| 43441 | 49350020 | Co-dydramol 10mg+500mg Tablet (Celltech Pharma Europe Ltd) |
| 34737 | 54661020 | Co-dydramol 10mg/500mg tablets (Teva UK Ltd) |
| 45231 | 62832020 | Co-proxamol 32.5mg+325mg Tablet (Regent Laboratories Ltd) |
| 34022 | 55237020 | Co-proxamol 32.5mg/325mg tablets (Teva UK Ltd) |
| 41407 | 60093020 | Co-proxamol 32.5mg/325mg tablets (Ranbaxy (UK) Ltd) |
| 34554 | 49368020 | Co-proxamol 32.5/325 Tablet (C P Pharmaceuticals Ltd) |
| 67755 | 69582020 | Co-proxamol 32.5mg/325mg tablets (Almus Pharmaceuticals Ltd) |
| 33995 | 49365020 | Co-proxamol 32.5/325 Tablet (Berk Pharmaceuticals Ltd) |
| 483 | 58898020 | Co-proxamol (dextropropoxyphene and paracetamol) 32.5mg with 325mg/5ml oral suspension sugar free |
| 34597 | 62331020 | Co-proxamol 32.5mg/325mg tablets (Kent Pharmaceuticals Ltd) |
| 53208 | 4308020 | Co-proxamol 32.5mg/325mg tablets (Lexon (UK) Ltd) |
| 34546 | 62885020 | Co-proxamol 32.5mg+325mg Tablet (Neo Laboratories Ltd) |
| 43536 | 55415020 | Co-proxamol 32.5/325 Tablet (Numark Management Ltd) |
| 28253 | 55651020 | Co-proxamol 32.5/325 Oral suspension (Rosemont Pharmaceuticals Ltd) |
| 69043 | 53442021 | Co-proxamol 32.5mg/325mg tablets (DE Pharmaceuticals) |
| 30954 | 51142020 | Co-proxamol 32.5mg/325mg tablets (Actavis UK Ltd) |
| 30966 | 51145020 | Co-proxamol 32.5/325 Tablet (Dista Products Ltd) |
| 69534 | 4306020 | Co-proxamol 32.5mg/325mg tablets (Alliance Healthcare (Distribution) Ltd) |
| 34349 | 57798020 | Co-proxamol 32.5mg/325mg tablets (IVAX Pharmaceuticals UK Ltd) |
| 45276 | 62237020 | Co-proxamol 32.5mg+325mg Tablet (Sigma Pharmaceuticals Plc) |
| 55245 | 4309020 | Co-proxamol 32.5mg/325mg tablets (Phoenix Healthcare Distribution Ltd) |
| 4 | 58897020 | Co-proxamol 32.5mg/325mg tablets |
| 34468 | 59367020 | Co-proxamol 32.5mg/325mg tablets (Sandoz Ltd) |
| 33647 | 49376020 | Co-proxamol 32.5mg/325mg tablets (A A H Pharmaceuticals Ltd) |
| 43891 | 92555020 | Co-proxamol 32.5mg/325mg/5ml oral suspension |
| 19550 | !1806301 | CO-PROXAMOL |
| 34319 | 49379020 | Co-proxamol 32.5mg/325mg tablets (Zentiva) |
| 34397 | 63582020 | Co-proxamol 32.5mg+325mg Tablet (M & A Pharmachem Ltd) |
| 4607 | 58599020 | Co-proxamol 32.5/325 Tablet (Dista Products Ltd) |
| 68989 | 71050021 | Buprenorphine 2mg oral lyophilisates sugar free |
| 6547 | 52971020 | Buprenorphine 2mg sublingual tablets sugar free |
| 40212 | 76007020 | Buprenorphine 8mg sublingual tablets sugar free (Teva UK Ltd) |
| 68988 | 71052021 | Buprenorphine 8mg oral lyophilisates sugar free |
| 64155 | 75999020 | Buprenorphine 400microgram sublingual tablets sugar free (Teva UK Ltd) |
| 6917 | 78619020 | Buprenorphine 52.5micrograms/hour transdermal patches |
| 6056 | 52972020 | Buprenorphine 8mg sublingual tablets sugar free |
| 63788 | 44850021 | Buprenorphine 1mg sublingual tablets sugar free |
| 62675 | 39813020 | Buprenorphine 200microgram sublingual tablets sugar free (A A H Pharmaceuticals Ltd) |
| 40211 | 76002020 | Buprenorphine 2mg sublingual tablets sugar free (Teva UK Ltd) |
| 62776 | 44852021 | Buprenorphine 4mg sublingual tablets sugar free |
| 7236 | 89596020 | Buprenorphine 10micrograms/hour transdermal patches |
| 67018 | 44279021 | Buprenorphine 35micrograms/hour transdermal patches (A A H Pharmaceuticals Ltd) |
| 62874 | 44854021 | Buprenorphine 6mg sublingual tablets sugar free |
| 66463 | 62207021 | Buprenorphine 15micrograms/hour transdermal patches |
| 69942 | 4372020 | Buprenorphine 400microgram sublingual tablets sugar free (Phoenix Healthcare Distribution Ltd) |
| 35681 | 92283020 | Buprenorphine 2mg / Naloxone 500microgram sublingual tablets sugar free |
| 35682 | 92291020 | Buprenorphine 8mg / Naloxone 2mg sublingual tablets sugar free |
| 7334 | 89594020 | Buprenorphine 5micrograms/hour transdermal patches |
| 40473 | 91997020 | Buprenorphine 300micrograms/1ml solution for injection ampoules |
| 396 | 64275020 | Buprenorphine 200microgram sublingual tablets sugar free |
| 11584 | 79157020 | Buprenorphine 70micrograms/hour transdermal patches |
| 320 | 64276020 | Buprenorphine HCl 300micrograms injection |
| 7238 | 89598020 | Buprenorphine 20micrograms/hour transdermal patches |
| 70283 | 69934021 | Buprenorphine 16mg / Naloxone 4mg sublingual tablets sugar free |
| 58273 | 76089020 | Buprenorphine 2mg sublingual tablets sugar free (A A H Pharmaceuticals Ltd) |
| 3064 | 64277020 | Buprenorphine 400microgram sublingual tablets sugar free |
| 62969 | 42183020 | Buprenorphine 8mg sublingual tablets sugar free (Zentiva) |
| 6879 | 83959020 | Buprenorphine 35micrograms/hour transdermal patches |
| 59970 | 4381020 | Buprenorphine 2mg sublingual tablets sugar free (Actavis UK Ltd) |
| 65157 | 4379020 | Buprenorphine 2mg sublingual tablets sugar free (Sigma Pharmaceuticals Plc) |
| 20077 | 71757020 | Paracetamol 400mg with codeine phosphate 10mg with diphenhydramine 5mg & caffeine 50mg tablet |
| 14912 | 81048020 | Codeine phosphate 8mg with Paracetamol 500mg capsules |
| 4805 | 59907020 | Codeine phosphate 15mg/5ml diabetic oral solution |
| 7542 | 61567020 | Codeine phosphate 8mg with paracetamol 500mg tablets |
| 24996 | 71309020 | Codeine phosphate 8mg with paracetamol 500mg with buclizine 6.25mg tablets |
| 8246 | 61568020 | Codeine phosphate 8mg with paracetamol 500mg effervescent tablets |
| 47081 | 98453020 | Paracetamol 500mg with codeine phosphate 12.8mg & caffeine 30mg effervescent tablet |
| 38363 | 94751020 | Codeine phosphate 12.8mg with paracetamol 500mg tablets |
| 68509 | 50959020 | Codeine phosphate 30mg Tablet (Celltech Pharma Europe Ltd) |
| 28606 | 261007 | ASPIRIN/CAFFEINE/CODEINE PHOSPHATE 300 MG TAB |
| 63600 | 53190020 | Codeine phosphate powder (Martindale Pharmaceuticals Ltd) |
| 38088 | 68500020 | Paracetamol 500mg with codeine phosphate 30 mg tablet |
| 8500 | 2810007 | CODEINE PHOSPHATE/PARACETAMOL/SODIUM CIT 8 MG CAP |
| 7770 | 262007 | ASPIRIN/CODEINE PHOSPHATE/PARACETAMOL 250 MG TAB |
| 306 | 69988020 | Aspirin with codeine phosphate and caffeine tablets |
| 14785 | 65987020 | Paracetamol500mg with codeine phosphate 15mg tablet |
| 19622 | 70130020 | Pseudoephedrine with brompheniramine & codeine phosphate paediatric oral solution |
| 18221 | 84466020 | Codeine phosphate 15mg with paracetamol 500mg tablets |
| 43504 | 49488020 | Codeine phosphate Oral solution (William Ransom) |
| 23420 | 53857020 | Codeine phosphate 60mg with paracetamol 1000mg effervescent powder sugar free |
| 47919 | 76335020 | Codeine phosphate 15mg Tablet (Wockhardt UK Ltd) |
| 9462 | 65988020 | Paracetamol 500mg with codeine phosphate 8mg effervescent tablet |
| 21693 | 70852020 | Codeine phosphate with dicycloverine with salts liquid |
| 10099 | 71315020 | Paracetamol 500mg with codeine phosphate 8mg & buclizine 6.25mg |
| 8053 | 3006007 | BROMPHENIRAMINE /CODEINE PHOSPHATE / 2 MG ELI |
| 23580 | 70266020 | Codeine phosphate with diphenhydramine with paracetamol with phenylephrine with caffeine with vitamin c tablets |
| 11325 | 77473020 | Paracetamol 500mg with codeine phosphate 30 mg tablet |
| 38085 | 81312020 | Paracetamol 500mg with codeine phosphate 10mg capsule |
| 2917 | 65993020 | Paracetamol 500mg with codeine phosphate 30 mg tablet |
| 20565 | 73512020 | Paracetamol 500mg with codeine phosphate 8mg & caffeine 30mg capsule |
| 15831 | 53858020 | Codeine phosphate 30mg with paracetamol 500mg effervescent powder sugar free |
| 9457 | 65992020 | Paracetamol 500mg with codeine phosphate 8mg tablet |
| 20127 | 77403020 | Codeine phosphate 8mg with aspirin 400mg with caffeine dispersible tablets |
| 11009 | 66001020 | Paracetamol 500mg with codeine phosphate 8mg & caffeine 30mg tablet |
| 21104 | 6402007 | PARACETAMOL/CODEINE PHOSPHATE 500 MG TAB |
| 2846 | 65994020 | Paracetamol 500mg with codeine phosphate 30mg effervescent tablet |
| 3185 | 68499020 | Paracetamol 500mg with codeine phosphate 30mg capsule |
| 13992 | 70129020 | Pseudoephedrine with brompheniramine & codeine phosphate oral solution |
| 14676 | 77683020 | Paracetamol 500mg with codeine phosphate 8mg & caffeine 30mg effervescent tablet |
| 25485 | 86018020 | Codeine phosphate with other ingredient pastilles |
| 16818 | 79777020 | Paracetamol 500mg with codeine phosphate 30mg effervescent powder sugar free |
| 6886 | 61569020 | Codeine phosphate 30mg with paracetamol 500mg tablets |
| 53999 | 77532020 | Codeine phosphate 60mg Tablet (Wockhardt UK Ltd) |
| 25529 | 77680020 | Paracetamol 500mg with codeine phosphate 8mg & caffeine 30mg tablet |
| 29828 | 218007 | ACETYLSALICYLIC ACID / CODEINE PHOSPHATE MG TAB |
| 25514 | 81311020 | Paracetamol 500mg with codeine phosphate 10mg tablet |
| 21703 | 79776020 | Paracetamol 1000mg with codeine phosphate 60mg effervescent powder sugar free |
| 16467 | 84870020 | Codeine phosphate 30mg with paracetamol 500mg effervescent tablets |
| 37348 | 71809020 | Codeine phosphate with paracetamol, doxylamine and caffeine 10mg with 450mg with 5mg with 30mg tablets |
| 24498 | 100007 | ASPIRIN/CODEINE PHOSPHATE/PARACETAMOL 300 MG TAB |
| 28784 | 82882020 | Codeine phosphate 8mg with aspirin 400mg tablets |
| 4487 | 2985007 | CAFFEINE CITRATE/CODEINE PHOSPHATE/PARAC 15 MG TAB |
| 21673 | 66002020 | Paracetamol 500mg with codeine phosphate 8mg & caffeine 30mg effervescent tablet |
| 31871 | 81313020 | Paracetamol 450mg with codeine phosphate 8.1mg tablet |
| 66807 | 72731020 | Paracetamol with codeine phosphate peadiatric elixir |
| 15360 | 52685020 | Codeine phosphate powder |
| 22627 | 5141007 | CODEINE PHOSPHATE 15/PARACETAMOL 500MG TAB |
| 24517 | 3834007 | CODEINE PHOSPHATE / MAGNESIUM CHLORIDE MG MIX |
| 2988 | 6690007 | CODEINE PHOSPHATE 15 MG ELI |
| 15937 | 70220020 | Diphenhydramine with codeine phosphate, sodium citrate and menthol 14mg with 5.7mg with 57mg with 1.1mg/5ml oral solution |
| 48066 | 92469020 | Codeine phosphate oral solution |
| 20256 | 71760020 | Paracetamol 450mg with codeine phosphate 10mg with doxylamine 5mg & caffeine 30mg tablet |
| 47952 | 76338020 | Codeine phosphate 30mg Tablet (Wockhardt UK Ltd) |
| 30021 | 68501020 | Paracetamol 500mg with codeine phosphate 13.5mg tablet |
| 9460 | 65989020 | Paracetamol 500mg with codeine phosphate 8mg capsule |
| 4671 | 68504020 | Codeine phosphate 30mg with Paracetamol 500mg capsules |
| 50421 | 53776020 | Codeine phosphate 15mg Tablet (Celltech Pharma Europe Ltd) |
| 32510 | !1417101 | CODEINE LINCTUS |
| 22450 | 3049007 | ASPIRIN & CODEINE 75 MG TAB |
| 19724 | 3888007 | ASPIRIN & CODEINE paed 75 MG TAB |
| 9202 | 6692007 | CODEINE PHOS/EPHEDRINE HYD/PROMETHAZINE LIN |
| 36846 | 81680020 | Guaiacol 75mg/5ml / Codeine 7mg/5ml oral solution |
| 33495 | 3002007 | BUCLIZINE HYD/CODEINE PHOS/PARACET / COD 6.25 MG TAB |
| 2250 | 6150007 | CODEINE & PARACETAMOL TAB |
| 24304 | 3832007 | CODEINE PHOS/GUAIPHENESIN/PSEUDOEPHEDRIN 3 MG ELI |
| 203 | 2818007 | CODEINE CO SOLUBLE TAB |
| 31498 | 6728007 | ASPIRIN / CAFFEINE CIT./ CODEINE PHOS./ 200 MG TAB |
| 13598 | 3414007 | ASPIRIN & CODEINE 500 MG TAB |
| 7696 | 6703007 | PARACETAMOL 500MG/CODEINE 10MG MG TAB |
| 8732 | 3877007 | BUCLIZINE HYD/CODEINE PHOS/DOCUSATE SOD/ 6.25 MG TAB |
| 24859 | 2811007 | CODEINE 8MG & PARACETAMOL 500MG SUP |
| 37298 | 2134007 | PARACETAMOL,CAFFEINE,CODEINE & NICOTINAM TAB |
| 27353 | 3871007 | CAFFEINE /CODEINE PHOS./DOXYLAMINE SUCCI 30 MG TAB |
| 27598 | 2983007 | CAFFEINE/CODEINE PHOS./DIPHENHYDRAMINE H 50 MG TAB |
| 142 | 4100007 | CODEINE SOLUBLE TAB |
| 7976 | 3267007 | PARACETAMOL 450MG/CODEINE 8.1MG TAB |
| 4349 | 4615007 | PARACETAMOL & CODEINE TAB |
| 2698 | 6691007 | CODEINE & PARACETAMOL 8 MG TAB |
| 241 | 4104007 | CODEINE CO TAB |
| 10519 | 4147007 | CODEINE PHOS/IBUPROFEN SR (20MG/300MG) TAB |
| 8835 | 3833007 | CODEINE PHOS /DIPHENHYDRAMINE HYD. /MENT 5.7 MG ELI |
| 17707 | 6689007 | CODEINE PHOS/GUAIPHENESIN/PSEUDOEPHEDRIN 7.5 MG ELI |
| 17734 | 62393020 | Dextromoramide 5mg/ml injection |
| 28805 | 62401020 | Dextromoramide 10mg suppository |
| 3990 | 62391020 | Dextromoramide 5mg tablets |
| 7998 | 62392020 | Dextromoramide 10mg tablets |
| 12076 | 72612020 | Dextropropoxyphene 60mg capsules |
| 1762 | 64851020 | Dextropropoxyphene HCl with paracetamol 32.5mg with 325mg tablets |
| 18482 | 66005020 | Paracetamol 325mg with dextropropoxyphene 32.5mg tablet |
| 3714 | 6705007 | DEXTROPROPOXYPHENE NAPSYLATE/PARACETAMOL MG TAB |
| 24733 | 3816007 | DEXTROPROPOXYPHENE HCl S/R 150 MG CAP |
| 25959 | 95007 | ASPIRIN/CAFFEINE/DEXTROPROPOXYPHENE NAPS PUL |
| 19069 | 62404020 | Dextropropoxyphene 60mg capsules |
| 44311 | 6730007 | DEXTROPROPOXYPHENE NAPSYLATE/ASPIRIN 100 MG TAB |
| 48912 | 36808020 | Diamorphine 30mg powder for solution for injection vials |
| 29500 | 73145020 | Diamorphine 5mg/5ml oral solution |
| 47671 | 54713020 | Diamorphine hydrochloride 10mg Injection (Approved Prescription Services Ltd) |
| 6459 | 59888020 | Diamorphine hydrochloride 100mg powder for injection solution |
| 19351 | 3807007 | DIAMORPHINE 3 GM INJ |
| 65372 | 77243020 | Diamorphine 5mg powder for solution for injection ampoules (A A H Pharmaceuticals Ltd) |
| 18792 | 55619020 | Diamorphine 10mg tablets |
| 48953 | 36804020 | Diamorphine 100mg powder for solution for injection vials |
| 10473 | 2728007 | DIAMORPHINE 10 MG LIN |
| 34489 | 49497020 | Diamorphine hydrochloride 10mg Injection (Hillcross Pharmaceuticals Ltd) |
| 48483 | 4428020 | Diamorphine 30mg powder for solution for injection ampoules |
| 58499 | 29879020 | Diamorphine 3mg/5ml oral solution |
| 48413 | 36812020 | Diamorphine 5mg powder for solution for injection vials |
| 31960 | 73147020 | Diamorphine 15mg/5ml oral solution |
| 7849 | 70494020 | Diamorphine 100mg Injection (Manufacturer unknown) |
| 18965 | 3806007 | DIAMORPHINE 2.5 GM INJ |
| 20752 | 2726007 | DIAMORPHINE 15 MG INJ |
| 5079 | 59881020 | Diamorphine hydrochloride 5mg powder for injection solution |
| 58279 | 4420020 | Diamorphine 10mg tablets (A A H Pharmaceuticals Ltd) |
| 55724 | 36810020 | Diamorphine 500mg powder for solution for injection vials |
| 26407 | 3363007 | DIAMORPHINE 1 GM SUP |
| 67796 | 77246020 | Diamorphine 10mg powder for solution for injection ampoules (A A H Pharmaceuticals Ltd) |
| 47672 | 49498020 | Diamorphine hydrochloride 30mg Injection (Hillcross Pharmaceuticals Ltd) |
| 32897 | 49451020 | Diamorphine 5mg powder for solution for injection ampoules (Novartis Vaccines and Diagnostics Ltd) |
| 32687 | 6687007 | DIAMORPHINE & TERPOIN LIN |
| 5668 | 59883020 | Diamorphine hydrochloride 30mg powder for injection solution |
| 8735 | 73131020 | Diamorphine 5mg/5ml Oral solution (Manufacturer unknown) |
| 13364 | 2727007 | DIAMORPHINE 20 MG ELI |
| 48259 | 4421020 | Diamorphine 5mg powder for solution for injection ampoules |
| 28396 | 2721007 | DIAMORPHINE 100 MG SUP |
| 9945 | 57278020 | Diamorphine 10mg Tablet (Aurum Pharmaceuticals Ltd) |
| 25649 | 2724007 | DIAMORPHINE 60 MG SUP |
| 23785 | 2720007 | DIAMORPHINE 10 MG SUP |
| 15793 | 52700020 | Diamorphine hydrochloride powder |
| 7999 | 70489020 | Diamorphine 5mg Injection (Manufacturer unknown) |
| 8460 | 2713007 | DIAMORPHINE 1.5 GM INJ |
| 5670 | 59882020 | Diamorphine hydrochloride 10mg powder for injection solution |
| 34786 | 49452020 | Diamorphine 10mg powder for solution for injection ampoules (Novartis Vaccines and Diagnostics Ltd) |
| 7114 | 70367020 | Diamorphine 3mg/5ml oral solution |
| 18977 | 3808007 | DIAMORPHINE HYDROCHLORIDE 20 MG INJ |
| 6458 | 59889020 | Diamorphine hydrochloride 500mg powder for injection solution |
| 8866 | 70488020 | Diamorphine 10mg Tablet (Manufacturer unknown) |
| 41722 | 57334020 | Diamorphine 30mg powder for solution for injection ampoules (Wockhardt UK Ltd) |
| 30761 | 73132020 | Diamorphine 10mg/5ml Oral solution (Manufacturer unknown) |
| 25830 | 2722007 | DIAMORPHINE 20 MG SUP |
| 48880 | 36806020 | Diamorphine 10mg powder for solution for injection vials |
| 28711 | 70364020 | Diamorphine hydrochloride bpc 1973 3mg/5ml oral solution |
| 29592 | 2725007 | DIAMORPHINE 40 MG SUP |
| 27352 | 3805007 | DIAMORPHINE 64 MG INJ |
| 53417 | 4427020 | Diamorphine 10mg powder for solution for injection ampoules (Actavis UK Ltd) |
| 17163 | 2712007 | DIAMORPHINE 1 MG INJ |
| 42913 | 57333020 | Diamorphine 10mg powder for solution for injection ampoules (Wockhardt UK Ltd) |
| 34787 | 49453020 | Diamorphine 30mg powder for solution for injection ampoules (Novartis Vaccines and Diagnostics Ltd) |
| 8823 | 2715007 | DIAMORPHINE HYDROCHLORIDE 60 MG INJ |
| 9126 | 2714007 | DIAMORPHINE 4 GM INJ |
| 29591 | 3809007 | DIAMORPHINE 25 MG SUP |
| 23778 | 2711007 | DIAMORPHINE 5 MG SUP |
| 53181 | 76866020 | Diamorphine 10mg powder for solution for injection vials (Teva UK Ltd) |
| 48158 | 76863020 | Diamorphine 5mg powder for solution for injection vials (Teva UK Ltd) |
| 9053 | 70495020 | Diamorphine 500mg Injection (Manufacturer unknown) |
| 20713 | 3810007 | DIAMORPHINE 30 MG SUP |
| 48913 | 4433020 | Diamorphine 100mg powder for solution for injection ampoules |
| 13420 | 73133020 | Diamorphine 15mg/5ml Oral solution (Manufacturer unknown) |
| 8040 | 70493020 | Diamorphine 30mg Injection (Manufacturer unknown) |
| 24840 | 3811007 | DIAMORPHINE 75 MG SUP |
| 55221 | 4436020 | Diamorphine 500mg powder for solution for injection ampoules |
| 3165 | 70490020 | Diamorphine 10mg Injection (Manufacturer unknown) |
| 15339 | 73146020 | Diamorphine 10mg/5ml oral solution |
| 31033 | 73149020 | Diamorphine hydrochloride 3mg/5ml oral solution |
| 60721 | 4423020 | Diamorphine 5mg powder for solution for injection ampoules (Actavis UK Ltd) |
| 48434 | 4424020 | Diamorphine 10mg powder for solution for injection ampoules |
| 66654 | 94623020 | Diamorphine 15mg powder for solution for injection ampoules |
| 21868 | 72609020 | Diamorphine hydrochloride and cocaine oral solution |
| 15514 | 3364007 | DIAMORPHINE 2.5 MG LIN |
| 24697 | 2710007 | DIAMORPHINE 50 MG SUP |
| 30320 | 2723007 | DIAMORPHINE 200 MG SUP |
| 17917 | 50341020 | Dihydrocodeine with paracetamol 20mg with 500mg effervescent tablets |
| 34008 | 53798020 | Dihydrocodeine 30mg tablets (IVAX Pharmaceuticals UK Ltd) |
| 40159 | 61081020 | Dihydrocodeine 10mg/5ml oral solution (Martindale Pharmaceuticals Ltd) |
| 39558 | 55276020 | Dihydrocodeine 30mg tablets (Zentiva) |
| 10023 | 62544020 | Dihydrocodeine with paracetamol 20mg+500mg tablets |
| 48133 | 75444020 | Dihydrocodeine 30mg tablets (Almus Pharmaceuticals Ltd) |
| 34662 | 59895020 | Dihydrocodeine 30mg tablets (Mylan) |
| 3653 | 72309020 | Dihydrocodeine 50mg/1ml solution for injection ampoules |
| 53 | 62538020 | Dihydrocodeine 30mg tablets |
| 21229 | 89742020 | Paracetamol 500mg / Dihydrocodeine 7.46mg effervescent tablets sugar free |
| 59989 | 60123020 | Dihydrocodeine 30mg tablets (Ranbaxy (UK) Ltd) |
| 50532 | 4444020 | Dihydrocodeine 30mg tablets (Bristol Laboratories Ltd) |
| 7989 | 2694007 | DIHYDROCODEINE TARTRATE/ASPIRIN 300 MG TAB |
| 28598 | 89020020 | Paracetamol with dihydrocodeine 500mg +10mg/5ml suspension sugar free |
| 4823 | 62540020 | Dihydrocodeine 40mg tablets |
| 33743 | 89813020 | Dihydrocodeine with paracetamol 7.46mg with 500mg effervescent tablets |
| 34579 | 49542020 | Dihydrocodeine 30mg tablets (Actavis UK Ltd) |
| 5955 | 76478020 | Paracetamol 500mg / Dihydrocodeine 30mg tablets |
| 4950 | 62545020 | Dihydrocodeine with paracetamol 30mg+500mg tablets |
| 191 | 62539020 | Dihydrocodeine 10mg/5ml oral solution |
| 33654 | 49539020 | Dihydrocodeine 30mg tablets (Wockhardt UK Ltd) |
| 30295 | 69749020 | Dihydrocodeine with paracetamol 7.46mg+500mg tablets |
| 34440 | 49549020 | Dihydrocodeine 30mg tablets (A A H Pharmaceuticals Ltd) |
| 2555 | 50340020 | Dihydrocodeine with paracetamol 10mg+500mg tablets |
| 10122 | 89018020 | Dihydrocodeine 10mg with paracetamol 500mg/5ml oral suspension sugar free |
| 64368 | 29690021 | Paracetamol 500mg / Dihydrocodeine 30mg tablets (Icarus Pharmaceuticals Ltd) |
| 65689 | 29446020 | Dihydrocodeine 30mg/5ml oral solution |
| 26653 | !1884501 | DIHYDROCODEINE 10mg/PARACETAMOL 500mg |
| 38521 | 54628020 | Dihydrocodeine 30mg tablets (Teva UK Ltd) |
| 9855 | 66016020 | Paracetamol 500mg / Dihydrocodeine 20mg tablets |
| 14688 | 66014020 | Paracetamol 500mg with dihydrocodeine 10mg tablet |
| 9313 | 62549020 | Dihydrocodeine 90mg modified-release tablets |
| 66121 | 19783020 | Dihydrocodeine 10mg/5ml oral suspension |
| 64079 | 47338020 | Dihydrocodeine 10mg/5ml oral solution (Waymade Healthcare Plc) |
| 6234 | 62550020 | Dihydrocodeine 120mg modified-release tablets |
| 2041 | 62548020 | Dihydrocodeine 60mg modified-release tablets |
| 34730 | 49536020 | Dihydrocodeine 30mg Tablet (Berk Pharmaceuticals Ltd) |
| 19206 | 66015020 | Paracetamol 500mg / Dihydrocodeine 7.46mg tablets |
| 59978 | 47337020 | Dihydrocodeine 30mg tablets (Waymade Healthcare Plc) |
| 21113 | 50342020 | Dihydrocodeine with paracetamol forte 30mg with 500mg effervescent tablets |
| 55425 | 99511020 | Dihydrocodeine 10mg tablets |
| 54354 | 4439020 | Dihydrocodeine 30mg tablets (Kent Pharmaceuticals Ltd) |
| 12020 | 62683020 | Dipipanone 10mg / Cyclizine 30mg tablets |
| 38301 | 61671020 | Cyclizine 30mg with dipipanone 10mg tablets |
| 5697 | 77204020 | Fentanyl 800microgram lozenges |
| 70988 | 9333020 | Fentanyl 50micrograms/hour transdermal patches (Phoenix Healthcare Distribution Ltd) |
| 56670 | 9325020 | Fentanyl 25micrograms/hour transdermal patches (Sigma Pharmaceuticals Plc) |
| 37968 | 94469020 | Fentanyl 40micrograms/dose transdermal system |
| 4691 | 76090020 | Fentanyl 100micrograms/hour transdermal patches |
| 48183 | 73996020 | Fentanyl 100micrograms/2ml solution for injection ampoules (Martindale Pharmaceuticals Ltd) |
| 42399 | 96307020 | Fentanyl 800microgram sublingual tablets sugar free |
| 59678 | 9347020 | Fentanyl 100micrograms/2ml solution for injection ampoules (A A H Pharmaceuticals Ltd) |
| 35853 | 92119020 | Fentanyl 500micrograms/10ml solution for injection ampoules |
| 51235 | 9327020 | Fentanyl 25micrograms/hour transdermal patches (Phoenix Healthcare Distribution Ltd) |
| 55752 | 9345020 | Fentanyl 100micrograms/hour transdermal patches (Phoenix Healthcare Distribution Ltd) |
| 65646 | 24152021 | Fentanyl 267microgram sublingual tablets sugar free |
| 39590 | 96299020 | Fentanyl 200microgram sublingual tablets sugar free |
| 24986 | 80288020 | Fentanyl 1.6mg lozenges |
| 47413 | 72774020 | Fentanyl 75micrograms/hr Transdermal patch (Sandoz Ltd) |
| 5651 | 76092020 | Fentanyl 400microgram lozenges |
| 35330 | 92117020 | Fentanyl 100micrograms/2ml solution for injection ampoules |
| 41135 | 97501020 | Fentanyl 50micrograms/dose nasal spray |
| 59482 | 21649021 | Fentanyl 37.5microgram/hour transdermal patches |
| 617 | 61829020 | Fentanyl 50microgram/ml Injection |
| 54979 | 71753020 | Fentanyl 50micrograms/hour transdermal patches (A A H Pharmaceuticals Ltd) |
| 40576 | 96373020 | Fentanyl 400microgram buccal tablets sugar free |
| 40098 | 96303020 | Fentanyl 400microgram sublingual tablets sugar free |
| 67830 | 53838021 | Fentanyl 12micrograms/hour transdermal patches (DE Pharmaceuticals) |
| 60477 | 71714020 | Fentanyl 25micrograms/hour transdermal patches (A A H Pharmaceuticals Ltd) |
| 63340 | 24150021 | Fentanyl 133microgram sublingual tablets sugar free |
| 620 | 76085020 | Fentanyl 25micrograms/hour transdermal patches |
| 39723 | 96369020 | Fentanyl 100microgram buccal tablets sugar free |
| 63139 | 21931021 | Fentanyl 12micrograms/hour transdermal patches (Phoenix Healthcare Distribution Ltd) |
| 50671 | 21754020 | Fentanyl 12micrograms/hour transdermal patches (A A H Pharmaceuticals Ltd) |
| 40018 | 96371020 | Fentanyl 200microgram buccal tablets sugar free |
| 45092 | 97505020 | Fentanyl 200micrograms/dose nasal spray |
| 757 | 76086020 | Fentanyl 50micrograms/hour transdermal patches |
| 40940 | 96301020 | Fentanyl 300microgram sublingual tablets sugar free |
| 11843 | 76091020 | Fentanyl 200microgram lozenges |
| 15350 | 2583007 | FENTANYL (10ML) 50 MCG/ML INJ |
| 6298 | 76087020 | Fentanyl 75micrograms/hour transdermal patches |
| 41348 | 96305020 | Fentanyl 600microgram sublingual tablets sugar free |
| 7126 | 89905020 | Fentanyl 12micrograms/hour transdermal patches |
| 67766 | 9339020 | Fentanyl 75micrograms/hour transdermal patches (Phoenix Healthcare Distribution Ltd) |
| 61156 | 20345021 | Fentanyl 12micrograms/hour transdermal patches (Waymade Healthcare Plc) |
| 15337 | 77205020 | Fentanyl 1.2mg lozenges |
| 59057 | 20689021 | Fentanyl 400microgram buccal films sugar free |
| 63398 | 16370021 | Fentanyl 2.5mg/50ml solution for infusion vials |
| 45894 | 97503020 | Fentanyl 100micrograms/dose nasal spray |
| 60766 | 20266021 | Fentanyl 25micrograms/hour transdermal patches (Waymade Healthcare Plc) |
| 65437 | 71720020 | Fentanyl 100micrograms/hour transdermal patches (A A H Pharmaceuticals Ltd) |
| 47759 | 98961020 | Fentanyl 400micrograms/dose nasal spray |
| 5696 | 77203020 | Fentanyl 600microgram lozenges |
| 59443 | 20687021 | Fentanyl 200microgram buccal films sugar free |
| 46555 | 96377020 | Fentanyl 800microgram buccal tablets sugar free |
| 24457 | 61832020 | Fentanyl with droperidol 500microgramwith2.5mg/ml Injection |
| 39469 | 96297020 | Fentanyl 100microgram sublingual tablets sugar free |
| 52178 | 9349020 | Fentanyl 100micrograms/2ml solution for injection ampoules (AMCo) |
| 42538 | 96375020 | Fentanyl 600microgram buccal tablets sugar free |
| 53062 | 38912020 | Hydromorphone 10mg/1ml solution for injection ampoules |
| 15798 | 84391020 | Hydromorphone 8mg modified-release capsules |
| 19972 | 84393020 | Hydromorphone 16mg modified-release capsules |
| 9325 | 84390020 | Hydromorphone 4mg modified-release capsules |
| 69063 | 70391021 | Hydromorphone 2mg/1ml solution for injection ampoules |
| 5137 | 84396020 | Hydromorphone 2.6mg capsules |
| 15792 | 84389020 | Hydromorphone 2mg modified-release capsules |
| 5138 | 84397020 | Hydromorphone 1.3mg capsules |
| 24736 | 84394020 | Hydromorphone 24mg modified-release capsules |
| 3239 | 69566020 | Meptazinol 200mg tablets |
| 11801 | 64421020 | Meptazinol 100mg/1ml solution for injection ampoules |
| 21337 | 3672007 | METHADONE 15 MG SUP |
| 60944 | 38790020 | Methadone 5mg capsules |
| 33475 | 64467020 | Methadone 35mg/ml Injection |
| 17671 | 79122020 | Methadone 50mg/1ml solution for injection ampoules |
| 67342 | 15260020 | Methadone 50mg/1ml solution for injection ampoules (Alliance Healthcare (Distribution) Ltd) |
| 33832 | 50026020 | Methadone 1mg/ml oral solution (Martindale Pharmaceuticals Ltd) |
| 30531 | 60731020 | Methadone 1mg/ml oral solution sugar free (Rosemont Pharmaceuticals Ltd) |
| 12132 | 2265007 | METHADONE 5 MG/ML INJ |
| 36436 | 91660020 | Methadone 50mg/5ml solution for injection ampoules |
| 23158 | 81972020 | Methadone 20mg/ml oral solution sugar free |
| 32526 | !8505564 | METHADONE GREEN S/F |
| 6441 | 64465020 | Methadone 5mg tablets |
| 37518 | 91658020 | Methadone 35mg/3.5ml solution for injection ampoules |
| 24584 | 87427020 | Methadone 50mg/2ml solution for injection ampoules |
| 66921 | 47340020 | Methadone 1mg/ml oral solution sugar free (Waymade Healthcare Plc) |
| 68959 | 30916020 | Methadone 20mg/5ml oral solution |
| 62708 | 53150020 | Methadone hydrochloride powder |
| 41608 | 60266020 | Methadone 1mg/ml oral solution (Rosemont Pharmaceuticals Ltd) |
| 59295 | 31679020 | Methadone 100mg capsules |
| 69053 | 86761020 | Pinadone methadone 1mg/ml Oral solution sugar free (Pinewood Healthcare) |
| 29769 | 50027020 | Methadone 2mg/5ml Oral solution (Martindale Pharmaceuticals Ltd) |
| 70267 | 30894020 | Methadone 15mg/5ml oral solution |
| 14086 | 64466020 | Methadone 10mg/ml Injection |
| 41720 | 54093020 | Methadone 1mg/ml Mixture (Macarthy Medical Ltd) |
| 55825 | 66692020 | Methadone 1mg/ml oral solution sugar free (Thornton & Ross Ltd) |
| 28861 | 3290007 | METHADONE 50 MG SUP |
| 24446 | 2269007 | METHADONE 100 MG SUP |
| 5211 | 55631020 | Methadone 2mg/5ml linctus |
| 33068 | 91654020 | Methadone 10mg/1ml solution for injection ampoules |
| 23948 | 2266007 | METHADONE 20 MG SUP |
| 25046 | 82413020 | Methadone diluent Liquid |
| 2952 | 72887020 | Methadone 1mg/ml oral solution |
| 47706 | 50028020 | Methadone 1mg/ml oral solution sugar free (Martindale Pharmaceuticals Ltd) |
| 26801 | 77794020 | Methadone colourant for Liquid |
| 64463 | 31690020 | Methadone 30mg capsules |
| 9728 | 72888020 | Methadone 1mg/ml oral solution sugar free |
| 29911 | 54090020 | Methadone 2mg/5ml linctus (Thornton & Ross Ltd) |
| 63077 | 30900020 | Methadone 1mg/5ml oral suspension |
| 23947 | 3671007 | METHADONE 30 MG SUP |
| 43260 | 94182020 | Methadone Oral solution |
| 11722 | 81971020 | Methadone 10mg/ml oral solution sugar free |
| 23769 | 2268007 | METHADONE 25 MG SUP |
| 36994 | 93940020 | Methadone 5mg/ml oral solution |
| 35506 | 91656020 | Methadone 20mg/2ml solution for injection ampoules |
| 15449 | 2267007 | METHADONE 40 MG SUP |
| 20008 | 6957007 | MORPHINE SULPHATE 20 MG CAP |
| 47753 | 96061020 | Morphine 90mg modified-release capsules |
| 14050 | 83128020 | Morphine sulphate 12 100mg Modified-release capsule |
| 40563 | 96053020 | Morphine 30mg modified-release capsules |
| 26805 | 2219007 | MORPHINE SULPHATE 50 MG SUP |
| 12602 | 59815020 | Morphine sulfate 10mg suppositories |
| 22622 | 3283007 | MORPHINE TARTRATE/CYCLIZINE TARTRATE 15 MG INJ |
| 53273 | 31027020 | Morphine hydrochloride 10mg/5ml oral solution (Special Order) |
| 63423 | 23776021 | Morphine sulfate 10mg/1ml solution for injection ampoules (DE Pharmaceuticals) |
| 45736 | 95525020 | Morphine 60mg modified-release capsules |
| 31650 | 90413020 | Morphine sulfate 30mg/30ml solution for infusion vials |
| 41974 | 51519020 | Morphine sulfate 10mg/1ml solution for injection ampoules (Martindale Pharmaceuticals Ltd) |
| 12583 | 3158007 | MORPHINE SULPHATE CR 5 MG TAB |
| 15781 | 81628020 | Morphine sulphate 24 90mg Modified-release capsule |
| 148 | 3652007 | MORPHINE 60 MG SUP |
| 49976 | 9739020 | Morphine sulfate 10mg/1ml solution for injection ampoules (A A H Pharmaceuticals Ltd) |
| 22026 | 78230020 | Rhotard Morphine SR 30mg tablets (Sovereign Medical Ltd) |
| 54406 | 51526020 | Morphine sulfate 30mg suppositories (Martindale Pharmaceuticals Ltd) |
| 17825 | 6649007 | MORPHINE HCL 5MG/CHLOROFORM WATER TO 5ML SOL |
| 31599 | 2220007 | MORPHINE SULPHATE 4 MG INJ |
| 27298 | 3650007 | MORPHINE SULPHATE EPIDURAL 2 MG INJ |
| 61744 | 40104020 | Morphine sulfate 100mg/50ml solution for infusion vials (A A H Pharmaceuticals Ltd) |
| 54017 | 94727020 | Morphine sulphate Capsule |
| 458 | 70481020 | Morphine 15mg Suppository |
| 12011 | 3161007 | MORPHINE SULPHATE CR 60 MG TAB |
| 22690 | 81630020 | Morphine sulphate 24 120mg Modified-release capsule |
| 14935 | 88732020 | Apomorphine 50mg/10ml solution for infusion pre-filled syringes |
| 47949 | 96067020 | Morphine 120mg modified-release capsules |
| 12608 | 3162007 | MORPHINE SULPHATE CR 200 MG TAB |
| 7517 | 59813020 | Morphine sulfate 15mg suppositories |
| 17271 | 2221007 | MORPHINE SULPHATE 5 MG SUP |
| 16335 | 66055020 | Morphine tartrate 10mg/1ml / Cyclizine tartrate 50mg/1ml solution for injection ampoules |
| 824 | 52832020 | Morphine hydrochloride powder |
| 9183 | 64810020 | Morphine 100mg modified-release tablets |
| 43657 | 96073020 | Morphine 200mg modified-release capsules |
| 8822 | 64807020 | Morphine 60mg modified-release tablets |
| 13997 | 79896020 | Morphine sulphate 100mg Modified-release capsule |
| 4266 | 74008020 | Morphine 10mg tablets |
| 66336 | 50131020 | Morphine sulphate 10mg/ml Injection (Celltech Pharma Europe Ltd) |
| 5563 | 83126020 | Morphine sulphate 12 20mg Modified-release capsule |
| 2425 | 55754020 | Kaolin and Morphine mixture |
| 56202 | 94725020 | Morphine sulphate Injection |
| 13995 | 81632020 | Morphine sulphate 24 200mg Modified-release capsule |
| 8075 | 59714020 | Morphine sulphate 30mg/ml Injection |
| 6269 | 72523020 | Morphine sulfate 20mg/ml oral solution sugar free |
| 56329 | 31888020 | Morphine sulfate 10mg/2ml solution for injection ampoules |
| 12591 | 75453020 | Morphine 60mg modified-release granules sachets sugar free |
| 25503 | 3056007 | AMMONIUM CHLORIDE & MORPHINE MIX |
| 58290 | 4346020 | Morphine sulfate 10mg suppositories (Martindale Pharmaceuticals Ltd) |
| 32459 | 6958007 | MORPHINE SULPHATE 50 MG CAP |
| 10749 | 71718020 | Ipecacuanha and morphine Mixture |
| 70274 | 60040021 | Morphine sulfate 5mg/5ml solution for injection ampoules (Torbay Pharmaceuticals) |
| 33133 | 92077020 | Apomorphine 50mg/5ml solution for injection ampoules |
| 5840 | 68485020 | Morphine sulfate 10mg/5ml oral solution |
| 27749 | 81631020 | Morphine sulphate 24 150mg Modified-release capsule |
| 68712 | 31024020 | Morphine hydrochloride 100mg/5ml oral solution |
| 22024 | 78227020 | Rhotard Morphine SR 10mg tablets (Sovereign Medical Ltd) |
| 11971 | 74663020 | Morphine and Cocaine elixir |
| 28837 | 6896007 | MORPHINE SULPHATE 200 MG CAP |
| 10631 | 64797020 | Morphine 10mg/ml Tincture |
| 31044 | 3651007 | MORPHINE SULPHATE 150 MG SUP |
| 13423 | 3146007 | MORPHINE SULPHATE BP GRANULES 30 MG |
| 55832 | 50132020 | Morphine sulphate 15mg/ml Injection (Celltech Pharma Europe Ltd) |
| 6002 | 79867020 | Morphine 10mg modified-release capsules |
| 9557 | 56797020 | Morphine 15mg modified-release tablets |
| 12219 | 59713020 | Morphine sulfate 15mg/1ml solution for injection ampoules |
| 23581 | 74227020 | Morphine hcl light kaolin, belladonna and aluminium hydroxide chewable tablet |
| 18626 | 3656007 | MORPHINE SULPHATE 200 MG TAB |
| 9960 | 79869020 | Morphine sulphate 12 60mg Modified-release capsule |
| 66815 | 60576021 | Morphine sulfate 10mg/10ml solution for injection ampoules (Hameln Pharmaceuticals Ltd) |
| 25650 | 2224007 | MORPHINE SULPHATE 60 MG SUP |
| 29379 | 2891007 | CHALK AROMAT & MORPHINE MIX |
| 28503 | 6948007 | MORPHINE SULPHATE 100 MG CAP |
| 47867 | 96069020 | Morphine 150mg modified-release capsules |
| 7872 | 5493007 | MORPHINE S/R 64 MG INJ |
| 17092 | 3654007 | MORPHINE HCl 10 MG INJ |
| 61918 | 31894020 | Morphine sulfate 20mg/2ml solution for injection ampoules |
| 25316 | 61674020 | Cyclizine tartrate with morphine tartrate 50mg+10mg/ml injection |
| 50513 | 73083020 | Morphine sulfate 10mg/1ml solution for injection ampoules (UCB Pharma Ltd) |
| 29020 | 75455020 | Morphine 200mg modified-release granules sachets sugar free |
| 8766 | 2229007 | MORPHINE HCl 30 MG INJ |
| 5487 | 74940020 | Apomorphine 2mg sublingual tablets sugar free |
| 18166 | 79897020 | Morphine sulphate 12 200mg Modified-release capsule |
| 9137 | 74009020 | Morphine 20mg tablets |
| 29970 | 73621020 | Morphine hcl and kaolin and belladonna Tablet |
| 23775 | 3920007 | MORPHINE SULPHATE 60 MG INJ |
| 23128 | 3057007 | AMMONIUM CHLOR.& MORPHINE DOUBLE STRENGT MIX |
| 17490 | 61675020 | Cyclizine tartrate with morphine tartrate 50mg+15mg/ml injection |
| 31407 | 2391007 | IPECACUANHA & MORPHINE CONC 1-4 MIX |
| 54520 | 94146020 | Morphine sulphate Oral solution |
| 13225 | 90607020 | Morphine sulfate 30mg/1ml solution for injection ampoules |
| 55365 | 40105020 | Morphine sulfate 50mg/50ml solution for infusion vials (A A H Pharmaceuticals Ltd) |
| 14226 | 56799020 | Morphine 30mg modified-release granules sachets sugar free |
| 41673 | 56575020 | Morphine sulphate 10mg Suppository (Aurum Pharmaceuticals Ltd) |
| 43315 | 96660020 | Morphine sulfate 10mg/1ml suspension for injection vials |
| 20005 | 90409020 | Morphine sulfate 10mg/10ml solution for injection pre-filled syringes |
| 24424 | 65271020 | Morphine hcl and activated attapulgite and attapulgite Tablet |
| 5681 | 64805020 | Morphine 10mg modified-release tablets |
| 20219 | 6905007 | MORPHINE SULPHATE 120 MG CAP |
| 7801 | 3921007 | MORPHINE SULPHATE SR 30 MG TAB |
| 715 | 78856020 | Morphine sulphate 1mg/ml Injection |
| 53106 | 31908020 | Morphine sulfate 5mg/5ml solution for injection ampoules |
| 6892 | 59712020 | Morphine sulphate 10mg/ml Injection |
| 63817 | 54422020 | Morphine hcl Powder (Celltech Pharma Europe Ltd) |
| 14156 | 68487020 | Morphine sulfate 30mg/5ml oral solution unit dose vials sugar free |
| 58215 | 49905020 | Kaolin and Morphine mixture (Thornton & Ross Ltd) |
| 9602 | 64812020 | Morphine 5mg modified-release tablets |
| 63593 | 46331020 | Morphine sulfate 10mg/5ml oral solution (Actavis UK Ltd) |
| 41668 | 48780020 | Morphine hcl 15mg Suppository (Martindale Pharmaceuticals Ltd) |
| 41674 | 50133020 | Morphine sulphate 15mg Suppository (Celltech Pharma Europe Ltd) |
| 10907 | 3159007 | MORPHINE SULPHATE CR 15 MG TAB |
| 34771 | 50378020 | Morphine sulphate 30mg/ml Injection (Celltech Pharma Europe Ltd) |
| 58836 | 31906020 | Morphine sulfate 5mg/1ml solution for injection ampoules |
| 64781 | 46683020 | Morphine sulfate 50mg/5ml solution for injection ampoules |
| 7875 | 64806020 | Morphine 30mg modified-release tablets |
| 55206 | 4587020 | Morphine 20mg modified-release capsules |
| 8420 | 2227007 | MORPHINE SULPHATE 5 MG INJ |
| 35093 | 90415020 | Morphine sulfate 50mg/50ml solution for infusion vials |
| 19738 | 4040007 | MORPHINE,COCAINE & CHLORPROMAZINE MIX |
| 64780 | 20060021 | Morphine sulfate 50mg/1ml solution for injection ampoules |
| 15064 | 6650007 | MORPHINE HCL 10MG/CHLOROFORM WATER > 5ML SOL |
| 29898 | 52762020 | Morphine sulfate powder |
| 6736 | 56798020 | Morphine 20mg modified-release granules sachets sugar free |
| 64417 | 31078020 | Morphine sulfate 2mg/5ml oral solution |
| 56544 | 60696020 | Morphine sulfate 50mg/50ml solution for infusion vials (Martindale Pharmaceuticals Ltd) |
| 42380 | 96059020 | Morphine sulphate 10mg Modified-release capsule |
| 8867 | 3160007 | MORPHINE SULPHATE CR 30 MG TAB |
| 17398 | 66056020 | Morphine tartrate 15mg/1ml / Cyclizine tartrate 50mg/1ml solution for injection ampoules |
| 30049 | 84105020 | Morphine sulphate rapiject 1mg/ml Injection (International Medication Systems (UK) Ltd) |
| 9672 | 75454020 | Morphine 100mg modified-release granules sachets sugar free |
| 8220 | 2228007 | MORPHINE ANHYDROUS 8.4 MG ELI |
| 61241 | 28067020 | Morphine sulfate 50mg/50ml solution for infusion vials (Alliance Healthcare (Distribution) Ltd) |
| 7729 | 50127020 | Morphine hcl Oral solution (Thornton and Ross Ltd) |
| 30597 | 75261020 | Ipecacuanha and Morphine mixture BP 1980 |
| 60507 | 20090021 | Morphine 0.1% in Intrasite gel |
| 34477 | 63736020 | Morphine sulfate 10mg/5ml oral solution (Martindale Pharmaceuticals Ltd) |
| 20783 | 59814020 | Morphine sulfate 30mg suppositories |
| 15815 | 74010020 | Morphine 50mg tablets |
| 24830 | 73053020 | Morphine sulphate 20mg/ml Injection |
| 56788 | 4351020 | Morphine sulfate 10mg/5ml oral solution (A A H Pharmaceuticals Ltd) |
| 23063 | 74224020 | Morphine 1mg/5ml / Peppermint oil 1.5microlitres/5ml oral solution |
| 35255 | 92075020 | Apomorphine 30mg/3ml solution for injection pre-filled disposable devices |
| 32460 | 6895007 | MORPHINE SULPHATE 150 MG CAP |
| 18727 | 3145007 | MORPHINE SULPHATE BP GRANULES 20 MG |
| 61942 | 31892020 | Morphine sulfate 2.5mg/5ml solution for injection ampoules |
| 61423 | 4580020 | Morphine 30mg modified-release tablets (Sigma Pharmaceuticals Plc) |
| 32357 | 2222007 | MORPHINE SULPHATE 300 MG SUP |
| 71462 | 60579021 | Morphine sulfate 1mg/1ml solution for injection ampoules (Torbay Pharmaceuticals) |
| 8740 | 55628020 | Morphine hydrochloride 30mg suppositories |
| 7197 | 79868020 | Morphine sulphate 12 30mg Modified-release capsule |
| 354 | 78857020 | Morphine sulfate 100mg/50ml solution for infusion vials |
| 61584 | 72627020 | Morphine sulfate 50mg/50ml solution for infusion vials (Torbay Pharmaceuticals) |
| 12508 | 3904007 | MORPHINE SULPHATE CR 100 MG TAB |
| 26144 | 71721020 | Morphine with ipecacuanha Mixture |
| 48604 | 9738020 | Morphine sulfate 10mg/1ml solution for injection ampoules |
| 18639 | 3655007 | MORPHINE SULPHATE SR 100 MG TAB |
| 9701 | 74939020 | Apomorphine 10mg/ml injection |
| 30252 | 64802020 | Morphine 8.4mg/ml elixir |
| 58710 | 16243021 | Morphine sulfate 1mg/1ml solution for injection ampoules |
| 655 | 68486020 | Morphine sulfate 10mg/5ml oral solution unit dose vials sugar free |
| 71171 | 60078021 | Morphine sulfate 10mg/10ml solution for injection ampoules (Torbay Pharmaceuticals) |
| 27338 | 73649020 | Morphine hcl and light kaolin and calcium carbonate Tablet |
| 47555 | 90411020 | Morphine sulfate 10mg/10ml solution for injection Minijet pre-filled syringes (UCB Pharma Ltd) |
| 64860 | 46299020 | Morphine sulfate 10mg/10ml solution for injection ampoules |
| 5535 | 74941020 | Apomorphine 3mg sublingual tablets sugar free |
| 9484 | 81627020 | Morphine sulphate 24 60mg Modified-release capsule |
| 21972 | !4629102 | MORPHINE ANHYDROUS |
| 23777 | 3649007 | MORPHINE SULPHATE 100 MG SUP |
| 16189 | 91452020 | Morphine sulphate 10mg/ml Injection |
| 57623 | 73089020 | Morphine sulfate 15mg/1ml solution for injection ampoules (UCB Pharma Ltd) |
| 24816 | 3653007 | MORPHINE ANHYDROUS 8.4 MG INJ |
| 22571 | 57477020 | Morphine sulphate and atropine 10mg + 600microgram/ml Injection |
| 58879 | 31026020 | Morphine hydrochloride 10mg/5ml oral solution |
| 62689 | 46291020 | Morphine hydrochloride 1mg/1ml solution for injection ampoules |
| 29019 | 54447020 | Bismuth with Morphine mixture |
| 61400 | 9357020 | Morphine sulfate 30mg/1ml solution for injection ampoules (A A H Pharmaceuticals Ltd) |
| 61506 | 63148020 | Morphine sulfate 10mg/1ml solution for injection ampoules (Wockhardt UK Ltd) |
| 60518 | 31702020 | Morphine sulfate 500micrograms/5ml oral solution |
| 11698 | 81626020 | Morphine sulphate 24 30mg Modified-release capsule |
| 659 | 70482020 | Morphine 30mg Suppository |
| 27436 | 6904007 | MORPHINE SULPHATE 90 MG CAP |
| 24808 | 2223007 | MORPHINE SULPHATE SUP |
| 43652 | 96063020 | Morphine 100mg modified-release capsules |
| 11838 | 64811020 | Morphine 200mg modified-release tablets |
| 33781 | 92073020 | Apomorphine 20mg/2ml solution for injection ampoules |
| 19291 | 54364020 | Morphine sulfate 100mg/5ml oral solution unit dose vials sugar free |
| 13172 | 78858020 | Morphine sulfate 20mg/1ml solution for injection ampoules |
| 20815 | 56944020 | Morphine sulfate 20mg suppositories |
| 8959 | 54007 | MORPHINE SULPHATE 15 MG TAB |
| 25234 | 52659020 | Chloroform and Morphine tincture |
| 59584 | 4352020 | Morphine sulfate 10mg/5ml oral solution (Alliance Healthcare (Distribution) Ltd) |
| 53918 | 9741020 | Morphine sulfate 10mg/1ml solution for injection ampoules (Hameln Pharmaceuticals Ltd) |
| 60082 | 31904020 | Morphine sulfate 5mg/10ml solution for injection ampoules |
| 53639 | 4578020 | Morphine 10mg modified-release tablets (Sigma Pharmaceuticals Plc) |
| 13588 | 3946007 | MORPHINE SULPHATE susp C/R 30 MG |
| 57750 | 51513020 | Morphine sulfate 15mg/1ml solution for injection ampoules (Wockhardt UK Ltd) |
| 22051 | 52761020 | Morphine sulphate Crystals |
| 60950 | 31053020 | Morphine sulfate 5mg/5ml oral solution |
| 13280 | 90609020 | Morphine sulfate 60mg/2ml solution for injection ampoules |
| 5664 | 55627020 | Morphine hydrochloride 15mg suppositories |
| 5652 | 83127020 | Morphine sulphate 12 50mg Modified-release capsule |
| 19119 | 64909020 | Nalbuphine hc 10mg/ml Injection |
| 6790 | 80382020 | Oxycodone 5mg capsules |
| 45929 | 99590020 | Oxycodone 60mg modified-release tablets |
| 11405 | 76931020 | Oxycodone 10mg/ml oral solution sugar free |
| 27548 | !5251301 | OXYCODONE |
| 40785 | 97290020 | Oxycodone 40mg / Naloxone 20mg modified-release tablets |
| 40688 | 97157020 | Oxycodone 50mg/1ml solution for injection ampoules |
| 35341 | 92085020 | Oxycodone 10mg/1ml solution for injection ampoules |
| 45827 | 99588020 | Oxycodone 30mg modified-release tablets |
| 71335 | 25490021 | Oxycodone 20mg modified-release tablets (Teva UK Ltd) |
| 6414 | 83857020 | Oxycodone hydrochloride 10mg/ml injection |
| 6608 | 79351020 | Oxycodone 20mg modified-release tablets |
| 39498 | 96518020 | Oxycodone 20mg / Naloxone 10mg modified-release tablets |
| 35085 | 92087020 | Oxycodone 20mg/2ml solution for injection ampoules |
| 10866 | 71496020 | Oxycodone hydrochloride 30mg suppositories |
| 45790 | 99586020 | Oxycodone 15mg modified-release tablets |
| 64965 | 78387020 | Oxycodone 5mg/5ml oral solution sugar free (Wockhardt UK Ltd) |
| 6708 | 79352020 | Oxycodone 40mg modified-release tablets |
| 40616 | 97288020 | Oxycodone 5mg / Naloxone 2.5mg modified-release tablets |
| 39475 | 96516020 | Oxycodone 10mg / Naloxone 5mg modified-release tablets |
| 5843 | 79350020 | Oxycodone 10mg modified-release tablets |
| 5585 | 80383020 | Oxycodone 10mg capsules |
| 69474 | 57313021 | Oxycodone 5mg/5ml oral solution sugar free (DE Pharmaceuticals) |
| 58039 | 94116020 | Oxycodone 5mg/5ml oral solution |
| 6609 | 76930020 | Oxycodone 5mg/5ml oral solution sugar free |
| 7275 | 80384020 | Oxycodone 20mg capsules |
| 46187 | 99560020 | Oxycodone 120mg modified-release tablets |
| 6769 | 73071020 | Oxycodone 5mg modified-release tablets |
| 6948 | 76929020 | Oxycodone 80mg modified-release tablets |
| 18261 | 70309020 | Aspirin 500mg with Papaveretum 7.71mg dispersible tablets |
| 166 | 3621007 | PAPAVERETUM 20 MG INJ |
| 11129 | 68940020 | Papaveretum 15.4mg/1ml solution for injection ampoules |
| 19764 | 68941020 | Papaveretum 10mg tablet |
| 6226 | 70312020 | Aspirin 500mg / Papaveretum 7.71mg dispersible tablets sugar free |
| 15353 | 68939020 | Papaveretum 7.7mg/1ml solution for injection ampoules |
| 28732 | 68945020 | Papaveretum with hyoscine 7.7mg with 400 micrograms/ml injection |
| 31584 | 90813020 | Pentazocine 60mg/2ml solution for injection ampoules |
| 328 | 71386020 | Pentazocine 50mg capsules |
| 36472 | 66011020 | Paracetamol 500 mg+ pentazocine 15mg tablet |
| 71170 | 57085020 | Pentazocine 25mg tablets (Actavis UK Ltd) |
| 31582 | 90811020 | Pentazocine 30mg/1ml solution for injection ampoules |
| 17863 | 73655020 | Pentazocine 50mg suppositories |
| 7450 | 65567020 | Pentazocine 15mg with paracetamol 500mg tablet |
| 10583 | 65561020 | Pentazocine 30mg/ml injection |
| 38092 | 60096020 | Pentazocine 30mg/ml Injection (Sterwin Medicines) |
| 2367 | 65564020 | Pentazocine 25mg tablets |
| 53929 | 9366020 | Pethidine 50mg/1ml solution for injection ampoules (A A H Pharmaceuticals Ltd) |
| 57027 | 4480020 | Pethidine 50mg tablets (A A H Pharmaceuticals Ltd) |
| 42708 | 69172020 | Pethidine 50mg/ml intramuscular injection (Roche Products Ltd) |
| 30319 | 53208020 | Pethidine powder |
| 55852 | 51617020 | Pethidine 10mg/ml Injection (Martindale Pharmaceuticals Ltd) |
| 54085 | 94386020 | Pethidine capsule |
| 19116 | 69221020 | Pethidine 100mg/2ml / Promethazine 50mg/2ml solution for injection ampoules |
| 234 | 69166020 | Pethidine 25mg tablet |
| 55839 | 50249020 | Pethidine 50mg/ml Injection (Roche Products Ltd) |
| 38013 | 94823020 | Pethidine 50mg capsules |
| 24867 | 69218020 | Pethidine with levallorphan tartrate injection |
| 33954 | 79285020 | Promethazine hydrochloride 50mg with pethidine 100mg/2ml injection |
| 31885 | 90771020 | Pethidine 100mg/10ml solution for injection ampoules |
| 23442 | 90769020 | Pethidine 50mg/5ml solution for injection ampoules |
| 45325 | 94388020 | Pethidine injection |
| 423 | 69168020 | Pethidine 50mg/ml injection |
| 48148 | 72877020 | Pethidine 100mg/2ml solution for injection ampoules (Actavis UK Ltd) |
| 40239 | 51618020 | Pethidine 50mg tablets (Martindale Pharmaceuticals Ltd) |
| 52400 | 9363020 | Pethidine 100mg/10ml solution for injection ampoules (Alliance Healthcare (Distribution) Ltd) |
| 53709 | 94114020 | Pethidine oral liquid |
| 41550 | 51608020 | Pethidine 100mg/2ml Injection (C P Pharmaceuticals Ltd) |
| 31253 | 64216020 | Pethidine 50mg/ml Injection (Auden McKenzie (Pharma Division) Ltd) |
| 31935 | 50251020 | Pethidine 50mg Tablet (Roche Products Ltd) |
| 17386 | 69171020 | Pethidine 50mg Tablet (Roche Products Ltd) |
| 826 | 90765020 | Pethidine 50mg/1ml solution for injection ampoules |
| 63182 | 9365020 | Pethidine 50mg/1ml solution for injection ampoules (Alliance Healthcare (Distribution) Ltd) |
| 67599 | 72622020 | Pethidine 100mg/2ml solution for injection ampoules (AMCo) |
| 2966 | 51090020 | Pethidine 50mg/ml injection |
| 29426 | 51616020 | Pethidine 50mg/ml Injection (Martindale Pharmaceuticals Ltd) |
| 58190 | 9361020 | Pethidine 50mg/5ml solution for injection ampoules (A A H Pharmaceuticals Ltd) |
| 56022 | 75279020 | Pethidine 50mg Capsule (Martindale Pharmaceuticals Ltd) |
| 17043 | 90767020 | Pethidine 100mg/2ml solution for injection ampoules |
| 249 | 51091020 | Pethidine 10mg/ml injection |
| 58737 | 4481020 | Pethidine 50mg tablets (Alliance Healthcare (Distribution) Ltd) |
| 22896 | 6401007 | PETHIDINE CO 50 MG INJ |
| 48128 | 72883020 | Pethidine 100mg/2ml solution for injection ampoules (Martindale Pharmaceuticals Ltd) |
| 32831 | 50250020 | Pethidine 100mg/2ml Injection (Roche Products Ltd) |
| 54790 | 46333020 | Pethidine 50mg tablets (Teva UK Ltd) |
| 37703 | 72617020 | Pethidine 50mg/1ml solution for injection ampoules (AMCo) |
| 38103 | 53205020 | Pethidine 25mg Tablet (Roche Products Ltd) |
| 2450 | 69167020 | Pethidine 50mg tablets |
| 11046 | 79629020 | Ipratropium bromide with salbutamol 500micrograms + 2.5mg/2.5ml |
| 29195 | 58769020 | Opium 30c Tablet (Weleda (UK) Ltd) |
| 45918 | 95693020 | Opium papaver somniferum D30 Oral drops |
| 9270 | 51108020 | Ipratropium bromide with fenoterol hydrobromide 500micrograms + 1.25mg/4ml |
| 20803 | !3550103 | IPRATROPIUM BROMIDE NEBULISER SOLUTION |
| 25020 | !3550102 | IPRATROPIUM BROMIDE (FORTE) |
| 6772 | 88591020 | Ipratropium bromide 250micrograms/1ml nebuliser liquid unit dose vials |
| 12808 | 61826020 | Fenoterol 100micrograms/dose / Ipratropium bromide 40micrograms/dose breath actuated inhaler |
| 68030 | 70195020 | Ipratropium bromide 250microgram/ml Nebuliser liquid (Approved Prescription Services Ltd) |
| 23961 | 56682020 | Ipratropium bromide 250microgram/ml Inhalation vapour (Galen Ltd) |
| 71346 | 36409020 | Salbutamol 2.5mg/2.5ml / Ipratropium bromide 500micrograms/2.5ml nebuliser liquid ampoules (A A H Pharmaceuticals Ltd) |
| 37791 | 63617020 | Ipratropium bromide 250microgram/ml |
| 12822 | 80090020 | Salbutamol 2.5mg with ipratropium bromide 500micrograms/2.5ml unit dose nebuilser solution |
| 2437 | 74071020 | Oxitropium bromide 100micrograms/dose inhaler |
| 9658 | 74072020 | Oxitropium bromide 100micrograms/dose breath actuated inhaler |
| 44302 | 60757020 | Tropium 10mg tablets (Dr Reddy's Laboratories (UK) Ltd) |
| 6719 | 88593020 | Ipratropium bromide 500micrograms/2ml nebuliser liquid unit dose vials |
| 4639 | 69503020 | Ipratropium bromide 21micrograms/dose nasal spray |
| 18299 | 80152020 | Fenoterol 1.25mg/4ml / Ipratropium 500micrograms/4ml nebuliser liquid unit dose vials |
| 29006 | 3275007 | OPIUM TINCTURE LIQ |
| 34078 | 53111020 | Camphorated opium tincture |
| 53174 | 69151020 | Ipratropium bromide 500micrograms/2ml nebuliser liquid unit dose vials (A A H Pharmaceuticals Ltd) |
| 64232 | 44715021 | Tiotropium bromide 2.5micrograms/dose solution for inhalation cartridge with device CFC free (AM Distributions (Yorkshire) Ltd) |
| 32520 | 53112020 | Opium tincture |
| 27505 | 63621020 | Ipratropium bromide with fenoterol hydrobromide 40micrograms + 100micrograms/actuation |
| 26616 | 63620020 | Ipratropium bromide with fenoterol hydrobromide 0micrograms + 100micrograms/actuation |
| 68530 | 71062021 | Tiotropium bromide 10microgram inhalation powder capsules with device |
| 1415 | 75830020 | Steri-neb ipratropium 250microgram/ml Nebuliser liquid (IVAX Pharmaceuticals UK Ltd) |
| 64509 | 53187021 | Tiotropium bromide 2.5micrograms/dose / Olodaterol 2.5micrograms/dose solution for inhalation cartridge with device CFC free |
| 35011 | 92695020 | Tiotropium bromide 18microgram inhalation powder capsules |
| 40832 | 96099020 | Ipratropium 500micrograms/2ml nebuliser liquid unit dose Steripoule vials (Galen Ltd) |
| 2097 | 69502020 | Ipratropium bromide 20micrograms/metered dose |
| 3786 | 61825020 | Fenoterol 100micrograms/dose / Ipratropium 40micrograms/dose inhaler |
| 1409 | 63608020 | Ipratropium bromide 20micrograms/dose inhaler |
| 48607 | 2813020 | Salbutamol 2.5mg/2.5ml / Ipratropium bromide 500micrograms/2.5ml nebuliser liquid unit dose vials |
| 22183 | 2880007 | CHALK + OPIUM AROMATIC MIX |
| 30229 | 69142020 | Ipratropium bromide 250microgram/ml Nebuliser liquid (Galen Ltd) |
| 1410 | 63615020 | Ipratropium bromide 0.25mg/ml |
| 27880 | 60752020 | Tropium 5mg capsules (Dr Reddy's Laboratories (UK) Ltd) |
| 24691 | 4797007 | LEAD + OPIUM LOT |
| 68039 | 55751020 | Chalk with Opium mixture aromatic BP 1988 |
| 23709 | 88636020 | Ipratropium 500micrograms/2ml nebuliser liquid Steri-Neb unit dose vials (Teva UK Ltd) |
| 6522 | 87822020 | Ipratropium bromide 20micrograms/dose inhaler CFC free |
| 4268 | 63612020 | Ipratropium bromide 40micrograms/dose inhaler |
| 40637 | 96091020 | Ipratropium 250micrograms/1ml nebuliser liquid unit dose Steripoule vials (Galen Ltd) |
| 11779 | 72265020 | Ipratropium bromide 40microgram inhalation powder capsules with device |
| 48410 | 36408020 | Salbutamol 2.5mg/2.5ml / Ipratropium bromide 500micrograms/2.5ml nebuliser liquid ampoules |
| 6081 | 63609020 | Ipratropium bromide 20micrograms/dose breath actuated inhaler |
| 11195 | 85545020 | Opium papaver somniferum 30c Tablet |
| 23276 | 54457020 | Chalk with Opium BP 1988 aromatic mixture |
| 30273 | 60754020 | Tropium 5mg tablets (Dr Reddy's Laboratories (UK) Ltd) |
| 12909 | 80973020 | Salbutamol 100micrograms/dose / Ipratropium 20micrograms/dose inhaler |
| 8333 | 72266020 | Ipratropium bromide 40microgram inhalation powder capsules |
| 6758 | 88632020 | Ipratropium 250micrograms/1ml nebuliser liquid Steri-Neb unit dose vials (Teva UK Ltd) |
| 36864 | 94071020 | Tiotropium bromide 2.5micrograms/dose solution for inhalation cartridge with device CFC free |
| 746 | 73050020 | Tiotropium 18 microgram Capsule |
| 2152 | 75512020 | Ipratropium bromide with salbutamol 20mcg + 100mcg |
| 40177 | 60390020 | Ipratropium bromide 250microgram/ml Nebuliser liquid (Hillcross Pharmaceuticals Ltd) |
| 35014 | 92693020 | Tiotropium bromide 18microgram inhalation powder capsules with device |
| 1411 | 63616020 | Ipratropium bromide 250micrograms/ml |
| 60759 | 24114021 | Tapentadol 20mg/ml oral solution sugar free |
| 47399 | 99825020 | Tapentadol 250mg modified-release tablets |
| 45800 | 99823020 | Tapentadol 200mg modified-release tablets |
| 46019 | 99821020 | Tapentadol 150mg modified-release tablets |
| 45811 | 99801020 | Tapentadol 50mg tablets |
| 46461 | 99803020 | Tapentadol 75mg tablets |
| 46018 | 99819020 | Tapentadol 100mg modified-release tablets |
| 46021 | 99817020 | Tapentadol 50mg modified-release tablets |
| 58316 | 15654021 | Tramadol 50mg modified-release capsules (DE Pharmaceuticals) |
| 4115 | 79961020 | Tramadol 100mg modified-release tablets |
| 43198 | 77140020 | Tramadol sr 50mg Capsule (Hillcross Pharmaceuticals Ltd) |
| 37867 | 92641020 | Tramadol (roi) Tablet |
| 38528 | 66344020 | Tramadol 50mg Capsule (Tillomed Laboratories Ltd) |
| 11748 | 82223020 | Tramadol 400mg modified-release tablets |
| 34260 | 65703020 | Tramadol sr 100mg Modified-release tablet (Winthrop Pharmaceuticals Ltd) |
| 34422 | 57153020 | Tramadol 50mg capsules (Actavis UK Ltd) |
| 64496 | 29694021 | Tramadol 100mg modified-release capsules (Ennogen Healthcare Ltd) |
| 65266 | 4485020 | Tramadol 50mg capsules (Kent Pharmaceuticals Ltd) |
| 11471 | 75787020 | Tramadol 100mg/2ml solution for injection ampoules |
| 68210 | 27275021 | Tramadol 100mg modified-release tablets (Elite Pharma (Surrey) Ltd) |
| 11549 | 79983020 | Tramadol 75mg modified-release tablets |
| 46587 | 78021 | Tramadol 100mg/ml oral drops |
| 48090 | 70246020 | Tramadol 200mg modified-release tablets (A A H Pharmaceuticals Ltd) |
| 61775 | 4489020 | Tramadol 50mg capsules (Sigma Pharmaceuticals Plc) |
| 61610 | 23065021 | Tramadol 50mg capsules (Morningside Healthcare Ltd) |
| 50947 | 4527020 | Tramadol 100mg modified-release capsules (Alliance Healthcare (Distribution) Ltd) |
| 63047 | 20202021 | Tramadol 100mg modified-release capsules (Waymade Healthcare Plc) |
| 5257 | 79962020 | Tramadol 12 Modified-release tablet |
| 52495 | 4490020 | Tramadol 50mg capsules (Bristol Laboratories Ltd) |
| 29860 | 56511020 | Tramadol 50mg capsules (IVAX Pharmaceuticals UK Ltd) |
| 64459 | 45235020 | Tramadol 37.5mg / Paracetamol 325mg tablets (A A H Pharmaceuticals Ltd) |
| 4834 | 79975020 | Tramadol 150mg modified-release capsules |
| 16076 | 87620020 | Paracetamol 325mg with tramadol 37.5 mg tablet |
| 54023 | 4524020 | Tramadol 50mg modified-release capsules (A A H Pharmaceuticals Ltd) |
| 71358 | 4507020 | Tramadol 150mg modified-release tablets (Sigma Pharmaceuticals Plc) |
| 3378 | 75788020 | Tramadol 50mg soluble tablets sugar free |
| 4999 | 82219020 | Tramadol 24 Modified-release tablet |
| 43513 | 60117020 | Tramadol 50mg capsules (Zentiva) |
| 52977 | 4529020 | Tramadol 100mg modified-release capsules (A A H Pharmaceuticals Ltd) |
| 11746 | 82221020 | Tramadol 300mg modified-release tablets |
| 11734 | 87129020 | Tramadol 50mg orodispersible tablets sugar free |
| 68427 | 23070021 | Tramadol 50mg modified-release capsules (CST Pharma Ltd) |
| 52605 | 39814020 | Tramadol 50mg capsules (Accord Healthcare Ltd) |
| 34281 | 65711020 | Tramadol sr 200mg Modified-release tablet (Winthrop Pharmaceuticals Ltd) |
| 11559 | 78590020 | Tramadol 50mg effervescent powder sachets sugar free |
| 8416 | 79963020 | Tramadol 12 Modified-release tablet |
| 34639 | 57229020 | Tramadol 50mg capsules (Genus Pharmaceuticals Ltd) |
| 4114 | 79974020 | Tramadol 100mg modified-release capsules |
| 41976 | 70238020 | Tramadol 100mg modified-release tablets (A A H Pharmaceuticals Ltd) |
| 35347 | 92192020 | Tramadol 24 Modified-release tablet |
| 63898 | 15653021 | Tramadol 50mg modified-release capsules (J M McGill Ltd) |
| 64731 | 27289021 | Tramadol 100mg modified-release capsules (Icarus Pharmaceuticals Ltd) |
| 47854 | 92639020 | Tramadol (roi) Tablet |
| 40718 | 68881020 | Tramadol 50mg capsules (Almus Pharmaceuticals Ltd) |
| 5028 | 82220020 | Tramadol 24 Modified-release tablet |
| 40166 | 61111020 | Tramadol 50mg capsules (Niche Generics Ltd) |
| 6558 | 87618020 | Tramadol 37.5mg / Paracetamol 325mg tablets |
| 46279 | 76711020 | Tramadol 200mg modified-release capsules (A A H Pharmaceuticals Ltd) |
| 701 | 79973020 | Tramadol 50mg modified-release capsules |
| 65954 | 47349020 | Tramadol 50mg modified-release capsules (Cubic Pharmaceuticals Ltd) |
| 34065 | 65707020 | Tramadol sr 150mg Modified-release tablet (Winthrop Pharmaceuticals Ltd) |
| 36732 | 93848020 | Tramadol 50mg modified-release tablets |
| 71355 | 4535020 | Tramadol 150mg modified-release capsules (Waymade Healthcare Plc) |
| 37021 | 90670020 | Tramadol 200mg modified-release tablets |
| 86 | 75786020 | Tramadol 50mg capsules |
| 6215 | 79982020 | Tramadol 200mg modified-release capsules |
| 42280 | 97487020 | Tramadol 37.5mg / Paracetamol 325mg effervescent tablets sugar free |
| 34570 | 59770020 | Tramadol 50mg capsules (Teva UK Ltd) |
| 67197 | 44900021 | Tramadol 50mg capsules (DE Pharmaceuticals) |
| 68833 | 15657021 | Tramadol 100mg modified-release capsules (DE Pharmaceuticals) |
| 9739 | 78591020 | Tramadol 100mg effervescent powder sachets sugar free |
| 60121 | 20201021 | Tramadol 50mg modified-release capsules (Waymade Healthcare Plc) |
| 37020 | 90668020 | Tramadol 150mg modified-release tablets |
| 67161 | 76708020 | Tramadol 150mg modified-release capsules (A A H Pharmaceuticals Ltd) |
| 34521 | 60506020 | Tramadol 50mg capsules (A A H Pharmaceuticals Ltd) |
| 61272 | 4492020 | Tramadol 50mg capsules (Phoenix Healthcare Distribution Ltd) |
| 42798 | 70243020 | Tramadol 150mg modified-release tablets (A A H Pharmaceuticals Ltd) |
| 34808 | 61634020 | Tramadol 50mg capsules (PLIVA Pharma Ltd) |
| 32165 | 59495020 | Tramadol 50mg Capsule (Generics (UK) Ltd) |
| 68989 | 71050021 | Buprenorphine 2mg oral lyophilisates sugar free |
| 6547 | 52971020 | Buprenorphine 2mg sublingual tablets sugar free |
| 40212 | 76007020 | Buprenorphine 8mg sublingual tablets sugar free (Teva UK Ltd) |
| 68988 | 71052021 | Buprenorphine 8mg oral lyophilisates sugar free |
| 64155 | 75999020 | Buprenorphine 400microgram sublingual tablets sugar free (Teva UK Ltd) |
| 6917 | 78619020 | Buprenorphine 52.5micrograms/hour transdermal patches |
| 6056 | 52972020 | Buprenorphine 8mg sublingual tablets sugar free |
| 63788 | 44850021 | Buprenorphine 1mg sublingual tablets sugar free |
| 62675 | 39813020 | Buprenorphine 200microgram sublingual tablets sugar free (A A H Pharmaceuticals Ltd) |
| 40211 | 76002020 | Buprenorphine 2mg sublingual tablets sugar free (Teva UK Ltd) |
| 62776 | 44852021 | Buprenorphine 4mg sublingual tablets sugar free |
| 7236 | 89596020 | Buprenorphine 10micrograms/hour transdermal patches |
| 67018 | 44279021 | Buprenorphine 35micrograms/hour transdermal patches (A A H Pharmaceuticals Ltd) |
| 62874 | 44854021 | Buprenorphine 6mg sublingual tablets sugar free |
| 66463 | 62207021 | Buprenorphine 15micrograms/hour transdermal patches |
| 69942 | 4372020 | Buprenorphine 400microgram sublingual tablets sugar free (Phoenix Healthcare Distribution Ltd) |
| 35681 | 92283020 | Buprenorphine 2mg / Naloxone 500microgram sublingual tablets sugar free |
| 35682 | 92291020 | Buprenorphine 8mg / Naloxone 2mg sublingual tablets sugar free |
| 7334 | 89594020 | Buprenorphine 5micrograms/hour transdermal patches |
| 40473 | 91997020 | Buprenorphine 300micrograms/1ml solution for injection ampoules |
| 396 | 64275020 | Buprenorphine 200microgram sublingual tablets sugar free |
| 11584 | 79157020 | Buprenorphine 70micrograms/hour transdermal patches |
| 320 | 64276020 | Buprenorphine HCl 300micrograms injection |
| 7238 | 89598020 | Buprenorphine 20micrograms/hour transdermal patches |
| 70283 | 69934021 | Buprenorphine 16mg / Naloxone 4mg sublingual tablets sugar free |
| 58273 | 76089020 | Buprenorphine 2mg sublingual tablets sugar free (A A H Pharmaceuticals Ltd) |
| 3064 | 64277020 | Buprenorphine 400microgram sublingual tablets sugar free |
| 62969 | 42183020 | Buprenorphine 8mg sublingual tablets sugar free (Zentiva) |
| 6879 | 83959020 | Buprenorphine 35micrograms/hour transdermal patches |
| 59970 | 4381020 | Buprenorphine 2mg sublingual tablets sugar free (Actavis UK Ltd) |
| 65157 | 4379020 | Buprenorphine 2mg sublingual tablets sugar free (Sigma Pharmaceuticals Plc) |
| 21337 | 3672007 | METHADONE 15 MG SUP |
| 60944 | 38790020 | Methadone 5mg capsules |
| 33475 | 64467020 | Methadone 35mg/ml Injection |
| 17671 | 79122020 | Methadone 50mg/1ml solution for injection ampoules |
| 67342 | 15260020 | Methadone 50mg/1ml solution for injection ampoules (Alliance Healthcare (Distribution) Ltd) |
| 33832 | 50026020 | Methadone 1mg/ml oral solution (Martindale Pharmaceuticals Ltd) |
| 30531 | 60731020 | Methadone 1mg/ml oral solution sugar free (Rosemont Pharmaceuticals Ltd) |
| 12132 | 2265007 | METHADONE 5 MG/ML INJ |
| 36436 | 91660020 | Methadone 50mg/5ml solution for injection ampoules |
| 23158 | 81972020 | Methadone 20mg/ml oral solution sugar free |
| 32526 | !8505564 | METHADONE GREEN S/F |
| 6441 | 64465020 | Methadone 5mg tablets |
| 37518 | 91658020 | Methadone 35mg/3.5ml solution for injection ampoules |
| 24584 | 87427020 | Methadone 50mg/2ml solution for injection ampoules |
| 66921 | 47340020 | Methadone 1mg/ml oral solution sugar free (Waymade Healthcare Plc) |
| 68959 | 30916020 | Methadone 20mg/5ml oral solution |
| 62708 | 53150020 | Methadone hydrochloride powder |
| 41608 | 60266020 | Methadone 1mg/ml oral solution (Rosemont Pharmaceuticals Ltd) |
| 59295 | 31679020 | Methadone 100mg capsules |
| 69053 | 86761020 | Pinadone methadone 1mg/ml Oral solution sugar free (Pinewood Healthcare) |
| 29769 | 50027020 | Methadone 2mg/5ml Oral solution (Martindale Pharmaceuticals Ltd) |
| 70267 | 30894020 | Methadone 15mg/5ml oral solution |
| 14086 | 64466020 | Methadone 10mg/ml Injection |
| 41720 | 54093020 | Methadone 1mg/ml Mixture (Macarthy Medical Ltd) |
| 55825 | 66692020 | Methadone 1mg/ml oral solution sugar free (Thornton & Ross Ltd) |
| 28861 | 3290007 | METHADONE 50 MG SUP |
| 24446 | 2269007 | METHADONE 100 MG SUP |
| 5211 | 55631020 | Methadone 2mg/5ml linctus |
| 33068 | 91654020 | Methadone 10mg/1ml solution for injection ampoules |
| 23948 | 2266007 | METHADONE 20 MG SUP |
| 25046 | 82413020 | Methadone diluent Liquid |
| 2952 | 72887020 | Methadone 1mg/ml oral solution |
| 47706 | 50028020 | Methadone 1mg/ml oral solution sugar free (Martindale Pharmaceuticals Ltd) |
| 26801 | 77794020 | Methadone colourant for Liquid |
| 64463 | 31690020 | Methadone 30mg capsules |
| 9728 | 72888020 | Methadone 1mg/ml oral solution sugar free |
| 29911 | 54090020 | Methadone 2mg/5ml linctus (Thornton & Ross Ltd) |
| 63077 | 30900020 | Methadone 1mg/5ml oral suspension |
| 23947 | 3671007 | METHADONE 30 MG SUP |
| 43260 | 94182020 | Methadone Oral solution |
| 11722 | 81971020 | Methadone 10mg/ml oral solution sugar free |
| 23769 | 2268007 | METHADONE 25 MG SUP |
| 36994 | 93940020 | Methadone 5mg/ml oral solution |
| 35506 | 91656020 | Methadone 20mg/2ml solution for injection ampoules |
| 15449 | 2267007 | METHADONE 40 MG SUP |

Population selection – SUI

| Medical code | Read code | Read term |
| --- | --- | --- |
| 15918 | 1593 | H/O: stress incontinence |
| 5196 | 16F..00 | Double incontinence |
| 109624 | 16F0.00 | Functional urinary and faecal incontinence |
| 6161 | 1A23.00 | Incontinence of urine |
| 110001 | 1A23000 | Functional urinary incontinence |
| 1929 | 1A24.00 | Stress incontinence |
| 5844 | 1A24.11 | Stress incontinence - symptom |
| 5959 | 1A25.00 | Urgency |
| 583 | 1A25.11 | Urgency of micturition |
| 3887 | 1A26.00 | Urge incontinence of urine |
| 93952 | 1A27.00 | Urge to pass urine again shortly after finishing voiding |
| 20728 | 317A.00 | Pad test for incontinence |
| 13424 | 394..11 | Bladder-incontinence assessmnt |
| 13423 | 394..12 | Bladder- continence assessment |
| 13421 | 3940 | Bladder: incontinent |
| 40789 | 39H..00 | Continence assessment |
| 49417 | 39H0.00 | Continence reassessment |
| 29039 | 679H.00 | Health education - continence |
| 29040 | 679H.11 | Promotion of continence |
| 98767 | 7B33800 | Insertion retropubic device stress urinary incontinence NEC |
| 94021 | 7B33C00 | Insertion retropubic dev fem stress urinary incontinence NEC |
| 2739 | 8C14.00 | Incontinence care |
| 12138 | 8C14.11 | Continence care |
| 30981 | 8D7..00 | Urinary bladder control |
| 17637 | 8D7..12 | Incontinence control |
| 48601 | 8D71.00 | Incontinence control |
| 47963 | 8D7Z.00 | Urinary bladder control NOS |
| 9020 | 8E97.00 | Bladder training |
| 29192 | 8H7w.00 | Referral to continence nurse |
| 25899 | 8HTX.00 | Referral to incontinence clinic |
| 106548 | 9NgY.00 | Continence care equipment available at home |
| 94673 | 9Nl8.00 | Seen by continence nurse |
| 3182 | K198.00 | Stress incontinence |
| 17620 | K586.00 | Stress incontinence - female |
| 52763 | Kyu5A00 | [X]Other specified urinary incontinence |
| 107787 | M129500 | Incontinence-associated dermatitis |
| 7649 | R002400 | [D]Micturition syncope |
| 3283 | R083.00 | [D]Incontinence of urine |
| 31220 | R083100 | [D]Urethral sphincter incontinence |
| 17320 | R083200 | [D] Urge incontinence |
| 15400 | R083z00 | [D]Incontinence of urine NOS |
| 8028 | R086200 | [D] Urgency of micturition |
| 73228 | Ryu4000 | [X]Other difficulties with micturition |
| 46614 | Z1J..00 | Procedures to aid continence |
| 45495 | Z9EA.00 | Provision of incontinence appliance |
| 45492 | ZL22400 | Under care of continence nurse |
| 25901 | ZL62400 | Referral to continence nurse |
| 22095 | ZLA2400 | Seen by continence nurse |
| 43234 | ZV65900 | [V] Admission for bladder training |

Population selection – POP

| Medical code | Read code | Read term |
| --- | --- | --- |
| 9803 | 1594 | H/O: genital prolapse |
| 28602 | 7D17.00 | Repair of vaginal prolapse and amputation of cervix uteri |
| 69708 | 7D17y00 | Repair of vaginal prolapse & amputation of cervix uteri OS |
| 15703 | 7D17z00 | Repair of vaginal prolapse & amputation of cervix uteri NOS |
| 11863 | 7D18.00 | Other repair of vaginal prolapse |
| 18606 | 7D18y00 | Other specified other repair of vaginal prolapse |
| 28040 | 7D18z00 | Other repair of vaginal prolapse NOS |
| 6819 | K51..00 | Genital prolapse |
| 1903 | K510.00 | Vaginal wall prolapse without uterine prolapse |
| 211 | K510000 | Cystocele without uterine prolapse |
| 25278 | K510100 | Cystourethrocele without uterine prolapse |
| 2285 | K510200 | Rectocele without uterine prolapse |
| 37918 | K510211 | Proctocele without uterine prolapse |
| 4575 | K510300 | Urethrocele without uterine prolapse |
| 22659 | K510400 | Vaginal prolapse unspecified without uterine prolapse |
| 17481 | K510z00 | Vaginal prolapse without uterine prolapse NOS |
| 1345 | K511.00 | Uterine prolapse without vaginal wall prolapse |
| 22757 | K511000 | First degree uterine prolapse |
| 6882 | K511100 | Second degree uterine prolapse |
| 37790 | K511200 | Third degree uterine prolapse |
| 37185 | K511z00 | Uterine prolapse without vaginal wall prolapse NOS |
| 7870 | K512.00 | Uterovaginal prolapse, incomplete |
| 30419 | K512000 | Cystocele with first degree uterine prolapse |
| 12359 | K512100 | Cystocele with second degree uterine prolapse |
| 9356 | K513.00 | Uterovaginal prolapse, complete |
| 25974 | K513000 | Cystocele with third degree uterine prolapse |
| 1057 | K514.00 | Uterovaginal prolapse, unspecified |
| 12845 | K514000 | Cystocele with unspecified uterine prolapse |
| 10888 | K515.00 | Post hysterectomy vaginal vault prolapse |
| 23941 | K51y.00 | Other genital prolapse |
| 41895 | K51yz00 | Other genital prolapse NOS |
| 33440 | K51z.00 | Genital prolapse NOS |
| 42845 | K534.00 | Prolapse of the ovary and fallopian tube |
| 3578 | K534000 | Prolapse of the ovary |
| 99174 | K534z00 | Ovarian and fallopian tube prolapse NOS |
| 97649 | Kyu9100 | [X]Other female genital prolapse |

Procedures SUI

| Medical code | Read code | Read term | opcs4 |
| --- | --- | --- | --- |
| 11197 | 7B32200 | Introduction of tension free vaginal tape | M53.3 |
| 57283 | 7B32500 | Introduction of transobturator tape | M53.6 |
| 49097 | 7B32y00 | Vaginal operation to support outlet of female bladder OS | M53.8 |
| 48900 | 7B32z00 | Vaginal operation to support outlet of female bladder NOS | M53.8 |
| 33558 | 7B32.00 | Vaginal operations to support outlet of female bladder | M53.8 |
| 60075 | 7M01100 | Insertion of prosthesis into organ NOC | Y02.2 |
| 15696 | 7B31000 | Suprapubic sling operation | M52.1 |
| 21250 | 7B31011 | Aldridge suprapubic sling | M52.1 |
| 4202 | 7B31200 | Colposuspension of bladder neck | M52.3 |
| 36539 | 7B34200 | Endoscopic suburethral injection of inert substance - female | M56.3 |
| 18434 | 7B34211 | Endoscopic suburethral injection of collagen in female | M56.3 |
| 46542 | 7B34212 | Endoscopic suburethral teflon injection in female | M56.3 |
| 51659 | 7B31.00 | Abdominal operations to support outlet of female bladder | M52.8 |
| 97857 | 7B31y00 | Abdominal operation to support outlet of female bladder OS | M52.8 |
| 71003 | 7B31z00 | Abdominal operation to support outlet of female bladder NOS | M52.8 |
| 54329 | 7B35y00 | Other specified other operation on outlet of female bladder | M52.8 |
| 42107 | 7M31100 | Secondary operation NOC | Y71.2 |
| 18256 | 7M31200 | Revisional operation NOC | Y71.3 |
| 45776 | 7M31700 | Second revisional operation NOC | Y71.6 |
| 110714 | 7M31800 | Third or greater revisional operation NOC | Y71.7 |
| 102035 | 7M02000 | Maintenance of prosthesis in organ NOC | Y03.1 |
| 111061 | 7M02100 | Renewal of prosthesis in organ NOC | Y03.2 |
| 60431 | 7M02600 | Correction of displacement of prosthesis NOC | Y03.3 |
| 63624 | 7M02200 | Resiting of prosthesis in organ NOC | Y03.4 |
| 68453 | 7M02400 | Adjustment to prosthesis in organ NOC | Y03.6 |
| 99506 | ZV52y00 | [V]Fitting or adjustment of other specified prosthesis | Y03.6 |
| 39978 | ZV52.00 | [V]Fitting and adjustment of prosthesis | Y03.6 |
| 101516 | ZV52z00 | [V]Fitting or adjustment of unspecified prosthesis | Y03.6 |
| 62250 | ZV52.11 | [V]Prosthesis adjustment | Y03.6 |
| 70457 | 7M02y00 | Attention to prosthesis NOC OS | Y03.8 |
| 98877 | 7M02z00 | Attention to prosthesis NOC NOS | Y03.9 |
| 29132 | 7M02.00 | Attention to prosthesis NOC | Y03.9 |
| 97037 | 7B38900 | Introduction of transobturator sling |  |

Removals SUI

| Medical code | | Read code | | Read term |
| --- | --- | --- | --- | --- |
| medcode | readcode | | readterm | |
| 85494 | 7B32400 | | Partial removal of tension-free vaginal tape | |
| 93869 | 7B32600 | | Removal of transobturator tape | |
| 51857 | 7M02500 | | Removal of prosthesis from organ NOC | |
| 67686 | 7M0J800 | | Removal of other repair material from organ NOC | |
| 49097 | 7B32y00 | | Vaginal operation to support outlet of female bladder OS | |
| 48900 | 7B32z00 | | Vaginal operation to support outlet of female bladder NOS | |
| 33558 | 7B32.00 | | Vaginal operations to support outlet of female bladder | |

Procedures POP

| Medical code | | Read code | Read term | |  |  |
| --- | --- | --- | --- | --- | --- | --- |
| 15911 | 7 | | | Amputation of cervix uteri | |  |
| 28602 | 7D17.00 | | | Repair of vaginal prolapse and amputation of cervix uteri | |  |
| 21695 | 7D17.11 | | | Colporrhaphy and amputation of cervix uteri | |  |
| 7919 | 7D17000 | | | Ant and post colporrhaphy and amputation of cervix uteri | | P22.1 |
| 33955 | 7D17100 | | | Anterior colporrhaphy and amputation of cervix uteri NEC | | P22.2 |
| 4233 | 7D17111 | | | Fothergill anterior colporrhaphy and amputation of cervix | |  |
| 20632 | 7D17200 | | | Posterior colporrhaphy and amputation of cervix uteri NEC | | P22.3 |
| 69708 | 7D17y00 | | | Repair of vaginal prolapse & amputation of cervix uteri OS | | P22.8 |
| 15703 | 7D17z00 | | | Repair of vaginal prolapse & amputation of cervix uteri NOS | | P22.9 |
| 11863 | 7D18.00 | | | Other repair of vaginal prolapse | |  |
| 2286 | 7D18.11 | | | Colporrhaphy | |  |
| 19050 | 7D18000 | | | Anterior and posterior colporrhaphy NEC | | P23.1 |
| 2447 | 7D18100 | | | Anterior colporrhaphy NEC | |  |
| 3145 | 7D18200 | | | Posterior colporrhaphy NEC | | P23.3 |
| 3420 | 7D18300 | | | Repair of enterocele NEC | | P23.4 |
| 72070 | 7D18311 | | | McCall repair of enterocele | |  |
| 67132 | 7D18312 | | | Moschowitz repair of enterocele | |  |
| 37274 | 7D18400 | | | Colporrhaphy NEC | |  |
| 44748 | 7D18500 | | | Anterior mesh vaginal repair | |  |
| 89963 | 7D18600 | | | Paravaginal repair | | P23.5 |
| 89708 | 7D18700 | | | Anterior colporrhaphy with mesh reinforcement | | P23.6 |
| 84375 | 7D18800 | | | Posterior colporrhaphy with mesh reinforcement | | P23.7 |
| 18606 | 7D18y00 | | | Other specified other repair of vaginal prolapse | | P23.8 |
| 28040 | 7D18z00 | | | Other repair of vaginal prolapse NOS | | P23.9 |
| 17020 | 7D19.00 | | | Repair of vault of vagina | |  |
| 66379 | 7D19000 | | | Repair vaginal vault combined abdominal & vaginal approach | |  |
| 52417 | 7D19100 | | | Repair of vault of vagina using abdominal approach NEC | |  |
| 57237 | 7D19200 | | | Repair of vault of vagina using vaginal approach NEC | |  |
| 16175 | 7D19300 | | | Sacrocolpopexy | | P24.2 |
| 1652 | 7D19400 | | | Suspension of vagina NEC | |  |
| 18931 | 7D19500 | | | Sacrospinous fixation of vaginal vault | | P247 |
| 96345 | 7D19600 | | | Repair of vault of vagina with mesh using abdominal approach | | P24.5 |
| 46339 | 7D19700 | | | Repair of vault of vagina with mesh using vaginal approach | | P24.6 |
| 27604 | 7D19y00 | | | Other specified repair of vault of vagina | |  |
| 48218 | 7D19z00 | | | Repair of vault of vagina NOS | |  |
| 8083 | 7D1Ay00 | | | Other specified other repair of vagina | |  |
| 14874 | 7E28000 | | | Suspension of uterus | |  |
| 3835 | 7E28011 | | | Gilliam suspension of uterus | |  |
| 54641 | 7E28300 | | | Endoscopic suspension of uterus | |  |
| 100058 | 7E28500 | | | Suspension of uterus using mesh | | Q54.4 |
| 90156 | 7E28600 | | | Suspension of uterus NEC | |  |
| 102644 | 7E28700 | | | Sacrohysteropexy | | Q54.5 |
| 103999 | 7E28800 | | | Infracoccygeal hysteropexy | | Q54.6 |
| 42107 | 7M31100 | | | Secondary operation NOC | | Y71.2 |
| 18256 | 7M31200 | | | Revisional operation NOC | | Y71.3 |
| 45776 | 7M31700 | | | Second revisional operation NOC | | Y71.6 |
| 110714 | 7M31800 | | | Third or greater revisional operation NOC | | Y71.7 |
| 102035 | 7M02000 | | | Maintenance of prosthesis in organ NOC | | Y03.1 |
| 111061 | 7M02100 | | | Renewal of prosthesis in organ NOC | | Y03.2 |
| 60431 | 7M02600 | | | Correction of displacement of prosthesis NOC | | Y03.3 |
| 63624 | 7M02200 | | | Resiting of prosthesis in organ NOC | | Y03.4 |
| 68453 | 7M02400 | | | Adjustment to prosthesis in organ NOC | | Y03.6 |
| 99506 | ZV52y00 | | | [V]Fitting or adjustment of other specified prosthesis | | Y03.6 |
| 39978 | ZV52.00 | | | [V]Fitting and adjustment of prosthesis | | Y03.6 |
| 101516 | ZV52z00 | | | [V]Fitting or adjustment of unspecified prosthesis | | Y03.6 |
| 62250 | ZV52.11 | | | [V]Prosthesis adjustment | | Y03.6 |
| 70457 | 7M02y00 | | | Attention to prosthesis NOC OS | | Y03.8 |
| 98877 | 7M02z00 | | | Attention to prosthesis NOC NOS | | Y03.9 |
| 29132 | 7M02.00 | | | Attention to prosthesis NOC | | Y03.9 |

Removals POP

| Medical code | | Read code | | Read term |  |  |
| --- | --- | --- | --- | --- | --- | --- |
| 8083 | 7D1Ay00 | | Other specified other repair of vagina | | |  |
| 1504 | 7D1Az00 | | Other repair of vagina NOS | | |  |
| 27604 | 7D19y00 | | Other specified repair of vault of vagina | | | P24.8 |
| 28040 | 7D18z00 | | Other repair of vaginal prolapse NOS | | | P23.8 |
| 18606 | 7D18y00 | | Other specified other repair of vaginal prolapse | | | P23.8 |
| 11863 | 7D18.00 | | Other repair of vaginal prolapse | | | P23.8 |
| 97867 | 7E28y00 | | Other specified operation on other uterine ligament | | | Q54.8 |
| 67017 | 7E28z00 | | Operation on other uterine ligament NOS | | | Q54.8 |
| 67686 | 7M0J800 | | Removal of other repair material from organ NOC | | | Y26.4 |
|  |  | | **OPCS4 text** | | |  |
|  |  | | Total removal of prosthetic material from previous repair of vaginal prolapse | | | P28.1 |
|  |  | | Partial removal of prosthetic material from previous repair of vaginal prolapse | | | P28.2 |
|  |  | | Total removal of prosthetic material from previous repair of vaginal vault prolapse | | | P30.1 |
|  |  | | Partial removal of prosthetic material from previous repair of vaginal vault prolapse | | | P30.2 |
|  |  | | Total removal of prosthetic material from previous suspension of uterus | | | Q54.7 |
|  |  | | Partial removal of prosthetic material from previous suspension of uterus | | | Q57.1 |
